# Sex specificity in DNA Methylation Profiles of Neonates Following Early Preterm Birth

**DOI:** 10.64898/2026.11.12.26367906

**Authors:** Nojus Jampolskis, Hannah R Elliott, Michellle A. Williams, Sixto Sanchez, Raul Ortega, Maria Luisa Jimenez Sanchez, Bizu Gelaye, Diana Juvinao-Quintero

## Abstract

Preterm birth (PTB) affects 11% of pregnancies worldwide, yet the biological mechanisms underlying its etiology remain unclear. We conducted a sex-stratified epigenome-wide association study of newborn saliva DNA methylation in 273 mother-child pairs from the Peruvian Placental Abruption Genetic Epidemiology (PAGE) study. Robust linear regression models adjusted for maternal, cellular, and technical factors identified 318 and 542 ePTB-associated CpG sites and 784 and 1,401 differentially methylated regions in boys and girls, respectively, with limited overlap between sexes. Functional analyses revealed enrichment of signals in Open Sea regions and pathways related to neurodevelopment, neural function, fetal growth, cellular organization, transcription factors, and microRNAs. Several associations replicated findings from prior studies of PTB, gestational age, and post-menstrual age. These results identify sex-specific epigenetic signatures of ePTB in an Admixed-American population and highlight developmental and regulatory pathways associated with ePTB.

## Introduction

Preterm birth (PTB) is a leading cause of neonatal and infant morbidity and mortality. PTB is associated with long-term health impairments for the newborn, including respiratory distress syndrome, necrotizing enterocolitis, intraventricular hemorrhage, neurodevelopmental delays and mortality in adulthood.^1–5^ PTB affects around 11% of pregnancies worldwide, totaling nearly 15 million births.^6^ Despite its substantial public health impact, the biological mechanisms underlying PTB and early PTB (ePTB), a more extreme outcome representing births occurring 34 weeks of gestation, remain insufficiently understood. Epigenetic processes, particularly DNA methylation (DNAm), are increasingly recognized as central in fetal development and may provide insight into biological pathways associated with PTB and its subsequent health consequences.

Maternal psychosocial stressors, including history of trauma exposure and mood disorders, are known risk factors for PTB and adverse neonatal outcomes.^1,7–9^ These exposures have been linked to DNAm variation in pathways related to stress regulation, immune function, and neurodevelopment, suggesting a plausible biological route through which maternal experiences shape offspring health.^10–13^ Although these maternal factors are not the primary focus of the present paper, they provide important context for interpreting DNAm signatures associated with PTB, particularly in populations with high psychosocial burden.

Prior epigenome-wide association studies (EWAS) have identified DNAm variation associated with gestational age,^14,15^ PTB,^16–18^ postnatal morbidity,^19^ and early-life neurobehavioral outcomes.^20^ However, DNAm associations with ePTB specifically remain understudied. Additionally, evidence from high-risk and under-represented populations is particularly limited, even though there is a disproportionate burden of ePTB in many low-and middle-income settings.^1,21,22^

To this end, we conducted an EWAS to identify DNAm differences between infants born early preterm compared to term-born peers using neonatal saliva DNAm collected at delivery. Our study is conducted in a deeply phenotyped cohort recruited in Lima, Peru, a population that is characterized by high rates of both maternal psychosocial stressors and of ePTB.^23,24^ By characterizing DNAm signatures of ePTB, we sought to identify biological pathways represented among ePTB-associated signals and provide a foundation for future work examining links among maternal stress, DNAm variation, and neonatal health.

## Results

### Baseline characteristics of the study population

The 273 mother–child dyads included in the analytic sample were similar to eligible PAGE participants on most baseline characteristics, with differences observed in newborn sex, cesarean delivery, and perceived maternal health during pregnancy (Supplementary Table 1). Description of baseline characteristics of participants included in this study is shown in Table 1. Specifically, women with an early preterm pregnancy were on average older than controls and were more likely to give birth to a boy than a girl. Infants born early preterm were on average smaller, more frequently remained in a neonatal intensive care unit (NICU), had a lower APGAR score, and were more prone to experience neonatal death than their term-born counterparts.

**Table 1.**
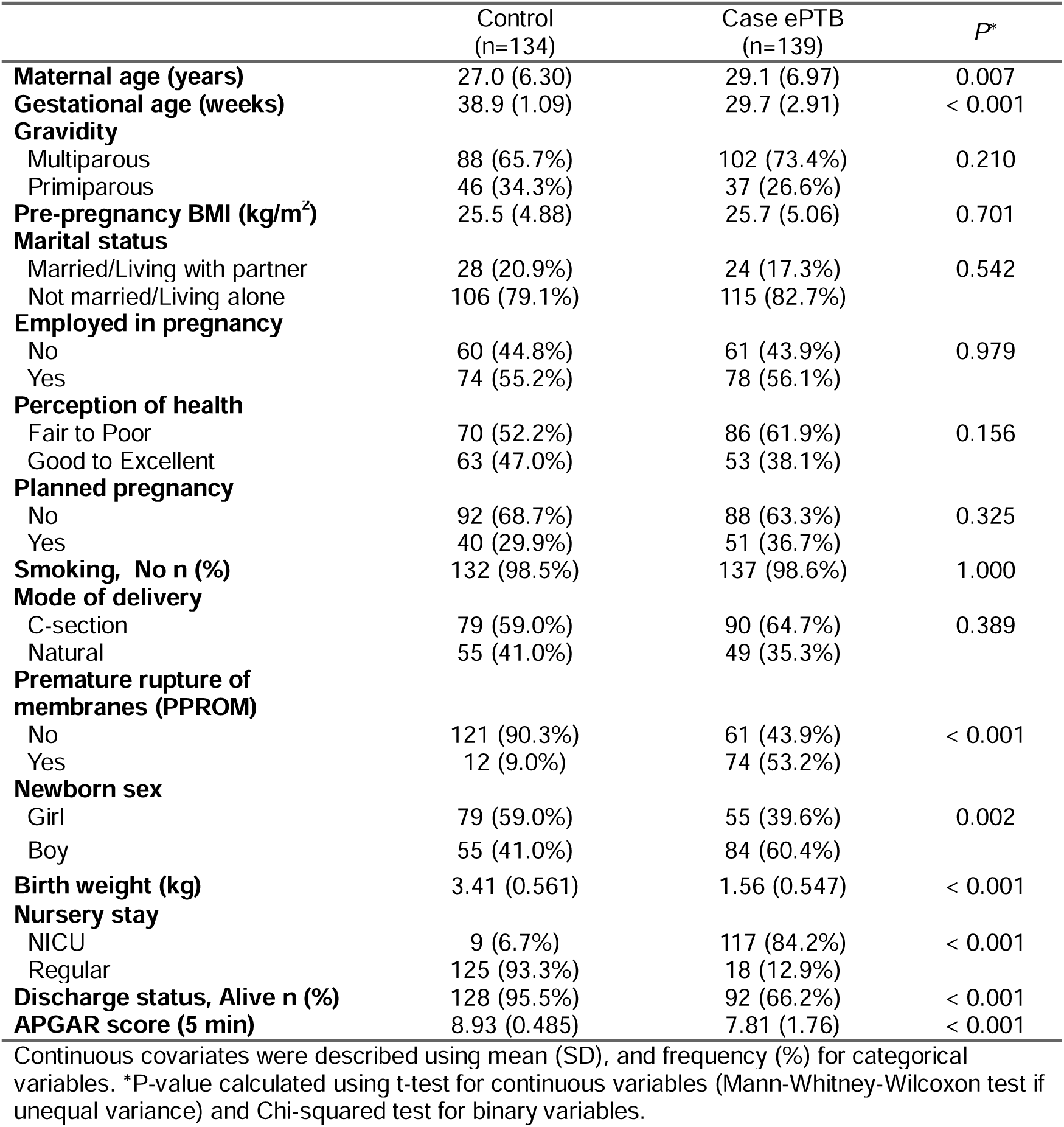
Description of study population at baseline (N=273).

|  | Control<br>(n=134) | Case ePTB<br>(n=139) | *P*<sup>\*</sup> |
|---|---|---|---|
| **Maternal age (years)** | 27.0 (6.30) | 29.1 (6.97) | 0.007 |
| **Gestational age (weeks)** | 38.9 (1.09) | 29.7 (2.91) | < 0.001 |
| **Gravidity** |  |  |  |
| Multiparous | 88 (65.7%) | 102 (73.4%) | 0.210 |
| Primiparous | 46 (34.3%) | 37 (26.6%) |  |
| **Pre-pregnancy BMI (kg/m<sup>2</sup>)** | 25.5 (4.88) | 25.7 (5.06) | 0.701 |
| **Marital status** |  |  |  |
| Married/Living with partner | 28 (20.9%) | 24 (17.3%) | 0.542 |
| Not married/Living alone | 106 (79.1%) | 115 (82.7%) |  |
| **Employed in pregnancy** |  |  |  |
| No | 60 (44.8%) | 61 (43.9%) | 0.979 |
| Yes | 74 (55.2%) | 78 (56.1%) |  |
| **Perception of health** |  |  |  |
| Fair to Poor | 70 (52.2%) | 86 (61.9%) | 0.156 |
| Good to Excellent | 63 (47.0%) | 53 (38.1%) |  |
| **Planned pregnancy** |  |  |  |
| No | 92 (68.7%) | 88 (63.3%) | 0.325 |
| Yes | 40 (29.9%) | 51 (36.7%) |  |
| **Smoking, No n (%)** | 132 (98.5%) | 137 (98.6%) | 1.000 |
| **Mode of delivery** |  |  |  |
| C-section | 79 (59.0%) | 90 (64.7%) | 0.389 |
| Natural | 55 (41.0%) | 49 (35.3%) |  |
| **Premature rupture of membranes (PPROM)** |  |  |  |
| No | 121 (90.3%) | 61 (43.9%) | < 0.001 |
| Yes | 12 (9.0%) | 74 (53.2%) |  |
| **Newborn sex** |  |  |  |
| Girl | 79 (59.0%) | 55 (39.6%) | 0.002 |
| Boy | 55 (41.0%) | 84 (60.4%) |  |
| **Birth weight (kg)** | 3.41 (0.561) | 1.56 (0.547) | < 0.001 |
| **Nursery stay** |  |  |  |
| NICU | 9 (6.7%) | 117 (84.2%) | < 0.001 |
| Regular | 125 (93.3%) | 18 (12.9%) |  |
| **Discharge status, Alive n (%)** | 128 (95.5%) | 92 (66.2%) | < 0.001 |
| **APGAR score (5 min)** | 8.93 (0.485) | 7.81 (1.76) | < 0.001 |

Continuous covariates were described using mean (SD), and frequency (%) for categorical variables. \*P-value calculated using t-test for continuous variables (Mann-Whitney-Wilcoxon test if unequal variance) and Chi-squared test for binary variables.

### Genome-wide difference in newborn DNAm is associated with ePTB

We conducted EWAS stratified by sex to account for underlying variation in DNAm driven by this variable in our sample. In the boys’ analysis, we identified 318 CpGs differentially methylated in association with ePTB ( =1.41, Fig 1A) with effect estimates ranging from - 0.22 to 0.17. There was an almost equal distribution between hypomethylated and hypermethylated signals (55% vs 45%). The strongest association based on the *p*-value was detected at cg00861010, located near the long non-coding RNA *SLFNL1-AS1*. In the girls’ analysis, we identified 542 CpGs ( =1.61, Fig 1B), most of them hypomethylated in relation to ePTB (64%). The effect estimates ranged from -0.22 to 0.21, and the strongest signal based on the *p*-value was seen at cg00221747, mapping near the maternally expressed transcript of the long non-coding RNA *H19*. We identified 24 CpGs in common between boys and girls, with the smallest *p-*value seen at cg13933718, mapping near the *UNQ6494* gene. Overall, for top CpGs, there was low correlation in the effect estimates across sex-analyses (*r* ranged from 0.03 to 0.08, *p* > 0.10). Complete results of the sex-stratified EWAS of ePTB are presented in Supplementary Tables 2-3, while Table 2 shows the top ten signals selected by *p*-value and effect size (absolute effect estimate > 0.1) for each sex-stratified analysis.

**Figure 1.**
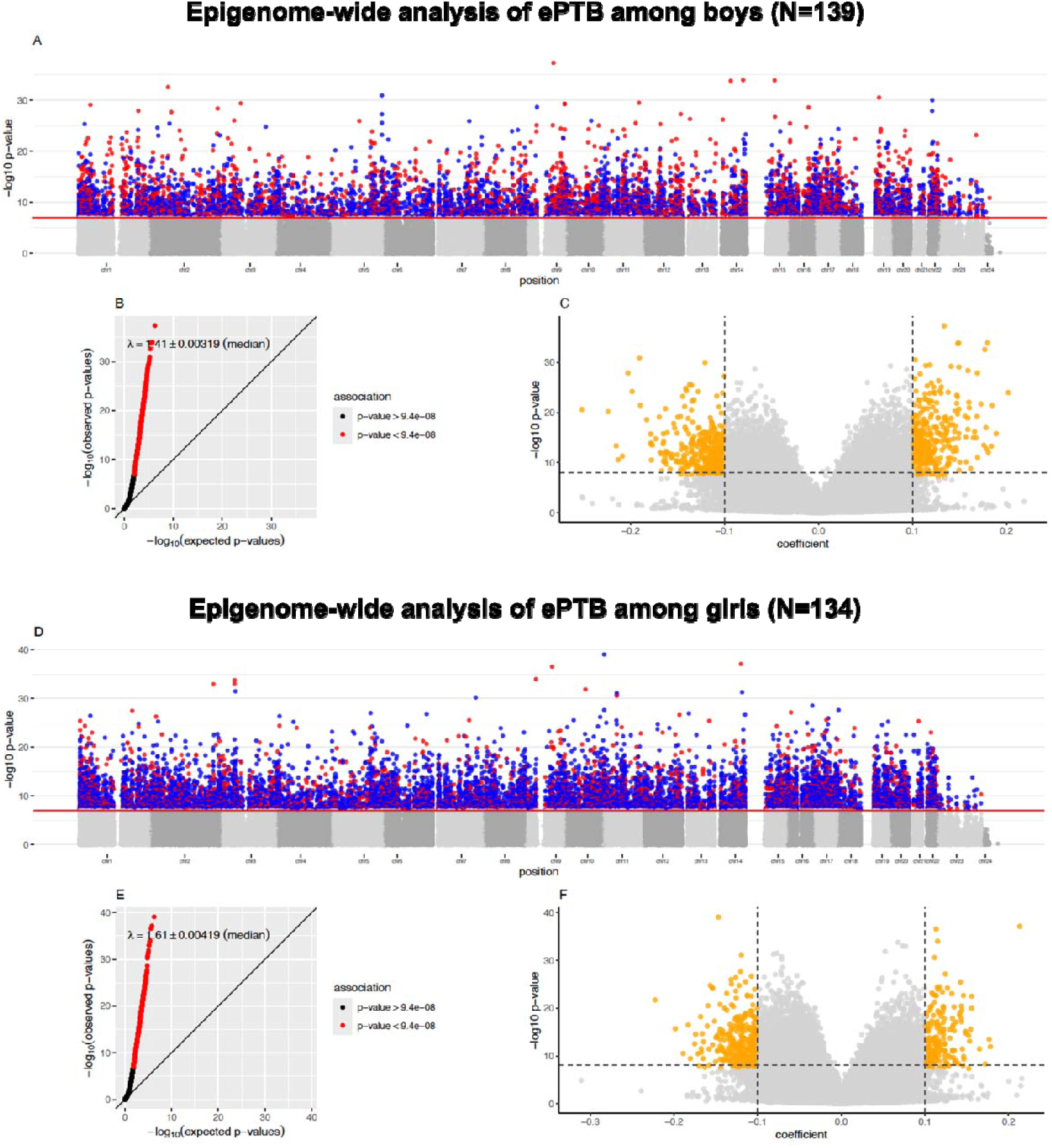
Epigenome-wide analysis after *bacon* adjustment of early preterm birth stratified by sex among participants from the PAGE study (N=273). (a-c) Manhattan, Q-Q plot, and volcano plot in the boys’ analysis (n=139); (d-f) similar plots in the girls’ analysis (n=134). P-threshold for epigenome-wide significance was set at *p* < 9.4x10^-8^ (solid horizontal red line in Manhattan plots). Top CpG sites were also selected based on an absolute effect estimate > 0.1.

**Table 2.** Top associations identified with epigenome-significance in the sex-stratified EWAS of early preterm birth (Boys N=139, Girls N=134). Describing top 10 most significant CpGs that were unique to the boys’ analysis (n=295 CpGs), the girls’ analysis (n=519 CpGs), and signals identified in common between analyses (n=24 CpGs). Associations were deemed significant at *p* < 9.4x10^-8^.

| **CpG** | **Chr** | **Position** | **Near Gene** | **Gene context** | **CG island context** | **Boys** |  | **Girls** |  |
|---|---|---|---|---|---|---|---|---|---|
|  |  |  |  |  |  | **Effect size (95% CI)** | ***P*** | **Effect size (95% CI)** | ***P*** |
| *Boys' unique associations* |  |  |  |  |  |  |  |  |  |
| cg00861010 | 1 | 41500442 | *SLFNL1-AS1* | lncRNA | S_Shelf | 0.13(0.11,0.15) | 8.7x10<sup>-29</sup> | 0.05(0.03,0.06) | 2.4x10<sup>-12</sup> |
| cg04308291 | 2 | 234189621 | *ATG16L1* | intronic | Open sea | 0.14(0.12,0.17) | 2.5x10<sup>-26</sup> | -0.04(-0.06,-0.03) | 2.5x10<sup>-12</sup> |
| cg02458632 | 2 | 62450946 | *B3GNT2* | UTR3 | Open sea | 0.16(0.13,0.19) | 6.4x10<sup>-26</sup> | -0.05(-0.07,-0.04) | 1.2x10<sup>-15</sup> |
| cg11343820 | 5 | 174617516 | *LINC01951;DRD1* | intergenic | Open sea | -0.17(-0.2,-0.14) | 3.6x10<sup>-27</sup> | 0.08(0.06,0.1) | 3.9x10<sup>-12</sup> |
| cg14871997 | 5 | 174618039 | *LINC01951;DRD1* | intergenic | Open sea | -0.11(-0.13,-0.09) | 5.0x10<sup>-26</sup> | -0.05(-0.06,-0.04) | 1.5x10<sup>-16</sup> |
| cg13413384 | 9 | 134410385 | *UCK1;PRRT1B* | intergenic | S_Shelf | 0.13(0.1,0.15) | 9.3x10<sup>-25</sup> | -0.09(-0.11,-0.07) | 5.3x10<sup>-17</sup> |
| cg14887720 | 10 | 33118141 | *CCDC7* | intronic | Open sea | 0.11(0.08,0.13) | 2.9x10<sup>-23</sup> | -0.06(-0.08,-0.05) | 4.5x10<sup>-15</sup> |
| cg01714756 | 14 | 94314063 | *PRIMA1;FAM181A-AS1* | intergenic | Open sea | 0.16(0.13,0.19) | 1.6x10<sup>-24</sup> | -0.09(-0.11,-0.07) | 1.2x10<sup>-16</sup> |
| cg19489600 | 14 | 51337991 | *ABHD12B* | upstream | Open sea | 0.12(0.1,0.14) | 2.2x10<sup>-23</sup> | -0.04(-0.05,-0.03) | 7.8x10<sup>-10</sup> |
| cg07222631 | 22 | 27809086 | *LINC02554;MN1* | intergenic | Open sea | -0.18(-0.22,-0.15) | 1.4x10<sup>-26</sup> | -0.06(-0.08,-0.05) | 5.5x10<sup>-17</sup> |
| *Girls' unique associations* |  |  |  |  |  |  |  |  |  |
| cg02635407 | 4 | 8198822 | *ABLIM2;SH3TC1* | Intergenic | Open Sea | 0.06(0.04,0.08) | 2.4x10<sup>-10</sup> | 0.11(0.09,0.14) | 4.1x10<sup>-27</sup> |
| cg00221747 | 11 | 2131198 | *H19;IGF2* | Intergenic | N_Shore | -0.12(-0.16,-0.09) | 1.1x10<sup>-12</sup> | -0.15(-0.17,-0.12) | 8.7x10<sup>-40</sup> |
| cg01668279 | 11 | 2135469 | *H19;IGF2* | Intergenic | S_Shelf | -0.11(-0.13,-0.08) | 2.9x10<sup>-16</sup> | -0.12(-0.14,-0.1) | 2.2x10<sup>-28</sup> |
| cg21370660 | 11 | 45249998 | *PRDM11* | UTR3 | Open Sea | 0.06(0.05,0.08) | 3.5x10<sup>-13</sup> | -0.11(-0.14,-0.09) | 3.1x10<sup>-26</sup> |
| cg17266233 | 11 | 46361175 | *DGKZ* | Intronic | Open Sea | -0.07(-0.09,-0.06) | 3.4x10<sup>-19</sup> | 0.11(0.09,0.13) | 2.7x10<sup>-31</sup> |
| cg02865822 | 11 | 46361591 | *DGKZ* | Intronic | Open Sea | -0.09(-0.1,-0.07) | 3.1x10<sup>-23</sup> | -0.12(-0.14,-0.1) | 7.4x10<sup>-32</sup> |
| cg16943230 | 14 | 91466411 | *RPS6KA5* | Intronic | Open Sea | 0.14(0.1,0.18) | 3.2x10<sup>-11</sup> | 0.21(0.18,0.25) | 7.9x10<sup>-38</sup> |
| cg06285241 | 14 | 105478318 | *CDCA4* | Intronic | S_Shelf | -0.06(-0.08,-0.04) | 2.6x10<sup>-8</sup> | -0.11(-0.13,-0.09) | 2.0x10<sup>-27</sup> |
| cg19752649 | 16 | 11114847 | *CLEC16A* | Intronic | Open Sea | -0.09(-0.12,-0.06) | 1.5x10<sup>-10</sup> | 0.12(0.1,0.15) | 7.0x10<sup>-28</sup> |
| cg00810262 | 17 | 50673068 | *LINC01982;LINC02089* | Intergenic | OpenSea | 0.05(0.03,0.07) | 6.1x10<sup>-10</sup> | -0.13(-0.16,-0.11) | 1.1x10<sup>-26</sup> |
| *Common associations* |  |  |  |  |  |  |  |  |  |
| cg21902516 | 3 | 13767651 | *LINC00620* | ncRNA | Open sea | -0.1(-0.13,-0.08) | 3.8x10<sup>-19</sup> | -0.12(-0.15,-0.09) | 4.9x10<sup>-17</sup> |
| cg03876023 | 8 | 103348635 | *UBR5* | Intronic | Open sea | -0.22(-0.28,-0.17) | 2.5x10<sup>-16</sup> | 0.15(0.11,0.18) | 7.1x10<sup>-16</sup> |
| cg13933718 | 9 | 92725373 | *UNQ6494;LOC101927847* | Intergenic | Open sea | 0.11(0.08,0.13) | 1.3x10<sup>-21</sup> | 0.11(0.10,0.13) | 2.8x10<sup>-37</sup> |
| cg16109426 | 9 | 36566818 | *RNF38;MELK* | Intergenic | Open sea | 0.11(0.09,0.13) | 7.5x10<sup>-21</sup> | 0.12(0.10,0.13) | 1.0x10<sup>-34</sup> |
| cg10355819 | 9 | 121464001 | *LINC02578* | ncRNA | Open sea | 0.12(0.09,0.15) | 5.5x10<sup>-18</sup> | 0.12(0.10,0.14) | 4.0x10<sup>-24</sup> |
| cg18295744 | 10 | 79113169 | *KCNMA1-AS3* | ncRNA | Open sea | 0.12(0.09,0.14) | 8.2x10<sup>-20</sup> | -0.11(-0.14,-0.08) | 4.7x10<sup>-12</sup> |
| cg02723558 | 11 | 27514526 | *LOC105376671;LIN7C* | Intergenic | Open Sea | 0.17(0.13,0.21) | 8.2x10<sup>-17</sup> | -0.11(-0.14,-0.09) | 4.7x10<sup>-16</sup> |
| cg19944895 | 11 | 62914623 | *SLC22A24;SLC22A25* | Intergenic | Open sea | -0.12(-0.15,-0.09) | 3.5x10<sup>-15</sup> | 0.11(0.08,0.14) | 2.3x10<sup>-14</sup> |
| cg05616242 | 12 | 54389632 | *HOXC-AS2* | ncRNA | Open sea | 0.10(0.08,0.12) | 9.8x10<sup>-16</sup> | -0.11(-0.13,-0.08) | 4.0x10<sup>-20</sup> |
| cg21854374 | 16 | 84737742 | *USP10* | Intronic | Open sea | 0.13(0.09,0.16) | 3.3x10<sup>-15</sup> | 0.12(0.1,0.15) | 6.9x10<sup>-20</sup> |

The negative control analysis showed lambda values considerably smaller than in the main analyses ( =1.05 vs 1.4-1.6 in sex-stratified EWAS), and only two sites surpassed EWAS significance (cg20654161 and cg02294020), none of them overlapping with ePTB sites detected in our main analyses. These findings provide evidence that the main EWAS results were unlikely to be explained entirely by systematic features of the data detectable using this negative-control approach.

### Identification of DMRs

We performed a DMR screen with Bonferroni correction for each sex-stratified EWAS result. This analysis revealed 1,401 DMRs in girls and 784 in boys, collectively comprising 3,482 and 2,197 CpG sites, respectively (Supplementary Tables 4-5). Among male samples, the largest DMR mapped to *EGFL8* and encompassed 16 probes, all of which exhibited negative coefficients. Notably, one probe within this region was independently associated with ePTB in boys. For females, the two largest DMRs mapped to *KCNQ1* and *APBB1IP*, each harboring 12 probes; however, neither of these DMRs contained probes significantly linked to ePTB in girls.

### Enrichment analysis for genomic regions and biological pathways

Compared to other CpGs analyzed across the genome, the top CpGs associated with ePTB in the boys’ and girls’ analyses were more likely to be found in open sea regions than expected by chance (*p*<0.005; Supplementary Fig 1), rather than within CpG islands, or any other genomic region. In the boys’ analysis, inspection across different functional databases revealed enrichment of ePTB-associated CpGs for various Gene Ontology (GO) biological processes (neuron development, neuron projection development, neuron differentiation, signal transduction, and cell communication), molecular functions (protein binding), cellular components (synapse, cell junction, cytoplasm), and for transcription factors (*HES2, ZXDL*, *MAZ, ED2F3, CKROX*), micro RNAs, and pathways affecting insulin-like growth factor *IGF1* Akt signaling (Supplementary Fig 2, Supplementary Table 6). In the girls’ analysis, genes mapping to ePTB-associated CpGs were particularly enriched for transcription factors (n=100 terms), micro-RNAs (n=28 terms), cerebral cortex and hippocampus proteins (n=4 terms), GO cellular components (cytosol, nucleoplasm, cytoplasm, cell junction) and biological processes (cellular response to stimulus and organelle organization). Enrichment was also seen for the *KCNQ2* and *KCNQ3* related epilepsy pathway in girls (Supplementary Fig 3, Supplementary Table 7).

### Comparison of ePTB-associated CpGs with previous EWAS

We compared signals in common between our sex-stratified EWAS of ePTB and previous EWAS of gestational age (GA) and post-menstrual age (PMA) in very preterm infants (< 30 gestational weeks)^25^, PTB^6,26^, and gestational age continuous.^27,28^ PMA is a measure of aging and neonatal development defined as the period from conception to saliva sample collection at NICU discharge.^25^ We identified the largest overlap with associations reported with GA (n=35 sites in common) and PMA (n=348) in the study by Hodge *et al*.^25^. Most of the signals identified in common with GA were directionally consistent with our study, particularly when the comparison was done with the boys-identified ePTB associations. Some GA CpGs in common to highlight are cg07164639 (*DDO*), cg21658153 (*TPM4*), and cg00187327 (*MN1*), mapping to known age-related and PTB loci (Supplementary Table 8). As expected, there was disparity in the direction of associations for CpGs detected in common between ePTB and PMA across studies. Some PMA CpGs in common to highlight are cg07164639 (*DDO*), cg00187327, cg07222631, cg19034820 (*MN1*), cg09253641 (*SMIM2*), cg03185144 (*KIAA0513*), and cg17806418 (*GABBR1*), all located in known loci related with age, neurodevelopment, and PTB.^25,26,29^ When looking at signals previously reported in a large meta-analysis of GA continuous (27-42 weeks) using cord-blood DNAm^28^, we found 16 CpGs in common with our ePTB study (Supplementary Table 9). More than half of these 16 CpGs showed opposite direction of association with our study, and absolute effect estimates were consistently larger in our EWAS.

We also identified some overlap between our ePTB signals and PTB-related DMRs reported by Winchester *et al*.^6^ using buccal cell DNAm from triads. These PTB-DMR genes have a role in transcription (*LDB2, CAMTA1, ZNF717, ZFHX3*), cytoskeleton (*CTNNA3, BTBD11*), receptors (*MERTK*), cellular signaling (*PRKDC*), or other roles (*CRTAC1, GPC6, LINC01170*); however, there were few directional consistencies in these associations with our study. Similarly, we found an overlap at the *SMIM2* gene reported by Weather *et al*. ^27^ in association with GA. Even though CpGs detected at *SMIM2* differed between studies, there was consistency in the direction of associations at this locus across studies. Lastly, in the comparison with findings from Fernando *et al.*^26^, loci but not specific CpGs associated with PTB, were found in common at *KRT78, CHN2*, *TBCD, KIAA0513, UVRAG, GALNT9, UVRAG*, and *DNAH10*; 66% of these loci showed the same direction of association as in our study.

## Discussion

Our study analyzed data from 273 mother–child dyads, of which 139 were cases of ePTB. Sex-stratified analyses identified 318 and 542 CpG sites with saliva methylation levels associated with ePTB in boys and girls, respectively. Most of these sites were hypomethylated, which is consistent with the literature, as increasing gestational age is associated with more hypermethylated sites.^25^ Only 24 of these sites overlapped between sexes, indicating substantial sex heterogeneity in the observed associations. In secondary analyses, we identified 784 and 1,401 DMRs associated with ePTB for boys and girls, respectively. Enrichment analyses revealed further distinctions: boys exhibited enrichment of ePTB-associated pathways across well-defined biological processes, with a focus on neurodevelopment, whereas girls predominantly showed enrichment for regulatory pathways, with transcription factors and micro-RNAs being the most common pathways identified. We discuss the results separately for each sex.

Forty-five percent of the CpG sites significantly associated with ePTB in boys were hypermethylated. The most significant hypermethylated CpG site by p-value is a novel site for ePTB mapping to a long non-coding RNA region near *SLFNL1 Antisense RNA 1* (*SLFNL1-AS1*). Experimental evidence has shown that *SLFNL1-AS1* interacts with proteins encoded by *HNRNPH1*, a gene that plays a critical role in cancer development.^30^ The two most significant hypomethylated sites were also previously unreported CpG sites, both mapping to an intergenic region near the *Dopamine Receptor D1* (*DRD1*), which encodes the most abundant dopamine receptor in the brain and has been implicated in mental and neuronal diseases^31^. The largest DMR in boys mapped to *EGF Like Domain Multiple 8* (*EGFL8*), a protein-coding gene associated with immune system-related diseases such as asthma and Type 1 diabetes mellitus through genome-wide association studies (GWAS).^32,33^ Up to 209 ePTB-associated CpG sites identified in boys overlapped with CpGs that were previously reported in an EWAS of GA and PMA by Hodge *et al*.^34^, and 12% of them showed consistent directionality between studies. Genes replicated in our study include *DDO*, which controls neurological homeostasis^35^, and *MN1*, which has evidence of tumor suppression.^36^ Enrichment analysis of pathways identified many GO biological processes related to neurodevelopment in boys. Some of these pathways, such as neuron projection development and cell morphogenesis involved in neuron differentiation, were replicated by Hodge *et al.*^25^, while others, including neuron development and neuron differentiation, were previously unreported but clearly linked to known associated pathways. Lastly, a pathway related to insulin-like growth factor 1 (*IGF1*) and Akt signaling was found with enrichment among ePTB-associated CpGs in boys. This pathway is important because proteins in the IGF-axis are associated with cellular growth, proliferation, and metabolism throughout the life course, and *IGF1* is particularly related with fetal growth and development ^37^, and with postnatal growth.^38^ These findings suggest that DNAm differences associated with ePTB may involve pathways relevant to fetal and postnatal growth.

In girls, thirty-six percent of CpG sites associated with ePTB were hypermethylated. The most significant hypermethylated CpG site was a previously unreported site mapping to an intergenic region near *Ribosomal Protein S6 Kinase A5* (*RPS6KA5*), which regulates inflammatory response and early gene expression and can promote neuronal death.^39^ The most significant hypomethylated site was a previously unreported CpG mapping to an intergenic region near the *H19 Imprinted Maternally Expressed Transcript* (*H19*) and the paternally expressed *Insulin Like Growth Factor 2* (*IGF2*) genes. These genes are implicated in tumor suppression and fetal growth, respectively, thus pointing to their interrelated role in postnatal development and growth.^40^ *H19*/*IGF2* were also overrepresented in several significant DMRs, providing more evidence of their involvement in fetal-growth regulatory pathways linked to ePTB. The two largest DMRs, with 12 CpGs each, were nearest to *KCNQ1* and *APBB1IP*. *KCNQ1* is associated with Jervell and Lange-Nielsen Syndrome 1, a fetal cardiovascular disease.^41^ *APBB1IP* plays a central role in T-cell activation and thus affects immune response, as well as being linked to cancer.^42^ There was less replication and directional consistency of associations identified in girls with previous studies of PTB and GA. Nonetheless, we found notable overlap with Hodge *et al.*^34^ for *GABBR1.* This gene has previously been linked to epilepsy.^43^ Pathway enrichment analysis for ePTB-associated CpGs in girls mainly identified enrichment for micro-RNAs and transcription factors. Most micro-RNAs identified in this way were replicated by Winchester *et al*.^16^. Lastly, we identified a *KCNQ2* and *KCNQ3*-related epilepsy pathway. *KCNQ2* and *KCNQ3* are part of a family of voltage-gated potassium channel proteins with a function in cellular excitatory activity, particularly in the brain.^44^ Pathogenic variants in these genes have been associated with neonatal-onset developmental and epileptic encephalopathy (NEO-DEE), self-limited (benign) familial neonatal epilepsy (SLFNE), and with neurodevelopmental disorder with or without seizures (NDD)^45^. Benign forms of epilepsies may also occur in preterm infants.^46^ Thus, findings among girls highlight epigenetic signals involving pathways relevant to fetal growth, energy regulation, and neurodevelopment.

Our study provides novel CpG sites, DMRs, and pathways associated with ePTB in an Admixed-American population. Additionally, several signals overlapped with findings from studies conducted predominantly in populations of European ancestry, suggesting that some ePTB-associated DNAm patterns may be shared across population groups. To ensure our results were robust, we controlled for cell composition and batch effects and used Bacon adjustment to reduce genomic inflation. We also thoroughly inspected for data structure and reduced latent variation in DNAm by stratifying our data and analyses by sex. To verify the reliability of our findings, we applied a negative control analysis, corrected for multiple testing using an epigenome-wide significance threshold recommended for the EPIC array, and compared our results with existing evidence. To the best of our knowledge, this is the first such study in a South American population, helping to reduce gaps in epigenetic research across populations.

Our study also has some limitations. Due to the cross-sectional study design, we cannot make causal claims, as we cannot determine from these data when DNAm differences occurred, whether before or after delivery. Even though timing of sample collection relative to delivery was known for these participants (within 72 hours), precise assessment of PMA and other markers of aging and development among preterms was not possible as we did not have information about time of discharge from the NICU. In relation to the above, differences in DNAm observed could have been associated with other pregnancy complications related to ePTB, or with stressful exposures occurring at or around delivery rather than by ePTB alone; this possibility warrants further examination. The methylation data were obtained from saliva, which may not reflect methylation in other tissues, like brain, blood, or placenta which are more often utilized for the study of fetal neurodevelopment. We identified many novel associations that may be attributable to population differences^47^ and/or to higher rates of maternal prenatal stressors that could influence the relationship between ePTB and dysregulation in newborn DNAm. These possibilities require further study in similar populations.

Our study identified multiple novel epigenetic associations with ePTB in an admixed-Amerindian sample of Peruvian women, while also replicating some associations previously reported in European samples. These results suggest that ePTB-associated DNAm patterns include both signals shared with previous population groups and signals that may be population-specific or phenotype-specific. We also observed sex-specific associations with ePTB, with few signals shared between boys and girls. In both cases, most differentially methylated sites were hypomethylated in relation to ePTB, which is consistent with the existing literature (hypermethylation with increasing GA or aging). Although pathways enriched in differentially methylated genes for ePTB consistently pointed to neurodevelopment, cellular growth and function, immune response, and neurological outcomes, our results showed that for girls these processes were more influenced by regulatory elements like transcription factors and micro-RNAs, while for boys it was by the biological process or molecular function itself, suggesting sex-specific patterns in the epigenetic features associated with ePTB. Replication in independent and diverse populations, ideally with longitudinal biospecimen collection, will be important for determining the timing, reproducibility, and potential biological relevance of these associations.

## Materials and Methods

### Study population

Mother-child pairs included in the study were part of the Placental Abruption Genetic Epidemiology (PAGE) study, a case-control study conducted in Lima, Peru. Description of the study can be found elsewhere.^48,49^ Briefly, eligible women were recruited at delivery from seven hospitals in Lima, Peru, by reviewing admission logbooks for the emergency room, labor and delivery wards, prenatal wards, emergency, and surgery rooms of the study hospitals, between March 2013 and December 2015. Eligibility criteria included speaking Spanish, being > 18 years old, having a singleton pregnancy, and complete medical information to define ePTB status. All study participants gave their written informed consent; the study protocols were approved by the IRB of the seven participating hospitals in Lima, the Swedish Medical Center, Seattle, WA, and the Harvard T.H. Chan School of Public Health.

### Data collection

Information on sociodemographic (age, employment, marital status), lifestyle (smoking and alcohol use in pregnancy), behavioral, and anthropometric variables (pre-pregnancy BMI), were gathered using structured questionnaires. Obstetric history and medical information of the current pregnancy (gestational age at delivery, gravidity, planned pregnancy, mode of delivery, birthweight, sex, discharge status, and APGAR score) were abstracted from medical records.

### Definition of ePTB

Gestational age at delivery (weeks) was calculated using information from the date of the last menstrual period, and from an early fetal ultrasound and fundal height examinations performed < 20 weeks of gestation. Early preterm birth was defined following the American College of Obstetrics and Gynecology criteria as a delivery occurring 20-34 weeks of gestation.^50^ Based on their pathophysiology,^51^ cases are either grouped as spontaneous (45%) or with preterm premature rupture of fetal membranes (PPROM; 54%). There were no cases of medically induced PTB. Controls were women who delivered at term (> 37 weeks and < 42 weeks of gestation) and were randomly selected from the participating hospitals within 48 hours of the identification of a case.

### Analytic Population

For the current study, we randomly selected 152 mom-child dyads with ePTB and 152 matched controls from the entire cohort with complete covariates of interest, and archived newborn biosamples with the highest quality of genomic DNA extracted from saliva samples collected 72 hours post-delivery (total eligible *N*=1,009 , *n*=468 with ePTB).

### Processing of DNA methylation data

Newborn buccal cells were collected post-delivery, plated, and stored using the Oragene™saliva cell collection kits (OGR250, DNA Genotek Ottawa, Canada). Samples were processed at Roswell Park Comprehensive Cancer Center (Buffalo, NY) for DNA extraction using Qiagen DNeasy™ system (Qiagen, Valencia, CA), and DNA methylation quantification using llumina MethylationEPIC v2 Beadarray (Illumina, San Diego, CA). For DNAm assessment, 500 ng of genomic DNA from each sample was plated at random into 96-well plates, followed by bisulphite conversion treatment using the Zymo EZ DNA methylation kit. This was followed by amplification, fragmentation, hybridization, single-base extension, and staining. Methylation levels were analyzed at 937,055 CpGs across the genome.

Before analyses, DNAm data were pre-processed using the *meffil* R package,^52^ where probes were quantile normalized using the first twenty principal components (PC) from the control probe matrix, and samples excluded based on outliers for the median methylated or unmethylated signal (n=1), sex-mismatch between reported and predicted sex (n=15), and X-Y ratio outliers (n=5). Probes with detection p-value > 0.01 (n=1,937) and bead number lower than three (n=90) in more than 20% of the samples were excluded. In addition, n=824 probes (nv-GRCh3 probes) with missing values for all the samples were excluded. After exclusions, 284 samples and 927,761 probes remained for the analysis. The proportion of epithelial cells and seven immune cell types was calculated from DNAm using a saliva reference panel.^53^ We inspected for factors driving underlying variation in DNAm by plotting the first two PCs derived from the 20,000 most variable probes, against different confounding factors. We identified that sex was driving a large proportion of the underlying variation in DNAm. Thus, our main analysis was stratified by sex.

### Covariates

We selected covariates a priori based on supporting literature, and by inspection of variability and multicollinearity. We included as covariates maternal age (missing n=2), gravidity (missing n=3), pre-pregnancy BMI (Kg/m^2^) (missing n=5), and employment in pregnancy (missing n=1), as these are established confounders in prenatal epigenetic studies, and known risk factors for PTB.^28,54,55^ In addition, we adjusted for batch effects and the proportion of epithelial cells and seven immune cell types predicted using DNAm data.^53^ We did not include ethnicity (most participants were mixed Amerindian and European descent by genetic ancestry analysis)^48^ and maternal smoking (< 3% smokers) as covariates, as they showed low variability in our sample. Newborn sex was used as the stratification factor in our main analysis. After exclusion of missing values for covariates, 273 (ePTB=139, controls=134) samples with high-quality DNAm remained in the study for analysis.

### Statistical Analysis

We described the characteristics of participants using the mean and standard deviation (SD), or frequency and proportions. We conducted epigenome-wide assessment of differential methylation at the individual CpG sites in relation to ePTB using fitted robust linear regressions stratified by sex (boys=139, girls=134) in the *ewaff* R package.^56^ In this analysis, DNAm Beta (range 0 to 1) were modelled as the response variable, and the binary event of ePTB as the predictor. The confounders described above were included in the models. We controlled for batch effects and non-biological variability remaining in the DNAm matrix using a surrogate variable analysis (n=10 SVs).^57^ Fitted robust linear regression was used to account for potential heteroskedasticity in the DNAm distribution. We calculated the genomic inflation factor (γ) using the ratio of the median observed versus the median expected chi-square test statistic distribution from the EWAS p-values. To reduce remaining variance in the test statistic and the likelihood of false positives, estimates of the EWAS were further corrected using *bacon* adjustment.^58^ To account for multiple testing, statistical significance was defined using a *p-value* threshold < 9.4x10^-8^ , consistent with recommendations for the Illumina EPIC array.^59^ “Top CpGs” were defined based on *p*-value and an absolute effect estimate > 0.1. EWAS results were visualized using Manhattan, volcano, and Q-Q plots generated using the *meffil* and *ggplot2* R packages. Association estimates were presented for top CpGs unique to each sex-stratified analysis, and for signals shared between analyses. All analyses were conducted in R 4.5.1.

In a sensitivity analysis, we checked for systematic sources of bias in our data by conducting a negative control analysis. In this analysis, we used the complete sample (N=273 dyads), permuted ePTB status, adjusted for the same covariates as in the main analysis, and tested for genome-wide associations. Obtaining similar results between the negative control analysis and the main analysis would suggest the presence of residual confounding in the main analysis or other systematic sources of bias still present in the data.

### Secondary analyses

We examined the biological function of signals detected in the EWAS of ePTB by conducting a series of functional inspections. First, to account for the correlation between CpGs across the genome, and to more comprehensively examine associations, we looked at genomic regions rather than individual CpGs by conducting a differentially methylated regional (DMR) analysis using *dmrff* R package.^60^ We selected robustly associated DMRs at regional Bonferroni-adjusted *p* < 0.05 and CpG count above one. For top CpGs of the sex-stratified EWAS, we performed enrichment analysis for genomic regions using Illumina annotations, and enrichment for genes and biological pathways using *gprofiler2* R package.^61^ Enrichment was deemed at FDR < 0.05 after multiple-testing correction from log_2_(OR) tests. We also identified overlap between top CpGs identified in our EWAS, and top sites reported in previous epigenetic studies of PTB and/or ePTB and gestational age.^6,25–27^ For associations identified in common, we compared directionality consistency between studies.

## Data Availability

The phenotype and methylation data that support the findings of this study are available in Figshare with the identifier https://doi.org/10.6084/m9.figshare.34036587.62

## Data Availability

The phenotype and methylation data that support the findings of this study are available in Figshare with the identifier https://doi.org/10.6084/m9.figshare.34036587

## Acknowledgements

The authors are grateful to participants from the different hospitals who contributed to the PAGE study. We also want to recognize the technical and administrative contributions to the study of Ms. Elena Sanchez and research staff from the Association Civil PROESA. This study was supported by awards from the *Eunice Kennedy Shriver* National Institute of Child Health and Human Development, National Institutes of Health (1R21HD113774-01A1: Z99 HD999999). HRE works in the Medical Research Council Integrative Epidemiology Unit at the University of Bristol, UK, which is supported by the Medical Research Council and the University of Bristol (MC_UU_00032/5). For the purpose of open access, the authors have applied a ’Creative Commons Attribution (CC BY) public copyright license to any Author Accepted Manuscript (AAM) version arising from this submission

## Author Contributions

DJQ, BG, SS and HRE designed and conceptualized the study. NJ and DJQ performed the analyses. NJ was co-supervised by DJQ and HRE. DJQ, NJ and HRE drafted the initial version of the manuscript. BG, SS, RO, MW, MJ reviewed the contents of the manuscript. All authors contributed to the interpretation of findings and are responsible for the intellectual content of the published article. The contributions of Dr. Gelaye were made as part of his official duties as a National Institutes of Health (NIH) federal employee, are in compliance with agency policy requirements, and are considered Works of the United States Government. However, the findings and conclusions presented in this paper are those of the author(s) and do not necessarily reflect the views of the NIH or the U.S. Department of Health and Human Services.

## Competing Interests

The authors declare no competing interests.

## Supplementary Tables and Figures legend

**Supplementary Table 1.** Comparison of baseline characteristics between our analytic sample for the EWAS of ePTB (N=273), and the eligible sample from the PAGE study (n=1,009). Eligible participants were those who met the inclusion criteria for the study and had genomic DNA extracted from newborn saliva samples for DNA methylation analysis.

**Supplementary Table 2.** Associations identified with epigenome-significance in the EWAS of early preterm birth among boys (N=139). Showing effect estimates observed for same signals in the girls’ analysis for comparison. Associations were deemed significant at *p* < 9.4x10^-8^.

**Supplementary Table 3.** Associations identified with epigenome-significance in the EWAS of early preterm birth among girls (N=134). Showing effect estimates observed for same signals in the boys’ analysis for comparison. Associations were deemed significant at *p* < 9.4x10^-8^.

**Supplementary Table 4.** Differentially methylated regions associated with ePTB in boys (N=139). DMRs were selected at regional Bonferroni-adjusted *p* < 0.05.

**Supplementary Table 5.** Differentially methylated regions associated with ePTB in girls (N=134). DMRs were selected at regional Bonferroni-adjusted *p* < 0.05.

**Supplementary Table 6.** Functional enrichment analysis for top CpGs identified in the EWAS of ePTB in boys (n=318 CpGs at *p* < 9.4x10^-8^). Analysis performed using the *KnowyourCG* R package with functional terms observed across multiple databases. Enrichment of a functional term based on FDR < 0.05.

**Supplementary Table 7.** Functional enrichment analysis for top CpGs identified in the EWAS of ePTB in girls (n=542 CpGs at *p* < 9.4x10^-8^). Analysis performed using the *KnowyourCG* R package. Enrichment deemed at FDR < 0.05.

**Supplementary Table 8.** Look-up of CpGs in common between our sex-stratified EWAS of ePTB, and the EWAS of gestational age and pre-menstrual age (PMA) among very preterm infants (< 30 weeks) reported by Hodge *et al.* 2024.

**Supplementary Table 9.** Look-up of CpGs in common between our sex-stratified EWAS of ePTB and the meta-analysis of gestational age reported by Merid *et al.* 2020.

**Supplementary Fig 1.** Functional enrichment analysis for genomic regions and CpG island context for CpG sites detected in the EWAS of ePTB in a) boys, b) girls, c) signals in common between analyses. DHS: CpGs identified across analyses were primarily enriched in Open Sea regions (*p* < 0.05). DNAse Hypersensitive regions.

**Supplementary Fig 2.** Graphical view of pathways, biological processes and other molecular functions enriched among CpGs identified in the EWAS of ePTB in boys (n=318 CpGs at *p* < 9.4x10^-8^). Analysis performed using *gProfiler*. Presenting top terms with the lowest adjusted-*p* for enrichment across multiple functional databases.

**Supplementary Fig 3.** Graphical view of pathways, biological processes and other molecular functions enriched among CpGs identified in the EWAS of ePTB in girls (n=542 CpGs at *p* < 9.4x10^-8^). Analysis performed using *gProfiler*. Presenting top terms with the lowest adjusted-*p* for enrichment across multiple functional databases.

## Notes

### Competing Interest Statement

The authors have declared no competing interest.

### Author Declarations

All study participants gave their written informed consent; the study protocols were approved by the IRB of the seven participating hospitals in Lima, the Swedish Medical Center, Seattle, WA, and the Harvard T.H. Chan School of Public Health.

