## Supplemental Figures for "Sex specificity in DNA Methylation Profiles of Neonates Following Early Preterm Birth"

**Supplementary Fig 1.** Functional enrichment analysis for genomic regions and CpG island context for CpG sites detected in the EWAS of ePTB in a) boys, b) girls, c) signals in common between analyses. DHS: CpGs identified across analyses were primarily enriched in Open Sea regions ($p < 0.05$). DNAse Hypersensitive regions.

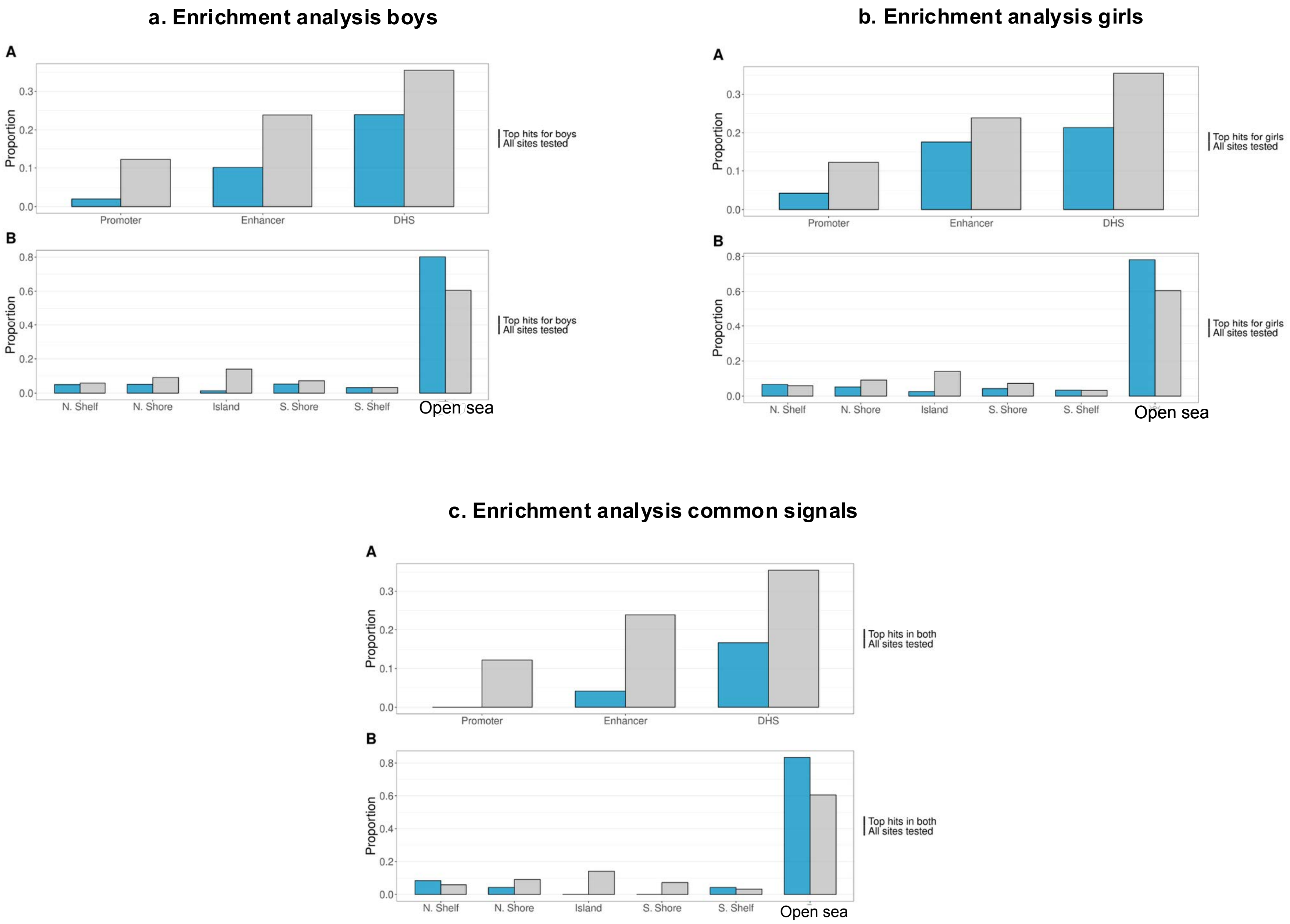

**Supplementary Fig 2.** Graphical view of pathways, biological processes and other molecular functions enriched among CpGs identified in the EWAS of ePTB in boys (n=318 CpGs at $p < 9.4 \times 10^{-8}$). Analysis performed using *gProfiler*. Presenting top terms with the lowest adjusted-$p$ for enrichment across multiple functional databases.

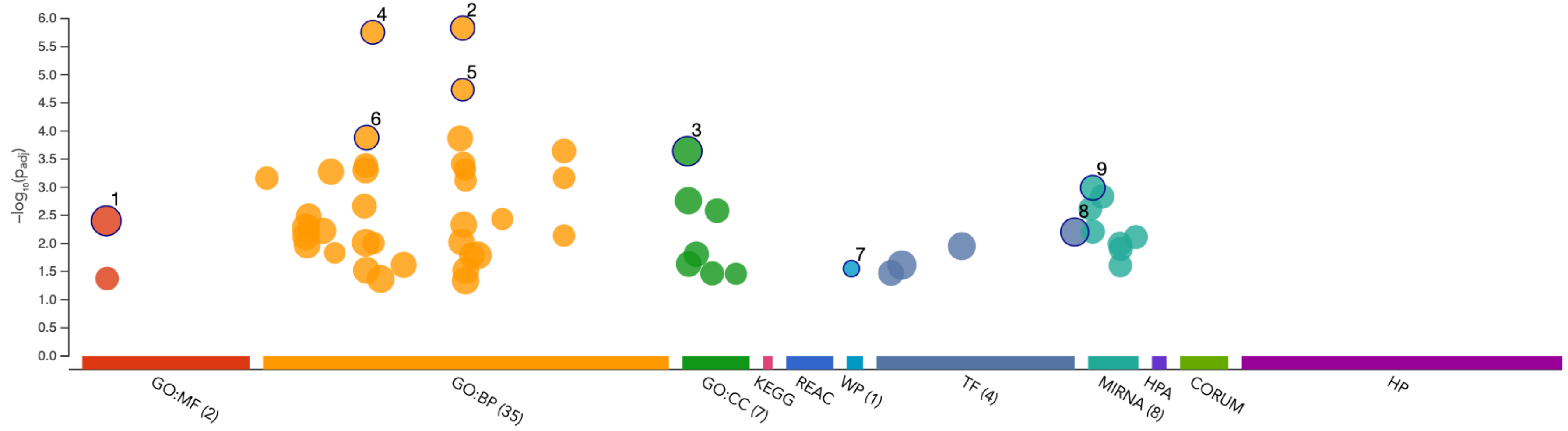


| ID | Source | Term ID | Term Name | $p_{adj}$ (query_1) |
|---|---|---|---|---|
| 1 | GO:MF | GO:0005515 | protein binding | $3.958 \times 10^{-3}$ |
| 2 | GO:BP | GO:0048666 | neuron development | $1.475 \times 10^{-6}$ |
| 3 | GO:CC | GO:0005737 | cytoplasm | $2.285 \times 10^{-4}$ |
| 4 | GO:BP | GO:0031175 | neuron projection development | $1.758 \times 10^{-6}$ |
| 5 | GO:BP | GO:0048667 | cell morphogenesis involved in neuron differentiation | $1.842 \times 10^{-5}$ |
| 6 | GO:BP | GO:0030182 | neuron differentiation | $1.317 \times 10^{-4}$ |
| 7 | WP | WP:WP3850 | Factors and pathways affecting insulin like growth factor IGF1 Akt signaling | $2.798 \times 10^{-2}$ |
| 8 | TF | TF:M12722_1 | Factor: ZXDL; motif: GSGSCNNGGGMRGCNCCGGGS; match class: 1 | $6.250 \times 10^{-3}$ |
| 9 | MIRNA | MIRNA:hsa-mir-543 | hsa-mir-543 | $1.022 \times 10^{-3}$ |

**Supplementary Fig 3.** Graphical view of pathways, biological processes and other molecular functions enriched among CpGs identified in the EWAS of ePTB in girls (n=542 CpGs at $p < 9.4 \times 10^{-8}$). Analysis performed using *gProfiler*. Presenting top terms with the lowest adjusted-*p* for enrichment across multiple functional databases.

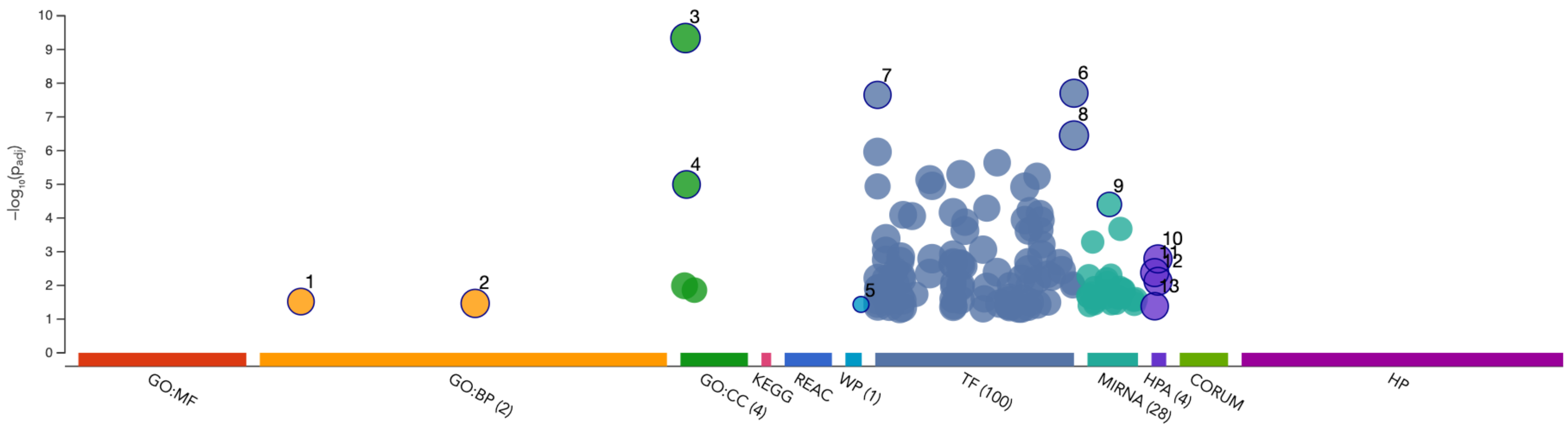


| ID | Source | Term ID | Term Name | $p_{adj}$ (query_1) |
|---|---|---|---|---|
| 1 | GO:BP | GO:0006996 | organelle organization | $3.018 \times 10^{-2}$ |
| 2 | GO:BP | GO:0051716 | cellular response to stimulus | $3.425 \times 10^{-2}$ |
| 3 | GO:CC | GO:0005737 | cytoplasm | $4.589 \times 10^{-10}$ |
| 4 | GO:CC | GO:0005829 | cytosol | $1.012 \times 10^{-5}$ |
| 5 | WP | WP:WP5599 | KCNQ2 and KCNQ3 related epilepsy | $3.671 \times 10^{-2}$ |
| 6 | TF | TF:M12722_1 | Factor: ZXDL; motif: GSGSCNNGGGMRGCNCCGGGS; match class: 1 | $1.988 \times 10^{-8}$ |
| 7 | TF | TF:M01857_1 | Factor: AP-2alpha; motif: NGCCYSNNGSN; match class: 1 | $2.226 \times 10^{-8}$ |
| 8 | TF | TF:M12722 | Factor: ZXDL; motif: GSGSCNNGGGMRGCNCCGGGS | $3.548 \times 10^{-7}$ |
| 9 | MIRNA | MIRNA:hsa-mir-326 | hsa-mir-326 | $3.959 \times 10^{-5}$ |
| 10 | HPA | HPA:0250000 | Hippocampus | $1.638 \times 10^{-3}$ |
| 11 | HPA | HPA:0100131 | Cerebral cortex; neuronal cells[≥Low] | $4.173 \times 10^{-3}$ |
| 12 | HPA | HPA:0250131 | Hippocampus; neuronal cells[≥Low] | $7.554 \times 10^{-3}$ |
| 13 | HPA | HPA:0100132 | Cerebral cortex; neuronal cells[≥Medium] | $4.155 \times 10^{-2}$ |
