## Supplemental Tables for "Sex specificity in DNA Methylation Profiles of Neonates Following Early Preterm Birth"

**Supplementary Table 1**

Comparison of baseline characteristics between the subset of 273 samples included in the EWAS of early preterm birth (ePTB), and the full eligible sample from the case-control study. Eligible participants were those who met the inclusion criteria for the study, and additionally had genomic DNA extracted from saliva. Eligible participants were those who met the inclusion criteria for the study, and additionally had genomic DNA extracted from saliva. Eligible participants were those who met the inclusion criteria for the study, and additionally had genomic DNA extracted from saliva.

| | **Eligible sample (N=1,009)** | | **Samples in EWAS** | |
|---|---|---|---|---|
| | **ePTB (N=468)** | **Control (N=541)** | **ePTB (n=139)** | **Controls (n=134)** |
| Maternal age (years) | 28.6 (6.84) | 27.1 (6.68) | 29.1 (6.97) | 27.0 (6.30) |
| Gestational age (weeks) | 333 (71.2%) | 357 (66.0%) | 29.7 (2.91) | 38.9 (1.09) |
| Gravidity | | | | |
| Multiparous | 333 (71.2%) | 357 (66.0%) | 102 (73.4%) | 88 (65.7%) |
| Primiparous | 132 (28.2%) | 179 (33.1%) | 37 (26.6%) | 46 (34.3%) |
| Pre-pregnancy BMI (kg/m2) | 25.2 (5.15) | 25.2 (4.52) | 25.7 (5.06) | 25.5 (4.88) |
| Employed in pregnancy | | | | |
| No | 209 (44.7%) | 244 (45.1%) | 61 (43.9%) | 60 (44.8%) |
| Yes | 256 (54.7%) | 293 (54.2%) | 78 (56.1%) | 74 (55.2%) |
| Marital status | | | | |
| Married/Living with partner | 75 (16.0%) | 86 (15.9%) | 24 (17.3%) | 28 (20.9%) |
| Not married/Living alone | 391 (83.5%) | 452 (83.5%) | 115 (82.7%) | 106 (79.1%) |
| Perception of health | | | | |
| Fair to Poor | 297 (63.5%) | 242 (44.7%) | 86 (61.9%) | 70 (52.2%) |
| Good to Excellent | 170 (36.3%) | 294 (54.3%) | 53 (38.1%) | 63 (47.0%) |
| Planned pregnancy | | | | |
| No | 321 (68.6%) | 383 (70.8%) | 88 (63.3%) | 92 (68.7%) |
| Yes | 143 (30.6%) | 152 (28.1%) | 51 (36.7%) | 40 (29.9%) |
| Smoked in pregnancy | | | | |
| No | 462 (98.7%) | 533 (98.5%) | 137 (98.6%) | 132 (98.5%) |
| Yes | 6 (1.3%) | 7 (1.3%) | 2 (1.4%) | 2 (1.5%) |
| Mode of delivery | | | | |
| Natural | 172 (36.8%) | 273 (50.5%) | 49 (35.3%) | 55 (41.0%) |
| C-section | 294 (62.8%) | 266 (49.2%) | 90 (64.7%) | 79 (59.0%) |
| Premature rupture of membranes (PPROM) | | | | |
| No | 210 (44.9%) | 468 (86.5%) | 61 (43.9%) | 121 (90.3%) |
| Yes | 252 (53.8%) | 64 (11.8%) | 74 (53.2%) | 12 (9.0%) |
| Newborn sex | | | | |
| Girl | 197 (42.1%) | 257 (47.5%) | 55 (39.6%) | 79 (59.0%) |
| Boy | 265 (56.6%) | 277 (51.2%) | 84 (60.4%) | 55 (41.0%) |
| Birth weight (kg) | 1.63 (0.573) | 3.39 (0.553) | 1.56 (0.547) | 3.41 (0.561) |
| Nursery stay | | | | |
| NICU | 360 (76.9%) | 17 (3.1%) | 117 (84.2%) | 9 (6.7%) |
| regular | 95 (20.3%) | 521 (96.3%) | 18 (12.9%) | 125 (93.3%) |
| Discharge status | | | | |
| Alive | 359 (76.7%) | 526 (97.2%) | 92 (66.2%) | 128 (95.5%) |
| Dead | 45 (9.6%) | 3 (0.6%) | 18 (12.9%) | 2 (1.5%) |
| APGAR score (5 min) | 7.98 (1.68) | 8.94 (0.385) | 7.81 (1.76) | 8.93 (0.485) |

P calculated using t-test for continuous variables (Mann-Whitney-Wilcoxon test if unequal variance); Chi-squared test for binary variables; Cochran–Armitage test for ordinal variables

**Supplementary Table 2**

Associations identified with epigenome-significance in the EWAS of early preterm birth (ePTB) among boys (N=139). Showing effect estimates observed for same signals in the girls' analysis for comparison. Associations were deemed significant at $p < 9.4 \times 10^{-8}$.

| | | | | | | Boys analysis | | Girls analysis | |
|---|---|---|---|---|---|---|---|---|---|
| **CpG** | **CHR** | **POS** | **Near Gene** | **Gene context** | **CG island context** | **Effect size (95% CI)** | ***P*** | **Effect size (95% CI)** | ***P*** |
| cg00861010 | 1 | 41500442 | *SLFNL1-AS1* | ncRNA_intronic | S_Shelf | 0.13(0.11,0.15) | 8.7E-29 | 0.05(0.03,0.06) | 2.4E-12 |
| cg02603733 | 1 | 151474886 | *POGZ;CGN* | intergenic | S_Shore | -0.11(-0.14,-0.09) | 1.4E-21 | -0.07(-0.08,-0.06) | 4.8E-23 |
| cg03515773 | 1 | 206796226 | *EIF2D;DYRK3* | intergenic | OpenSea | -0.18(-0.22,-0.15) | 4.3E-21 | -0.03(-0.04,-0.02) | 2.4E-09 |
| cg01886180 | 1 | 207867500 | *CR1L* | intronic | N_Shore | 0.15(0.12,0.19) | 1.4E-20 | -0.05(-0.06,-0.04) | 2.3E-20 |
| cg01206832 | 1 | 178653198 | *MIR4424;RALGPS2* | intergenic | OpenSea | 0.11(0.08,0.13) | 5.2E-19 | 0.09(0.07,0.11) | 4.7E-18 |
| cg16041784 | 1 | 204286824 | *PLEKHA6* | intronic | OpenSea | 0.11(0.08,0.13) | 9.2E-19 | -0.06(-0.08,-0.05) | 3.2E-13 |
| cg01118127 | 1 | 99647850 | *LOC100129620;PLPPR4* | intergenic | OpenSea | 0.11(0.09,0.14) | 8.5E-17 | -0.06(-0.08,-0.04) | 5.9E-11 |
| cg09896211 | 1 | 107689799 | *NTNG1* | intronic | OpenSea | 0.15(0.11,0.18) | 3.8E-16 | 0.04(0.03,0.05) | 1.5E-12 |
| cg15060258 | 1 | 167663761 | *RCSD1* | intronic | OpenSea | 0.13(0.1,0.17) | 4.0E-16 | 0.08(0.06,0.1) | 1.1E-21 |
| cg01652584 | 1 | 178919806 | *RALGPS2;FAM20B* | intergenic | OpenSea | -0.11(-0.14,-0.08) | 5.0E-16 | -0.04(-0.05,-0.02) | 4.1E-09 |
| cg24541835 | 1 | 12591527 | *VPS13D;DHRS3* | intergenic | OpenSea | 0.14(0.11,0.18) | 5.9E-16 | -0.05(-0.07,-0.03) | 1.1E-09 |
| cg19250709 | 1 | 152811013 | *LCE1A;LCE6A* | intergenic | OpenSea | 0.12(0.09,0.14) | 1.1E-15 | -0.04(-0.06,-0.03) | 4.1E-12 |
| cg10424513 | 1 | 99657534 | *LOC100129620;PLPPR4* | intergenic | OpenSea | 0.14(0.11,0.18) | 5.2E-15 | 0.07(0.05,0.09) | 2.3E-18 |
| cg13907014 | 1 | 87715234 | *LINC02801* | intronic | OpenSea | -0.11(-0.14,-0.08) | 9.6E-14 | 0.02(0.01,0.03) | 1.1E-04 |
| cg08895932 | 1 | 152806104 | *LCE1A;LCE6A* | intergenic | OpenSea | 0.11(0.08,0.15) | 1.7E-13 | -0.09(-0.13,-0.05) | 3.0E-06 |
| cg24933740 | 1 | 228143715 | *WNT9A;LOC107985355* | intergenic | S_Shelf | 0.11(0.08,0.15) | 3.6E-13 | 0.05(0.04,0.07) | 1.7E-11 |
| cg26897909 | 1 | 206455850 | *CTSE;SRGAP2* | intergenic | OpenSea | -0.1(-0.13,-0.08) | 5.2E-13 | -0.04(-0.05,-0.03) | 4.1E-16 |
| cg24793045 | 1 | 162690226 | *DDR2* | intronic | OpenSea | 0.1(0.07,0.13) | 1.6E-12 | 0.06(0.04,0.08) | 4.4E-12 |
| cg06713314 | 1 | 201264212 | *PKP1* | intronic | OpenSea | 0.1(0.07,0.13) | 2.0E-12 | -0.04(-0.06,-0.03) | 6.7E-10 |
| cg27438425 | 1 | 41486550 | *SLFNL1-AS1* | ncRNA_intronic | OpenSea | -0.1(-0.13,-0.07) | 7.7E-12 | -0.04(-0.05,-0.02) | 5.7E-07 |
| cg21278764 | 1 | 38016093 | *SNIP1* | intronic | S_Shelf | 0.1(0.07,0.13) | 3.2E-11 | -0.06(-0.08,-0.04) | 6.9E-10 |
| cg23656872 | 1 | 167664347 | *RCSD1* | intronic | OpenSea | 0.13(0.09,0.17) | 4.8E-11 | 0.06(0.04,0.07) | 1.5E-21 |
| cg02036963 | 1 | 225817687 | *ENAH* | intronic | OpenSea | 0.1(0.07,0.13) | 7.0E-11 | 0.07(0.05,0.09) | 7.6E-14 |
| cg10307152 | 1 | 12580650 | *VPS13D;DHRS3* | intergenic | OpenSea | -0.15(-0.19,-0.1) | 1.3E-10 | -0.02(-0.03,-0.01) | 1.2E-07 |
| cg16295156 | 1 | 9406269 | *SPSB1* | intronic | OpenSea | 0.11(0.08,0.14) | 1.7E-10 | 0.03(0.02,0.04) | 2.8E-10 |
| cg17083209 | 1 | 81808889 | *ADGRL2* | intronic | OpenSea | -0.13(-0.17,-0.09) | 5.7E-10 | 0.06(0.05,0.08) | 7.4E-12 |

| | | | | | | | | | |
|---|---|---|---|---|---|---|---|---|---|
| cg01617632 | 1 | 174116288 | *RABGAP1L-DT* | ncRNA_intronic | OpenSea | -0.11(-0.15,-0.08) | 8.9E-10 | -0.04(-0.05,-0.03) | 4.5E-18 |
| cg02605091 | 1 | 7497450 | *CAMTA1* | intronic | OpenSea | 0.1(0.07,0.14) | 1.2E-09 | -0.08(-0.11,-0.06) | 1.3E-10 |
| cg01699305 | 1 | 183882582 | *RGL1* | intronic | OpenSea | -0.12(-0.16,-0.08) | 1.3E-09 | -0.05(-0.06,-0.04) | 3.4E-16 |
| cg09364316 | 1 | 9405637 | *SPSB1* | intronic | OpenSea | 0.12(0.08,0.16) | 1.9E-09 | 0.05(0.04,0.07) | 1.4E-16 |
| cg23017345 | 1 | 241589805 | *RGS7;FH* | intergenic | OpenSea | -0.11(-0.15,-0.07) | 3.5E-09 | -0.06(-0.07,-0.04) | 3.9E-09 |
| cg14980042 | 1 | 230154208 | *LINC01736;GALNT2* | intergenic | OpenSea | -0.12(-0.16,-0.08) | 4.9E-09 | -0.09(-0.12,-0.07) | 1.2E-12 |
| cg13495130 | 1 | 230153971 | *LINC01736;GALNT2* | intergenic | OpenSea | -0.11(-0.14,-0.07) | 6.0E-09 | -0.04(-0.06,-0.03) | 6.1E-10 |
| cg00815112 | 1 | 56867865 | *LINC01755;LINC01767* | intergenic | OpenSea | -0.1(-0.14,-0.07) | 2.3E-08 | -0.01(-0.02,0) | 1.7E-03 |
| cg04321197 | 1 | 206772311 | *EIF2D* | intronic | OpenSea | -0.1(-0.14,-0.07) | 5.1E-08 | 0.05(0.04,0.06) | 3.1E-16 |
| cg21159128 | 1 | 54228260 | *GLIS1;NDC1* | intergenic | OpenSea | -0.1(-0.14,-0.06) | 6.0E-08 | -0.05(-0.07,-0.04) | 2.7E-12 |
| cg01000824 | 1 | 83818553 | *LINC01362;LINC01712* | intergenic | OpenSea | -0.11(-0.14,-0.07) | 6.1E-08 | 0.04(0.03,0.06) | 7.9E-12 |
| cg00314861 | 1 | 179838533 | *TOR1AIP2* | intronic | OpenSea | -0.11(-0.15,-0.07) | 7.0E-08 | -0.02(-0.03,-0.01) | 1.4E-06 |
| cg04308291 | 2 | 234189621 | *ATG16L1* | intronic | OpenSea | 0.14(0.12,0.17) | 2.5E-26 | -0.04(-0.06,-0.03) | 2.5E-12 |
| cg02458632 | 2 | 62450946 | *B3GNT2* | UTR3 | OpenSea | 0.16(0.13,0.19) | 6.4E-26 | -0.05(-0.07,-0.04) | 1.2E-15 |
| cg27293447 | 2 | 67130663 | *LINC01799* | upstream | OpenSea | -0.12(-0.15,-0.1) | 1.7E-22 | -0.04(-0.05,-0.03) | 1.6E-09 |
| cg11712893 | 2 | 121156075 | *INHBB;LINC01101* | intergenic | OpenSea | 0.1(0.08,0.12) | 2.0E-22 | -0.03(-0.04,-0.03) | 1.3E-20 |
| cg21361437 | 2 | 106561756 | *NCK2;ECRG4* | intergenic | OpenSea | 0.1(0.08,0.12) | 6.0E-21 | 0.09(0.07,0.11) | 3.6E-19 |
| cg04813347 | 2 | 218072455 | *LINC01921;DIRC3-AS1* | intergenic | OpenSea | -0.12(-0.15,-0.1) | 2.5E-20 | -0.06(-0.07,-0.05) | 6.7E-17 |
| cg04404503 | 2 | 240403358 | *HDAC4-AS1;LOC150935* | intergenic | OpenSea | -0.12(-0.14,-0.09) | 1.2E-17 | -0.06(-0.08,-0.04) | 2.1E-08 |
| cg04470497 | 2 | 1285487 | *SNTG2* | intronic | S_Shelf | -0.13(-0.17,-0.1) | 3.7E-17 | -0.03(-0.04,-0.01) | 3.2E-04 |
| cg23173746 | 2 | 102186692 | *RFX8;MAP4K4* | intergenic | N_Shore | 0.11(0.08,0.14) | 3.7E-16 | -0.05(-0.07,-0.04) | 4.9E-15 |
| cg24867727 | 2 | 64994448 | *SERTAD2;LINC01800* | intergenic | OpenSea | 0.15(0.11,0.18) | 1.0E-14 | -0.04(-0.05,-0.03) | 1.1E-11 |
| cg04342509 | 2 | 177192977 | *MTX2* | intronic | OpenSea | 0.16(0.12,0.2) | 2.0E-14 | 0.09(0.06,0.11) | 1.0E-12 |
| cg03826123 | 2 | 174603308 | *CDCA7;SP3* | intergenic | OpenSea | 0.11(0.08,0.14) | 2.1E-14 | 0.06(0.04,0.08) | 7.0E-12 |
| cg04281920 | 2 | 232316147 | *B3GNT7;NCL* | intergenic | OpenSea | 0.12(0.09,0.15) | 1.6E-13 | -0.06(-0.07,-0.04) | 1.9E-23 |
| cg11244641 | 2 | 240403608 | *HDAC4-AS1;LOC150935* | intergenic | OpenSea | -0.12(-0.16,-0.09) | 1.2E-12 | -0.02(-0.02,-0.01) | 1.5E-06 |
| cg02069095 | 2 | 133262469 | *GPR39* | intronic | N_Shelf | -0.12(-0.16,-0.09) | 1.9E-12 | -0.14(-0.18,-0.1) | 4.9E-11 |
| cg08406348 | 2 | 75473440 | *TACR1;EVA1A* | intergenic | OpenSea | -0.1(-0.13,-0.07) | 2.0E-11 | -0.03(-0.05,-0.02) | 1.2E-05 |
| cg04800041 | 2 | 186481111 | *LOC105373782;FSIP2-AS2* | intergenic | OpenSea | 0.12(0.09,0.16) | 5.8E-11 | 0.05(0.03,0.07) | 5.2E-10 |
| cg13908635 | 2 | 218815041 | *TNS1;LOC105373878* | intergenic | OpenSea | 0.1(0.07,0.13) | 3.0E-10 | -0.04(-0.05,-0.02) | 2.7E-07 |
| cg07141356 | 2 | 43171892 | *HAAO;LINC01819* | intergenic | S_Shore | -0.12(-0.16,-0.08) | 5.8E-10 | -0.05(-0.06,-0.03) | 2.5E-13 |
| cg21503763 | 2 | 62444115 | *B3GNT2* | intronic | OpenSea | 0.13(0.09,0.17) | 9.2E-10 | -0.15(-0.19,-0.11) | 7.4E-15 |
| cg19374905 | 2 | 217978471 | *LINC01921;DIRC3-AS1* | intergenic | N_Shore | -0.1(-0.13,-0.07) | 1.6E-09 | 0.02(0.01,0.03) | 1.7E-05 |
| cg12862395 | 2 | 152537517 | *NEB* | intronic | OpenSea | -0.13(-0.17,-0.08) | 5.7E-09 | -0.03(-0.04,-0.02) | 2.7E-05 |

| | | | | | | | | | |
|---|---|---|---|---|---|---|---|---|---|
| cg03825906 | 2 | 174573807 | *CDCA7;SP3* | intergenic | OpenSea | 0.14(0.09,0.19) | 9.9E-09 | -0.03(-0.04,-0.02) | 1.5E-07 |
| cg02559003 | 2 | 25205114 | *DNAJC27-AS1* | ncRNA_intronic | S_Shore | -0.1(-0.14,-0.07) | 1.3E-08 | -0.04(-0.05,-0.02) | 4.4E-07 |
| cg04040637 | 2 | 241885697 | *CROCC2* | intronic | S_Shelf | -0.11(-0.15,-0.07) | 2.0E-08 | -0.06(-0.07,-0.04) | 5.1E-14 |
| cg10347254 | 2 | 112680968 | *MERTK* | intronic | OpenSea | 0.11(0.07,0.15) | 6.6E-08 | -0.03(-0.04,-0.02) | 6.0E-12 |
| cg05103200 | 3 | 72836974 | *SHQ1* | intronic | OpenSea | 0.1(0.08,0.12) | 1.9E-22 | 0.04(0.03,0.05) | 1.3E-17 |
| cg05513060 | 3 | 129869013 | *LINC02021;COL6A4P2* | intergenic | OpenSea | -0.11(-0.13,-0.09) | 6.9E-20 | -0.06(-0.07,-0.04) | 8.1E-11 |
| cg23982461 | 3 | 159237238 | *IQCJ-SCHIP1;SCHIP1* | intronic | OpenSea | -0.13(-0.15,-0.1) | 7.0E-20 | -0.06(-0.08,-0.04) | 6.1E-12 |
| cg21902516 | 3 | 13767651 | *LINC00620* | ncRNA_intronic | OpenSea | -0.1(-0.13,-0.08) | 3.8E-19 | -0.12(-0.15,-0.09) | 4.9E-17 |
| cg23173566 | 3 | 30007623 | *RBMS3* | intronic | OpenSea | -0.16(-0.2,-0.12) | 5.3E-17 | 0.05(0.03,0.07) | 1.6E-05 |
| cg08248287 | 3 | 171307052 | *TNIK;PLD1* | intergenic | OpenSea | 0.12(0.09,0.15) | 5.4E-13 | -0.04(-0.05,-0.03) | 3.2E-14 |
| cg22866411 | 3 | 54898860 | *CACNA2D3* | intronic | OpenSea | -0.13(-0.16,-0.09) | 9.8E-13 | 0.07(0.05,0.09) | 5.1E-11 |
| cg22797643 | 3 | 78496655 | *LINC02077;ROBO1* | intergenic | OpenSea | -0.11(-0.14,-0.08) | 1.9E-12 | 0.07(0.05,0.09) | 1.4E-12 |
| cg07649206 | 3 | 171024305 | *TNIK* | intronic | N_Shelf | -0.1(-0.13,-0.07) | 7.5E-12 | -0.04(-0.05,-0.03) | 8.6E-13 |
| cg00191665 | 3 | 128759459 | *EFCC1* | UTR3 | OpenSea | -0.14(-0.18,-0.1) | 6.7E-11 | -0.07(-0.09,-0.05) | 5.0E-10 |
| cg15649559 | 3 | 40766191 | *ZNF621;CTNNB1* | intergenic | OpenSea | 0.15(0.11,0.2) | 3.8E-10 | -0.12(-0.15,-0.09) | 1.1E-17 |
| cg01049503 | 3 | 123993353 | *KALRN* | intronic | S_Shore | -0.11(-0.15,-0.08) | 4.3E-10 | 0.04(0.02,0.06) | 4.3E-06 |
| cg25839267 | 3 | 77042494 | *ZNF717;ROBO2* | intergenic | S_Shelf | -0.11(-0.15,-0.07) | 1.8E-09 | -0.05(-0.07,-0.03) | 2.0E-05 |
| cg05783189 | 3 | 167840567 | *GOLIM4;EGFEM1P* | intergenic | OpenSea | 0.1(0.07,0.13) | 2.7E-09 | -0.03(-0.04,-0.02) | 1.5E-07 |
| cg27024488 | 3 | 49409077 | *RHOA* | intronic | N_Shelf | -0.11(-0.15,-0.07) | 2.6E-08 | 0.01(0.01,0.02) | 5.9E-07 |
| cg22792232 | 3 | 55364520 | *LINC02017;WNT5A* | intergenic | OpenSea | -0.11(-0.15,-0.07) | 4.7E-08 | 0.04(0.02,0.06) | 1.4E-04 |
| cg08583624 | 4 | 97366529 | *LINC02267* | ncRNA_intronic | OpenSea | -0.11(-0.13,-0.08) | 1.1E-16 | 0.05(0.02,0.08) | 1.3E-03 |
| cg21956384 | 4 | 41189840 | *APBB2* | intronic | OpenSea | -0.11(-0.14,-0.08) | 2.0E-14 | -0.06(-0.08,-0.05) | 6.5E-15 |
| cg06380971 | 4 | 26205012 | *SMIM20;RBPJ* | intergenic | OpenSea | 0.1(0.08,0.13) | 2.7E-13 | -0.05(-0.06,-0.04) | 1.5E-14 |
| cg23543824 | 4 | 69518966 | *UGT2B15* | intronic | OpenSea | -0.16(-0.21,-0.11) | 1.8E-11 | -0.02(-0.03,0) | 7.1E-03 |
| cg06413218 | 4 | 31089889 | *PCDH7* | intronic | OpenSea | -0.11(-0.15,-0.08) | 9.0E-11 | -0.07(-0.09,-0.04) | 1.8E-09 |
| cg14022913 | 4 | 177308497 | *SPCS3;VEGFC* | intergenic | N_Shore | 0.13(0.09,0.18) | 2.0E-10 | -0.06(-0.07,-0.04) | 1.6E-11 |
| cg14538218 | 4 | 123554544 | *IL21-AS1* | ncRNA_intronic | OpenSea | -0.12(-0.15,-0.08) | 3.9E-10 | -0.08(-0.1,-0.06) | 1.8E-13 |
| cg06848823 | 4 | 94568077 | *GRID2* | intronic | OpenSea | 0.15(0.11,0.2) | 4.9E-10 | 0.06(0.03,0.08) | 4.2E-07 |
| cg02726826 | 4 | 177109306 | *SPATA4* | exonic | OpenSea | -0.1(-0.14,-0.07) | 5.3E-10 | -0.02(-0.04,0) | 2.4E-02 |
| cg27060294 | 4 | 16837781 | *LDB2* | intronic | OpenSea | 0.1(0.07,0.14) | 7.6E-10 | -0.04(-0.06,-0.03) | 2.7E-06 |
| cg25114899 | 4 | 67747105 | *MIR1269A;LOC101927237* | intergenic | OpenSea | -0.14(-0.19,-0.1) | 1.3E-09 | -0.03(-0.04,-0.02) | 7.8E-10 |
| cg04111054 | 4 | 130479179 | *C4orf33;LINC02466* | intergenic | OpenSea | -0.13(-0.17,-0.09) | 2.5E-09 | -0.02(-0.02,-0.01) | 2.1E-04 |
| cg14900031 | 4 | 6693650 | *LINC002481;S100P* | intergenic | OpenSea | -0.1(-0.14,-0.06) | 4.0E-08 | -0.05(-0.07,-0.03) | 2.2E-05 |
| cg11343820 | 5 | 174617516 | *LINC01951;DRD1* | intergenic | OpenSea | -0.17(-0.2,-0.14) | 3.6E-27 | 0.08(0.06,0.1) | 3.9E-12 |

| | | | | | | | | | |
|---|---|---|---|---|---|---|---|---|---|
| cg14871997 | 5 | 174618039 | *LINC01951;DRD1* | intergenic | OpenSea | -0.11(-0.13,-0.09) | 5.0E-26 | -0.05(-0.06,-0.04) | 1.5E-16 |
| cg06325472 | 5 | 174618083 | *LINC01951;DRD1* | intergenic | OpenSea | -0.12(-0.15,-0.1) | 7.7E-22 | -0.1(-0.12,-0.08) | 1.8E-17 |
| cg14880229 | 5 | 95882301 | *LOC101929710* | ncRNA_intronic | OpenSea | 0.1(0.08,0.13) | 3.1E-18 | -0.02(-0.03,-0.01) | 2.3E-07 |
| cg03142049 | 5 | 140219836 | *PCDHA1;PCDHA2* | intronic | OpenSea | 0.11(0.09,0.14) | 2.4E-17 | -0.06(-0.07,-0.05) | 5.4E-25 |
| cg09364402 | 5 | 139912477 | *NKHD1;ANKHD1-EIF4EBF* | intronic | OpenSea | 0.1(0.08,0.13) | 1.7E-15 | -0.07(-0.1,-0.05) | 4.3E-11 |
| cg00251358 | 5 | 82635682 | *XRCC4* | intronic | OpenSea | -0.13(-0.16,-0.09) | 1.1E-13 | -0.04(-0.05,-0.03) | 1.8E-12 |
| cg09999715 | 5 | 53948938 | *SNX18;LOC102467080* | intergenic | OpenSea | -0.1(-0.13,-0.07) | 2.1E-13 | -0.06(-0.08,-0.04) | 6.7E-08 |
| cg08431289 | 5 | 12636543 | *LINC01194* | ncRNA_intronic | OpenSea | -0.2(-0.26,-0.15) | 1.3E-12 | -0.14(-0.18,-0.11) | 3.0E-15 |
| cg08667430 | 5 | 146047637 | *PPP2R2B* | intronic | OpenSea | 0.1(0.07,0.13) | 1.7E-11 | 0.07(0.05,0.08) | 1.4E-13 |
| cg22026677 | 5 | 172861442 | *MIR8056;LOC285593* | intergenic | OpenSea | 0.1(0.07,0.13) | 2.8E-11 | -0.11(-0.15,-0.08) | 1.0E-11 |
| cg06013117 | 5 | 174726315 | *LINC01951;DRD1* | intergenic | S_Shore | -0.15(-0.2,-0.11) | 3.7E-11 | -0.09(-0.11,-0.07) | 5.4E-15 |
| cg03242909 | 5 | 174617887 | *LINC01951;DRD1* | intergenic | OpenSea | -0.12(-0.15,-0.08) | 3.5E-10 | -0.06(-0.08,-0.04) | 8.4E-10 |
| cg20324727 | 5 | 126600144 | *C5orf63;MEGF10* | intergenic | N_Shore | 0.13(0.09,0.17) | 1.3E-09 | 0(0,0) | 7.4E-04 |
| cg26873042 | 5 | 68067510 | *PIK3R1;LINC02198* | intergenic | OpenSea | -0.1(-0.14,-0.07) | 2.6E-09 | 0.05(0.04,0.06) | 8.5E-15 |
| cg08315524 | 5 | 58246631 | *RAB3C;PDE4D* | intergenic | OpenSea | -0.11(-0.15,-0.08) | 7.3E-09 | -0.02(-0.03,-0.01) | 1.2E-03 |
| cg16283919 | 5 | 75044668 | *LOC441087;SV2C* | intergenic | OpenSea | 0.12(0.07,0.16) | 6.1E-08 | 0.03(0.02,0.04) | 1.1E-08 |
| cg10340304 | 6 | 158738149 | *TULP4* | intronic | OpenSea | 0.1(0.08,0.13) | 9.0E-20 | -0.05(-0.06,-0.04) | 7.8E-14 |
| cg23873636 | 6 | 47198171 | *ADGRF1;TNFRSF21* | intergenic | OpenSea | 0.12(0.09,0.15) | 1.4E-15 | -0.08(-0.1,-0.05) | 4.3E-10 |
| cg09920931 | 6 | 106732794 | *ATG5* | intronic | OpenSea | 0.11(0.08,0.14) | 1.4E-15 | -0.04(-0.05,-0.03) | 6.6E-16 |
| cg05906420 | 6 | 47308295 | *TNFRSF21;CD2AP* | intergenic | N_Shore | 0.11(0.09,0.14) | 2.1E-15 | -0.03(-0.04,-0.02) | 6.3E-06 |
| cg07164639 | 6 | 110415755 | *GPR6;WASF1* | intergenic | OpenSea | 0.1(0.08,0.13) | 5.2E-15 | 0.08(0.07,0.1) | 1.6E-22 |
| cg06593560 | 6 | 148819347 | *SASH1* | intronic | OpenSea | -0.12(-0.15,-0.09) | 5.4E-14 | -0.03(-0.04,-0.02) | 3.5E-12 |
| cg05382185 | 6 | 53361488 | *GCLC* | downstream | S_Shore | 0.1(0.08,0.13) | 2.1E-13 | -0.06(-0.08,-0.04) | 6.8E-12 |
| cg17470895 | 6 | 13984718 | *RNF182;CD83* | intergenic | OpenSea | -0.13(-0.17,-0.1) | 3.7E-13 | -0.02(-0.03,-0.01) | 4.0E-08 |
| cg06764229 | 6 | 14875854 | *LINC01108;JARID2* | intergenic | OpenSea | -0.11(-0.14,-0.08) | 1.4E-10 | 0.05(0.03,0.06) | 2.0E-10 |
| cg10121133 | 6 | 134331353 | *SLC2A12* | intronic | OpenSea | 0.1(0.07,0.13) | 1.4E-10 | -0.02(-0.03,-0.01) | 6.8E-04 |
| cg07317567 | 6 | 81767081 | *BCKDHB;TENT5A* | intergenic | OpenSea | -0.13(-0.18,-0.09) | 1.8E-10 | -0.05(-0.06,-0.04) | 3.4E-25 |
| cg23063382 | 6 | 121142324 | *MIR3144;TBC1D32* | intergenic | OpenSea | 0.12(0.08,0.16) | 2.6E-09 | -0.07(-0.1,-0.05) | 1.2E-07 |
| cg07478690 | 6 | 118351568 | *SLC35F1* | intronic | OpenSea | 0.12(0.08,0.16) | 3.8E-09 | -0.05(-0.06,-0.04) | 5.0E-16 |
| cg10166855 | 6 | 139518130 | *HECA;TXLNB* | intergenic | OpenSea | -0.1(-0.14,-0.07) | 7.6E-09 | -0.05(-0.06,-0.04) | 1.1E-16 |
| cg13918754 | 6 | 30071603 | *TRIM31* | intronic | Island;N_Shelf | -0.13(-0.18,-0.09) | 2.2E-08 | -0.03(-0.04,-0.02) | 2.5E-07 |
| cg06204531 | 7 | 150383888 | *GIMAP2* | intronic | OpenSea | -0.13(-0.16,-0.1) | 3.7E-19 | -0.02(-0.03,-0.01) | 1.1E-06 |
| cg09276050 | 7 | 102097941 | *ALKBH4* | exonic | OpenSea | -0.12(-0.15,-0.09) | 1.5E-15 | -0.1(-0.12,-0.08) | 4.0E-21 |
| cg02451350 | 7 | 116805334 | *ST7* | intronic | OpenSea | 0.1(0.08,0.13) | 4.0E-15 | -0.06(-0.09,-0.04) | 1.0E-07 |

| | | | | | | | | | |
|---|---|---|---|---|---|---|---|---|---|
| cg09031352 | 7 | 137324634 | *DGKI* | intronic | OpenSea | -0.17(-0.21,-0.12) | 5.9E-15 | -0.03(-0.05,-0.02) | 7.3E-07 |
| cg11550335 | 7 | 108007740 | *NRCAM* | intronic | OpenSea | -0.13(-0.17,-0.1) | 4.3E-14 | -0.04(-0.05,-0.03) | 9.7E-10 |
| cg10644659 | 7 | 102097612 | *ALKBH4* | UTR3 | OpenSea | -0.11(-0.14,-0.08) | 3.2E-13 | -0.04(-0.05,-0.02) | 3.6E-09 |
| cg16813564 | 7 | 94650348 | *PPP1R9A* | intronic | OpenSea | -0.14(-0.18,-0.1) | 4.9E-13 | -0.04(-0.06,-0.02) | 1.2E-06 |
| cg07205403 | 7 | 106860409 | *COG5* | intronic | OpenSea | -0.1(-0.13,-0.07) | 5.4E-12 | -0.03(-0.03,-0.02) | 1.8E-09 |
| cg13420277 | 7 | 28725358 | *CREB5* | intronic | OpenSea | 0.14(0.1,0.18) | 1.1E-11 | 0.07(0.05,0.1) | 1.5E-09 |
| cg11489261 | 7 | 101848193 | *CUX1* | intronic | OpenSea | 0.17(0.12,0.23) | 7.0E-11 | -0.02(-0.03,-0.01) | 2.6E-06 |
| cg20087585 | 7 | 150383885 | *GIMAP2* | intronic | OpenSea | -0.12(-0.16,-0.08) | 1.1E-10 | -0.03(-0.04,-0.02) | 2.2E-08 |
| cg11874977 | 7 | 149067707 | *LOC155060;ZNF777* | intergenic | S_Shore;N_Shelf | -0.12(-0.16,-0.09) | 1.6E-10 | 0.05(0.03,0.06) | 2.6E-10 |
| cg25832180 | 7 | 78279878 | *MAGI2* | intronic | OpenSea | -0.11(-0.14,-0.07) | 1.9E-10 | -0.08(-0.13,-0.04) | 1.2E-04 |
| cg08300570 | 7 | 92724140 | *LOC101927497;SAMD9* | intergenic | OpenSea | -0.11(-0.14,-0.07) | 4.5E-10 | -0.08(-0.1,-0.06) | 6.5E-13 |
| cg05916479 | 7 | 112199089 | *LSMEM1;LOC100996249* | intergenic | OpenSea | -0.11(-0.14,-0.07) | 9.7E-10 | -0.01(-0.02,-0.01) | 8.0E-06 |
| cg01942816 | 7 | 5495832 | *FBXL18* | intronic | S_Shore | 0.1(0.07,0.13) | 1.0E-09 | -0.04(-0.06,-0.03) | 1.8E-08 |
| cg19919244 | 7 | 69967992 | *AUTS2* | intronic | OpenSea | -0.12(-0.16,-0.08) | 7.2E-09 | -0.06(-0.08,-0.04) | 5.7E-07 |
| cg03078488 | 7 | 23467029 | *IGF2BP3* | intronic | N_Shore | -0.13(-0.18,-0.09) | 1.2E-08 | -0.05(-0.06,-0.04) | 7.9E-13 |
| cg19307584 | 7 | 29204763 | *CHN2;CPVL* | intronic | OpenSea | -0.11(-0.15,-0.07) | 2.0E-08 | -0.03(-0.05,-0.01) | 2.2E-04 |
| cg05527149 | 7 | 74463042 | *RCC1L* | intronic | OpenSea | -0.1(-0.14,-0.06) | 9.2E-08 | 0.07(0.05,0.09) | 7.1E-09 |
| cg13018311 | 8 | 116502783 | *TRPS1* | intronic | OpenSea | -0.12(-0.14,-0.09) | 3.6E-21 | -0.05(-0.07,-0.04) | 8.4E-14 |
| cg00120416 | 8 | 48872958 | *MCM4;PRKDC* | upstream | S_Shelf | 0.11(0.09,0.14) | 7.5E-18 | 0.08(0.06,0.1) | 6.9E-19 |
| cg03876023 | 8 | 103348635 | *UBR5* | intronic | OpenSea | -0.22(-0.28,-0.17) | 2.5E-16 | 0.15(0.11,0.18) | 7.1E-16 |
| cg12833469 | 8 | 8858071 | *MFHAS1;ERI1* | intergenic | OpenSea | 0.11(0.08,0.14) | 3.4E-16 | -0.04(-0.05,-0.03) | 9.7E-11 |
| cg09750115 | 8 | 33561278 | *DUSP26;LINC01288* | intergenic | OpenSea | 0.12(0.09,0.14) | 4.3E-16 | -0.06(-0.08,-0.05) | 6.7E-13 |
| cg13105150 | 8 | 127306245 | *LINC00861;LOC101927657* | intergenic | OpenSea | -0.1(-0.13,-0.08) | 7.4E-15 | 0.08(0.05,0.1) | 4.2E-09 |
| cg19238575 | 8 | 140309372 | *COL22A1;KCNK9* | intergenic | OpenSea | -0.15(-0.19,-0.11) | 8.7E-15 | -0.07(-0.09,-0.05) | 1.2E-09 |
| cg11889960 | 8 | 119102827 | *EXT1* | intronic | OpenSea | 0.11(0.08,0.13) | 3.9E-14 | -0.05(-0.07,-0.03) | 6.5E-10 |
| cg20599118 | 8 | 17648954 | *MTUS1* | intronic | OpenSea | 0.12(0.09,0.15) | 5.0E-13 | -0.05(-0.06,-0.04) | 9.9E-16 |
| cg12457255 | 8 | 40059189 | *TCIM;SIRLNT* | intergenic | OpenSea | -0.1(-0.13,-0.07) | 2.1E-12 | -0.03(-0.05,-0.02) | 1.8E-04 |
| cg04657644 | 8 | 40172988 | *TCIM;SIRLNT* | intergenic | OpenSea | -0.13(-0.17,-0.1) | 5.0E-12 | -0.05(-0.06,-0.04) | 2.7E-14 |
| cg25212737 | 8 | 48303255 | *SPIDR* | intronic | OpenSea | 0.13(0.09,0.17) | 1.6E-11 | 0.03(0.02,0.04) | 1.7E-10 |
| cg01413249 | 8 | 127893880 | *LRATD2;PCAT1* | intergenic | OpenSea | 0.13(0.09,0.17) | 6.6E-11 | 0.04(0.03,0.05) | 2.3E-10 |
| cg13029379 | 8 | 118032301 | *SLC30A8* | intronic | OpenSea | 0.1(0.07,0.13) | 1.0E-10 | 0.08(0.07,0.1) | 2.5E-20 |
| cg16871855 | 8 | 129958372 | *LINC00824;LINC00976* | intergenic | OpenSea | -0.11(-0.15,-0.08) | 1.6E-10 | 0.07(0.05,0.09) | 3.0E-10 |
| cg09063809 | 8 | 40099277 | *TCIM;SIRLNT* | intergenic | OpenSea | -0.16(-0.21,-0.11) | 1.9E-10 | -0.03(-0.04,-0.02) | 5.5E-07 |
| cg27085598 | 8 | 40098361 | *TCIM;SIRLNT* | intergenic | OpenSea | -0.12(-0.15,-0.08) | 2.1E-10 | -0.07(-0.09,-0.04) | 5.7E-07 |

| | | | | | | | | | |
|---|---|---|---|---|---|---|---|---|---|
| cg23538418 | 8 | 40099760 | *TCIM;SIRLNT* | intergenic | OpenSea | -0.14(-0.18,-0.09) | 9.6E-10 | 0.08(0.06,0.11) | 7.6E-10 |
| cg13632752 | 8 | 36984771 | *MIR1268A* | ncRNA_intronic | OpenSea | -0.1(-0.14,-0.07) | 1.4E-09 | -0.03(-0.04,-0.01) | 1.2E-05 |
| cg15671775 | 8 | 40100231 | *TCIM;SIRLNT* | intergenic | OpenSea | -0.11(-0.15,-0.08) | 1.6E-09 | -0.05(-0.07,-0.04) | 5.8E-11 |
| cg01451808 | 8 | 48764003 | *PRKDC* | intronic | OpenSea | -0.12(-0.15,-0.08) | 1.9E-09 | -0.05(-0.08,-0.03) | 7.1E-05 |
| cg10387952 | 8 | 19310842 | *CSGALNACT1* | intronic | N_Shelf | -0.1(-0.14,-0.07) | 1.1E-08 | -0.03(-0.04,-0.02) | 1.3E-07 |
| cg03728930 | 8 | 133298475 | *KCNQ3* | intronic | S_Shore | 0.17(0.11,0.23) | 4.7E-08 | -0.03(-0.04,-0.01) | 7.0E-06 |
| cg02046552 | 8 | 22056776 | *BMP1* | intronic | Island | -0.12(-0.17,-0.08) | 7.7E-08 | -0.03(-0.04,-0.02) | 9.8E-12 |
| cg13413384 | 9 | 134410385 | *UCK1;PRRT1B* | intergenic | S_Shelf;S_Shore | 0.13(0.1,0.15) | 9.3E-25 | -0.09(-0.11,-0.07) | 5.3E-17 |
| cg13933718 | 9 | 92725373 | *UNQ6494;LOC101927847* | intergenic | OpenSea | 0.11(0.08,0.13) | 1.3E-21 | 0.11(0.1,0.13) | 2.8E-37 |
| cg16109426 | 9 | 36566818 | *RNF38;MELK* | intergenic | OpenSea | 0.11(0.09,0.13) | 7.5E-21 | 0.12(0.1,0.13) | 1.0E-34 |
| cg13832290 | 9 | 35649909 | *SIT1* | exonic | S_Shelf | 0.1(0.08,0.13) | 2.0E-18 | -0.09(-0.11,-0.06) | 7.0E-13 |
| cg10355819 | 9 | 121464001 | *LINC02578* | ncRNA_intronic | OpenSea | 0.12(0.09,0.15) | 5.5E-18 | 0.12(0.1,0.14) | 4.0E-24 |
| cg14200004 | 9 | 121743668 | *LINC02578;BRINP1* | intergenic | OpenSea | 0.11(0.08,0.13) | 1.6E-17 | -0.05(-0.06,-0.04) | 5.9E-19 |
| cg08017357 | 9 | 71756971 | *TJP2* | intronic | OpenSea | -0.1(-0.13,-0.08) | 6.5E-17 | -0.07(-0.09,-0.05) | 4.8E-10 |
| cg08224960 | 9 | 95082818 | *NOL8* | intronic | N_Shelf | 0.12(0.09,0.14) | 4.4E-16 | -0.08(-0.1,-0.06) | 2.1E-14 |
| cg13961684 | 9 | 94786745 | *ROR2;SPTLC1* | intergenic | OpenSea | 0.12(0.09,0.15) | 1.5E-15 | -0.06(-0.08,-0.05) | 2.1E-16 |
| cg05891562 | 9 | 91192491 | *NXNL2;MIR4289* | intergenic | N_Shore | -0.15(-0.18,-0.11) | 2.7E-13 | 0.02(0.01,0.03) | 1.4E-03 |
| cg10364271 | 9 | 136352918 | *SLC2A6;MYMK* | intergenic | S_Shore | -0.1(-0.14,-0.07) | 1.6E-09 | -0.02(-0.03,-0.02) | 6.4E-08 |
| cg13630153 | 9 | 38043932 | *SHB* | intronic | OpenSea | 0.11(0.07,0.14) | 1.8E-08 | -0.04(-0.07,-0.02) | 2.1E-04 |
| cg09340066 | 9 | 128339743 | *MAPKAP1* | intronic | N_Shore | 0.1(0.06,0.14) | 3.3E-08 | 0.02(0.02,0.03) | 3.5E-08 |
| cg02872583 | 9 | 129339062 | *MVB12B;LMX1B* | intergenic | S_Shore | -0.1(-0.14,-0.07) | 3.6E-08 | -0.04(-0.06,-0.03) | 6.8E-12 |
| cg14887720 | 10 | 33118141 | *CCDC7* | intronic | OpenSea | 0.11(0.08,0.13) | 2.9E-23 | -0.06(-0.08,-0.05) | 4.5E-15 |
| cg18295744 | 10 | 79113169 | *KCNMA1-AS3* | ncRNA_intronic | OpenSea | 0.12(0.09,0.14) | 8.2E-20 | -0.11(-0.14,-0.08) | 4.7E-12 |
| cg20463533 | 10 | 132892351 | *TCERG1L* | intronic | OpenSea | 0.11(0.08,0.13) | 5.1E-16 | 0.08(0.06,0.1) | 2.4E-15 |
| cg25368317 | 10 | 99697490 | *CRTAC1* | intronic | OpenSea | 0.13(0.09,0.17) | 3.0E-12 | -0.09(-0.11,-0.07) | 6.7E-23 |
| cg08431336 | 10 | 110759156 | *LINC01435;XPNPEP1* | intergenic | OpenSea | 0.1(0.07,0.13) | 1.0E-11 | -0.02(-0.02,-0.01) | 3.3E-08 |
| cg19864972 | 10 | 20962028 | *MIR4675;NEBL* | intergenic | OpenSea | 0.11(0.07,0.14) | 9.9E-11 | -0.04(-0.05,-0.02) | 5.1E-08 |
| cg08474901 | 10 | 13050035 | *CCDC3* | intronic | OpenSea | -0.13(-0.16,-0.09) | 1.1E-10 | -0.02(-0.04,-0.01) | 2.5E-04 |
| cg20744163 | 10 | 79240084 | *KCNMA1* | intronic | N_Shelf | -0.18(-0.23,-0.12) | 1.9E-10 | -0.07(-0.08,-0.06) | 3.0E-22 |
| cg11902170 | 10 | 90593473 | *ANKRD22* | intronic | OpenSea | -0.11(-0.14,-0.07) | 4.1E-09 | -0.08(-0.12,-0.04) | 2.8E-05 |
| cg12519998 | 10 | 338346 | *DIP2C* | intronic | N_Shelf | 0.1(0.07,0.14) | 1.9E-08 | -0.04(-0.05,-0.03) | 1.2E-10 |
| cg17889259 | 10 | 79288587 | *KCNMA1* | intronic | OpenSea | 0.12(0.08,0.16) | 1.9E-08 | -0.03(-0.05,-0.01) | 6.5E-04 |
| cg02806631 | 10 | 34909833 | *PARD3* | intronic | OpenSea | -0.1(-0.14,-0.07) | 5.3E-08 | -0.04(-0.05,-0.02) | 9.6E-07 |
| cg18107573 | 10 | 11814507 | *ECHDC3;PROSER2* | intergenic | OpenSea | -0.11(-0.14,-0.07) | 5.4E-08 | -0.06(-0.08,-0.05) | 3.2E-21 |

| | | | | | | | | | |
|---|---|---|---|---|---|---|---|---|---|
| cg07920092 | 11 | 46361501 | *DGKZ* | intronic | OpenSea | -0.11(-0.13,-0.08) | 1.2E-18 | -0.07(-0.08,-0.06) | 1.6E-31 |
| cg16986026 | 11 | 120241281 | *ARHGEF12* | intronic | S_Shore | 0.11(0.08,0.13) | 1.5E-18 | -0.08(-0.1,-0.07) | 8.3E-19 |
| cg24183187 | 11 | 2135231 | *H19;IGF2* | intergenic | S_Shelf;N_Shelf | -0.1(-0.12,-0.08) | 1.8E-17 | -0.08(-0.1,-0.06) | 4.1E-16 |
| cg09144398 | 11 | 118679776 | *DDX6;CXCR5* | intergenic | OpenSea | 0.12(0.09,0.15) | 3.3E-17 | 0.09(0.06,0.13) | 2.1E-07 |
| cg21728792 | 11 | 2135444 | *H19;IGF2* | intergenic | S_Shelf;N_Shelf | -0.12(-0.15,-0.09) | 4.0E-17 | -0.06(-0.08,-0.04) | 5.9E-13 |
| cg02723558 | 11 | 27514526 | *LOC105376671;LIN7C* | intergenic | OpenSea | 0.17(0.13,0.21) | 8.2E-17 | -0.11(-0.14,-0.09) | 4.7E-16 |
| cg26683426 | 11 | 121653408 | *SORL1;MIR100HG* | intergenic | OpenSea | -0.1(-0.13,-0.08) | 1.2E-15 | -0.04(-0.05,-0.03) | 3.9E-13 |
| cg19944895 | 11 | 62914623 | *SLC22A24;SLC22A25* | intergenic | OpenSea | -0.12(-0.15,-0.09) | 3.5E-15 | 0.11(0.08,0.14) | 2.3E-14 |
| cg27609217 | 11 | 46236488 | *LOC101928894;CREB3L1* | intergenic | N_Shore;N_Shelf | -0.11(-0.14,-0.09) | 1.2E-14 | -0.04(-0.06,-0.03) | 1.2E-07 |
| cg16003049 | 11 | 2168904 | *INS-IGF2* | exonic | S_Shore | 0.16(0.12,0.2) | 3.5E-14 | -0.07(-0.09,-0.05) | 1.7E-12 |
| cg00221747 | 11 | 2131198 | *H19;IGF2* | intergenic | N_Shore | -0.12(-0.16,-0.09) | 1.1E-12 | -0.15(-0.17,-0.12) | 8.7E-40 |
| cg22206745 | 11 | 69669789 | *FGF3;LINC02753* | intergenic | OpenSea | 0.13(0.09,0.16) | 1.8E-12 | 0.05(0.03,0.06) | 2.3E-12 |
| cg25274540 | 11 | 43895808 | *HSD17B12;ALKBH3* | intergenic | OpenSea | -0.11(-0.14,-0.08) | 1.2E-11 | 0(-0.01,0) | 1.6E-01 |
| cg16983824 | 11 | 120089541 | *OAF* | intronic | OpenSea | 0.11(0.08,0.15) | 1.7E-10 | -0.06(-0.08,-0.04) | 6.3E-10 |
| cg20016416 | 11 | 66722050 | *PC* | intronic | OpenSea | -0.1(-0.14,-0.07) | 3.0E-10 | -0.03(-0.04,-0.02) | 7.0E-12 |
| cg08849509 | 11 | 9815661 | *SBF2-AS1* | ncRNA_intronic | OpenSea | 0.11(0.08,0.15) | 1.7E-09 | -0.13(-0.16,-0.11) | 9.1E-26 |
| cg27255502 | 11 | 75371754 | *MAP6* | intronic | OpenSea | 0.11(0.07,0.15) | 5.0E-09 | -0.04(-0.06,-0.03) | 3.2E-10 |
| cg08257257 | 11 | 68750336 | *MRGPRD;MRGPRF* | intergenic | Island | -0.11(-0.14,-0.07) | 1.2E-08 | -0.05(-0.06,-0.04) | 3.7E-14 |
| cg11201229 | 11 | 17169688 | *PIK3C2A* | intronic | OpenSea | -0.1(-0.14,-0.07) | 1.9E-08 | -0.01(-0.02,0) | 4.9E-03 |
| cg06679296 | 11 | 1927802 | *LINC01150* | ncRNA_intronic | Shore;Island;N_Sh | -0.11(-0.14,-0.07) | 2.0E-08 | -0.04(-0.05,-0.03) | 2.6E-09 |
| cg16781933 | 11 | 95549248 | *CEP57* | intronic | OpenSea | -0.11(-0.15,-0.07) | 5.6E-08 | 0.01(0,0.01) | 6.0E-03 |
| cg05616242 | 12 | 54389632 | *HOXC-AS2* | ncRNA_exonic | OpenSea | 0.1(0.08,0.12) | 9.8E-16 | -0.11(-0.13,-0.08) | 4.0E-20 |
| cg17603826 | 12 | 51013725 | *DIP2B* | intronic | OpenSea | 0.12(0.09,0.15) | 4.9E-15 | -0.13(-0.17,-0.1) | 2.4E-12 |
| cg17562253 | 12 | 47903846 | *LINC02156;RPAP3* | intergenic | N_Shore | 0.12(0.09,0.16) | 3.2E-14 | -0.07(-0.09,-0.06) | 1.8E-24 |
| cg07066860 | 12 | 11779155 | *LINC01252;ETV6* | intergenic | OpenSea | -0.2(-0.25,-0.14) | 1.7E-13 | -0.05(-0.06,-0.03) | 2.5E-11 |
| cg21919117 | 12 | 12979600 | *DDX47* | intronic | OpenSea | -0.1(-0.13,-0.08) | 2.7E-13 | 0.03(0.02,0.05) | 2.7E-04 |
| cg13538965 | 12 | 15852530 | *EPS8* | intronic | OpenSea | -0.13(-0.17,-0.1) | 4.1E-13 | 0.07(0.05,0.09) | 1.1E-10 |
| cg17537502 | 12 | 44619739 | *TMEM117* | intronic | OpenSea | -0.1(-0.13,-0.07) | 1.7E-11 | -0.04(-0.05,-0.02) | 6.4E-08 |
| cg05268743 | 12 | 93563962 | *LOC643339* | ncRNA_intronic | OpenSea | 0.11(0.07,0.14) | 2.2E-11 | -0.1(-0.13,-0.07) | 9.9E-11 |
| cg17891246 | 12 | 88538783 | *TMTC3* | intronic | OpenSea | 0.11(0.08,0.14) | 4.0E-11 | -0.04(-0.05,-0.03) | 1.7E-11 |
| cg00100361 | 12 | 110314497 | *GLTP* | intronic | OpenSea | -0.11(-0.14,-0.07) | 9.3E-11 | -0.07(-0.09,-0.06) | 4.3E-16 |
| cg17894886 | 12 | 89212475 | *KITLG;LINC02458* | intergenic | OpenSea | -0.13(-0.18,-0.09) | 3.5E-10 | -0.1(-0.12,-0.08) | 1.1E-24 |
| cg02083251 | 12 | 27039986 | *ITPR2;INTS13* | intergenic | OpenSea | -0.11(-0.14,-0.07) | 4.4E-10 | -0.01(-0.02,-0.01) | 6.5E-04 |
| cg14641582 | 12 | 53257943 | *KRT78;KRT8* | intergenic | OpenSea | -0.1(-0.14,-0.07) | 9.8E-10 | -0.02(-0.03,-0.02) | 1.2E-11 |

| | | | | | | | | | |
|---|---|---|---|---|---|---|---|---|---|
| cg17772773 | 12 | 69065914 | *RAP1B* | downstream | OpenSea | -0.11(-0.15,-0.07) | 3.0E-08 | -0.07(-0.09,-0.05) | 2.1E-14 |
| cg00777315 | 13 | 94481085 | *GPC6-AS2* | ncRNA_intronic | OpenSea | 0.11(0.08,0.13) | 2.7E-16 | -0.03(-0.04,-0.02) | 1.4E-09 |
| cg08780680 | 13 | 24066824 | *LINC00327;TNFRSF19* | intergenic | OpenSea | 0.11(0.08,0.14) | 4.0E-16 | -0.07(-0.08,-0.06) | 1.1E-27 |
| cg03715269 | 13 | 94433198 | *GPC6* | intronic | OpenSea | -0.1(-0.13,-0.07) | 5.9E-14 | -0.11(-0.14,-0.08) | 4.2E-12 |
| cg19121101 | 13 | 106468446 | *LINC00343;LINC00460* | intergenic | OpenSea | 0.1(0.07,0.13) | 1.2E-12 | -0.03(-0.04,-0.02) | 3.5E-10 |
| cg18774577 | 13 | 48505919 | *LINC00562* | ncRNA_exonic | OpenSea | 0.14(0.1,0.18) | 1.7E-12 | -0.04(-0.05,-0.02) | 6.6E-10 |
| cg04067341 | 13 | 110508360 | *IRS2;LINC00396* | intergenic | N_Shelf | 0.12(0.09,0.16) | 6.9E-12 | 0.11(0.08,0.14) | 6.7E-12 |
| cg23941537 | 13 | 73101881 | *DACH1;MZT1* | intergenic | OpenSea | -0.13(-0.17,-0.09) | 1.4E-10 | -0.04(-0.05,-0.02) | 3.7E-08 |
| cg26169783 | 13 | 33797700 | *STARD13* | intronic | OpenSea | 0.11(0.08,0.15) | 3.0E-10 | -0.03(-0.05,-0.02) | 5.8E-05 |
| cg12678751 | 13 | 112719229 | *SOX1-OT* | ncRNA_intronic | OpenSea | -0.12(-0.16,-0.08) | 1.2E-08 | -0.02(-0.03,-0.01) | 7.0E-06 |
| cg22749750 | 13 | 84137510 | *NONE;SLITRK1* | intergenic | OpenSea | 0.12(0.08,0.16) | 4.9E-08 | -0.06(-0.08,-0.04) | 7.5E-13 |
| cg01423044 | 13 | 46164185 | *ERICH6B* | intronic | OpenSea | -0.13(-0.18,-0.08) | 8.7E-08 | -0.02(-0.03,-0.01) | 5.4E-04 |
| cg01714756 | 14 | 94314063 | *PRIMA1;FAM181A-AS1* | intergenic | OpenSea | 0.16(0.13,0.19) | 1.6E-24 | -0.09(-0.11,-0.07) | 1.2E-16 |
| cg19489600 | 14 | 51337991 | *ABHD12B* | upstream | OpenSea | 0.12(0.1,0.14) | 2.2E-23 | -0.04(-0.05,-0.03) | 7.8E-10 |
| cg18925726 | 14 | 23513265 | *PSMB11* | UTR3 | OpenSea | 0.11(0.08,0.13) | 5.0E-17 | -0.04(-0.05,-0.03) | 2.4E-15 |
| cg19004811 | 14 | 75476770 | *EIF2B2* | UTR3 | OpenSea | -0.11(-0.14,-0.08) | 1.8E-15 | 0.07(0.06,0.08) | 8.1E-21 |
| cg05883794 | 14 | 99413316 | *C14orf177;BCL11B* | intergenic | OpenSea | -0.16(-0.21,-0.12) | 5.6E-14 | -0.08(-0.1,-0.07) | 2.2E-24 |
| cg17334357 | 14 | 99617073 | *C14orf177;BCL11B* | intergenic | OpenSea | -0.13(-0.16,-0.09) | 3.1E-13 | 0.08(0.06,0.1) | 1.3E-21 |
| cg16943230 | 14 | 91466411 | *RPS6KA5* | intronic | OpenSea | 0.14(0.1,0.18) | 3.2E-11 | 0.21(0.18,0.25) | 7.9E-38 |
| cg20485781 | 14 | 103552398 | *LBHD2* | exonic | OpenSea | 0.11(0.07,0.14) | 7.6E-09 | -0.04(-0.05,-0.03) | 1.4E-17 |
| cg17326611 | 14 | 34482574 | *EGLN3;SPTSSA* | intergenic | OpenSea | -0.11(-0.14,-0.07) | 1.3E-08 | 0.09(0.07,0.1) | 2.6E-21 |
| cg19743728 | 14 | 50242813 | *KLHDC2* | intronic | OpenSea | -0.11(-0.15,-0.07) | 3.8E-08 | -0.03(-0.05,-0.02) | 6.5E-05 |
| cg19580937 | 15 | 70448090 | *TLE3;LINC02205* | intergenic | OpenSea | -0.14(-0.18,-0.11) | 4.7E-18 | -0.05(-0.07,-0.04) | 7.4E-17 |
| cg15661950 | 15 | 81880426 | *TMC3-AS1;MEX3B* | intergenic | OpenSea | 0.12(0.09,0.15) | 7.4E-16 | 0.05(0.04,0.07) | 4.7E-09 |
| cg20383223 | 15 | 51052020 | *SPPL2A* | intronic | OpenSea | 0.12(0.09,0.15) | 6.8E-14 | -0.03(-0.05,-0.02) | 1.9E-05 |
| cg24401660 | 15 | 66819312 | *ZWILCH* | intronic | OpenSea | 0.11(0.08,0.14) | 2.5E-12 | -0.09(-0.11,-0.07) | 1.1E-13 |
| cg20508648 | 15 | 64567036 | *CSNK1G1* | intronic | OpenSea | -0.11(-0.15,-0.08) | 3.1E-11 | -0.03(-0.04,-0.02) | 1.3E-09 |
| cg20268564 | 15 | 39621215 | *C15orf54;THBS1* | intergenic | OpenSea | -0.11(-0.15,-0.08) | 4.1E-10 | -0.09(-0.12,-0.07) | 2.8E-12 |
| cg17883068 | 15 | 56985436 | *ZNF280D* | intronic | OpenSea | 0.15(0.1,0.19) | 5.1E-10 | -0.05(-0.06,-0.03) | 5.6E-08 |
| cg13286635 | 15 | 48385141 | *LINC01491;SLC24A5* | intergenic | OpenSea | 0.1(0.07,0.14) | 1.1E-08 | -0.07(-0.09,-0.05) | 2.0E-18 |
| cg01030757 | 15 | 73573987 | *NEO1* | intronic | OpenSea | 0.11(0.07,0.15) | 5.6E-08 | -0.05(-0.06,-0.04) | 1.9E-19 |
| cg21854374 | 16 | 84737742 | *USP10* | intronic | OpenSea | 0.13(0.09,0.16) | 3.3E-15 | 0.12(0.1,0.15) | 6.9E-20 |
| cg07532353 | 16 | 85575053 | *MIR5093;GSE1* | intergenic | OpenSea | -0.12(-0.15,-0.09) | 2.5E-14 | 0.05(0.03,0.07) | 5.4E-09 |
| cg21128374 | 16 | 10930096 | *TVP23A;CIITA* | intergenic | OpenSea | 0.11(0.08,0.14) | 4.6E-12 | -0.03(-0.04,-0.02) | 6.3E-08 |

| | | | | | | | | | |
|---|---|---|---|---|---|---|---|---|---|
| cg07595197 | 16 | 79670023 | *MAF;MAFTRR* | intergenic | OpenSea | 0.11(0.07,0.15) | 9.7E-09 | 0.06(0.04,0.09) | 2.0E-07 |
| cg11766101 | 16 | 66481318 | *BEAN1* | intronic | OpenSea | -0.11(-0.14,-0.07) | 1.1E-08 | -0.03(-0.04,-0.02) | 2.2E-06 |
| cg21895560 | 16 | 86664553 | *FOXL1;LINC02189* | intergenic | OpenSea | -0.14(-0.19,-0.09) | 2.4E-08 | -0.04(-0.06,-0.03) | 1.5E-07 |
| cg23565609 | 17 | 81843050 | *RPL23AP87;NONE* | intergenic | S_Shore;N_Shelf | 0.13(0.1,0.16) | 2.1E-19 | -0.04(-0.05,-0.03) | 1.5E-13 |
| cg22482278 | 17 | 82331755 | *RPL23AP87;NONE* | intergenic | N_Shore | 0.14(0.11,0.17) | 2.2E-19 | -0.09(-0.11,-0.07) | 4.1E-16 |
| cg03837680 | 17 | 79993730 | *DCXR* | downstream | S_Shore | -0.13(-0.16,-0.1) | 1.7E-18 | -0.04(-0.06,-0.03) | 3.0E-10 |
| cg04998091 | 17 | 77267267 | *RBFOX3* | intronic | OpenSea | -0.1(-0.13,-0.07) | 1.1E-14 | -0.08(-0.1,-0.06) | 8.5E-15 |
| cg01890836 | 17 | 17152270 | *COPS3* | exonic | OpenSea | 0.1(0.07,0.13) | 3.1E-13 | -0.03(-0.04,-0.02) | 2.2E-09 |
| cg27244773 | 17 | 45405750 | *EFCAB13* | intron | N_Shore | -0.11(-0.14,-0.08) | 6.1E-13 | -0.04(-0.05,-0.02) | 2.3E-09 |
| cg26980152 | 17 | 78680591 | *RPTOR* | intronic | OpenSea | -0.14(-0.18,-0.1) | 6.1E-12 | -0.05(-0.06,-0.04) | 7.6E-13 |
| cg22487572 | 17 | 38443442 | *CDC6* | upstream | OpenSea | -0.1(-0.13,-0.07) | 7.8E-12 | -0.04(-0.05,-0.03) | 4.0E-14 |
| cg22660070 | 17 | 49616812 | *LINC02073;CA10* | intergenic | OpenSea | 0.12(0.08,0.16) | 4.7E-10 | -0.04(-0.05,-0.02) | 1.4E-08 |
| cg00231966 | 17 | 82419423 | *RPL23AP87;NONE* | intergenic | S_Shelf;S_Shore | 0.1(0.07,0.14) | 1.7E-09 | -0.03(-0.04,-0.02) | 3.0E-08 |
| cg16667827 | 17 | 59725937 | *NACA2;BRIP1* | intergenic | OpenSea | -0.16(-0.21,-0.11) | 2.3E-09 | 0.07(0.05,0.1) | 9.3E-12 |
| cg05688017 | 17 | 40622703 | *ATP6V0A1* | intronic | OpenSea | -0.11(-0.14,-0.07) | 1.1E-08 | -0.07(-0.08,-0.05) | 6.4E-24 |
| cg24414383 | 17 | 41042692 | *LINC00671* | ncRNA_intronic | OpenSea | 0.13(0.08,0.17) | 2.0E-08 | 0.02(0.02,0.03) | 1.4E-08 |
| cg11824764 | 17 | 80838687 | *TBCD* | intronic | N_Shore | -0.1(-0.14,-0.07) | 2.4E-08 | -0.07(-0.09,-0.05) | 2.7E-10 |
| cg15742818 | 17 | 27586429 | *NUFIP2* | UTR3 | OpenSea | -0.12(-0.16,-0.08) | 7.6E-08 | 0.02(0.01,0.03) | 2.4E-07 |
| cg03862381 | 19 | 18464963 | *PGPEP1* | intronic | S_Shelf | 0.1(0.08,0.13) | 1.8E-21 | -0.09(-0.12,-0.06) | 3.9E-10 |
| cg00984412 | 19 | 13001548 | *GCDH* | upstream | Island | 0.12(0.09,0.15) | 2.8E-15 | 0.06(0.04,0.08) | 9.6E-09 |
| cg24625605 | 19 | 46303573 | *RSPH6A* | intronic | N_Shore | -0.12(-0.15,-0.09) | 2.3E-14 | -0.08(-0.11,-0.06) | 1.1E-12 |
| cg21658153 | 19 | 16067760 | *OR10H4;LINC00661* | intergenic | Island | 0.11(0.08,0.14) | 4.7E-14 | -0.08(-0.1,-0.06) | 9.5E-14 |
| cg01809838 | 19 | 46303413 | *RSPH6A* | intronic | N_Shore | -0.14(-0.18,-0.11) | 1.7E-13 | -0.08(-0.1,-0.06) | 1.1E-14 |
| cg27548752 | 19 | 46760357 | *IGFL1;HIF3A* | intergenic | S_Shelf | 0.12(0.08,0.15) | 5.5E-13 | 0.09(0.07,0.11) | 1.2E-19 |
| cg19014858 | 19 | 50968555 | *MYBPC2* | intronic | OpenSea | -0.1(-0.13,-0.07) | 7.6E-11 | -0.05(-0.06,-0.04) | 5.3E-26 |
| cg18522395 | 19 | 12105113 | *ZNF433-AS1* | ncRNA_intronic | OpenSea | 0.12(0.08,0.16) | 5.4E-10 | 0(-0.01,0) | 2.3E-04 |
| cg24765446 | 20 | 45539335 | *EYA2* | intronic | OpenSea | 0.11(0.09,0.14) | 2.6E-15 | 0.1(0.07,0.13) | 4.9E-13 |
| cg25227018 | 20 | 37349615 | *ARHGAP40;SLC32A1* | intergenic | S_Shelf | 0.12(0.09,0.15) | 1.3E-14 | -0.15(-0.2,-0.1) | 1.8E-09 |
| cg18039268 | 20 | 3980435 | *RNF24* | intronic | OpenSea | 0.14(0.11,0.18) | 5.4E-14 | -0.06(-0.07,-0.04) | 2.4E-18 |
| cg14153983 | 20 | 39826203 | *ZHX3* | intronic | OpenSea | 0.16(0.12,0.2) | 3.2E-13 | -0.06(-0.07,-0.04) | 2.5E-15 |
| cg25074484 | 20 | 23145501 | *LINC00656;NXT1* | intergenic | OpenSea | 0.11(0.08,0.14) | 4.5E-12 | -0.08(-0.1,-0.05) | 9.4E-11 |
| cg04302055 | 20 | 52493501 | *SUMO1P1;BCAS1* | intergenic | OpenSea | -0.11(-0.14,-0.07) | 2.0E-09 | -0.01(-0.02,0) | 5.3E-03 |
| cg25263251 | 20 | 41061465 | *PTPRT* | intronic | OpenSea | 0.11(0.07,0.14) | 7.4E-08 | -0.03(-0.04,-0.01) | 2.9E-04 |
| cg25982653 | 21 | 46189039 | *UBE2G2* | UTR3 | OpenSea | -0.1(-0.13,-0.07) | 1.5E-13 | -0.1(-0.13,-0.07) | 2.3E-14 |

| | | | | | | | | | |
|---|---|---|---|---|---|---|---|---|---|
| cg07374077 | 21 | 41220593 | *IGSF5;PCP4* | intergenic | OpenSea | -0.11(-0.14,-0.07) | 9.5E-11 | 0.07(0.04,0.09) | 4.3E-09 |
| cg05297739 | 21 | 36071951 | *CLIC6* | intronic | S_Shore | 0.11(0.07,0.15) | 7.2E-08 | -0.05(-0.06,-0.04) | 2.4E-13 |
| cg07222631 | 22 | 27809086 | *LINC02554;MN1* | intergenic | OpenSea | -0.18(-0.22,-0.15) | 1.4E-26 | -0.06(-0.08,-0.05) | 5.5E-17 |
| cg19034820 | 22 | 28147187 | *MN1* | intronic | OpenSea | -0.1(-0.13,-0.08) | 1.3E-17 | -0.03(-0.05,-0.02) | 1.1E-08 |
| cg25909582 | 22 | 30506826 | *HORMAD2* | intronic | S_Shore | 0.11(0.08,0.13) | 1.2E-14 | -0.02(-0.04,-0.01) | 9.4E-06 |
| cg00187327 | 22 | 27795793 | *LINC02554;MN1* | intergenic | N_Shore | -0.12(-0.16,-0.09) | 6.8E-14 | 0.08(0.06,0.09) | 9.8E-19 |
| cg24493244 | 22 | 20546491 | *FAM230G* | ncRNA_intronic | OpenSea | -0.11(-0.14,-0.08) | 5.1E-13 | -0.06(-0.09,-0.04) | 1.3E-09 |
| cg26467624 | 22 | 46084652 | *ATXN10* | intronic | N_Shore | 0.11(0.08,0.14) | 5.3E-13 | -0.07(-0.09,-0.04) | 7.0E-08 |
| cg23825480 | 22 | 30940798 | *SEC14L6* | intronic | OpenSea | -0.1(-0.13,-0.07) | 2.1E-12 | 0.02(0,0.04) | 2.2E-02 |
| cg17744019 | 22 | 47038726 | *GRAMD4* | intronic | OpenSea | -0.1(-0.14,-0.07) | 1.7E-10 | -0.04(-0.05,-0.02) | 2.2E-08 |
| cg18168435 | 22 | 36156886 | *RBFOX2* | intronic | OpenSea | 0.1(0.07,0.13) | 3.5E-10 | -0.05(-0.07,-0.04) | 3.2E-14 |
| cg08935541 | 22 | 46040669 | *LINC01589;ATXN10* | intergenic | N_Shelf | 0.1(0.07,0.14) | 3.9E-10 | 0.05(0.03,0.07) | 5.4E-08 |
| cg22413388 | 22 | 45971737 | *FBLN1* | intronic | Island;N_Shelf | -0.11(-0.15,-0.07) | 7.1E-08 | -0.03(-0.04,-0.02) | 1.2E-06 |
| cg17281810 | X | 40148696 | *LOC101927476;ATP6AP2* | intergenic | S_Shore;N_Shelf | -0.11(-0.13,-0.08) | 8.0E-17 | -0.01(-0.02,-0.01) | 7.2E-04 |
| cg01466576 | X | 129653149 | *DENND10P1;ENOX2* | intergenic | N_Shore | 0.12(0.09,0.16) | 2.8E-14 | -0.04(-0.05,-0.03) | 8.6E-12 |
| cg09161311 | X | 23770255 | *ACOT9;SAT1* | intergenic | OpenSea | 0.11(0.08,0.14) | 1.4E-12 | -0.02(-0.03,-0.01) | 7.1E-04 |
| cg23864546 | X | 9656386 | *TBL1X* | intronic | OpenSea | 0.12(0.08,0.15) | 7.2E-12 | -0.04(-0.05,-0.03) | 7.2E-11 |
| cg08940231 | X | 13009063 | *TMSB4X;FAM9C* | intergenic | OpenSea | 0.12(0.09,0.16) | 4.8E-11 | -0.03(-0.04,-0.02) | 2.9E-07 |
| cg25996557 | X | 12140277 | *FRMPD4* | intronic | S_Shore | -0.12(-0.17,-0.08) | 3.2E-08 | -0.02(-0.04,-0.01) | 1.3E-03 |
| cg26939342 | X | 40300785 | *LOC101927476;ATP6AP2* | intergenic | OpenSea | -0.11(-0.15,-0.07) | 4.2E-08 | -0.07(-0.1,-0.04) | 6.2E-06 |

**Supplementary Table 3**

Associations identified with epigenome-significance in the EWAS of early preterm birth (ePTB) among girls (N=134). Showing effect estimates observed for same signals in the boys' analysis for comparison. Associations were deemed significant at p < 9.4x10-8.

| CpG | CHR | POS | Near Gene | Gene context | CG island context | Girls analysis Effect size (95% CI) | *P* | Boys analysis Effect size (95% CI) | *P* |
|---|---|---|---|---|---|---|---|---|---|
| cg20465027 | 1 | 19045582 | *PAX7* | intronic | OpenSea | 0.11(0.09,0.14) | 4.0E-25 | -0.07(-0.09,-0.05) | 5.3E-10 |
| cg04266340 | 1 | 206080002 | *SLC26A9;FAM72A* | intergenic | OpenSea | 0.13(0.1,0.15) | 6.4E-25 | -0.07(-0.08,-0.05) | 5.1E-19 |
| cg08655953 | 1 | 2494449 | *TNFRSF14* | intronic | OpenSea | -0.22(-0.27,-0.18) | 2.0E-22 | -0.07(-0.1,-0.05) | 3.9E-08 |
| cg25193276 | 1 | 41435975 | *CITED4;CTPS1* | intergenic | OpenSea | -0.14(-0.17,-0.12) | 1.7E-21 | -0.07(-0.08,-0.05) | 4.0E-20 |
| cg00748668 | 1 | 50620998 | *ELAVL4* | intronic | OpenSea | 0.12(0.1,0.15) | 4.7E-21 | -0.06(-0.07,-0.04) | 7.7E-17 |
| cg06170720 | 1 | 6477549 | *HES2* | UTR3 | OpenSea | 0.12(0.1,0.15) | 1.4E-20 | -0.08(-0.1,-0.06) | 1.9E-11 |
| cg06322310 | 1 | 2504775 | *LOC100996583* | ncRNA_intronic | S_Shelf;S_Shore | -0.14(-0.17,-0.11) | 2.8E-20 | -0.03(-0.04,-0.02) | 4.4E-08 |
| cg17499490 | 1 | 226606906 | *PARP1;STUM* | intergenic | OpenSea | 0.11(0.09,0.13) | 3.2E-19 | -0.04(-0.05,-0.03) | 5.6E-13 |
| cg24986815 | 1 | 226941273 | *ITPKB;PSEN2* | intergenic | S_Shore | -0.13(-0.16,-0.1) | 3.0E-18 | 0.06(0.05,0.08) | 2.0E-17 |
| cg21406409 | 1 | 10353642 | *KIF1B* | intronic | OpenSea | 0.11(0.09,0.14) | 6.0E-18 | -0.05(-0.07,-0.04) | 4.7E-11 |
| cg24038105 | 1 | 15352803 | *KAZN* | intronic | OpenSea | -0.1(-0.13,-0.08) | 1.7E-16 | 0.06(0.04,0.07) | 4.2E-14 |
| cg07350078 | 1 | 206464119 | *CTSE;SRGAP2* | intergenic | OpenSea | -0.2(-0.25,-0.15) | 2.0E-16 | 0.09(0.07,0.11) | 1.4E-21 |
| cg27526774 | 1 | 234963342 | *PP2672;LOC101927851* | intergenic | OpenSea | -0.13(-0.16,-0.1) | 2.4E-16 | -0.1(-0.14,-0.07) | 1.3E-08 |
| cg01948134 | 1 | 214659048 | *PTPN14* | intronic | OpenSea | -0.11(-0.13,-0.08) | 3.5E-16 | -0.03(-0.04,-0.02) | 6.8E-08 |
| cg12828504 | 1 | 2128552 | *FAAP20* | intronic | N_Shelf | -0.13(-0.17,-0.1) | 1.4E-15 | -0.07(-0.09,-0.05) | 3.2E-10 |
| cg07915117 | 1 | 31955530 | *SERINC2;LDC1P* | intergenic | OpenSea | -0.12(-0.15,-0.09) | 4.0E-15 | -0.12(-0.16,-0.07) | 7.4E-08 |
| cg06906523 | 1 | 220335698 | *RAB3GAP2* | intronic | OpenSea | 0.11(0.08,0.13) | 1.5E-14 | 0.11(0.08,0.14) | 1.7E-11 |
| cg09336794 | 1 | 2477900 | *HES5;TNFRSF14-AS1* | intergenic | N_Shore;N_Shore | -0.11(-0.13,-0.08) | 3.5E-14 | -0.05(-0.07,-0.04) | 1.2E-13 |
| cg12633102 | 1 | 35210888 | *MIR552;GJB5* | intergenic | OpenSea | -0.12(-0.15,-0.09) | 1.4E-13 | -0.02(-0.03,-0.01) | 3.9E-04 |
| cg27596556 | 1 | 93751922 | *CCDC18;CCDC18-AS1* | intergenic | OpenSea | -0.11(-0.13,-0.08) | 1.8E-13 | 0.06(0.04,0.08) | 1.1E-11 |
| cg02087739 | 1 | 192512642 | *RGS21;RGS1* | intergenic | OpenSea | -0.1(-0.13,-0.08) | 4.1E-13 | -0.03(-0.05,-0.01) | 3.5E-03 |
| cg13555278 | 1 | 26021993 | *MAN1C1* | intronic | OpenSea | -0.12(-0.15,-0.09) | 6.2E-13 | -0.03(-0.05,-0.02) | 8.6E-05 |
| cg00552781 | 1 | 33244345 | *YARS* | intronic | OpenSea | 0.11(0.08,0.13) | 2.1E-12 | -0.04(-0.06,-0.02) | 1.7E-06 |
| cg20858526 | 1 | 15438569 | *TMEM51-AS1* | ncRNA_exonic | OpenSea | -0.12(-0.16,-0.09) | 2.8E-12 | -0.02(-0.04,-0.01) | 3.0E-04 |
| cg18836974 | 1 | 51668152 | *LINC01562* | ncRNA_intronic | OpenSea | 0.17(0.12,0.21) | 3.4E-12 | -0.04(-0.06,-0.03) | 8.5E-09 |
| cg25468058 | 1 | 150981204 | *PRUNE1* | intronic | OpenSea | -0.11(-0.14,-0.08) | 4.6E-12 | -0.03(-0.04,-0.02) | 2.9E-10 |
| cg00004905 | 1 | 115461677 | *SYCP1* | intronic | OpenSea | 0.11(0.08,0.14) | 4.9E-12 | 0.05(0.04,0.07) | 2.6E-15 |
| cg00657048 | 1 | 41753089 | *SCMH1;FOXO6* | intergenic | OpenSea | -0.11(-0.14,-0.08) | 7.0E-12 | 0.04(0.01,0.06) | 1.1E-03 |
| cg04205753 | 1 | 204476069 | *PIK3C2B;MDM4* | intergenic | OpenSea | -0.1(-0.13,-0.07) | 7.7E-12 | -0.01(-0.02,0) | 3.5E-03 |

| | | | | | | | | | |
|---|---|---|---|---|---|---|---|---|---|
| cg07937270 | 1 | 44806961 | *ERI3* | intronic | S_Shore | -0.12(-0.16,-0.09) | 7.7E-12 | 0.08(0.06,0.1) | 2.9E-13 |
| cg11968713 | 1 | 204196981 | *PLEKHA6* | intronic | OpenSea | -0.15(-0.19,-0.11) | 1.1E-11 | -0.02(-0.03,-0.01) | 5.4E-04 |
| cg02045580 | 1 | 226350096 | *ACBD3* | intronic | OpenSea | 0.11(0.08,0.14) | 2.2E-11 | -0.05(-0.08,-0.03) | 5.7E-06 |
| cg04286677 | 1 | 93581334 | *MTF2* | intronic | OpenSea | -0.13(-0.17,-0.09) | 2.5E-11 | -0.05(-0.07,-0.04) | 2.8E-09 |
| cg00624255 | 1 | 39174693 | *LINC01685;RRAGC* | intergenic | OpenSea | -0.17(-0.22,-0.12) | 3.3E-11 | -0.03(-0.05,-0.01) | 3.6E-03 |
| cg02519744 | 1 | 232529486 | *TSNAX-DISC1;SIPA1L2* | intergenic | OpenSea | 0.11(0.08,0.14) | 7.0E-11 | 0.05(0.03,0.07) | 1.9E-06 |
| cg21749436 | 1 | 207849427 | *CR1L* | intronic | OpenSea | 0.11(0.08,0.14) | 1.6E-10 | 0.04(0.01,0.07) | 5.2E-03 |
| cg26663783 | 1 | 183293518 | *NMNAT2* | intronic | OpenSea | -0.11(-0.15,-0.08) | 1.8E-10 | 0.03(0.02,0.04) | 4.3E-07 |
| cg04738197 | 1 | 19787238 | *CAPZB* | intronic | OpenSea | 0.11(0.07,0.14) | 8.1E-10 | -0.03(-0.05,-0.02) | 3.5E-06 |
| cg00065572 | 1 | 2556842 | *MMEL1* | intronic | OpenSea | 0.14(0.09,0.18) | 8.1E-10 | 0.04(0.03,0.06) | 1.0E-10 |
| cg00140004 | 1 | 205538519 | *MFSD4A* | intronic | OpenSea | 0.1(0.07,0.14) | 1.3E-09 | -0.04(-0.07,-0.02) | 2.8E-04 |
| cg07888841 | 1 | 25503069 | *MIR4425;SYF2* | intergenic | OpenSea | 0.12(0.08,0.16) | 1.5E-09 | -0.05(-0.07,-0.03) | 4.7E-09 |
| cg09838159 | 1 | 7726014 | *CAMTA1* | intronic | OpenSea | 0.1(0.07,0.14) | 2.6E-09 | -0.01(-0.01,0) | 2.1E-01 |
| cg22016234 | 1 | 205246580 | *TMCC2;NUAK2* | intergenic | OpenSea | -0.12(-0.16,-0.08) | 3.1E-09 | -0.11(-0.15,-0.08) | 3.2E-10 |
| cg23222898 | 1 | 42152576 | *HIVEP3* | intronic | OpenSea | 0.1(0.07,0.14) | 3.8E-09 | 0.03(0.02,0.04) | 6.4E-07 |
| cg09858176 | 1 | 16018798 | *PLEKHM2* | intronic | OpenSea | 0.11(0.07,0.15) | 4.4E-09 | -0.01(-0.02,0) | 7.9E-04 |
| cg20414082 | 1 | 203033465 | *PPFIA4* | intronic | OpenSea | -0.1(-0.14,-0.07) | 4.5E-09 | -0.02(-0.03,0) | 1.3E-02 |
| cg04938922 | 1 | 6321842 | *GPR153* | upstream | OpenSea | -0.1(-0.14,-0.07) | 5.2E-09 | -0.01(-0.02,0) | 2.1E-02 |
| cg09422942 | 1 | 203753502 | *LAX1;ZC3H11A* | intergenic | OpenSea | 0.1(0.07,0.14) | 4.1E-08 | -0.03(-0.04,-0.01) | 6.8E-05 |
| cg18270687 | 1 | 38570310 | *MIR3659;LINC01343* | intergenic | OpenSea | -0.1(-0.14,-0.06) | 5.2E-08 | -0.06(-0.08,-0.04) | 1.1E-09 |
| cg23707274 | 2 | 20188779 | *WDR35* | intronic | OpenSea | -0.16(-0.19,-0.13) | 3.1E-21 | -0.04(-0.05,-0.03) | 1.5E-10 |
| cg06101959 | 2 | 8475624 | *LINC00299;LINC01814* | intergenic | OpenSea | -0.13(-0.16,-0.1) | 4.4E-21 | -0.07(-0.09,-0.05) | 2.6E-15 |
| cg00760938 | 2 | 216492436 | *LINC00607* | ncRNA_intronic | OpenSea | -0.11(-0.13,-0.09) | 4.0E-20 | -0.09(-0.12,-0.06) | 2.8E-11 |
| cg23604266 | 2 | 6960890 | *LINC00487;NRIR* | intergenic | OpenSea | -0.11(-0.14,-0.09) | 1.5E-19 | 0.08(0.06,0.11) | 7.6E-12 |
| cg20734664 | 2 | 47752560 | *KCNK12* | intronic | OpenSea | -0.11(-0.13,-0.08) | 3.1E-19 | -0.08(-0.11,-0.06) | 1.5E-10 |
| cg03163262 | 2 | 235836470 | *LINC01173;SH3BP4* | intergenic | OpenSea | -0.11(-0.13,-0.08) | 3.6E-19 | -0.06(-0.08,-0.04) | 4.3E-10 |
| cg10094994 | 2 | 239240627 | *TRAF3IP1* | intronic | OpenSea | 0.14(0.11,0.18) | 1.2E-18 | -0.03(-0.04,-0.02) | 3.6E-06 |
| cg24801254 | 2 | 4141289 | *LOC105373394;LINC01249* | intergenic | OpenSea | 0.1(0.08,0.13) | 1.8E-17 | -0.04(-0.06,-0.01) | 1.7E-03 |
| cg17364007 | 2 | 128362748 | *MYO7B* | intronic | OpenSea | 0.13(0.1,0.16) | 7.9E-17 | -0.07(-0.09,-0.05) | 4.3E-14 |
| cg19206131 | 2 | 26002937 | *ASXL2* | intronic | OpenSea | 0.12(0.09,0.15) | 2.4E-16 | 0.07(0.05,0.09) | 5.2E-12 |
| cg18570243 | 2 | 241764257 | *KIF1A;AGXT* | intergenic | OpenSea | -0.11(-0.13,-0.08) | 5.8E-15 | -0.1(-0.13,-0.06) | 1.3E-08 |
| cg21503763 | 2 | 62444115 | *B3GNT2* | intronic | OpenSea | -0.15(-0.19,-0.11) | 7.4E-15 | 0.13(0.09,0.17) | 9.2E-10 |
| cg13883684 | 2 | 202687315 | *CDK15* | intronic | OpenSea | -0.12(-0.15,-0.09) | 9.6E-15 | 0.05(0.02,0.07) | 9.3E-04 |
| cg23970275 | 2 | 207143328 | *ZDBF2* | intronic | OpenSea | -0.15(-0.19,-0.11) | 1.4E-14 | -0.04(-0.06,-0.03) | 8.5E-07 |
| cg18245735 | 2 | 219579058 | *TTLL4* | intronic | OpenSea | 0.13(0.1,0.16) | 1.8E-14 | -0.09(-0.12,-0.06) | 1.0E-07 |
| cg14864829 | 2 | 37362076 | *EIF2AK2* | intronic | OpenSea | 0.12(0.09,0.15) | 2.3E-14 | 0.1(0.07,0.12) | 1.1E-14 |
| cg03015311 | 2 | 74745831 | *DQX1* | intronic | OpenSea | 0.1(0.08,0.13) | 2.3E-14 | -0.02(-0.03,-0.01) | 4.0E-03 |

| | | | | | | | | | |
|---|---|---|---|---|---|---|---|---|---|
| cg15150970 | 2 | 25250660 | *DNAJC27-AS1* | ncRNA_intronic | N_Shore;N_Shelf | -0.11(-0.14,-0.08) | 3.2E-14 | -0.04(-0.05,-0.03) | 5.3E-09 |
| cg19302297 | 2 | 66305786 | *SPRED2;MIR4778* | intergenic | OpenSea | 0.11(0.08,0.13) | 6.8E-14 | 0.05(0.03,0.07) | 4.8E-07 |
| cg04705280 | 2 | 8501341 | *LINC00299;LINC01814* | intergenic | OpenSea | -0.12(-0.15,-0.09) | 7.6E-14 | -0.03(-0.04,-0.02) | 4.4E-08 |
| cg23371680 | 2 | 31288164 | *GALNT14* | intronic | OpenSea | 0.11(0.08,0.14) | 3.5E-13 | 0.04(0.02,0.06) | 8.0E-05 |
| cg09694782 | 2 | 96743062 | *GPAT2;ADRA2B* | intergenic | S_Shelf | 0.1(0.07,0.13) | 4.2E-13 | -0.05(-0.07,-0.03) | 1.2E-06 |
| cg03032690 | 2 | 109062907 | *SULT1C4;GCC2* | intergenic | OpenSea | -0.1(-0.13,-0.07) | 5.7E-13 | -0.01(-0.02,0) | 1.4E-01 |
| cg02622498 | 2 | 30301737 | *ALK;YPEL5* | intergenic | OpenSea | -0.1(-0.13,-0.07) | 1.6E-12 | -0.05(-0.06,-0.03) | 1.5E-06 |
| cg02548816 | 2 | 24410313 | *FAM228A* | intronic | OpenSea | -0.1(-0.13,-0.08) | 2.7E-12 | -0.03(-0.04,-0.01) | 1.7E-04 |
| cg27072594 | 2 | 176764024 | *ATP5MC3;LNPK* | intergenic | OpenSea | 0.1(0.07,0.13) | 4.1E-12 | 0.05(0.03,0.06) | 2.6E-10 |
| cg13158936 | 2 | 44282988 | *LRPPRC;PPM1B* | intergenic | OpenSea | 0.14(0.1,0.17) | 7.2E-12 | 0.1(0.06,0.13) | 5.9E-09 |
| cg24196622 | 2 | 14615963 | *LINC00276;LRATD1* | intergenic | OpenSea | 0.11(0.08,0.14) | 1.4E-11 | -0.03(-0.05,-0.02) | 1.2E-05 |
| cg02699666 | 2 | 38945705 | *GALM* | intronic | OpenSea | -0.12(-0.16,-0.09) | 1.5E-11 | -0.03(-0.05,-0.01) | 6.4E-04 |
| cg25183214 | 2 | 221495706 | *MIR4268;EPHA4* | intergenic | OpenSea | -0.1(-0.13,-0.07) | 2.7E-11 | -0.06(-0.1,-0.02) | 2.1E-03 |
| cg15714846 | 2 | 207766960 | *FASTKD2;CPO* | intergenic | Island | -0.19(-0.24,-0.13) | 3.0E-11 | -0.08(-0.11,-0.04) | 6.0E-05 |
| cg02317642 | 2 | 74051203 | *C2orf78;STAMBP* | intergenic | OpenSea | -0.1(-0.13,-0.07) | 3.1E-11 | -0.02(-0.03,-0.01) | 5.2E-03 |
| cg23093954 | 2 | 169514136 | *CERS6* | intronic | OpenSea | -0.1(-0.13,-0.07) | 3.5E-11 | -0.05(-0.08,-0.03) | 2.3E-06 |
| cg02069095 | 2 | 133262469 | *GPR39* | intronic | N_Shelf | -0.14(-0.18,-0.1) | 4.9E-11 | -0.12(-0.16,-0.09) | 1.9E-12 |
| cg03804880 | 2 | 172339903 | *DCAF17* | UTR3 | OpenSea | -0.12(-0.16,-0.09) | 5.0E-11 | 0.04(0.02,0.06) | 3.1E-05 |
| cg04372478 | 2 | 120887860 | *EPB41L5* | intronic | OpenSea | -0.1(-0.13,-0.07) | 1.1E-10 | -0.04(-0.06,-0.02) | 5.0E-06 |
| cg15418831 | 2 | 237905363 | *ACKR3;LOC93463* | intergenic | OpenSea | -0.11(-0.14,-0.07) | 1.2E-10 | -0.02(-0.04,-0.01) | 1.3E-03 |
| cg03239958 | 2 | 102057937 | *RFX8* | intronic | OpenSea | 0.13(0.09,0.18) | 2.3E-10 | -0.03(-0.04,-0.01) | 9.1E-05 |
| cg14632054 | 2 | 226795357 | *NYAP2;LOC646736* | intergenic | S_Shelf;N_Shelf | -0.1(-0.13,-0.07) | 2.6E-10 | -0.04(-0.05,-0.03) | 1.5E-12 |
| cg20405062 | 2 | 30366611 | *ALK;YPEL5* | intergenic | OpenSea | 0.11(0.07,0.14) | 2.6E-10 | -0.03(-0.04,-0.02) | 1.6E-07 |
| cg23564664 | 2 | 113240340 | *TTL* | intronic | S_Shelf | -0.18(-0.24,-0.12) | 3.6E-10 | 0.01(0,0.02) | 6.9E-03 |
| cg02578106 | 2 | 201127718 | *MAIP1;SPATS2L* | intergenic | OpenSea | 0.11(0.07,0.14) | 4.7E-10 | 0.03(0.02,0.04) | 2.3E-07 |
| cg19661658 | 2 | 28363249 | *BABAM2* | intronic | OpenSea | -0.14(-0.19,-0.1) | 4.8E-10 | -0.05(-0.06,-0.03) | 7.1E-11 |
| cg02446816 | 2 | 162093347 | *LOC101929512* | ncRNA_intronic | OpenSea | -0.14(-0.18,-0.09) | 5.7E-10 | -0.08(-0.12,-0.05) | 2.6E-06 |
| cg21117965 | 2 | 219460647 | *CNOT9* | UTR3 | OpenSea | -0.1(-0.14,-0.07) | 1.3E-09 | -0.07(-0.09,-0.04) | 2.9E-07 |
| cg20128773 | 2 | 236014890 | *SH3BP4;AGAP1* | intergenic | OpenSea | -0.1(-0.14,-0.07) | 2.2E-09 | -0.01(-0.02,-0.01) | 1.5E-03 |
| cg00137918 | 2 | 190986183 | *MSTN;C2orf88* | intergenic | OpenSea | -0.11(-0.15,-0.08) | 2.2E-09 | -0.02(-0.03,-0.01) | 3.1E-06 |
| cg03328004 | 2 | 111777324 | *ACOXL* | intronic | OpenSea | -0.11(-0.15,-0.08) | 4.3E-09 | -0.01(-0.02,0.01) | 4.6E-01 |
| cg04410886 | 2 | 36515341 | *MIR548AD;CRIM1-DT* | intergenic | OpenSea | -0.12(-0.16,-0.08) | 6.1E-09 | -0.04(-0.06,-0.03) | 6.3E-07 |
| cg06638795 | 2 | 42492793 | *EML4* | intronic | Island | -0.12(-0.16,-0.08) | 1.0E-08 | -0.08(-0.11,-0.05) | 3.7E-07 |
| cg16444777 | 2 | 46337962 | *PRKCE* | intronic | OpenSea | 0.1(0.07,0.14) | 2.2E-08 | -0.02(-0.04,0) | 1.2E-01 |
| cg20219891 | 2 | 120739299 | *PTPN4* | UTR3 | S_Shore;N_Shelf | -0.11(-0.14,-0.07) | 2.3E-08 | -0.05(-0.07,-0.02) | 3.2E-05 |
| cg04971583 | 3 | 57151164 | *IL17RD* | intronic | OpenSea | -0.1(-0.12,-0.08) | 1.5E-20 | -0.06(-0.08,-0.04) | 1.7E-10 |
| cg10922765 | 3 | 8593671 | *LMCD1* | intronic | OpenSea | 0.1(0.08,0.12) | 4.9E-19 | -0.07(-0.09,-0.05) | 4.8E-12 |

| | | | | | | | | | |
|---|---|---|---|---|---|---|---|---|---|
| cg15608892 | 3 | 127002743 | *PRR20G* | UTR3 | OpenSea | 0.12(0.09,0.15) | 5.1E-19 | -0.03(-0.04,-0.02) | 3.1E-07 |
| cg09825286 | 3 | 127582044 | *MGLL;KBTBD12* | intergenic | OpenSea | -0.12(-0.15,-0.09) | 5.2E-19 | -0.03(-0.04,-0.02) | 7.1E-08 |
| cg15649559 | 3 | 40766191 | *ZNF621;CTNNB1* | intergenic | OpenSea | -0.12(-0.15,-0.09) | 1.1E-17 | 0.15(0.11,0.2) | 3.8E-10 |
| cg21902516 | 3 | 13767651 | *LINC00620* | ncRNA_intronic | OpenSea | -0.12(-0.15,-0.09) | 4.9E-17 | -0.1(-0.13,-0.08) | 3.8E-19 |
| cg08510718 | 3 | 12978149 | *IQSEC1* | exonic | OpenSea | 0.11(0.08,0.13) | 5.4E-17 | -0.07(-0.11,-0.04) | 4.1E-06 |
| cg04638085 | 3 | 21257345 | *LOC101927829;VENTXP7* | intergenic | OpenSea | -0.14(-0.18,-0.11) | 5.2E-16 | -0.07(-0.09,-0.05) | 5.7E-10 |
| cg02011912 | 3 | 53893006 | *IL17RB* | intronic | S_Shore | -0.12(-0.15,-0.09) | 8.6E-16 | -0.05(-0.06,-0.03) | 1.2E-08 |
| cg16736924 | 3 | 112115450 | *CD200;BTLA* | intergenic | OpenSea | 0.12(0.09,0.15) | 1.2E-15 | -0.08(-0.1,-0.06) | 9.2E-11 |
| cg07585771 | 3 | 32844096 | *CNOT10;TRIM71* | intergenic | OpenSea | -0.13(-0.17,-0.1) | 3.5E-15 | -0.05(-0.07,-0.03) | 7.9E-07 |
| cg05315047 | 3 | 138047885 | *NME9* | intronic | OpenSea | -0.14(-0.17,-0.1) | 4.3E-15 | -0.06(-0.08,-0.04) | 1.7E-12 |
| cg04481360 | 3 | 5006880 | *BHLHE40-AS1* | ncRNA_intronic | OpenSea | -0.1(-0.13,-0.07) | 3.3E-14 | -0.05(-0.06,-0.03) | 4.7E-11 |
| cg27199647 | 3 | 142275882 | *ATR* | intronic | OpenSea | -0.14(-0.18,-0.11) | 3.6E-14 | -0.06(-0.08,-0.04) | 4.8E-08 |
| cg17051399 | 3 | 100045899 | *TBC1D23;NIT2* | intergenic | OpenSea | -0.11(-0.14,-0.08) | 4.9E-14 | -0.02(-0.04,-0.01) | 4.1E-04 |
| cg12120178 | 3 | 188385496 | *LPP* | intronic | OpenSea | -0.11(-0.14,-0.08) | 5.0E-14 | -0.09(-0.12,-0.05) | 3.5E-06 |
| cg08362543 | 3 | 150248549 | *TSC22D2;SERP1* | intergenic | OpenSea | -0.13(-0.16,-0.09) | 7.8E-14 | -0.05(-0.07,-0.03) | 4.2E-07 |
| cg10710607 | 3 | 59252187 | *C3orf67;LOC339902* | intergenic | OpenSea | 0.12(0.09,0.16) | 3.5E-13 | 0.05(0.04,0.07) | 2.2E-09 |
| cg04443299 | 3 | 13421122 | *NUP210* | exonic | S_Shore | 0.11(0.08,0.14) | 8.8E-13 | -0.06(-0.07,-0.04) | 1.2E-10 |
| cg02116058 | 3 | 134252313 | *CEP63* | intronic | S_Shore | 0.1(0.07,0.13) | 1.5E-12 | -0.05(-0.07,-0.04) | 7.7E-11 |
| cg11429281 | 3 | 38028145 | *CTDSPL;VILL* | intergenic | S_Shelf;N_Shore | -0.13(-0.16,-0.09) | 3.0E-12 | 0.03(0.02,0.05) | 1.3E-06 |
| cg08633861 | 3 | 49814432 | *IP6K1* | intronic | OpenSea | 0.1(0.07,0.13) | 3.6E-12 | 0.06(0.04,0.07) | 9.8E-14 |
| cg04585783 | 3 | 14462687 | *SLC6A6* | intronic | OpenSea | 0.11(0.08,0.14) | 7.2E-12 | -0.1(-0.13,-0.07) | 5.4E-11 |
| cg05771324 | 3 | 171614731 | *TMEM212-AS1* | ncRNA_intronic | OpenSea | 0.12(0.09,0.16) | 7.4E-12 | -0.04(-0.05,-0.02) | 2.1E-07 |
| cg18482576 | 3 | 127631346 | *MGLL;KBTBD12* | intergenic | S_Shore | 0.12(0.08,0.15) | 1.0E-11 | -0.03(-0.04,-0.02) | 1.6E-06 |
| cg05150709 | 3 | 78992309 | *ROBO1* | intronic | OpenSea | -0.14(-0.18,-0.1) | 1.1E-11 | -0.05(-0.07,-0.04) | 1.6E-09 |
| cg02600660 | 3 | 143362737 | *SLC9A9* | intronic | OpenSea | -0.1(-0.13,-0.07) | 1.5E-11 | -0.03(-0.04,-0.02) | 7.4E-11 |
| cg00109834 | 3 | 142091054 | *XRN1* | intronic | OpenSea | 0.1(0.07,0.13) | 2.3E-11 | -0.09(-0.11,-0.07) | 4.3E-13 |
| cg21499981 | 3 | 194194701 | *ATP13A3* | intronic | OpenSea | -0.11(-0.15,-0.08) | 8.0E-11 | -0.04(-0.05,-0.03) | 5.9E-09 |
| cg01769483 | 3 | 47443023 | *PTPN23* | intronic | OpenSea | -0.13(-0.17,-0.09) | 1.3E-10 | 0.05(0.03,0.07) | 2.4E-07 |
| cg20569075 | 3 | 182118117 | *LINC01206;LINC01994* | intergenic | OpenSea | -0.11(-0.15,-0.08) | 1.5E-10 | -0.04(-0.07,-0.02) | 1.3E-03 |
| cg05784401 | 3 | 168040845 | *EGFEM1P* | ncRNA_intronic | OpenSea | 0.11(0.08,0.15) | 5.7E-10 | -0.02(-0.05,0.01) | 2.7E-01 |
| cg26334888 | 3 | 58667469 | *FAM3D;C3orf67* | intergenic | OpenSea | -0.13(-0.18,-0.09) | 9.0E-10 | -0.05(-0.06,-0.03) | 1.1E-07 |
| cg13002603 | 3 | 150248905 | *TSC22D2;SERP1* | intergenic | OpenSea | -0.11(-0.14,-0.07) | 1.6E-09 | -0.04(-0.06,-0.02) | 3.7E-05 |
| cg18069880 | 3 | 195005360 | *ACAP2* | intronic | OpenSea | -0.13(-0.18,-0.09) | 1.8E-09 | -0.02(-0.03,-0.01) | 3.4E-04 |
| cg19760612 | 3 | 197270695 | *BDH1* | intronic | OpenSea | -0.1(-0.14,-0.07) | 2.2E-09 | -0.01(-0.03,0) | 5.7E-02 |
| cg03745746 | 3 | 52315323 | *WDR82;GLYCTK* | intergenic | OpenSea | 0.11(0.07,0.14) | 3.9E-09 | -0.01(-0.02,0) | 4.9E-03 |
| cg02733814 | 3 | 61685995 | *PTPRG* | intronic | OpenSea | 0.13(0.08,0.17) | 4.6E-09 | 0.05(0.03,0.06) | 1.0E-06 |
| cg14056226 | 3 | 64291387 | *PRICKLE2;ADAMTS9* | intergenic | OpenSea | -0.13(-0.17,-0.08) | 9.1E-09 | 0.05(0.03,0.07) | 9.4E-07 |

| | | | | | | | | | |
|---|---|---|---|---|---|---|---|---|---|
| cg15653350 | 3 | 52841359 | *ITIH3* | intronic | OpenSea | 0.12(0.07,0.16) | 4.9E-08 | -0.01(-0.02,0) | 2.3E-01 |
| cg25981343 | 3 | 39531752 | *MOBP* | intronic | OpenSea | -0.11(-0.16,-0.07) | 6.9E-08 | -0.08(-0.11,-0.04) | 1.3E-05 |
| cg22389845 | 3 | 14540448 | *GRIP2* | intronic | OpenSea | -0.11(-0.15,-0.07) | 7.9E-08 | -0.03(-0.04,-0.01) | 2.8E-04 |
| cg02635407 | 4 | 8198822 | *ABLIM2;SH3TC1* | intergenic | OpenSea | 0.13(0.1,0.17) | 4.1E-27 | 0.06(0.04,0.08) | 2.4E-10 |
| cg05899788 | 4 | 8198808 | *ABLIM2;SH3TC1* | intergenic | OpenSea | 0.12(0.1,0.14) | 3.7E-25 | 0.08(0.05,0.1) | 2.2E-09 |
| cg05455266 | 4 | 1366013 | *UVSSA* | intronic | OpenSea | 0.1(0.08,0.12) | 7.5E-20 | -0.06(-0.09,-0.03) | 6.0E-05 |
| cg14323984 | 4 | 6693476 | *LINC002481;S100P* | intergenic | OpenSea | -0.14(-0.18,-0.11) | 2.7E-15 | -0.05(-0.07,-0.03) | 3.1E-06 |
| cg07904475 | 4 | 150579779 | *IQCM* | intronic | Island;N_Shelf | -0.13(-0.16,-0.1) | 1.8E-14 | -0.03(-0.04,-0.02) | 2.7E-07 |
| cg06213122 | 4 | 7344865 | *SORCS2* | intronic | OpenSea | -0.11(-0.14,-0.08) | 1.9E-13 | -0.05(-0.07,-0.03) | 1.2E-06 |
| cg25965776 | 4 | 52957676 | *SPATA18* | intronic | OpenSea | -0.11(-0.13,-0.08) | 2.6E-13 | -0.03(-0.04,-0.01) | 3.7E-04 |
| cg07244414 | 4 | 7905496 | *AFAP1* | intronic | OpenSea | -0.12(-0.16,-0.09) | 2.7E-12 | -0.1(-0.13,-0.08) | 2.2E-16 |
| cg01484685 | 4 | 153265807 | *FBXW7* | intronic | OpenSea | 0.13(0.1,0.17) | 5.6E-12 | -0.05(-0.07,-0.03) | 9.5E-06 |
| cg10808122 | 4 | 6523569 | *PPP2R2C* | intronic | OpenSea | 0.13(0.09,0.17) | 2.2E-10 | -0.03(-0.06,-0.01) | 6.7E-03 |
| cg04030971 | 4 | 121189957 | *MAD2L1;PRDM5* | intergenic | OpenSea | -0.1(-0.13,-0.07) | 2.4E-10 | 0.04(0.03,0.05) | 1.0E-13 |
| cg09114672 | 4 | 7026311 | *TBC1D14* | intronic | S_Shore | 0.1(0.07,0.13) | 3.7E-10 | -0.07(-0.09,-0.04) | 1.0E-06 |
| cg24029426 | 4 | 25993874 | *SMIM20;RBPJ* | intergenic | OpenSea | -0.14(-0.19,-0.1) | 1.4E-09 | -0.05(-0.06,-0.04) | 4.6E-14 |
| cg27106766 | 4 | 157043259 | *CTSO;PDGFC* | intergenic | OpenSea | -0.13(-0.17,-0.09) | 8.9E-09 | -0.01(-0.02,0) | 8.4E-02 |
| cg08719697 | 5 | 151745676 | *LINC01933;NMUR2* | intergenic | OpenSea | 0.11(0.09,0.13) | 3.6E-21 | 0.06(0.04,0.08) | 1.8E-08 |
| cg23112946 | 5 | 10761982 | *DAP* | upstream | S_Shore | -0.12(-0.15,-0.09) | 3.0E-18 | -0.1(-0.14,-0.06) | 1.2E-07 |
| cg07897352 | 5 | 38979703 | *RICTOR* | intronic | OpenSea | -0.12(-0.15,-0.1) | 4.5E-18 | 0.05(0.03,0.07) | 1.6E-08 |
| cg08266095 | 5 | 123814022 | *LINC01170;ZNF608* | intergenic | OpenSea | 0.12(0.09,0.15) | 9.6E-18 | 0.07(0.05,0.1) | 1.7E-07 |
| cg19938826 | 5 | 145838618 | *TCERG1* | exonic | S_Shelf | -0.11(-0.13,-0.08) | 6.0E-16 | -0.1(-0.12,-0.07) | 8.4E-15 |
| cg08431289 | 5 | 12636543 | *LINC01194* | ncRNA_intronic | OpenSea | -0.14(-0.18,-0.11) | 3.0E-15 | -0.2(-0.26,-0.15) | 1.3E-12 |
| cg05763702 | 5 | 77437389 | *AP3B1* | intronic | OpenSea | -0.11(-0.13,-0.08) | 9.3E-15 | -0.07(-0.09,-0.05) | 1.1E-12 |
| cg18477820 | 5 | 135349105 | *LECT2;TGFBI* | intergenic | OpenSea | -0.15(-0.2,-0.11) | 4.4E-13 | 0.05(0.02,0.08) | 2.5E-04 |
| cg23449231 | 5 | 179812690 | *GFPT2;CNOT6* | intergenic | N_Shelf | -0.14(-0.18,-0.11) | 8.3E-13 | 0.09(0.06,0.11) | 1.4E-11 |
| cg08780718 | 5 | 159572060 | *PWWP2A;FABP6* | intergenic | OpenSea | -0.1(-0.13,-0.07) | 1.6E-12 | -0.03(-0.05,-0.02) | 4.0E-05 |
| cg08087091 | 5 | 68504801 | *CENPH* | intronic | OpenSea | -0.14(-0.19,-0.1) | 7.0E-12 | -0.05(-0.08,-0.03) | 8.3E-05 |
| cg22026677 | 5 | 172861442 | *MIR8056;LOC285593* | intergenic | OpenSea | -0.11(-0.15,-0.08) | 1.0E-11 | 0.1(0.07,0.13) | 2.8E-11 |
| cg21023952 | 5 | 60487208 | *SMIM15-AS1* | ncRNA_intronic | OpenSea | -0.1(-0.13,-0.07) | 7.7E-11 | -0.06(-0.08,-0.04) | 4.2E-10 |
| cg08354494 | 5 | 107395981 | *FBXL17* | intronic | OpenSea | 0.15(0.1,0.19) | 1.4E-10 | -0.01(-0.03,0) | 3.9E-02 |
| cg08247299 | 5 | 89902094 | *ADGRV1* | intronic | OpenSea | 0.1(0.07,0.14) | 2.8E-10 | 0.06(0.04,0.08) | 9.5E-10 |
| cg06564875 | 5 | 157926772 | *LINC02227;EBF1* | intergenic | OpenSea | -0.1(-0.13,-0.07) | 3.0E-10 | -0.08(-0.1,-0.05) | 1.9E-08 |
| cg20941894 | 5 | 17147749 | *BASP1-AS1* | ncRNA_intronic | OpenSea | -0.12(-0.15,-0.08) | 3.4E-10 | -0.05(-0.07,-0.04) | 2.3E-12 |
| cg03284061 | 5 | 127858650 | *FBN2* | intronic | OpenSea | 0.11(0.08,0.15) | 4.5E-10 | 0.04(0.01,0.08) | 7.7E-03 |
| cg26222075 | 5 | 151958557 | *NMUR2;LINC01470* | intergenic | OpenSea | -0.11(-0.15,-0.08) | 4.7E-10 | -0.08(-0.11,-0.05) | 5.2E-08 |
| cg10030197 | 5 | 178281606 | *AACSP1;ZNF354B* | intergenic | OpenSea | -0.1(-0.14,-0.07) | 1.6E-08 | -0.01(-0.02,0) | 6.4E-02 |

| | | | | | | | | | |
|---|---|---|---|---|---|---|---|---|---|
| cg01868577 | 5 | 92927320 | *NR2F1* | intronic | OpenSea | 0.12(0.08,0.16) | 2.2E-08 | -0.07(-0.1,-0.04) | 5.9E-07 |
| cg23948167 | 5 | 134574717 | *C5orf66;C5orf66-AS2* | ncRNA_intronic | OpenSea | 0.15(0.1,0.21) | 4.3E-08 | 0.02(0.01,0.03) | 1.8E-04 |
| cg08695296 | 5 | 149632761 | *CAMK2A* | intronic | OpenSea | -0.12(-0.16,-0.08) | 5.8E-08 | 0.02(0.01,0.03) | 2.8E-03 |
| cg20254998 | 6 | 136655645 | *BCLAF1;MAP7* | intergenic | OpenSea | 0.11(0.09,0.13) | 5.3E-22 | -0.09(-0.12,-0.07) | 1.9E-11 |
| cg21737106 | 6 | 44665016 | *LOC105375075;SUPT3H* | intergenic | OpenSea | -0.1(-0.13,-0.08) | 6.6E-19 | -0.09(-0.12,-0.06) | 3.9E-08 |
| cg10156772 | 6 | 138563916 | *ARFGEF3* | intronic | OpenSea | -0.12(-0.14,-0.09) | 7.7E-19 | -0.05(-0.07,-0.03) | 1.3E-08 |
| cg17716056 | 6 | 73421876 | *KCNQ5* | intronic | OpenSea | 0.1(0.08,0.12) | 7.2E-17 | 0.08(0.06,0.1) | 6.9E-17 |
| cg07878585 | 6 | 136501722 | *PDE7B* | intronic | OpenSea | 0.11(0.08,0.14) | 1.5E-14 | -0.01(-0.02,0) | 1.1E-02 |
| cg17940495 | 6 | 14282651 | *LINC01108* | ncRNA_exonic | OpenSea | -0.14(-0.18,-0.1) | 3.7E-14 | -0.03(-0.04,-0.02) | 2.8E-05 |
| cg09509966 | 6 | 43266907 | *SLC22A7* | intronic | S_Shelf;N_Shelf | 0.1(0.07,0.13) | 2.6E-13 | -0.02(-0.03,-0.01) | 6.6E-04 |
| cg10374523 | 6 | 118299304 | *SLC35F1* | intronic | OpenSea | -0.18(-0.22,-0.13) | 2.8E-13 | 0.04(0.02,0.06) | 2.5E-05 |
| cg17806418 | 6 | 29631542 | *MOG* | intronic | S_Shelf;N_Shore | 0.12(0.09,0.15) | 5.2E-13 | -0.05(-0.06,-0.03) | 8.1E-08 |
| cg21704448 | 6 | 132089048 | *ENPP3;MIR548H5* | intergenic | OpenSea | -0.13(-0.16,-0.09) | 5.6E-13 | -0.03(-0.05,-0.01) | 8.2E-04 |
| cg07560287 | 6 | 41562150 | *FOXP4* | intronic | S_Shore | -0.11(-0.14,-0.08) | 2.6E-12 | -0.04(-0.06,-0.03) | 8.4E-09 |
| cg10171936 | 6 | 140206000 | *FILNC1;LOC100507477* | intergenic | OpenSea | -0.1(-0.13,-0.07) | 3.7E-12 | 0.05(0.04,0.06) | 1.9E-13 |
| cg09264886 | 6 | 21613854 | *SOX4;CASC15* | intergenic | OpenSea | -0.11(-0.14,-0.08) | 6.4E-12 | -0.04(-0.05,-0.02) | 7.2E-07 |
| cg04512899 | 6 | 89545599 | *RNGTT* | intronic | OpenSea | -0.15(-0.19,-0.11) | 6.7E-12 | -0.05(-0.07,-0.03) | 3.4E-05 |
| cg24050441 | 6 | 20907077 | *CDKAL1* | intronic | OpenSea | -0.11(-0.14,-0.08) | 2.0E-11 | -0.07(-0.1,-0.03) | 1.8E-04 |
| cg25498638 | 6 | 7996630 | *BLOC1S5-TXNDC5* | ncRNA_intronic | OpenSea | -0.13(-0.17,-0.09) | 3.1E-11 | -0.04(-0.06,-0.03) | 2.0E-08 |
| cg18304951 | 6 | 33746909 | *LEMD2* | intronic | OpenSea | -0.12(-0.16,-0.08) | 1.7E-10 | -0.02(-0.04,0) | 3.6E-02 |
| cg07610336 | 6 | 6759432 | *LY86;RREB1* | intergenic | OpenSea | -0.11(-0.14,-0.07) | 2.9E-10 | -0.03(-0.05,-0.01) | 4.3E-03 |
| cg06015449 | 6 | 128025931 | *C6orf58;THEMIS* | intergenic | OpenSea | -0.11(-0.15,-0.08) | 1.2E-09 | -0.05(-0.07,-0.03) | 5.9E-06 |
| cg27117853 | 6 | 21354311 | *CDKAL1;LINC00581* | intergenic | OpenSea | -0.1(-0.14,-0.07) | 1.8E-09 | -0.07(-0.08,-0.05) | 1.4E-15 |
| cg06281325 | 6 | 106798128 | *ATG5;CRYBG1* | intergenic | OpenSea | -0.11(-0.15,-0.07) | 2.8E-09 | -0.04(-0.06,-0.02) | 2.0E-04 |
| cg12764291 | 6 | 143396720 | *AIG1* | intronic | OpenSea | -0.1(-0.14,-0.07) | 3.5E-08 | -0.03(-0.05,-0.02) | 1.1E-04 |
| cg19803298 | 6 | 13267683 | *TBC1D7-LOC100130357* | UTR3 | OpenSea | 0.12(0.08,0.17) | 3.6E-08 | -0.09(-0.12,-0.06) | 2.3E-09 |
| cg18966495 | 6 | 22222508 | *CASC15;PRL* | intergenic | OpenSea | -0.1(-0.14,-0.07) | 3.8E-08 | -0.03(-0.05,0) | 2.4E-02 |
| cg10097240 | 6 | 131183143 | *EPB41L2* | intronic | OpenSea | -0.11(-0.15,-0.07) | 7.4E-08 | -0.03(-0.05,-0.02) | 3.7E-04 |
| cg09965127 | 6 | 111068166 | *CDK19* | intronic | OpenSea | -0.11(-0.15,-0.07) | 8.1E-08 | -0.04(-0.05,-0.02) | 1.6E-05 |
| cg14010305 | 7 | 2043326 | *MAD1L1* | intronic | OpenSea | -0.12(-0.15,-0.1) | 4.7E-17 | -0.06(-0.08,-0.04) | 2.0E-10 |
| cg20789819 | 7 | 130831614 | *MKLN1* | intronic | OpenSea | -0.16(-0.2,-0.12) | 5.6E-17 | 0.04(0.02,0.06) | 2.4E-05 |
| cg18978125 | 7 | 923414 | *GET4* | intronic | Shelf;S_Shelf;N_Sl | -0.1(-0.13,-0.08) | 3.6E-16 | -0.11(-0.15,-0.07) | 5.7E-08 |
| cg04744624 | 7 | 108001325 | *NRCAM* | intronic | N_Shore | -0.17(-0.22,-0.13) | 2.8E-15 | -0.06(-0.08,-0.03) | 3.1E-07 |
| cg10627562 | 7 | 5696084 | *RNF216* | intronic | OpenSea | -0.11(-0.13,-0.08) | 6.3E-15 | -0.04(-0.06,-0.02) | 1.4E-04 |
| cg20307391 | 7 | 75559765 | *POR* | intronic | Island | 0.1(0.08,0.13) | 2.1E-13 | -0.02(-0.03,0) | 1.1E-02 |
| cg19497702 | 7 | 2289111 | *NUDT1* | intronic | OpenSea | -0.11(-0.14,-0.08) | 7.7E-13 | -0.04(-0.05,-0.03) | 1.8E-10 |
| cg08675999 | 7 | 131009565 | *MKLN1-AS* | ncRNA_intronic | OpenSea | 0.12(0.09,0.16) | 1.1E-11 | 0.04(0.02,0.06) | 1.4E-04 |

| | | | | | | | | | |
|---|---|---|---|---|---|---|---|---|---|
| cg11747856 | 7 | 135176443 | *CNOT4* | intronic | OpenSea | -0.11(-0.15,-0.08) | 5.7E-11 | 0.05(0.03,0.07) | 2.7E-08 |
| cg00516678 | 7 | 30911198 | *INMT-MINDY4* | ncRNA_intronic | OpenSea | -0.1(-0.13,-0.07) | 1.9E-10 | -0.03(-0.05,-0.02) | 1.3E-07 |
| cg10153714 | 7 | 116511867 | *CAPZA2* | intronic | OpenSea | -0.11(-0.15,-0.08) | 2.1E-10 | 0.06(0.04,0.08) | 4.4E-10 |
| cg00560277 | 7 | 42227670 | *GLI3* | intronic | OpenSea | 0.11(0.08,0.15) | 3.1E-10 | -0.03(-0.05,-0.01) | 2.6E-03 |
| cg02355442 | 7 | 138008773 | *MIR4468;TRIM24* | intergenic | OpenSea | -0.1(-0.14,-0.07) | 3.0E-09 | -0.04(-0.06,-0.02) | 9.3E-05 |
| cg01058360 | 7 | 151745285 | *GALNT11* | intronic | OpenSea | -0.12(-0.16,-0.08) | 3.1E-09 | -0.05(-0.07,-0.03) | 4.4E-08 |
| cg13748800 | 7 | 107940526 | *NRCAM* | intronic | OpenSea | -0.11(-0.14,-0.07) | 2.5E-08 | 0(-0.01,0) | 4.0E-01 |
| cg10558261 | 7 | 2167149 | *MAD1L1* | intronic | OpenSea | -0.12(-0.16,-0.07) | 5.0E-08 | -0.03(-0.04,-0.02) | 4.2E-07 |
| cg25766247 | 8 | 58801606 | *LOC286178;LINC01602* | intergenic | OpenSea | -0.1(-0.12,-0.08) | 1.2E-24 | -0.04(-0.06,-0.03) | 2.3E-07 |
| cg05570141 | 8 | 37699441 | *ADGRA2* | exonic | S_Shore;N_Shore | -0.11(-0.13,-0.08) | 4.6E-19 | -0.04(-0.07,-0.02) | 2.2E-04 |
| cg13282461 | 8 | 143565283 | *ADGRB1* | intronic | N_Shore;N_Shelf | -0.1(-0.12,-0.08) | 1.4E-18 | 0.06(0.05,0.08) | 3.5E-16 |
| cg12769632 | 8 | 80279791 | *MIR12123;STMN2* | intergenic | OpenSea | -0.12(-0.15,-0.1) | 2.1E-18 | -0.07(-0.09,-0.05) | 6.7E-11 |
| cg12153259 | 8 | 9041103 | *PPP1R3B;LOC101929128* | intergenic | OpenSea | 0.14(0.11,0.17) | 8.2E-18 | -0.07(-0.09,-0.05) | 7.0E-12 |
| cg26412534 | 8 | 27617459 | *CCDC25* | intronic | S_Shelf | 0.12(0.09,0.14) | 4.4E-17 | 0.03(0.01,0.05) | 2.3E-04 |
| cg12427561 | 8 | 37336760 | *LINC01605* | ncRNA_intronic | OpenSea | 0.14(0.1,0.17) | 5.5E-16 | -0.09(-0.13,-0.06) | 9.9E-07 |
| cg13113226 | 8 | 128105173 | *PRNCR1* | downstream | OpenSea | -0.12(-0.16,-0.09) | 6.9E-16 | 0.05(0.03,0.07) | 9.4E-07 |
| cg03876023 | 8 | 103348635 | *UBR5* | intronic | OpenSea | 0.15(0.11,0.18) | 7.1E-16 | -0.22(-0.28,-0.17) | 2.5E-16 |
| cg09128169 | 8 | 8878459 | *ERI1* | intronic | OpenSea | -0.1(-0.13,-0.08) | 2.1E-14 | -0.03(-0.04,-0.01) | 1.6E-04 |
| cg13312986 | 8 | 144511579 | *MAFA* | exonic | S_Shore;N_Shore | -0.12(-0.15,-0.09) | 5.5E-14 | -0.02(-0.03,-0.01) | 1.6E-03 |
| cg01634958 | 8 | 139878035 | *COL22A1* | intronic | OpenSea | -0.13(-0.16,-0.1) | 1.6E-13 | -0.02(-0.04,-0.01) | 1.2E-04 |
| cg16292343 | 8 | 143463811 | *TSNARE1* | intronic | OpenSea | -0.12(-0.15,-0.09) | 5.1E-13 | -0.05(-0.06,-0.03) | 1.3E-09 |
| cg09234966 | 8 | 100713283 | *VPS13B* | intronic | OpenSea | -0.14(-0.18,-0.1) | 7.3E-12 | -0.06(-0.08,-0.04) | 1.5E-09 |
| cg26782908 | 8 | 122694501 | *HAS2-AS1;SMILR* | intergenic | OpenSea | -0.14(-0.18,-0.1) | 2.3E-11 | 0.06(0.04,0.07) | 1.8E-09 |
| cg17298326 | 8 | 141235378 | *TRAPPC9* | intronic | OpenSea | -0.1(-0.14,-0.07) | 2.6E-10 | -0.07(-0.1,-0.05) | 9.2E-12 |
| cg14735070 | 8 | 96575785 | *C8orf37-AS1* | ncRNA_intronic | OpenSea | 0.13(0.08,0.17) | 3.2E-09 | -0.08(-0.11,-0.05) | 4.6E-08 |
| cg05910407 | 8 | 92572608 | *SLC26A7;RUNX1T1* | intergenic | OpenSea | 0.13(0.08,0.17) | 8.4E-09 | 0.04(0.01,0.06) | 5.1E-03 |
| cg13221448 | 8 | 140645962 | *KCNK9* | intronic | OpenSea | -0.12(-0.16,-0.08) | 9.5E-09 | -0.03(-0.06,-0.01) | 6.1E-03 |
| cg06068343 | 8 | 58594479 | *LOC101929488;LOC286178* | intergenic | OpenSea | -0.12(-0.16,-0.08) | 1.0E-08 | 0.06(0.04,0.07) | 3.0E-11 |
| cg07290627 | 8 | 101541610 | *ANKRD46* | intronic | OpenSea | 0.1(0.07,0.14) | 2.5E-08 | 0.02(-0.01,0.04) | 2.3E-01 |
| cg13933718 | 9 | 92725373 | *UNQ6494;LOC101927847* | intergenic | OpenSea | 0.11(0.1,0.13) | 2.8E-37 | 0.11(0.08,0.13) | 1.3E-21 |
| cg16109426 | 9 | 36566818 | *RNF38;MELK* | intergenic | OpenSea | 0.12(0.1,0.13) | 1.0E-34 | 0.11(0.09,0.13) | 7.5E-21 |
| cg10355819 | 9 | 121464001 | *LINC02578* | ncRNA_intronic | OpenSea | 0.12(0.1,0.14) | 4.0E-24 | 0.12(0.09,0.15) | 5.5E-18 |
| cg01599826 | 9 | 113886158 | *LPAR1;MIR7702* | intergenic | OpenSea | 0.11(0.09,0.13) | 6.4E-24 | 0.08(0.06,0.1) | 2.6E-14 |
| cg13969295 | 9 | 95410042 | *IPPK* | intronic | OpenSea | 0.16(0.12,0.19) | 1.1E-20 | -0.07(-0.1,-0.04) | 4.6E-06 |
| cg14143030 | 9 | 114388073 | *LRRC37A5P;DNAJC25* | intergenic | OpenSea | -0.11(-0.14,-0.09) | 2.9E-19 | -0.06(-0.09,-0.03) | 9.5E-05 |
| cg14514516 | 9 | 137739787 | *LOC101448202* | ncRNA_intronic | OpenSea | 0.14(0.11,0.17) | 3.0E-19 | -0.06(-0.07,-0.04) | 3.7E-16 |
| cg14184527 | 9 | 120387039 | *ASTN2;LOC101928797* | intergenic | OpenSea | -0.17(-0.21,-0.13) | 6.0E-19 | -0.07(-0.09,-0.05) | 6.2E-11 |

| | | | | | | | | | |
|---|---|---|---|---|---|---|---|---|---|
| cg17388916 | 9 | 128607984 | *PBX3* | intronic | OpenSea | 0.11(0.08,0.13) | 4.8E-18 | 0.04(0.02,0.06) | 4.2E-06 |
| cg14080475 | 9 | 114388215 | *LRRC37A5P;DNAJC25* | intergenic | OpenSea | 0.1(0.08,0.13) | 1.7E-17 | -0.05(-0.07,-0.02) | 4.3E-04 |
| cg15422377 | 9 | 133979370 | *AIF1L* | intronic | Island | -0.11(-0.14,-0.09) | 3.5E-17 | -0.03(-0.04,-0.02) | 3.4E-08 |
| cg13905266 | 9 | 90095892 | *C9orf170;DAPK1* | intergenic | OpenSea | -0.14(-0.18,-0.11) | 2.5E-16 | -0.04(-0.05,-0.03) | 1.7E-08 |
| cg00698575 | 9 | 137494152 | *RXRA;COL5A1* | intergenic | N_Shore | -0.1(-0.13,-0.08) | 2.1E-15 | -0.08(-0.11,-0.05) | 4.0E-09 |
| cg14140249 | 9 | 114205416 | *ECPAS* | intronic | OpenSea | -0.1(-0.13,-0.08) | 2.7E-15 | -0.02(-0.03,-0.01) | 1.2E-05 |
| cg17331032 | 9 | 94608238 | *ROR2* | intronic | OpenSea | -0.11(-0.14,-0.08) | 3.9E-14 | -0.02(-0.04,-0.01) | 2.0E-05 |
| cg14529565 | 9 | 93942974 | *LINC00484* | ncRNA_intronic | N_Shelf | 0.1(0.07,0.13) | 3.2E-13 | -0.03(-0.04,-0.01) | 2.1E-03 |
| cg06914855 | 9 | 128867711 | *PBX3;LOC101929116* | intergenic | OpenSea | -0.13(-0.17,-0.1) | 4.2E-13 | -0.09(-0.12,-0.06) | 5.5E-08 |
| cg23460082 | 9 | 136060137 | *GBGT1;OBP2B* | intergenic | OpenSea | -0.13(-0.16,-0.09) | 4.6E-13 | -0.03(-0.05,-0.02) | 4.0E-10 |
| cg13900511 | 9 | 89667800 | *LOC440173;LOC494127* | intergenic | OpenSea | 0.18(0.13,0.23) | 1.1E-12 | -0.02(-0.03,0) | 5.7E-03 |
| cg17491920 | 9 | 36158164 | *GLIPR2* | intronic | OpenSea | -0.11(-0.14,-0.08) | 1.5E-12 | 0.04(0.01,0.06) | 4.0E-03 |
| cg13693256 | 9 | 129336032 | *MVB12B;LMX1B* | intergenic | N_Shore | 0.1(0.07,0.13) | 2.3E-12 | -0.04(-0.06,-0.03) | 3.6E-08 |
| cg13939387 | 9 | 93111922 | *LINC01508* | ncRNA_intronic | OpenSea | -0.11(-0.14,-0.08) | 5.9E-12 | -0.02(-0.03,-0.01) | 2.6E-05 |
| cg13895832 | 9 | 89359241 | *TUT7;GAS1* | intergenic | OpenSea | 0.11(0.07,0.14) | 6.3E-11 | 0.08(0.05,0.1) | 7.4E-08 |
| cg04382851 | 9 | 93397703 | *DIRAS2* | intronic | OpenSea | 0.11(0.08,0.14) | 7.6E-11 | -0.08(-0.11,-0.05) | 2.9E-07 |
| cg13571597 | 9 | 33375398 | *NFX1;AQP7* | intergenic | OpenSea | -0.13(-0.16,-0.09) | 1.7E-10 | -0.02(-0.03,-0.02) | 1.9E-08 |
| cg13396343 | 9 | 8365210 | *PTPRD* | intronic | OpenSea | -0.12(-0.16,-0.08) | 2.7E-10 | -0.08(-0.12,-0.05) | 1.0E-06 |
| cg00753593 | 9 | 70485496 | *CBWD5* | intronic | OpenSea | -0.13(-0.17,-0.09) | 7.6E-10 | -0.02(-0.03,-0.01) | 3.9E-03 |
| cg13372444 | 9 | 5451023 | *CD274* | intronic | S_Shore | 0.11(0.07,0.15) | 6.6E-09 | 0.03(0.01,0.06) | 3.5E-03 |
| cg06889514 | 9 | 98229166 | *LOC100507346* | ncRNA_intronic | OpenSea | -0.11(-0.15,-0.07) | 7.3E-09 | -0.01(-0.02,0) | 3.4E-02 |
| cg06642074 | 9 | 137299448 | *RXRA* | intronic | S_Shelf;N_Shore | -0.13(-0.17,-0.08) | 1.7E-08 | 0(-0.01,0.01) | 8.6E-01 |
| cg13675814 | 9 | 98167515 | *FANCC;PTCH1* | intergenic | OpenSea | -0.1(-0.14,-0.06) | 2.8E-08 | -0.02(-0.04,-0.01) | 4.8E-03 |
| cg07173719 | 10 | 114413356 | *VTI1A* | intronic | OpenSea | -0.15(-0.18,-0.12) | 5.1E-25 | 0.09(0.06,0.12) | 1.5E-10 |
| cg02769133 | 10 | 7625913 | *ITIH5* | intronic | OpenSea | -0.15(-0.18,-0.12) | 7.3E-25 | 0.05(0.02,0.07) | 3.3E-04 |
| cg15178824 | 10 | 69496748 | *CTNNA3;DNAJC12* | intergenic | OpenSea | -0.11(-0.13,-0.09) | 1.9E-23 | 0.07(0.05,0.08) | 1.3E-17 |
| cg23599026 | 10 | 100508789 | *HPSE2* | intronic | OpenSea | 0.12(0.09,0.14) | 8.3E-20 | -0.03(-0.04,-0.02) | 2.3E-07 |
| cg24435571 | 10 | 102676383 | *SLF2* | exonic | OpenSea | -0.11(-0.14,-0.09) | 2.0E-19 | -0.08(-0.11,-0.05) | 1.2E-08 |
| cg16412082 | 10 | 112850876 | *ADRA2A;HEAT2* | intergenic | OpenSea | -0.12(-0.15,-0.1) | 6.9E-19 | -0.03(-0.04,-0.02) | 2.1E-07 |
| cg24935786 | 10 | 71399884 | *FAM241B;COL13A1* | intergenic | S_Shore | -0.12(-0.15,-0.09) | 2.4E-16 | -0.05(-0.07,-0.03) | 6.6E-08 |
| cg12855310 | 10 | 132401531 | *GLRX3;MIR378C* | intergenic | S_Shelf;N_Shelf | 0.15(0.11,0.18) | 1.7E-15 | -0.06(-0.08,-0.04) | 7.9E-13 |
| cg15639387 | 10 | 370128 | *DIP2C* | intronic | S_Shore | -0.12(-0.15,-0.09) | 1.7E-14 | -0.03(-0.04,-0.01) | 3.0E-04 |
| cg25610720 | 10 | 104089967 | *GBF1* | intronic | OpenSea | -0.13(-0.16,-0.1) | 6.6E-14 | 0.05(0.03,0.07) | 1.5E-07 |
| cg14913773 | 10 | 35639680 | *CCNY* | intronic | Island | -0.11(-0.13,-0.08) | 5.8E-13 | -0.05(-0.08,-0.03) | 4.6E-05 |
| cg25570410 | 10 | 119462357 | *EMX2;RAB11FIP2* | intergenic | OpenSea | -0.12(-0.15,-0.09) | 8.8E-13 | -0.03(-0.05,-0.01) | 9.8E-04 |
| cg09372891 | 10 | 43348044 | *BMS1;LINC02623* | intergenic | N_Shelf | -0.16(-0.2,-0.11) | 3.5E-12 | -0.02(-0.04,-0.01) | 2.7E-03 |
| cg18295744 | 10 | 79113169 | *KCNMA1-AS3* | ncRNA_intronic | OpenSea | -0.11(-0.14,-0.08) | 4.7E-12 | 0.12(0.09,0.14) | 8.2E-20 |

| | | | | | | | | | |
|---|---|---|---|---|---|---|---|---|---|
| cg23904955 | 10 | 99523002 | *ZFYVE27;SFRP5* | intergenic | Shelf;S_Shore;Isla | -0.1(-0.14,-0.07) | 6.0E-11 | -0.03(-0.06,0) | 3.8E-02 |
| cg14804686 | 10 | 24618421 | *KIAA1217* | intronic | OpenSea | 0.1(0.07,0.14) | 8.1E-11 | -0.06(-0.08,-0.04) | 4.2E-08 |
| cg08097548 | 10 | 71086998 | *HK1* | intronic | OpenSea | -0.13(-0.17,-0.09) | 5.8E-10 | -0.03(-0.04,-0.01) | 5.3E-05 |
| cg15211694 | 10 | 71975685 | *PPA1* | intronic | OpenSea | -0.1(-0.14,-0.07) | 2.5E-09 | 0.03(0.02,0.04) | 4.5E-08 |
| cg15634806 | 10 | 10872250 | *CELF2* | intronic | OpenSea | -0.13(-0.17,-0.08) | 8.1E-09 | 0.04(0.02,0.06) | 1.3E-03 |
| cg15487644 | 10 | 101597804 | *ABCC2* | intronic | OpenSea | -0.11(-0.15,-0.07) | 1.9E-08 | -0.02(-0.03,-0.01) | 6.1E-07 |
| cg07226162 | 10 | 110533489 | *LINC01435;XPNPEP1* | intergenic | S_Shelf | -0.1(-0.14,-0.07) | 2.2E-08 | -0.05(-0.07,-0.03) | 7.5E-07 |
| cg07748741 | 10 | 97570299 | *ENTPD1-AS1* | ncRNA_intronic | Island | -0.16(-0.22,-0.11) | 2.5E-08 | 0.02(0,0.04) | 5.3E-02 |
| cg11988910 | 10 | 99526781 | *SFRP5* | UTR3 | 3helf;N_Shore;N_S | 0.11(0.07,0.14) | 3.6E-08 | -0.01(-0.02,0) | 1.3E-01 |
| cg02556345 | 10 | 122422449 | *LINC01561;WDR11-AS1* | intergenic | OpenSea | -0.12(-0.17,-0.08) | 6.6E-08 | -0.03(-0.05,-0.01) | 2.0E-03 |
| cg00221747 | 11 | 2131198 | *H19;IGF2* | intergenic | N_Shore | -0.15(-0.17,-0.12) | 8.7E-40 | -0.12(-0.16,-0.09) | 1.1E-12 |
| cg02865822 | 11 | 46361591 | *DGKZ* | intronic | OpenSea | -0.12(-0.14,-0.1) | 7.4E-32 | -0.09(-0.1,-0.07) | 3.1E-23 |
| cg17266233 | 11 | 46361175 | *DGKZ* | intronic | OpenSea | 0.11(0.09,0.13) | 2.7E-31 | -0.07(-0.09,-0.06) | 3.4E-19 |
| cg01668279 | 11 | 2135469 | *H19;IGF2* | intergenic | S_Shelf;N_Shelf | -0.12(-0.14,-0.1) | 2.2E-28 | -0.11(-0.13,-0.08) | 2.9E-16 |
| cg21370660 | 11 | 45249998 | *PRDM11* | UTR3 | OpenSea | -0.11(-0.14,-0.09) | 3.1E-26 | 0.06(0.05,0.08) | 3.5E-13 |
| cg08849509 | 11 | 9815661 | *SBF2-AS1* | ncRNA_intronic | OpenSea | -0.13(-0.16,-0.11) | 9.1E-26 | 0.11(0.08,0.15) | 1.7E-09 |
| cg27605998 | 11 | 46337661 | *CREB3L1* | intronic | OpenSea | -0.12(-0.15,-0.1) | 1.8E-25 | -0.1(-0.14,-0.06) | 1.7E-07 |
| cg17084793 | 11 | 129950381 | *APLP2* | intronic | OpenSea | 0.11(0.09,0.13) | 1.4E-23 | -0.08(-0.12,-0.05) | 8.3E-07 |
| cg16619139 | 11 | 46361092 | *DGKZ* | intronic | OpenSea | 0.11(0.09,0.13) | 2.0E-23 | -0.08(-0.11,-0.06) | 3.2E-13 |
| cg13048008 | 11 | 72014294 | *CLPB* | intronic | OpenSea | -0.11(-0.13,-0.08) | 2.2E-21 | 0.08(0.06,0.09) | 9.4E-24 |
| cg21017775 | 11 | 34513934 | *ELF5* | intronic | OpenSea | 0.14(0.11,0.17) | 3.9E-21 | -0.07(-0.09,-0.04) | 9.5E-08 |
| cg05205239 | 11 | 69249972 | *LOC102724265;LINC01488* | intergenic | OpenSea | -0.12(-0.15,-0.09) | 5.7E-18 | -0.09(-0.12,-0.06) | 7.1E-10 |
| cg22705929 | 11 | 57650501 | *CTNND1;OR9Q1* | intergenic | S_Shelf | -0.18(-0.23,-0.14) | 3.4E-17 | -0.06(-0.08,-0.04) | 5.7E-09 |
| cg07657954 | 11 | 75597838 | *UVRAG* | intronic | OpenSea | -0.11(-0.14,-0.08) | 1.1E-16 | -0.01(-0.02,0) | 6.4E-03 |
| cg18068443 | 11 | 69966008 | *ANO1* | intronic | N_Shore | -0.12(-0.15,-0.09) | 2.0E-16 | -0.03(-0.05,-0.02) | 1.8E-06 |
| cg08900426 | 11 | 75421424 | *MAP6;MOGAT2* | intergenic | N_Shelf | -0.13(-0.16,-0.1) | 2.2E-16 | -0.03(-0.04,-0.01) | 3.4E-04 |
| cg23657589 | 11 | 34526126 | *ELF5* | intronic | OpenSea | 0.13(0.1,0.16) | 2.9E-16 | -0.05(-0.07,-0.03) | 3.5E-09 |
| cg02723558 | 11 | 27514526 | *LOC105376671;LIN7C* | intergenic | OpenSea | -0.11(-0.14,-0.09) | 4.7E-16 | 0.17(0.13,0.21) | 8.2E-17 |
| cg13037081 | 11 | 64368403 | *SLC22A12* | exonic | Island;N_Shore | 0.15(0.11,0.19) | 4.9E-15 | -0.04(-0.05,-0.02) | 2.4E-06 |
| cg01444191 | 11 | 72014252 | *CLPB* | intronic | OpenSea | -0.1(-0.13,-0.08) | 5.7E-15 | 0.06(0.05,0.08) | 5.3E-20 |
| cg19944895 | 11 | 62914623 | *SLC22A24;SLC22A25* | intergenic | OpenSea | 0.11(0.08,0.14) | 2.3E-14 | -0.12(-0.15,-0.09) | 3.5E-15 |
| cg16609998 | 11 | 73157989 | *FAM168A* | intronic | OpenSea | 0.12(0.09,0.15) | 6.3E-14 | -0.05(-0.07,-0.04) | 1.1E-12 |
| cg16664812 | 11 | 78044069 | *GAB2* | intronic | OpenSea | 0.15(0.11,0.19) | 8.2E-14 | -0.02(-0.03,-0.01) | 1.9E-06 |
| cg23045350 | 11 | 75734574 | *UVRAG* | intronic | OpenSea | -0.1(-0.13,-0.07) | 9.7E-14 | -0.01(-0.02,0) | 7.1E-03 |
| cg17632226 | 11 | 94736072 | *KDM4D;KDM4E* | intergenic | OpenSea | -0.11(-0.14,-0.08) | 1.1E-13 | -0.04(-0.05,-0.02) | 9.8E-08 |
| cg06767142 | 11 | 64053928 | *GPR137* | intron | OpenSea | -0.12(-0.15,-0.08) | 2.9E-13 | -0.05(-0.07,-0.03) | 4.5E-08 |
| cg16073483 | 11 | 17765869 | *KCNC1* | intronic | OpenSea | -0.13(-0.16,-0.09) | 5.4E-13 | -0.03(-0.05,-0.02) | 7.9E-07 |

| | | | | | | | | | |
|---|---|---|---|---|---|---|---|---|---|
| cg19352801 | 11 | 64204294 | *RPS6KA4;LINC02724* | intergenic | N_Shelf | -0.13(-0.17,-0.09) | 8.2E-13 | -0.03(-0.04,-0.02) | 1.9E-06 |
| cg15451948 | 11 | 778638 | *LOC171391* | ncRNA_intronic | S_Shore | -0.12(-0.15,-0.08) | 2.9E-12 | -0.03(-0.04,-0.02) | 1.1E-06 |
| cg17667688 | 11 | 2474627 | *KCNQ1* | intronic | OpenSea | -0.12(-0.15,-0.08) | 4.8E-12 | -0.04(-0.05,-0.02) | 2.7E-08 |
| cg10978403 | 11 | 64102498 | *PRDX5;CCDC88B* | intergenic | OpenSea | -0.12(-0.16,-0.09) | 7.8E-12 | -0.05(-0.06,-0.03) | 2.7E-09 |
| cg16393295 | 11 | 60881089 | *CD5* | intronic | OpenSea | 0.12(0.08,0.15) | 7.2E-11 | -0.02(-0.03,-0.01) | 6.5E-04 |
| cg07606115 | 11 | 130198280 | *ZBTB44-DT* | ncRNA_intronic | OpenSea | 0.12(0.09,0.16) | 8.5E-11 | -0.06(-0.08,-0.05) | 1.1E-14 |
| cg16481602 | 11 | 65775676 | *BANF1;CST6* | intergenic | OpenSea | 0.13(0.09,0.17) | 8.9E-11 | -0.05(-0.07,-0.03) | 4.9E-07 |
| cg11490276 | 11 | 67675226 | *FAM86C2P;UNC93B1* | intergenic | OpenSea | 0.11(0.07,0.14) | 1.6E-10 | -0.01(-0.02,0) | 1.7E-02 |
| cg02828306 | 11 | 44973136 | *TP53I11* | upstream | OpenSea | 0.11(0.07,0.14) | 2.5E-10 | -0.04(-0.05,-0.02) | 2.2E-05 |
| cg16309752 | 11 | 47857727 | *NUP160* | intronic | OpenSea | -0.12(-0.16,-0.08) | 5.4E-10 | 0.03(0.01,0.06) | 2.2E-03 |
| cg18994049 | 11 | 69967719 | *ANO1* | intronic | S_Shore | -0.14(-0.19,-0.1) | 6.1E-10 | -0.04(-0.06,-0.03) | 7.3E-09 |
| cg13777907 | 11 | 16299989 | *SOX6* | intronic | OpenSea | 0.14(0.09,0.18) | 3.7E-09 | -0.02(-0.04,0) | 1.3E-01 |
| cg01599099 | 11 | 66882361 | *LOC107984341* | ncRNA_intronic | OpenSea | -0.12(-0.17,-0.08) | 5.8E-09 | -0.02(-0.03,0) | 5.3E-02 |
| cg17019994 | 11 | 123461213 | *GRAMD1B* | intronic | OpenSea | 0.12(0.08,0.16) | 1.3E-08 | 0.03(0.02,0.05) | 2.4E-04 |
| cg22698115 | 11 | 1750677 | *KRTAP5-6;IFITM10* | intergenic | S_Shore;N_Shelf | -0.11(-0.15,-0.07) | 1.4E-08 | 0.08(0.04,0.11) | 6.5E-06 |
| cg16965598 | 11 | 118878300 | *CCDC84* | intronic | OpenSea | 0.11(0.07,0.15) | 2.5E-08 | -0.01(-0.02,0.01) | 4.2E-01 |
| cg16534985 | 11 | 68436331 | *PPP6R3;GAL* | intergenic | S_Shelf;N_Shelf | -0.11(-0.15,-0.07) | 4.6E-08 | -0.01(-0.02,0) | 1.2E-02 |
| cg13955644 | 11 | 61041918 | *VWCE* | intronic | OpenSea | 0.11(0.07,0.15) | 5.4E-08 | -0.02(-0.04,-0.01) | 9.2E-05 |
| cg16745933 | 11 | 59184950 | *OR5AN1;OR5A2* | intergenic | OpenSea | -0.12(-0.16,-0.07) | 6.9E-08 | -0.01(-0.02,0) | 9.8E-03 |
| cg25383629 | 12 | 109829791 | *MYO1H* | intronic | N_Shelf | 0.11(0.09,0.14) | 2.3E-23 | -0.03(-0.04,-0.02) | 4.6E-09 |
| cg05616242 | 12 | 54389632 | *HOXC-AS2* | ncRNA_exonic | OpenSea | -0.11(-0.13,-0.08) | 4.0E-20 | 0.1(0.08,0.12) | 9.8E-16 |
| cg10369363 | 12 | 52010638 | *SCN8A* | intronic | S_Shelf;N_Shelf | 0.11(0.09,0.14) | 1.4E-16 | -0.03(-0.04,-0.01) | 1.9E-04 |
| cg05850869 | 12 | 132731778 | *GALNT9* | intronic | N_Shore | -0.11(-0.14,-0.08) | 1.6E-16 | -0.06(-0.07,-0.04) | 6.2E-08 |
| cg08888956 | 12 | 85874061 | *ALX1;RASSF9* | intergenic | OpenSea | -0.1(-0.13,-0.08) | 1.5E-15 | -0.01(-0.03,0) | 1.4E-02 |
| cg25289862 | 12 | 551949 | *CCDC77* | downstream | OpenSea | 0.11(0.09,0.14) | 1.0E-14 | 0.1(0.07,0.12) | 6.2E-13 |
| cg14391474 | 12 | 123079025 | *KNTC1* | intronic | OpenSea | -0.13(-0.17,-0.1) | 2.3E-14 | -0.02(-0.03,-0.01) | 9.8E-07 |
| cg08823894 | 12 | 107774192 | *BTBD11* | intronic | N_Shore | 0.11(0.08,0.14) | 7.4E-14 | -0.03(-0.04,-0.01) | 2.8E-03 |
| cg15318882 | 12 | 116978139 | *LINC00173;MAP1LC3B2* | intergenic | OpenSea | 0.11(0.08,0.14) | 8.0E-14 | -0.03(-0.04,-0.02) | 4.6E-08 |
| cg20924966 | 12 | 52010546 | *SCN8A* | intronic | S_Shelf | 0.12(0.09,0.15) | 1.5E-13 | -0.02(-0.04,-0.01) | 7.6E-05 |
| cg18030623 | 12 | 106205593 | *CASC18;NUAK1* | intergenic | OpenSea | -0.11(-0.14,-0.08) | 1.8E-13 | 0.08(0.06,0.1) | 1.1E-11 |
| cg06661904 | 12 | 120937300 | *DYNLL1;COQ5* | intergenic | OpenSea | -0.1(-0.13,-0.07) | 2.6E-13 | -0.07(-0.09,-0.05) | 9.5E-13 |
| cg02172262 | 12 | 57894274 | *MARS* | exonic | N_Shelf | -0.11(-0.14,-0.08) | 4.6E-13 | -0.1(-0.12,-0.07) | 6.1E-14 |
| cg13545247 | 12 | 52662533 | *KRT87P;KRT86* | intergenic | OpenSea | -0.11(-0.14,-0.08) | 6.7E-13 | -0.03(-0.04,-0.01) | 5.4E-05 |
| cg06130893 | 12 | 113333083 | *RPH3A* | intronic | N_Shore | -0.11(-0.13,-0.08) | 1.2E-12 | -0.06(-0.08,-0.05) | 2.1E-11 |
| cg17603826 | 12 | 51013725 | *DIP2B* | intronic | OpenSea | -0.13(-0.17,-0.1) | 2.4E-12 | 0.12(0.09,0.15) | 4.9E-15 |
| cg08333564 | 12 | 4278628 | *PARP11;CCND2-AS1* | intergenic | S_Shelf | 0.1(0.07,0.13) | 3.3E-12 | -0.05(-0.08,-0.03) | 4.1E-07 |
| cg21703255 | 12 | 106145231 | *CASC18;NUAK1* | intergenic | OpenSea | 0.13(0.09,0.17) | 1.0E-11 | -0.01(-0.03,0) | 2.5E-02 |

| | | | | | | | | | |
|---|---|---|---|---|---|---|---|---|---|
| cg14008271 | 12 | 68452005 | *IFNG-AS1;IFNG* | intergenic | OpenSea | -0.1(-0.13,-0.07) | 2.7E-11 | 0.09(0.06,0.11) | 3.3E-10 |
| cg10553109 | 12 | 115088900 | *TBX5-AS1;TBX3* | intergenic | OpenSea | 0.11(0.08,0.14) | 3.2E-11 | 0.04(0.03,0.06) | 6.0E-11 |
| cg12157156 | 12 | 124395627 | *DNAH10* | intronic | OpenSea | -0.1(-0.13,-0.07) | 5.2E-11 | -0.02(-0.03,0) | 1.7E-02 |
| cg05268743 | 12 | 93563962 | *LOC643339* | ncRNA_intronic | OpenSea | -0.1(-0.13,-0.07) | 9.9E-11 | 0.11(0.07,0.14) | 2.2E-11 |
| cg26641022 | 12 | 124516221 | *ZNF664-RFLNA* | intronic | Shore;N_Shelf;N_S | 0.11(0.07,0.14) | 1.4E-10 | 0.05(0.03,0.08) | 3.6E-05 |
| cg02192520 | 12 | 55720371 | *OR6C1;OR6C3* | intergenic | OpenSea | 0.11(0.07,0.14) | 5.1E-10 | 0.1(0.06,0.14) | 1.2E-06 |
| cg20086252 | 12 | 124535851 | *ZNF664-RFLNA* | intronic | OpenSea | -0.1(-0.13,-0.07) | 9.7E-10 | -0.03(-0.04,-0.02) | 4.5E-07 |
| cg08035822 | 12 | 121034167 | *POP5;CABP1* | intergenic | OpenSea | 0.17(0.11,0.23) | 4.9E-09 | 0.03(0.02,0.04) | 4.8E-10 |
| cg03454021 | 12 | 115433412 | *TBX3;MED13L* | intergenic | OpenSea | -0.11(-0.15,-0.07) | 6.0E-09 | -0.04(-0.06,-0.03) | 2.2E-07 |
| cg04404543 | 12 | 53625523 | *RARG* | intronic | N_Shore | -0.1(-0.14,-0.07) | 2.8E-08 | 0.03(0.02,0.04) | 9.3E-09 |
| cg07974084 | 12 | 104910071 | *CHST11* | intronic | OpenSea | 0.12(0.08,0.16) | 3.0E-08 | 0.01(-0.01,0.04) | 2.1E-01 |
| cg22049450 | 12 | 49956168 | *MCRS1* | intronic | S_Shore | -0.11(-0.15,-0.07) | 6.5E-08 | -0.03(-0.04,-0.01) | 6.2E-06 |
| cg09878710 | 13 | 94661199 | *GPC6* | intronic | OpenSea | 0.14(0.12,0.17) | 3.8E-26 | -0.06(-0.08,-0.04) | 2.5E-07 |
| cg15280369 | 13 | 95122354 | *DCT* | intronic | OpenSea | -0.14(-0.17,-0.11) | 9.8E-21 | -0.04(-0.05,-0.02) | 4.4E-05 |
| cg18523054 | 13 | 25753183 | *LINC00463* | ncRNA_intronic | OpenSea | 0.12(0.09,0.14) | 4.0E-20 | 0.07(0.05,0.09) | 3.2E-09 |
| cg18736682 | 13 | 50335014 | *KPNA3* | intronic | OpenSea | -0.13(-0.15,-0.1) | 9.2E-19 | -0.11(-0.15,-0.07) | 1.1E-08 |
| cg01268721 | 13 | 95123757 | *DCT* | intronic | OpenSea | -0.15(-0.19,-0.12) | 5.4E-18 | -0.04(-0.05,-0.02) | 1.7E-09 |
| cg09253641 | 13 | 44732742 | *SMIM2* | intronic | OpenSea | -0.1(-0.12,-0.08) | 1.0E-16 | -0.03(-0.04,-0.02) | 2.5E-06 |
| cg05231180 | 13 | 73704606 | *KLF5;LINC00392* | intergenic | OpenSea | 0.1(0.08,0.13) | 2.8E-16 | -0.02(-0.03,-0.01) | 1.6E-04 |
| cg14592487 | 13 | 94433792 | *GPC6* | intronic | OpenSea | -0.16(-0.21,-0.12) | 1.1E-13 | -0.06(-0.07,-0.04) | 1.5E-09 |
| cg03715269 | 13 | 94433198 | *GPC6* | intronic | OpenSea | -0.11(-0.14,-0.08) | 4.2E-12 | -0.1(-0.13,-0.07) | 5.9E-14 |
| cg04067341 | 13 | 110508360 | *IRS2;LINC00396* | intergenic | N_Shelf | 0.11(0.08,0.14) | 6.7E-12 | 0.12(0.09,0.16) | 6.9E-12 |
| cg21957311 | 13 | 45049196 | *TSC22D1* | intronic | OpenSea | -0.12(-0.15,-0.08) | 2.0E-11 | -0.05(-0.07,-0.03) | 3.1E-07 |
| cg26881318 | 13 | 95067415 | *GPC6;DCT* | intergenic | OpenSea | -0.12(-0.16,-0.08) | 6.0E-10 | -0.02(-0.03,-0.01) | 6.5E-06 |
| cg18916584 | 13 | 98840580 | *FARP1* | intronic | OpenSea | -0.11(-0.15,-0.08) | 1.3E-09 | -0.03(-0.04,-0.02) | 6.3E-06 |
| cg08409341 | 13 | 44816023 | *MIR8079;SERP2* | intergenic | OpenSea | -0.12(-0.15,-0.08) | 1.7E-09 | -0.04(-0.06,-0.03) | 2.3E-06 |
| cg18878787 | 13 | 73173561 | *DACH1;MZT1* | intergenic | OpenSea | -0.11(-0.15,-0.07) | 1.1E-08 | -0.09(-0.12,-0.05) | 1.9E-07 |
| cg16943230 | 14 | 91466411 | *RPS6KA5* | intronic | OpenSea | 0.21(0.18,0.25) | 7.9E-38 | 0.14(0.1,0.18) | 3.2E-11 |
| cg06285241 | 14 | 105478318 | *CDCA4* | intronic | S_Shelf;Island | -0.11(-0.13,-0.09) | 2.0E-27 | -0.06(-0.08,-0.04) | 2.6E-08 |
| cg18107268 | 14 | 69095862 | *RAD51B* | intronic | OpenSea | -0.13(-0.16,-0.1) | 2.6E-17 | -0.01(-0.03,0) | 2.2E-02 |
| cg01063461 | 14 | 100156476 | *CYP46A1* | intronic | N_Shelf | -0.12(-0.15,-0.09) | 9.9E-17 | 0.02(0.01,0.03) | 1.5E-04 |
| cg05268769 | 14 | 75267229 | *YLPM1* | intronic | OpenSea | 0.1(0.08,0.12) | 1.0E-16 | -0.07(-0.08,-0.05) | 7.2E-19 |
| cg06513971 | 14 | 64956575 | *ZBTB25* | intronic | OpenSea | -0.11(-0.14,-0.08) | 3.0E-14 | 0.08(0.06,0.11) | 1.0E-11 |
| cg20061948 | 14 | 105466145 | *CLBA1;CDCA4* | intergenic | hore;N_Shore;N_S | 0.11(0.08,0.14) | 5.4E-12 | -0.03(-0.04,-0.02) | 8.6E-08 |
| cg09013732 | 14 | 100406858 | *EML1* | UTR3 | OpenSea | -0.13(-0.17,-0.09) | 6.6E-12 | -0.04(-0.05,-0.02) | 2.3E-08 |
| cg01611656 | 14 | 100779378 | *MIR345;SLC25A47* | intergenic | OpenSea | -0.12(-0.15,-0.08) | 2.7E-11 | 0.02(0.01,0.04) | 5.1E-05 |
| cg26954761 | 14 | 81450355 | *TSHR* | intronic | OpenSea | 0.11(0.08,0.14) | 7.4E-11 | -0.07(-0.09,-0.05) | 6.8E-14 |

| | | | | | | | | | |
|---|---|---|---|---|---|---|---|---|---|
| cg03620691 | 14 | 102064158 | *DIO3;LINC00239* | intergenic | OpenSea | -0.11(-0.14,-0.08) | 1.1E-10 | -0.02(-0.03,-0.01) | 4.7E-03 |
| cg24381739 | 14 | 100779291 | *MIR345;SLC25A47* | intergenic | OpenSea | -0.11(-0.15,-0.08) | 2.4E-10 | 0.03(0.02,0.05) | 4.3E-05 |
| cg17306116 | 14 | 77061851 | *ESRRB;VASH1* | intergenic | OpenSea | -0.12(-0.16,-0.08) | 3.2E-10 | -0.04(-0.05,-0.03) | 4.3E-11 |
| cg04316096 | 14 | 81417251 | *CEP128;TSHR* | intergenic | OpenSea | 0.15(0.1,0.19) | 4.9E-10 | -0.05(-0.07,-0.04) | 5.4E-10 |
| cg02057912 | 14 | 75980090 | *JDP2;BATF* | intergenic | N_Shelf | -0.1(-0.13,-0.07) | 1.1E-09 | -0.03(-0.04,-0.01) | 6.6E-06 |
| cg20354873 | 14 | 61602312 | *SLC38A6;TMEM30B* | intergenic | OpenSea | 0.1(0.07,0.13) | 5.4E-09 | -0.01(-0.02,0) | 2.3E-02 |
| cg13643452 | 14 | 38256445 | *TTC6* | intronic | S_Shore | -0.13(-0.17,-0.09) | 1.2E-08 | -0.05(-0.07,-0.03) | 3.0E-06 |
| cg20905794 | 15 | 101015481 | *CERS3* | intronic | OpenSea | 0.16(0.13,0.19) | 3.2E-23 | -0.03(-0.04,-0.01) | 3.0E-04 |
| cg20558585 | 15 | 68621047 | *ITGA11* | intronic | OpenSea | 0.12(0.1,0.15) | 5.7E-21 | -0.04(-0.06,-0.03) | 1.3E-10 |
| cg19348484 | 15 | 90870006 | *NGRN;GABARAPL3* | intergenic | N_Shore;N_Shore | 0.12(0.09,0.15) | 3.3E-15 | -0.06(-0.07,-0.04) | 1.9E-17 |
| cg20818326 | 15 | 92634719 | *SLCO3A1* | intronic | OpenSea | -0.1(-0.13,-0.08) | 4.2E-15 | -0.07(-0.1,-0.05) | 1.3E-08 |
| cg11398451 | 15 | 90592837 | *ZNF710* | intronic | OpenSea | 0.18(0.13,0.22) | 3.3E-14 | -0.12(-0.17,-0.07) | 8.8E-07 |
| cg04673385 | 15 | 40338170 | *SRP14-AS1* | ncRNA_exonic | OpenSea | 0.11(0.08,0.14) | 4.7E-13 | -0.05(-0.07,-0.04) | 1.1E-10 |
| cg00409842 | 15 | 85532879 | *PDE8A* | intronic | OpenSea | -0.11(-0.14,-0.08) | 1.8E-12 | 0.1(0.06,0.14) | 4.0E-07 |
| cg20695941 | 15 | 80844790 | *ARNT2* | intronic | OpenSea | 0.13(0.1,0.17) | 7.0E-12 | -0.04(-0.06,-0.02) | 3.3E-04 |
| cg20570688 | 15 | 69730833 | *KIF23* | intronic | OpenSea | -0.17(-0.22,-0.12) | 1.7E-11 | -0.08(-0.11,-0.05) | 2.0E-09 |
| cg25432637 | 15 | 63382250 | *TPM1;LACTB* | intergenic | S_Shore | -0.1(-0.13,-0.07) | 2.3E-11 | 0.04(0.02,0.06) | 2.1E-04 |
| cg01152374 | 15 | 69524732 | *GLCE* | intronic | OpenSea | -0.11(-0.14,-0.08) | 3.1E-11 | -0.03(-0.04,-0.01) | 1.9E-05 |
| cg07146799 | 15 | 36978429 | *C15orf41* | intronic | OpenSea | 0.11(0.07,0.14) | 4.4E-11 | 0.03(0.01,0.06) | 3.6E-03 |
| cg21857123 | 15 | 77557398 | *PEAK1* | intronic | OpenSea | -0.14(-0.18,-0.1) | 3.3E-10 | -0.02(-0.03,-0.01) | 3.4E-03 |
| cg12915526 | 15 | 61049160 | *RORA* | intronic | OpenSea | -0.17(-0.22,-0.11) | 3.4E-10 | -0.03(-0.05,-0.01) | 2.9E-04 |
| cg08280562 | 15 | 68365319 | *PIAS1* | intronic | OpenSea | 0.12(0.08,0.16) | 6.7E-10 | 0.01(0,0.02) | 4.9E-03 |
| cg19143990 | 15 | 51736955 | *GLDN;DMXL2* | intergenic | N_Shore | 0.11(0.07,0.14) | 3.0E-09 | 0.02(0.01,0.03) | 1.2E-03 |
| cg20277551 | 15 | 40388969 | *BMF* | intronic | OpenSea | -0.14(-0.2,-0.09) | 3.7E-08 | -0.05(-0.07,-0.03) | 1.9E-06 |
| cg19752649 | 16 | 11114847 | *CLEC16A* | intronic | OpenSea | 0.12(0.1,0.15) | 7.0E-28 | -0.09(-0.12,-0.06) | 1.5E-10 |
| cg03185144 | 16 | 85072884 | *KIAA0513* | intronic | OpenSea | -0.13(-0.15,-0.1) | 2.6E-24 | 0.06(0.03,0.09) | 1.4E-04 |
| cg21320605 | 16 | 27560894 | *GTF3C1* | intronic | OpenSea | -0.14(-0.16,-0.11) | 8.1E-24 | 0.11(0.08,0.14) | 3.8E-14 |
| cg21694119 | 16 | 70753596 | *VAC14* | intronic | OpenSea | 0.15(0.12,0.17) | 2.3E-23 | 0.08(0.06,0.1) | 2.1E-14 |
| cg14994009 | 16 | 53910160 | *FTO* | intronic | OpenSea | 0.14(0.11,0.18) | 2.7E-20 | -0.05(-0.07,-0.04) | 9.5E-10 |
| cg21854374 | 16 | 84737742 | *USP10* | intronic | OpenSea | 0.12(0.1,0.15) | 6.9E-20 | 0.13(0.09,0.16) | 3.3E-15 |
| cg15547825 | 16 | 85485572 | *MIR5093;GSE1* | intergenic | Island | -0.1(-0.12,-0.08) | 1.1E-18 | -0.06(-0.08,-0.04) | 4.1E-09 |
| cg01948724 | 16 | 2988034 | *FLYWCH1* | intronic | OpenSea | 0.12(0.1,0.15) | 1.6E-18 | -0.07(-0.08,-0.05) | 3.5E-13 |
| cg10259077 | 16 | 456958 | *DECR2* | intronic | OpenSea | -0.11(-0.13,-0.08) | 5.9E-17 | -0.02(-0.04,0) | 2.5E-02 |
| cg06670039 | 16 | 12436389 | *SNX29* | intronic | OpenSea | 0.15(0.12,0.19) | 2.7E-16 | -0.07(-0.1,-0.04) | 1.7E-05 |
| cg14105153 | 16 | 66960132 | *RRAD* | upstream | OpenSea | -0.1(-0.13,-0.08) | 4.4E-15 | 0.07(0.05,0.09) | 6.6E-11 |
| cg20744302 | 16 | 58678958 | *CNOT1;SLC38A7* | intergenic | OpenSea | -0.11(-0.13,-0.08) | 2.8E-14 | 0.09(0.07,0.1) | 1.6E-25 |
| cg21139396 | 16 | 11597814 | *LOC400499* | intronic | OpenSea | -0.1(-0.13,-0.07) | 1.7E-12 | -0.07(-0.09,-0.05) | 2.9E-12 |

| | | | | | | | | | |
|---|---|---|---|---|---|---|---|---|---|
| cg14246480 | 16 | 50249047 | *TENT4B* | intronic | S_Shelf | -0.13(-0.16,-0.09) | 1.8E-12 | -0.05(-0.06,-0.03) | 9.7E-07 |
| cg03091585 | 16 | 53129622 | *CHD9* | intronic | N_Shore | -0.14(-0.18,-0.1) | 2.8E-12 | -0.05(-0.07,-0.02) | 8.4E-05 |
| cg00869550 | 16 | 58012633 | *TEPP* | intronic | OpenSea | -0.17(-0.21,-0.12) | 3.2E-12 | -0.03(-0.04,-0.01) | 4.2E-05 |
| cg07265549 | 16 | 27595697 | *KIAA0556* | intronic | OpenSea | -0.13(-0.17,-0.09) | 8.7E-11 | -0.02(-0.02,-0.01) | 4.2E-04 |
| cg06890610 | 16 | 12436257 | *SNX29* | intronic | OpenSea | -0.11(-0.14,-0.08) | 1.1E-10 | -0.03(-0.05,-0.02) | 4.0E-05 |
| cg21972289 | 16 | 89607687 | *SPG7* | intronic | OpenSea | -0.11(-0.14,-0.07) | 1.6E-10 | -0.04(-0.06,-0.02) | 6.4E-05 |
| cg07846061 | 16 | 74698536 | *RFWD3* | intron | N_Shore | -0.11(-0.15,-0.08) | 2.1E-10 | 0.08(0.05,0.1) | 1.3E-09 |
| cg06111946 | 16 | 70713936 | *MTSS2* | exonic | OpenSea | 0.1(0.07,0.13) | 3.0E-09 | -0.04(-0.05,-0.02) | 1.5E-05 |
| cg08449748 | 16 | 2299845 | *ECI1* | intronic | S_Shelf | -0.11(-0.15,-0.08) | 3.1E-09 | -0.03(-0.04,-0.02) | 2.6E-06 |
| cg21713033 | 16 | 72723868 | *LINC01572;ZFHX3* | intergenic | OpenSea | -0.11(-0.15,-0.08) | 7.1E-09 | 0.05(0.02,0.07) | 1.3E-04 |
| cg21678275 | 16 | 69840414 | *WWP2* | intronic | OpenSea | 0.15(0.1,0.2) | 1.1E-08 | 0.04(0.02,0.06) | 4.8E-05 |
| cg09681300 | 16 | 72896301 | *ZFHX3* | intronic | OpenSea | -0.1(-0.14,-0.07) | 1.3E-08 | -0.01(-0.02,-0.01) | 9.9E-04 |
| cg00810262 | 17 | 50673068 | *LINC01982;LINC02089* | intergenic | OpenSea | -0.13(-0.16,-0.11) | 1.1E-26 | 0.05(0.03,0.07) | 6.1E-10 |
| cg22656263 | 17 | 49371848 | *UTP18* | intronic | OpenSea | -0.16(-0.19,-0.13) | 1.8E-25 | -0.03(-0.04,-0.02) | 2.6E-06 |
| cg11477010 | 17 | 4175967 | *UBE2G1* | intronic | OpenSea | 0.11(0.08,0.13) | 5.6E-21 | -0.03(-0.04,-0.02) | 3.2E-08 |
| cg22357003 | 17 | 27319874 | *SEZ6* | intronic | OpenSea | -0.12(-0.14,-0.09) | 1.5E-18 | 0.06(0.05,0.08) | 4.3E-12 |
| cg15119221 | 17 | 19387442 | *RNF112;SLC47A1* | intergenic | OpenSea | 0.12(0.09,0.14) | 3.8E-17 | -0.04(-0.05,-0.02) | 3.4E-06 |
| cg03662398 | 17 | 55335467 | *MSI2* | intronic | OpenSea | -0.12(-0.14,-0.09) | 7.1E-17 | -0.07(-0.1,-0.04) | 1.7E-06 |
| cg26867637 | 17 | 1079155 | *ABR* | intronic | S_Shore | -0.1(-0.13,-0.08) | 3.6E-16 | -0.08(-0.09,-0.06) | 3.9E-17 |
| cg20404716 | 17 | 45305937 | *MYL4;ITGB3* | intergenic | OpenSea | 0.12(0.09,0.15) | 4.1E-16 | 0.04(0.02,0.05) | 1.0E-08 |
| cg04193923 | 17 | 40014647 | *KLHL11* | intronic | OpenSea | -0.1(-0.13,-0.08) | 1.1E-15 | -0.02(-0.03,-0.01) | 9.5E-04 |
| cg22991344 | 17 | 77329488 | *RBFOX3* | intronic | OpenSea | -0.15(-0.18,-0.11) | 1.3E-15 | 0.05(0.03,0.07) | 5.7E-07 |
| cg22842221 | 17 | 66715668 | *LINC01482;ABCA8* | intergenic | OpenSea | -0.15(-0.19,-0.11) | 1.2E-14 | 0.09(0.07,0.11) | 3.1E-15 |
| cg16824480 | 17 | 76193850 | *AFMID* | intronic | OpenSea | 0.11(0.08,0.13) | 5.5E-14 | -0.02(-0.03,0) | 1.0E-02 |
| cg22578231 | 17 | 44098209 | *MAPT* | intronic | OpenSea | 0.12(0.09,0.15) | 1.6E-13 | 0.07(0.05,0.08) | 2.5E-18 |
| cg01989275 | 17 | 82409151 | *RPL23AP87;NONE* | intergenic | Island | -0.12(-0.16,-0.09) | 2.7E-13 | -0.03(-0.04,-0.02) | 8.1E-10 |
| cg06186567 | 17 | 39995017 | *KLHL10* | intronic | OpenSea | -0.12(-0.16,-0.09) | 7.3E-13 | -0.04(-0.05,-0.02) | 1.9E-06 |
| cg15741102 | 17 | 2032493 | *SMG6* | intronic | S_Shore | 0.11(0.08,0.15) | 1.5E-12 | -0.01(-0.02,0) | 1.3E-01 |
| cg14177325 | 17 | 1638469 | *WDR81* | intronic | S_Shelf | -0.14(-0.18,-0.1) | 1.8E-12 | -0.07(-0.08,-0.05) | 1.2E-12 |
| cg00819441 | 17 | 78385856 | *RNF213-AS1* | ncRNA_intronic | OpenSea | -0.1(-0.13,-0.07) | 2.3E-12 | -0.08(-0.1,-0.05) | 5.7E-08 |
| cg07337480 | 17 | 41425660 | *CCDC200;LINC00910* | intergenic | OpenSea | -0.11(-0.13,-0.08) | 4.5E-12 | -0.04(-0.08,0) | 3.6E-02 |
| cg22991768 | 17 | 77356236 | *RBFOX3* | intronic | OpenSea | 0.13(0.09,0.17) | 7.8E-12 | 0.05(0.03,0.07) | 1.0E-06 |
| cg23122516 | 17 | 82611811 | *RPL23AP87;NONE* | intergenic | N_Shore | 0.11(0.08,0.15) | 2.2E-11 | -0.03(-0.04,-0.01) | 1.0E-04 |
| cg10992906 | 17 | 2032437 | *SMG6* | intronic | S_Shore | -0.12(-0.16,-0.08) | 1.1E-10 | -0.01(-0.02,0) | 2.7E-02 |
| cg25930644 | 17 | 8628597 | *MYH10;CCDC42* | intergenic | N_Shore | -0.13(-0.17,-0.09) | 2.0E-10 | -0.03(-0.04,-0.01) | 1.4E-04 |
| cg00897875 | 17 | 45314799 | *MYL4;ITGB3* | intergenic | N_Shore | -0.12(-0.16,-0.08) | 2.8E-10 | 0.03(0.01,0.05) | 6.5E-04 |
| cg23075265 | 17 | 81082880 | *METRNL;RPL23AP87* | intergenic | OpenSea | -0.1(-0.13,-0.07) | 3.5E-10 | 0.08(0.06,0.1) | 3.7E-15 |

| | | | | | | | | | |
|---|---|---|---|---|---|---|---|---|---|
| cg16076775 | 17 | 75250442 | *SEC14L1;SEPTIN9-DT* | intergenic | OpenSea | 0.11(0.08,0.15) | 8.0E-10 | -0.04(-0.06,-0.02) | 3.8E-05 |
| cg14428628 | 17 | 79833123 | *ARHGDIA;ALYREF* | intergenic | Island | -0.11(-0.14,-0.07) | 8.9E-10 | -0.03(-0.05,-0.01) | 1.5E-03 |
| cg06417243 | 17 | 73546973 | *LLGL2* | intronic | OpenSea | -0.11(-0.15,-0.08) | 1.2E-09 | 0.05(0.03,0.08) | 6.9E-06 |
| cg23006909 | 17 | 78179066 | *CARD14* | intronic | OpenSea | -0.1(-0.14,-0.07) | 3.9E-09 | -0.02(-0.03,0) | 6.5E-03 |
| cg22513981 | 17 | 39924293 | *JUP* | intronic | N_Shelf | 0.14(0.09,0.19) | 4.2E-09 | -0.01(-0.02,0) | 5.4E-03 |
| cg22994495 | 17 | 80688980 | *FN3KRP;FN3K* | intergenic | OpenSea | 0.11(0.07,0.15) | 1.0E-08 | -0.03(-0.05,-0.01) | 1.8E-04 |
| cg05669282 | 17 | 72137029 | *LINC00469;RPL38* | intergenic | OpenSea | -0.17(-0.23,-0.11) | 1.1E-08 | 0.04(0.01,0.08) | 2.4E-02 |
| cg09720939 | 17 | 82892906 | *RPL23AP87;NONE* | intergenic | OpenSea | 0.13(0.08,0.17) | 1.3E-08 | -0.03(-0.05,-0.01) | 5.5E-03 |
| cg07464291 | 17 | 40015202 | *KLHL11* | intronic | OpenSea | -0.14(-0.19,-0.09) | 3.2E-08 | -0.05(-0.06,-0.03) | 2.1E-13 |
| cg07531398 | 17 | 82392264 | *RPL23AP87;NONE* | intergenic | S_Shelf;N_Shore | 0.11(0.07,0.14) | 5.2E-08 | -0.01(-0.02,0) | 5.8E-03 |
| cg23609577 | 18 | 58345344 | *MC4R;CDH20* | intergenic | OpenSea | 0.12(0.09,0.14) | 1.1E-24 | -0.07(-0.08,-0.05) | 1.6E-17 |
| cg15257756 | 18 | 26470435 | *CDH2;MIR302F* | intergenic | OpenSea | 0.14(0.11,0.18) | 6.6E-18 | -0.03(-0.04,-0.01) | 2.6E-04 |
| cg16498391 | 18 | 12421073 | *PRELID3A* | intronic | S_Shore | 0.1(0.07,0.13) | 1.1E-13 | -0.05(-0.07,-0.03) | 2.0E-07 |
| cg08901202 | 18 | 59985553 | *RELCH;TNFRSF11A* | intergenic | OpenSea | -0.11(-0.15,-0.08) | 6.0E-10 | -0.05(-0.07,-0.03) | 2.0E-06 |
| cg14593760 | 18 | 63119739 | *LINC01924;CDH7* | intergenic | OpenSea | -0.1(-0.14,-0.07) | 2.0E-09 | 0.07(0.05,0.09) | 2.9E-09 |
| cg04164838 | 19 | 14480336 | *LINC01841;ADGRE5* | intergenic | Island | -0.13(-0.15,-0.1) | 7.0E-23 | -0.05(-0.07,-0.04) | 9.2E-11 |
| cg12148678 | 19 | 36120219 | *RBM42* | intronic | N_Shore | -0.13(-0.16,-0.1) | 4.5E-17 | -0.07(-0.08,-0.05) | 1.8E-21 |
| cg24731243 | 19 | 51093903 | *LRRC4B;SNAR-F* | intergenic | OpenSea | 0.1(0.08,0.13) | 2.5E-16 | -0.07(-0.1,-0.05) | 2.2E-07 |
| cg11037750 | 19 | 41331110 | *CYP2T1P;CYP2A6* | intergenic | Island | 0.12(0.09,0.15) | 6.5E-16 | -0.02(-0.04,-0.01) | 5.7E-03 |
| cg07986158 | 19 | 17516347 | *BST2* | exonic | S_Shelf | 0.1(0.08,0.13) | 6.8E-16 | -0.06(-0.09,-0.03) | 2.0E-04 |
| cg24214480 | 19 | 16203713 | *TPM4* | intronic | OpenSea | -0.14(-0.17,-0.1) | 2.7E-15 | -0.06(-0.08,-0.04) | 7.7E-11 |
| cg15863355 | 19 | 15293545 | *NOTCH3* | intronic | OpenSea | -0.12(-0.15,-0.09) | 2.0E-14 | 0.03(0.02,0.04) | 1.3E-08 |
| cg17175421 | 19 | 3025356 | *TLE2* | intronic | N_Shelf | -0.13(-0.17,-0.1) | 4.1E-14 | -0.06(-0.08,-0.04) | 1.9E-08 |
| cg27071707 | 19 | 1614335 | *TCF3* | intronic | Island;N_Shore | 0.11(0.08,0.14) | 5.2E-14 | -0.02(-0.03,-0.01) | 1.3E-04 |
| cg13922488 | 19 | 14434389 | *LINC01841* | ncRNA_intronic | S_Shelf;S_Shore | 0.12(0.08,0.15) | 1.6E-12 | 0.04(0.02,0.05) | 3.3E-09 |
| cg13941306 | 19 | 907556 | *R3HDM4* | intronic | OpenSea | 0.14(0.1,0.19) | 3.5E-12 | -0.07(-0.1,-0.05) | 3.4E-09 |
| cg24538509 | 19 | 40804101 | *AKT2;C19orf47* | intergenic | S_Shelf | 0.12(0.09,0.15) | 4.1E-12 | -0.09(-0.12,-0.06) | 5.9E-09 |
| cg00220152 | 19 | 46300655 | *RSPH6A* | intronic | S_Shelf;N_Shelf | -0.11(-0.15,-0.08) | 1.1E-10 | -0.1(-0.12,-0.07) | 8.6E-13 |
| cg24289580 | 19 | 19461133 | *MAU2* | intronic | OpenSea | -0.11(-0.15,-0.08) | 6.0E-10 | -0.02(-0.03,-0.01) | 6.4E-04 |
| cg25578690 | 19 | 7355545 | *INSR;ARHGEF18* | intergenic | OpenSea | -0.11(-0.15,-0.08) | 9.5E-10 | 0.04(0.02,0.06) | 2.0E-05 |
| cg12429481 | 20 | 54190219 | *LINC01441;CBLN4* | intergenic | OpenSea | -0.11(-0.13,-0.09) | 2.9E-23 | 0.12(0.09,0.15) | 4.5E-13 |
| cg09734183 | 20 | 19943057 | *RIN2* | intronic | OpenSea | 0.13(0.11,0.16) | 5.6E-21 | 0.1(0.07,0.12) | 4.4E-13 |
| cg26701500 | 20 | 37396211 | *ACTR5* | exonic | OpenSea | -0.1(-0.13,-0.08) | 1.0E-17 | -0.03(-0.04,-0.01) | 7.6E-05 |
| cg12494143 | 20 | 4167959 | *SMOX* | exonic | OpenSea | 0.11(0.09,0.14) | 1.4E-17 | -0.06(-0.08,-0.03) | 8.0E-06 |
| cg12223498 | 20 | 39291355 | *LINC01370;MAFB* | intergenic | OpenSea | -0.17(-0.21,-0.13) | 2.2E-15 | 0.06(0.04,0.09) | 5.2E-07 |
| cg10729019 | 20 | 19409017 | *SLC24A3* | intronic | OpenSea | 0.12(0.09,0.16) | 2.7E-13 | -0.08(-0.1,-0.05) | 1.5E-11 |
| cg25379272 | 20 | 50395452 | *ATP9A;SALL4* | intergenic | OpenSea | 0.1(0.08,0.13) | 3.2E-13 | -0.01(-0.02,0) | 2.0E-02 |

| | | | | | | | | | |
|---|---|---|---|---|---|---|---|---|---|
| cg02298220 | 20 | 37286505 | *ARHGAP40;SLC32A1* | intergenic | N_Shelf | -0.12(-0.16,-0.09) | 3.0E-12 | -0.08(-0.09,-0.06) | 5.6E-15 |
| cg25227018 | 20 | 37349615 | *ARHGAP40;SLC32A1* | intergenic | S_Shelf | -0.15(-0.2,-0.1) | 1.8E-09 | 0.12(0.09,0.15) | 1.3E-14 |
| cg10018632 | 20 | 23086396 | *CD93;LINC00656* | intergenic | OpenSea | -0.11(-0.14,-0.07) | 2.1E-09 | -0.02(-0.03,-0.01) | 1.8E-04 |
| cg24171822 | 20 | 46353665 | *SULF2* | intronic | OpenSea | -0.11(-0.14,-0.07) | 3.8E-09 | -0.05(-0.07,-0.03) | 8.4E-06 |
| cg10863113 | 20 | 25032709 | *ACSS1* | intronic | OpenSea | 0.1(0.06,0.14) | 9.2E-08 | -0.04(-0.05,-0.02) | 9.4E-05 |
| cg11767915 | 21 | 17438018 | *USP25;MIR99AHG* | intergenic | OpenSea | -0.11(-0.13,-0.09) | 2.8E-23 | -0.07(-0.09,-0.04) | 8.6E-07 |
| cg11359720 | 21 | 43826560 | *UBASH3A* | intronic | Island | 0.13(0.1,0.17) | 3.2E-17 | -0.05(-0.07,-0.04) | 2.4E-13 |
| cg25982653 | 21 | 46189039 | *UBE2G2* | UTR3 | OpenSea | -0.1(-0.13,-0.07) | 2.3E-14 | -0.1(-0.13,-0.07) | 1.5E-13 |
| cg20045250 | 21 | 29656743 | *LINC01695;LINC00161* | intergenic | OpenSea | 0.11(0.08,0.15) | 9.6E-11 | -0.1(-0.14,-0.07) | 7.7E-10 |
| cg19134305 | 21 | 42317044 | *DSCAM;LINC00323* | intergenic | OpenSea | -0.11(-0.15,-0.07) | 4.3E-08 | -0.06(-0.08,-0.03) | 1.1E-06 |
| cg26212541 | 22 | 30392930 | *MTMR3* | intronic | N_Shelf | 0.13(0.11,0.16) | 9.3E-24 | -0.06(-0.08,-0.04) | 4.5E-09 |
| cg06280150 | 22 | 17845343 | *CECR2* | intronic | S_Shelf | -0.13(-0.16,-0.1) | 2.0E-21 | 0.02(0,0.04) | 1.8E-02 |
| cg07387852 | 22 | 36253143 | *RBFOX2* | intronic | OpenSea | 0.11(0.08,0.13) | 2.3E-20 | 0.07(0.05,0.1) | 6.1E-10 |
| cg26286107 | 22 | 36318062 | *RBFOX2* | intronic | OpenSea | 0.14(0.11,0.17) | 9.8E-20 | 0.07(0.05,0.09) | 1.6E-16 |
| cg19547694 | 22 | 49780357 | *LINC01310;NONE* | intergenic | S_Shelf;N_Shelf | 0.13(0.1,0.16) | 1.5E-18 | -0.04(-0.05,-0.03) | 4.8E-12 |
| cg23659250 | 22 | 49780417 | *LINC01310;NONE* | intergenic | S_Shelf;N_Shelf | -0.1(-0.13,-0.08) | 2.0E-18 | -0.06(-0.08,-0.04) | 7.0E-11 |
| cg06705064 | 22 | 42695185 | *TCF20;LINC01315* | intergenic | S_Shore | -0.12(-0.15,-0.09) | 1.9E-14 | 0.05(0.03,0.07) | 4.2E-08 |
| cg26896611 | 22 | 29601566 | *EMID1* | upstream | N_Shore | -0.11(-0.13,-0.08) | 2.2E-14 | -0.05(-0.07,-0.02) | 3.3E-04 |
| cg19287501 | 22 | 39688719 | *PDGFB;RPL3* | intergenic | S_Shelf | -0.12(-0.15,-0.09) | 3.3E-14 | -0.04(-0.05,-0.02) | 1.0E-04 |
| cg01732982 | 22 | 38419704 | *POLR2F* | intronic | OpenSea | -0.12(-0.16,-0.09) | 3.6E-12 | -0.02(-0.03,-0.01) | 3.5E-06 |
| cg09806134 | 22 | 45897134 | *RIBC2;FBLN1* | intergenic | OpenSea | -0.14(-0.18,-0.1) | 2.2E-11 | -0.06(-0.08,-0.05) | 7.5E-20 |
| cg26466546 | 22 | 46056357 | *LINC01589;ATXN10* | intergenic | S_Shelf | -0.12(-0.16,-0.09) | 8.6E-11 | 0.03(0.02,0.05) | 6.8E-10 |
| cg19742138 | 22 | 42695747 | *TCF20;LINC01315* | intergenic | S_Shelf | -0.1(-0.13,-0.07) | 4.9E-10 | 0.04(0.02,0.05) | 5.1E-07 |
| cg04642170 | 22 | 26452322 | *MYO18B;SEZ6L* | intergenic | OpenSea | -0.12(-0.16,-0.09) | 5.5E-10 | -0.03(-0.04,-0.01) | 1.7E-04 |
| cg01781850 | 22 | 50446080 | *IL17REL* | intronic | N_Shore | -0.12(-0.16,-0.08) | 7.0E-10 | -0.02(-0.03,-0.01) | 7.4E-04 |
| cg20033091 | 22 | 39464236 | *APOBEC3F;APOBEC3G* | intergenic | OpenSea | 0.14(0.09,0.18) | 2.1E-09 | 0.02(0.02,0.03) | 2.5E-09 |
| cg19194233 | 22 | 50085372 | *C22orf34;BRD1* | intergenic | OpenSea | -0.13(-0.17,-0.09) | 2.1E-09 | -0.03(-0.05,-0.01) | 2.4E-04 |
| cg26209034 | 22 | 30204700 | *ASCC2* | intronic | OpenSea | -0.16(-0.21,-0.1) | 8.2E-09 | 0.05(0.04,0.06) | 6.1E-17 |
| cg26301801 | 22 | 37230172 | *PVALB;NCF4-AS1* | intergenic | OpenSea | -0.12(-0.16,-0.08) | 4.8E-08 | -0.05(-0.07,-0.03) | 1.9E-07 |
| cg26622247 | X | 2677017 | *XGY2* | ncRNA_intronic | OpenSea | 0.1(0.08,0.13) | 5.9E-16 | 0.05(0.03,0.07) | 5.7E-07 |
| cg06231618 | X | 40644807 | *MED14OS;LOC100132831* | intergenic | N_Shelf | -0.12(-0.15,-0.08) | 2.9E-12 | 0.09(0.06,0.12) | 5.9E-09 |
| cg21450164 | X | 71128623 | *LOC100132741;NHSL2* | intergenic | OpenSea | 0.1(0.07,0.14) | 3.9E-08 | -0.02(-0.03,-0.01) | 7.2E-03 |

**Supplementary Table 4**

Differentially methylated regions (DMRs) associated with early preterm birth (ePTB) in boys (N=139). DMRs were selected at regional Bonferroni-adjusted $p < 0.05$.

| DMR | #CpGs | Coefficient | Standard Error | *P* | Near Gene | Genomic context |
|---|---|---|---|---|---|---|
| chr5:174617516-174618083 | 5 | -0.88 | 0.06 | 4.73E-42 | | |
| chr2:219400113-219400672 | 4 | -0.72 | 0.057 | 1.45E-30 | *DNPEP* | TSS1500 |
| chr11:8810564-8811270 | 7 | 0.46 | 0.039 | 1.73E-25 | *DENND2B* | TSS1500 |
| chr6:14758502-14759038 | 4 | 0.28 | 0.025 | 8.06E-23 | | |
| chr16:2986197-2986208 | 2 | 0.73 | 0.066 | 3.65E-22 | | |
| chr17:7315706-7316084 | 4 | 0.67 | 0.061 | 4.79E-22 | *NEURL4* | 3UTR |
| chr6:111604241-111604602 | 2 | 0.4 | 0.037 | 9.86E-21 | | |
| chr16:2496264-2496748 | 3 | 1.1 | 0.1 | 1.82E-20 | *TBC1D24* | exon_2 |
| chr2:62450946-62451237 | 2 | 1 | 0.098 | 2.12E-19 | | |
| chr2:96858982-96859272 | 4 | -0.48 | 0.046 | 2.17E-19 | *ANKRD39* | TSS1500 |
| chr22:30881259-30881503 | 3 | -0.78 | 0.075 | 3.13E-19 | *OSBP2* | TSS1500 |
| chr14:23513012-23513320 | 4 | 0.89 | 0.087 | 9.11E-19 | *THTPA* | exon_3 |
| chr6:158738149-158738402 | 2 | 0.82 | 0.08 | 2.05E-18 | | |
| chr1:167664163-167664347 | 2 | 0.82 | 0.08 | 2.07E-18 | | |
| chr2:240403278-240403608 | 3 | -1.1 | 0.11 | 2.61E-18 | | |
| chr1:32882074-32882080 | 2 | 0.73 | 0.072 | 3.52E-18 | | |
| chr9:134410300-134410385 | 2 | 1 | 0.1 | 4.67E-18 | | |
| chr5:149999570-149999857 | 2 | 0.65 | 0.065 | 8.19E-18 | *HMGXB3* | TSS1500 |
| chr1:21322837-21323291 | 2 | -0.75 | 0.075 | 8.28E-18 | | |
| chr7:30911613-30911883 | 8 | -0.5 | 0.05 | 8.63E-18 | *AQP1* | TSS200 |
| chr10:33118133-33118141 | 2 | 1 | 0.1 | 2.28E-17 | *IATPR* | TSS1500 |
| chr1:156248190-156248549 | 6 | 0.4 | 0.04 | 3.12E-17 | *PAQR6* | TSS1500 |
| chr11:46361175-46361591 | 5 | -0.97 | 0.098 | 4.96E-17 | *DGKZ* | TSS200 |
| chr6:46325676-46326401 | 8 | -0.53 | 0.054 | 5.57E-17 | *RCAN2* | TSS1500 |
| chr2:67130508-67130663 | 2 | -1.2 | 0.12 | 7.18E-17 | | |
| chr4:97366509-97366529 | 2 | -0.95 | 0.097 | 1.01E-16 | *STPG2-AS1* | TSS1500 |
| chr1:206535516-206536022 | 3 | 0.57 | 0.059 | 3.14E-16 | | |
| chr19:18464596-18464963 | 2 | 1.2 | 0.13 | 3.33E-16 | | |
| chr3:195081078-195081353 | 2 | -0.97 | 0.1 | 1.48E-15 | | |
| chr17:81843050-81843575 | 4 | 0.87 | 0.092 | 1.64E-15 | *P4HB* | 3UTR |
| chr7:150383885-150384347 | 3 | -0.93 | 0.098 | 2.35E-15 | | |
| chr14:68359972-68360382 | 3 | 0.31 | 0.033 | 3.74E-15 | | |
| chr14:22492844-22494301 | 6 | 0.16 | 0.017 | 8.28E-15 | | |
| chr16:85394241-85394658 | 3 | -0.99 | 0.11 | 1.04E-14 | | |
| chr17:43530528-43531323 | 6 | -0.54 | 0.058 | 1.05E-14 | *ETV4* | TSS1500 |
| chr5:149728891-149729257 | 2 | 0.61 | 0.066 | 1.21E-14 | *PPARGC1B* | TSS1500 |
| chr1:149919103-149919548 | 2 | 0.77 | 0.083 | 1.29E-14 | *SV2A* | TSS1500 |
| chr2:218382129-218382332 | 4 | 0.49 | 0.053 | 1.34E-14 | *SLC11A1* | TSS200 |
| chr7:102097612-102097941 | 3 | -0.99 | 0.11 | 1.55E-14 | | |
| chr4:1365769-1366013 | 3 | -0.81 | 0.087 | 2.57E-14 | | |
| chr2:241762705-241763466 | 5 | -0.46 | 0.05 | 3.64E-14 | | |
| chr14:91244130-91244821 | 8 | 0.39 | 0.043 | 3.79E-14 | *GPR68* | 5UTR |
| chr9:108184917-108185415 | 2 | 1.1 | 0.12 | 5.92E-14 | | |
| chr6:47203911-47204079 | 3 | 0.31 | 0.034 | 6.99E-14 | | |
| chr20:49656476-49656657 | 4 | 0.23 | 0.025 | 9.20E-14 | *B4GALT5* | exon_2 |
| chr2:152486238-152486715 | 2 | -0.34 | 0.038 | 2.17E-13 | | |
| chr17:79993730-79993812 | 2 | -1.2 | 0.14 | 2.29E-13 | | |
| chr1:152810661-152811013 | 2 | 0.65 | 0.072 | 2.41E-13 | *LCE1B* | TSS1500 |
| chr1:206080031-206080075 | 2 | -0.81 | 0.09 | 2.58E-13 | | |
| chr7:2524036-2524622 | 4 | 0.26 | 0.029 | 3.88E-13 | | |

| | | | | | | |
|---|---|---|---|---|---|---|
| chr2:216607062-216607183 | 2 | -0.62 | 0.07 | 4.36E-13 | *LINC01280* | TSS1500 |
| chr9:93144237-93144446 | 2 | 0.44 | 0.049 | 5.39E-13 | | |
| chr7:2001149-2001216 | 2 | -0.94 | 0.11 | 6.03E-13 | | |
| chr10:71757055-71757640 | 5 | 0.22 | 0.024 | 6.12E-13 | | |
| chr14:51715984-51716124 | 2 | -0.48 | 0.054 | 7.39E-13 | *FRMD6* | TSS1500 |
| chr17:72623714-72623759 | 2 | -0.74 | 0.084 | 9.09E-13 | | |
| chr6:32166796-32167418 | 16 | -0.48 | 0.054 | 1.39E-12 | *EGFL8* | exon_6 |
| chr1:16981537-16981808 | 3 | -0.9 | 0.1 | 1.40E-12 | *MFAP2* | TSS1500 |
| chr17:1078490-1079263 | 7 | -0.86 | 0.098 | 2.84E-12 | *ABR* | 5UTR |
| chr1:204286824-204287021 | 2 | 0.97 | 0.11 | 4.00E-12 | | |
| chr19:29817228-29817512 | 3 | -0.82 | 0.095 | 5.44E-12 | *CCNE1* | exon_5 |
| chr2:25251026-25251458 | 2 | -1.2 | 0.14 | 6.27E-12 | | |
| chr3:169821102-169821196 | 2 | 0.49 | 0.057 | 6.54E-12 | | |
| chr17:964542-965041 | 2 | -0.69 | 0.08 | 7.71E-12 | | |
| chr7:836162-836508 | 2 | 0.54 | 0.063 | 8.52E-12 | | |
| chr16:81888761-81889238 | 6 | -0.51 | 0.059 | 8.70E-12 | *PLCG2* | exon_10 |
| chr4:6693196-6693971 | 9 | -0.22 | 0.026 | 9.14E-12 | *S100P* | TSS1500 |
| chr22:46087253-46087270 | 2 | 0.63 | 0.073 | 1.19E-11 | | |
| chr17:76480738-76480883 | 2 | -0.58 | 0.067 | 1.30E-11 | | |
| chr6:6758849-6759535 | 5 | -0.46 | 0.054 | 1.35E-11 | | |
| chr15:40439426-40439771 | 5 | -0.62 | 0.073 | 1.41E-11 | *BAHD1* | TSS1500 |
| chr17:83199997-83200930 | 7 | -0.45 | 0.053 | 2.14E-11 | | |
| chr1:25771427-25771819 | 3 | -0.93 | 0.11 | 2.20E-11 | | |
| chr7:5495671-5495951 | 3 | 0.55 | 0.065 | 2.24E-11 | *MIR589* | TSS200 |
| chr16:85575053-85575288 | 2 | -0.93 | 0.11 | 2.74E-11 | | |
| chr5:1089392-1089685 | 6 | -0.67 | 0.08 | 3.87E-11 | | |
| chr11:64103440-64103756 | 3 | -0.66 | 0.079 | 3.95E-11 | *FLRT1* | 5UTR |
| chr11:118622428-118622681 | 2 | 0.35 | 0.042 | 4.96E-11 | | |
| chr11:2135052-2135231 | 3 | -1 | 0.12 | 5.48E-11 | *MIR483* | TSS1500 |
| chr5:174618106-174618288 | 2 | -0.82 | 0.098 | 5.69E-11 | | |
| chr15:70448131-70448136 | 2 | -1.1 | 0.13 | 6.07E-11 | | |
| chr11:95299256-95299715 | 3 | -0.91 | 0.11 | 6.33E-11 | | |
| chr11:118679351-11867977[illegible] | 3 | 1 | 0.12 | 6.38E-11 | *TREH* | TSS200 |
| chr22:43236418-43236580 | 2 | 0.54 | 0.064 | 6.39E-11 | | |
| chr1:1342532-1342615 | 2 | -0.46 | 0.055 | 6.79E-11 | | |
| chr5:59548970-59549168 | 3 | -0.36 | 0.043 | 7.35E-11 | | |
| chr14:54858784-54858893 | 2 | -0.58 | 0.07 | 7.38E-11 | | |
| chr5:139912477-139912869 | 2 | 0.72 | 0.087 | 7.67E-11 | | |
| chr1:3732271-3733407 | 7 | -0.21 | 0.026 | 8.45E-11 | *TP73* | 3UTR |
| chr14:91277759-91278065 | 2 | 0.78 | 0.093 | 1.15E-10 | *CCDC88C* | exon_29 |
| chr17:7080491-7080868 | 4 | 0.44 | 0.053 | 1.26E-10 | *CLEC10A* | TSS1500 |
| chr14:24369016-24369163 | 3 | -0.91 | 0.11 | 1.47E-10 | *NFATC4* | 5UTR |
| chr19:40366379-40366820 | 2 | -0.8 | 0.097 | 1.54E-10 | *PLD3* | exon_5 |
| chr3:49813066-49813074 | 2 | 0.95 | 0.12 | 1.81E-10 | *UBA7* | exon_4 |
| chr11:6404799-6405664 | 9 | -0.5 | 0.061 | 1.91E-10 | *APBB1* | TSS1500 |
| chr12:56020658-56020706 | 2 | -1.2 | 0.14 | 2.23E-10 | *IKZF4* | TSS1500 |
| chr15:89628462-89628615 | 4 | -0.39 | 0.047 | 2.47E-10 | *KIF7* | exon_19 |
| chr6:30684994-30685955 | 12 | 0.61 | 0.075 | 2.53E-10 | *PPP1R18* | exon_1 |
| chr17:9179187-9179617 | 4 | -0.81 | 0.099 | 2.83E-10 | *OC10192826[illegible]* | TSS1500 |
| chr12:94074983-94075451 | 2 | 0.85 | 0.1 | 2.99E-10 | | |
| chr12:108846273-10884654[illegible] | 2 | 1 | 0.13 | 3.19E-10 | | |
| chr16:2212164-2212292 | 3 | 0.61 | 0.075 | 3.69E-10 | *PGP* | 3UTR |
| chr8:26635599-26635959 | 2 | -0.57 | 0.07 | 4.00E-10 | | |
| chr2:241603855-241603872 | 2 | -0.97 | 0.12 | 4.02E-10 | | |
| chr1:9717463-9717914 | 4 | -0.53 | 0.065 | 4.49E-10 | | |
| chr19:46303650-46303862 | 4 | -0.66 | 0.082 | 5.32E-10 | *HIF3A* | 5UTR |
| chr16:30947994-30948175 | 3 | -0.65 | 0.08 | 5.33E-10 | *FBXL19* | 3UTR |

| | | | | | | |
|---|---|---|---|---|---|---|
| chr22:46078351-46078509 | 3 | 0.63 | 0.077 | 5.49E-10 | | |
| chr15:41881415-41881887 | 3 | 0.45 | 0.055 | 5.59E-10 | | |
| chr1:22212428-22212752 | 3 | -0.27 | 0.033 | 8.47E-10 | | |
| chr4:109848094-109848291 | 3 | -0.49 | 0.061 | 1.13E-09 | *LRIT3* | exon_1 |
| chr16:84737742-84738060 | 2 | 0.89 | 0.11 | 1.44E-09 | | |
| chr1:107689789-107689799 | 2 | 0.95 | 0.12 | 1.48E-09 | *VAV3* | TSS1500 |
| chr1:201737235-201737320 | 2 | -0.59 | 0.074 | 1.77E-09 | | |
| chr9:136583287-136583401 | 2 | 1 | 0.13 | 1.97E-09 | | |
| chr6:33433759-33433765 | 2 | -0.78 | 0.098 | 2.65E-09 | | |
| chr2:10039880-10040161 | 2 | 0.61 | 0.078 | 3.38E-09 | | |
| chr5:139272753-139272830 | 2 | 0.54 | 0.069 | 4.18E-09 | *MATR3* | TSS1500 |
| chr9:121749349-121749831 | 2 | 0.47 | 0.059 | 4.41E-09 | | |
| chr1:10647877-10648034 | 4 | 0.41 | 0.052 | 4.95E-09 | *CASZ1* | exon_16 |
| chr19:44812315-44812464 | 2 | -0.87 | 0.11 | 5.59E-09 | *BCAM* | exon_3 |
| chr16:88939068-88939679 | 5 | 0.17 | 0.021 | 6.03E-09 | *OC10012969* | exon_1 |
| chr12:53103137-53103424 | 5 | -0.98 | 0.13 | 6.43E-09 | *SOAT2* | TSS200 |
| chr11:2533934-2534346 | 4 | -0.46 | 0.059 | 7.64E-09 | | |
| chr9:113579594-113580106 | 4 | -0.56 | 0.072 | 8.18E-09 | *RGS3* | TSS1500 |
| chr7:23473420-23473697 | 2 | -0.55 | 0.07 | 8.43E-09 | | |
| chr12:52181894-52182078 | 2 | -0.95 | 0.12 | 9.50E-09 | | |
| chr1:246005337-246005376 | 2 | 0.72 | 0.093 | 9.55E-09 | | |
| chr2:151161616-151161808 | 2 | -0.89 | 0.12 | 1.23E-08 | | |
| chr12:110349868-11034988 | 2 | 0.6 | 0.078 | 1.26E-08 | *ATP2A2* | 3UTR |
| chr16:2174165-2174310 | 2 | 1 | 0.13 | 1.36E-08 | *TRAF7* | exon_14 |
| chr19:49862983-49863124 | 2 | -0.9 | 0.12 | 1.37E-08 | | |
| chr10:112849539-112849961 | 2 | -0.82 | 0.11 | 1.40E-08 | | |
| chr15:89349026-89349045 | 2 | 0.82 | 0.11 | 1.56E-08 | | |
| chr12:6179096-6179405 | 2 | -0.79 | 0.1 | 1.63E-08 | | |
| chr11:2135464-2135469 | 2 | -0.93 | 0.12 | 1.67E-08 | *IGF2* | exon_2 |
| chr16:67147563-67147584 | 2 | 0.65 | 0.085 | 1.72E-08 | *PHAF1* | 3UTR |
| chr7:128860236-128860420 | 2 | 0.74 | 0.097 | 1.73E-08 | | |
| chr12:851093-851189 | 2 | 0.76 | 0.099 | 1.78E-08 | | |
| chr21:44083013-44083082 | 2 | -0.62 | 0.081 | 1.81E-08 | *TRAPPC10* | exon_14 |
| chr12:53173523-53173556 | 2 | -0.59 | 0.078 | 1.93E-08 | | |
| chr16:68328274-68328320 | 2 | -0.28 | 0.037 | 2.07E-08 | | |
| chr15:40054572-40054962 | 2 | -0.57 | 0.074 | 2.17E-08 | | |
| chr19:12997704-12997889 | 2 | 0.49 | 0.065 | 2.22E-08 | | |
| chr2:121699826-121700254 | 2 | 0.26 | 0.034 | 2.28E-08 | | |
| chr16:2989445-2990020 | 3 | -0.93 | 0.12 | 2.32E-08 | *GREP1* | TSS1500 |
| chr2:64994448-64994554 | 2 | 1 | 0.13 | 2.65E-08 | | |
| chr12:47216223-47216437 | 3 | -0.43 | 0.056 | 2.69E-08 | *PCED1B* | 5UTR |
| chr1:10835460-10836286 | 5 | -0.25 | 0.033 | 2.72E-08 | | |
| chr22:46085723-46085936 | 5 | 0.14 | 0.019 | 2.76E-08 | *MIRLET7BHG* | TSS200 |
| chr2:230925566-230926098 | 3 | -0.27 | 0.035 | 2.85E-08 | *GPR55* | TSS1500 |
| chr11:8709180-8709519 | 3 | -0.98 | 0.13 | 2.96E-08 | | |
| chr4:8228326-8228521 | 2 | 0.56 | 0.074 | 3.54E-08 | *SH3TC1* | exon_12 |
| chr1:35885869-35886240 | 2 | -0.66 | 0.088 | 3.86E-08 | | |
| chr17:73335060-73335744 | 4 | -0.19 | 0.026 | 4.18E-08 | *SDK2* | 3UTR |
| chr7:1900372-1900591 | 3 | -0.81 | 0.11 | 4.57E-08 | | |
| chr7:101237815-101238110 | 7 | -0.74 | 0.098 | 4.68E-08 | *CLDN15* | 5UTR |
| chr10:3804303-3804519 | 2 | -0.39 | 0.052 | 4.69E-08 | | |
| chr1:156509834-156509919 | 2 | -0.64 | 0.085 | 4.91E-08 | | |
| chr7:101818812-101818835 | 2 | -0.67 | 0.089 | 5.14E-08 | | |
| chr5:177448296-177448314 | 2 | 0.85 | 0.11 | 5.52E-08 | *PRR7-AS1* | TSS1500 |
| chr17:81842892-81842983 | 2 | 0.88 | 0.12 | 6.46E-08 | | |
| chr11:66876597-66876604 | 2 | -0.59 | 0.079 | 6.51E-08 | | |
| chr11:71488134-71488417 | 2 | -0.67 | 0.089 | 7.07E-08 | | |

| | | | | | | |
|---|---|---|---|---|---|---|
| chr22:46040669-46041398 | 3 | 1 | 0.14 | 7.10E-08 | *LINC00899* | exon_4 |
| chr16:4306531-4306821 | 2 | -0.58 | 0.077 | 7.64E-08 | | |
| chr9:93142078-93142216 | 2 | 1 | 0.14 | 8.28E-08 | | |
| chr20:10857420-10857491 | 2 | -0.88 | 0.12 | 8.57E-08 | | |
| chr19:46300655-46300863 | 2 | -0.93 | 0.12 | 8.65E-08 | | |
| chr11:845387-846237 | 4 | -0.55 | 0.074 | 9.88E-08 | | |
| chr4:3369564-3369793 | 3 | 0.21 | 0.028 | 1.02E-07 | *RGS12* | TSS1500 |
| chr5:149605727-149605771 | 2 | 0.44 | 0.059 | 1.09E-07 | | |
| chr20:47484420-47484633 | 5 | 0.7 | 0.094 | 1.14E-07 | | |
| chr15:41822237-41822643 | 2 | -0.32 | 0.043 | 1.18E-07 | *MAPKBP1* | exon_26 |
| chr2:128362318-128363217 | 3 | -0.83 | 0.11 | 1.18E-07 | | |
| chr19:851045-851650 | 4 | 0.33 | 0.044 | 1.23E-07 | *ELANE* | TSS1500 |
| chr19:38278450-38278516 | 2 | 0.42 | 0.056 | 1.24E-07 | | |
| chr11:2141248-2141315 | 5 | -1 | 0.14 | 1.27E-07 | *IGF2* | TSS1500 |
| chr22:27795793-27795926 | 2 | -1 | 0.14 | 1.34E-07 | | |
| chr19:16067704-16067760 | 2 | 0.61 | 0.083 | 1.41E-07 | *TPM4* | exon_1 |
| chr15:39788373-39788507 | 2 | -0.99 | 0.13 | 1.45E-07 | | |
| chr16:1655337-1656200 | 8 | 0.57 | 0.078 | 1.48E-07 | *CRAMP1* | exon_10 |
| chr8:117956956-117957102 | 2 | 0.5 | 0.068 | 1.60E-07 | | |
| chr10:103737008-103737064 | 2 | 0.41 | 0.056 | 1.65E-07 | | |
| chr11:72741437-72741722 | 2 | -0.34 | 0.046 | 1.70E-07 | | |
| chr7:151062477-151062546 | 3 | -0.44 | 0.059 | 1.72E-07 | *SLC4A2* | TSS200 |
| chr9:92717503-92717642 | 2 | 0.67 | 0.091 | 1.78E-07 | | |
| chr16:70701010-70701400 | 2 | 0.91 | 0.12 | 1.87E-07 | | |
| chr1:19645806-19646283 | 3 | 0.54 | 0.073 | 1.96E-07 | *NBL1* | 5UTR |
| chr17:29572241-29572250 | 2 | 0.8 | 0.11 | 2.16E-07 | *TP53I13* | exon_6 |
| chr3:78496655-78496721 | 2 | -1.1 | 0.14 | 2.20E-07 | | |
| chr17:19387040-19387807 | 8 | -0.48 | 0.065 | 2.22E-07 | *MFAP4* | TSS1500 |
| chr22:30300616-30300655 | 2 | 0.81 | 0.11 | 2.43E-07 | | |
| chr1:205246106-205246783 | 6 | -0.24 | 0.033 | 2.54E-07 | *TMCC2* | 5UTR |
| chr2:239267790-239267916 | 2 | -0.56 | 0.077 | 2.59E-07 | | |
| chr3:153159825-153159893 | 2 | 0.71 | 0.097 | 2.64E-07 | | |
| chr16:87780227-87780572 | 3 | 0.26 | 0.035 | 2.70E-07 | *OC10272446* | TSS1500 |
| chr14:76924708-76924714 | 2 | -0.68 | 0.093 | 2.79E-07 | | |
| chr11:66317701-66318487 | 5 | -0.63 | 0.087 | 2.92E-07 | *CD248* | TSS1500 |
| chr12:106205593-106205662 | 2 | 0.71 | 0.098 | 2.93E-07 | | |
| chr4:7371361-7371616 | 2 | -0.78 | 0.11 | 3.12E-07 | | |
| chr1:151372705-151372863 | 3 | 0.15 | 0.02 | 3.13E-07 | *SELENBP1* | 5UTR |
| chr1:150272362-150272651 | 3 | -0.65 | 0.089 | 3.18E-07 | *C1orf54* | TSS1500 |
| chr5:181198956-181199247 | 4 | -0.58 | 0.079 | 3.25E-07 | *TRIM7* | exon_4 |
| chr3:184320163-184320529 | 6 | 0.38 | 0.053 | 3.35E-07 | *EIF4G1* | 5UTR |
| chr5:179803002-179803091 | 2 | 0.74 | 0.1 | 3.47E-07 | *MGAT4B* | TSS1500 |
| chr12:124385848-124386274 | 3 | -0.61 | 0.083 | 3.68E-07 | *NCOR2* | exon_19 |
| chr6:30614397-30614639 | 5 | 0.25 | 0.034 | 3.72E-07 | | |
| chr2:134368283-134368302 | 2 | -0.92 | 0.13 | 3.74E-07 | | |
| chr16:85445163-85445302 | 2 | -0.58 | 0.08 | 3.75E-07 | | |
| chr2:232316147-232316640 | 2 | 0.83 | 0.11 | 3.83E-07 | | |
| chr3:129428698-129428710 | 2 | 0.094 | 0.013 | 3.98E-07 | *EFCAB12* | TSS200 |
| chr8:142375142-142375333 | 2 | 0.84 | 0.12 | 4.02E-07 | | |
| chr2:86036101-86036147 | 2 | -0.77 | 0.11 | 4.39E-07 | | |
| chr8:78805734-78805818 | 2 | 0.93 | 0.13 | 4.41E-07 | *IL7* | TSS1500 |
| chr17:17780696-17781299 | 3 | -0.64 | 0.088 | 4.97E-07 | *SMCR5* | TSS1500 |
| chr16:74447709-74447764 | 2 | -0.45 | 0.062 | 5.03E-07 | *GLG1* | 3UTR |
| chr3:49903386-49903420 | 2 | 0.52 | 0.072 | 5.41E-07 | *MST1R* | exon_1 |
| chr12:113333083-11333340 | 3 | -0.64 | 0.088 | 5.51E-07 | | |
| chr15:31352375-31352398 | 2 | 0.96 | 0.13 | 5.62E-07 | | |
| chr9:79974823-79975095 | 2 | 0.49 | 0.068 | 5.68E-07 | *LINC01507* | TSS1500 |

| | | | | | | |
|---|---|---|---|---|---|---|
| chr8:123658194-123658733 | 8 | -0.63 | 0.087 | 6.03E-07 | | |
| chr5:82640224-82640364 | 2 | -1.1 | 0.15 | 6.22E-07 | | |
| chr12:54352283-54352306 | 2 | -0.81 | 0.11 | 7.31E-07 | *OC10272405* | TSS1500 |
| chr17:83195593-83196268 | 4 | 0.82 | 0.11 | 7.65E-07 | | |
| chr6:29602730-29603297 | 4 | -0.54 | 0.076 | 8.79E-07 | *GABBR1* | 3UTR |
| chr15:77044232-77044596 | 2 | 0.85 | 0.12 | 9.06E-07 | *TSPAN3* | 3UTR |
| chr2:43039487-43039704 | 4 | 0.31 | 0.043 | 9.25E-07 | *LINC01819* | TSS200 |
| chr2:96146528-96147251 | 7 | -0.48 | 0.068 | 9.60E-07 | *DUSP2* | TSS1500 |
| chr6:3269268-3269347 | 3 | -0.78 | 0.11 | 9.75E-07 | *SLC22A23* | 3UTR |
| chr11:108788642-108788997 | 2 | -0.72 | 0.1 | 1.05E-06 | | |
| chr5:177400160-177400670 | 3 | -0.5 | 0.07 | 1.07E-06 | *PFN3* | exon_1 |
| chr1:27567375-27567379 | 2 | -0.24 | 0.033 | 1.16E-06 | | |
| chr2:219434690-219434937 | 6 | -0.45 | 0.063 | 1.20E-06 | *SPEG* | 5UTR |
| chr22:47144069-47144276 | 2 | 0.99 | 0.14 | 1.21E-06 | | |
| chr2:113234085-113234315 | 2 | 0.85 | 0.12 | 1.24E-06 | *PAX8-AS1* | TSS1500 |
| chr2:38097842-38098399 | 3 | -0.43 | 0.061 | 1.25E-06 | | |
| chr1:38816215-38816331 | 2 | -0.91 | 0.13 | 1.33E-06 | | |
| chr1:19643437-19643530 | 4 | 0.86 | 0.12 | 1.35E-06 | *NBL1* | TSS1500 |
| chr1:210328277-210328290 | 2 | 0.43 | 0.061 | 1.45E-06 | *HHAT* | TSS1500 |
| chr13:30157286-30157321 | 2 | -0.86 | 0.12 | 1.59E-06 | | |
| chr19:3383485-3383596 | 2 | 0.7 | 0.099 | 1.71E-06 | | |
| chr15:69593949-69594183 | 4 | 0.24 | 0.035 | 1.85E-06 | | |
| chr14:81450226-81450437 | 4 | -0.71 | 0.1 | 1.97E-06 | *LINC02308* | TSS200 |
| chr4:7393175-7393300 | 2 | -0.51 | 0.073 | 2.07E-06 | | |
| chr11:74673394-74673473 | 2 | -0.42 | 0.06 | 2.45E-06 | | |
| chr13:21033288-21033427 | 2 | 0.59 | 0.085 | 2.47E-06 | | |
| chr1:1431645-1431926 | 3 | -0.63 | 0.09 | 2.55E-06 | | |
| chr12:125149097-125149507 | 2 | 0.69 | 0.098 | 2.60E-06 | | |
| chr22:19905140-19905143 | 2 | -0.9 | 0.13 | 2.62E-06 | | |
| chr1:225817687-225817950 | 2 | 0.53 | 0.076 | 2.62E-06 | | |
| chr11:69669789-69669806 | 2 | 1 | 0.15 | 2.63E-06 | | |
| chr3:11136300-11136378 | 2 | 0.65 | 0.092 | 2.70E-06 | *HRH1* | TSS1500 |
| chr4:122826292-122826517 | 6 | -0.69 | 0.099 | 2.87E-06 | *FGF2* | TSS1500 |
| chr16:2084630-2084736 | 3 | -0.86 | 0.12 | 2.91E-06 | *TSC2* | exon_33 |
| chr1:6599145-6599293 | 3 | 0.73 | 0.1 | 3.00E-06 | *KLHL21* | exon_2 |
| chr1:43300273-43300535 | 2 | 0.2 | 0.029 | 3.09E-06 | *TIE1* | TSS1500 |
| chr5:146047637-146047789 | 2 | 1.2 | 0.17 | 3.18E-06 | *SH3RF2* | exon_6 |
| chr11:130198280-130198553 | 3 | -0.77 | 0.11 | 3.40E-06 | *ST14* | exon_13 |
| chr11:68004530-68004803 | 2 | -0.36 | 0.052 | 3.42E-06 | *UNC93B1* | TSS1500 |
| chr6:134663309-134663387 | 2 | -0.75 | 0.11 | 3.42E-06 | | |
| chr11:69216330-69216675 | 2 | 0.71 | 0.1 | 3.54E-06 | | |
| chr22:18064734-18064932 | 2 | -0.65 | 0.094 | 3.59E-06 | | |
| chr7:80646259-80646568 | 5 | -0.6 | 0.086 | 3.75E-06 | *CD36* | 5UTR |
| chr1:154407220-154407588 | 2 | 0.62 | 0.089 | 3.79E-06 | *IL6R-AS1* | TSS1500 |
| chr11:2889598-2889806 | 2 | -0.79 | 0.11 | 3.85E-06 | | |
| chr11:43895808-43896067 | 2 | -0.87 | 0.12 | 3.96E-06 | *SEC14L1P1* | TSS1500 |
| chr6:21354305-21354311 | 2 | -0.71 | 0.1 | 4.01E-06 | | |
| chr14:51704520-51704908 | 2 | -0.53 | 0.077 | 4.02E-06 | *FRMD6* | exon_6 |
| chr8:23494300-23494312 | 2 | -0.55 | 0.08 | 4.15E-06 | | |
| chr7:43144013-43144116 | 2 | 0.78 | 0.11 | 4.16E-06 | | |
| chr22:17167606-17167613 | 2 | 0.96 | 0.14 | 4.33E-06 | | |
| chr12:105814199-105814284 | 2 | -0.55 | 0.079 | 4.33E-06 | | |
| chr1:201127994-201128211 | 2 | -0.6 | 0.087 | 4.88E-06 | *ASCL5* | TSS1500 |
| chr2:3257098-3257553 | 6 | 0.15 | 0.022 | 4.96E-06 | *EIPR1* | exon_4 |
| chr5:122816880-122817045 | 3 | -0.41 | 0.059 | 5.35E-06 | *SNX2* | exon_9 |
| chr8:96327997-96328042 | 2 | -0.76 | 0.11 | 5.47E-06 | | |
| chr16:67429999-67430428 | 2 | 0.72 | 0.1 | 5.61E-06 | *HSD11B2* | TSS1500 |

| | | | | | | |
|---|---|---|---|---|---|---|
| chr17:77441430-77442051 | 4 | -0.46 | 0.067 | 5.98E-06 | | |
| chr11:114281245-11428127C | 2 | 0.68 | 0.099 | 6.19E-06 | | |
| chr1:31944266-31945077 | 7 | -0.25 | 0.037 | 6.24E-06 | | |
| chr15:70636742-70637024 | 2 | -0.59 | 0.085 | 6.31E-06 | *SALR2* | exon_1 |
| chr13:110508203-11050836C | 2 | 0.95 | 0.14 | 6.52E-06 | *COL4A2-AS1* | TSS200 |
| chr3:184691542-184692068 | 3 | -0.43 | 0.063 | 6.63E-06 | | |
| chr3:184317366-184317390 | 2 | -0.55 | 0.08 | 6.97E-06 | *EIF4G1* | exon_5 |
| chr1:207064629-207064872 | 2 | 0.63 | 0.092 | 7.05E-06 | | |
| chr19:58395237-58395895 | 5 | 0.18 | 0.027 | 7.07E-06 | *RNF225* | 5UTR |
| chr6:63571732-63571838 | 4 | -0.85 | 0.12 | 7.34E-06 | *PTP4A1* | TSS1500 |
| chr2:10344130-10344454 | 3 | -0.54 | 0.079 | 7.42E-06 | | |
| chr19:50968555-50968886 | 4 | -0.76 | 0.11 | 7.47E-06 | *KLK6* | 5UTR |
| chr6:56541166-56541538 | 3 | 0.22 | 0.033 | 7.98E-06 | | |
| chr6:30684460-30684619 | 6 | 0.65 | 0.095 | 8.35E-06 | *PPP1R18* | exon_1 |
| chr13:73101604-73101881 | 2 | -0.95 | 0.14 | 8.44E-06 | | |
| chr6:15816900-15817571 | 3 | -0.62 | 0.091 | 8.45E-06 | | |
| chr6:33278116-33278317 | 6 | 0.23 | 0.034 | 9.26E-06 | *B3GALT4* | exon_1 |
| chr2:219595363-219595792 | 2 | -0.64 | 0.094 | 9.46E-06 | | |
| chr1:2477896-2477900 | 2 | -0.63 | 0.093 | 9.48E-06 | | |
| chr16:85445447-85445695 | 3 | -0.63 | 0.093 | 9.62E-06 | | |
| chr19:13054140-13054149 | 2 | 0.6 | 0.088 | 9.89E-06 | | |
| chr12:53214132-53214214 | 2 | 0.68 | 0.099 | 1.02E-05 | *RARG* | exon_5 |
| chr6:31403711-31403773 | 3 | -0.046 | 0.0067 | 1.09E-05 | | |
| chr6:110415662-110415738 | 2 | 0.73 | 0.11 | 1.14E-05 | *DDO* | TSS200 |
| chr12:68447523-68447624 | 2 | -0.65 | 0.096 | 1.30E-05 | | |
| chr20:23145559-23145612 | 2 | 0.67 | 0.099 | 1.31E-05 | | |
| chr10:5560086-5560117 | 2 | -0.81 | 0.12 | 1.37E-05 | | |
| chr7:366043-366421 | 3 | -0.51 | 0.076 | 1.50E-05 | | |
| chr14:90726518-90726723 | 2 | 0.26 | 0.039 | 1.55E-05 | | |
| chr2:8475324-8475624 | 2 | -0.47 | 0.069 | 1.57E-05 | | |
| chr3:14267771-14267824 | 2 | -0.62 | 0.091 | 1.58E-05 | | |
| chr1:51404463-51404632 | 2 | -0.86 | 0.13 | 1.60E-05 | | |
| chr20:37396211-37396266 | 4 | -0.49 | 0.073 | 1.63E-05 | *SRC* | exon_8 |
| chr9:33431357-33431813 | 3 | 0.6 | 0.089 | 1.64E-05 | | |
| chr9:122394486-122394791 | 2 | 0.23 | 0.034 | 1.81E-05 | *PTGS1* | 3UTR |
| chr1:2305004-2305464 | 3 | -0.69 | 0.1 | 1.84E-05 | | |
| chr14:103709218-103709431 | 3 | 0.77 | 0.11 | 1.96E-05 | | |
| chr2:20184524-20184735 | 2 | 0.28 | 0.042 | 1.96E-05 | | |
| chr11:66935220-66935623 | 3 | -0.2 | 0.029 | 2.00E-05 | *MIR3163* | TSS1500 |
| chr11:65917791-65918316 | 3 | 0.73 | 0.11 | 2.03E-05 | *C11orf68* | exon_2 |
| chr6:6928065-6928352 | 2 | -0.2 | 0.03 | 2.04E-05 | | |
| chr12:41188253-41188265 | 3 | -0.56 | 0.083 | 2.06E-05 | *PDZRN4* | TSS200 |
| chr1:206539937-206540200 | 2 | 0.81 | 0.12 | 2.08E-05 | | |
| chr3:52402589-52402644 | 2 | 0.45 | 0.068 | 2.13E-05 | *BAP1* | exon_16 |
| chr22:46079768-46079819 | 2 | 0.67 | 0.099 | 2.25E-05 | | |
| chr14:91380360-91380650 | 2 | -0.81 | 0.12 | 2.26E-05 | | |
| chr1:13582457-13583128 | 4 | -1 | 0.15 | 2.28E-05 | *PDPN* | TSS1500 |
| chr1:53524818-53524882 | 2 | -0.54 | 0.081 | 2.28E-05 | *GLIS1* | exon_6 |
| chr3:65401633-65401866 | 3 | -0.69 | 0.1 | 2.34E-05 | | |
| chr16:88698266-88698663 | 2 | -0.72 | 0.11 | 2.39E-05 | | |
| chr8:546527-546618 | 2 | -0.81 | 0.12 | 2.43E-05 | *TDRP* | TSS1500 |
| chr9:129677730-129677819 | 2 | -0.53 | 0.079 | 2.54E-05 | | |
| chr17:78680161-78680221 | 2 | -0.9 | 0.13 | 2.56E-05 | *CYTH1* | exon_12 |
| chr22:17842003-17842009 | 2 | -0.64 | 0.095 | 2.57E-05 | *MICAL3* | exon_19 |
| chr3:37453675-37454315 | 3 | 0.31 | 0.047 | 2.76E-05 | | |
| chr4:8473070-8473134 | 2 | -0.91 | 0.14 | 2.86E-05 | | |
| chr12:3150617-3151008 | 2 | -0.58 | 0.087 | 2.95E-05 | | |

| | | | | | | |
|---|---|---|---|---|---|---|
| chr17:40015202-40015277 | 3 | -0.58 | 0.087 | 2.99E-05 | *CSF3* | TSS1500 |
| chr8:100960719-100960744 | 2 | -0.64 | 0.097 | 3.00E-05 | | |
| chr17:82493190-82493473 | 2 | 0.64 | 0.097 | 3.01E-05 | | |
| chr2:226795357-226796115 | 4 | -0.84 | 0.13 | 3.12E-05 | *IRS1* | exon_1 |
| chr21:43767112-43767186 | 2 | -0.81 | 0.12 | 3.23E-05 | | |
| chr22:46374298-46374706 | 5 | -0.62 | 0.094 | 3.29E-05 | | |
| chr3:9168432-9168552 | 3 | -0.49 | 0.074 | 3.42E-05 | | |
| chr17:40560954-40560990 | 2 | -0.68 | 0.1 | 3.46E-05 | *CCR7* | 5UTR |
| chr15:70559458-70559776 | 2 | -0.67 | 0.1 | 3.53E-05 | | |
| chr1:9847978-9848246 | 4 | -0.73 | 0.11 | 3.65E-05 | | |
| chr19:4338855-4339163 | 5 | -0.16 | 0.024 | 3.70E-05 | *STAP2* | TSS200 |
| chr11:126354534-12635509[illegible] | 3 | -0.9 | 0.14 | 3.74E-05 | *ST3GAL4* | TSS1500 |
| chr6:41639032-41639588 | 4 | -0.89 | 0.14 | 4.35E-05 | | |
| chr11:1782993-1783085 | 2 | -0.56 | 0.086 | 4.36E-05 | | |
| chr1:40421368-40421804 | 2 | -0.45 | 0.068 | 4.37E-05 | | |
| chr12:109914965-10991513[illegible] | 2 | 0.84 | 0.13 | 4.48E-05 | | |
| chr12:52926522-52926810 | 2 | -0.35 | 0.053 | 4.53E-05 | *KRT8* | TSS1500 |
| chr12:112134733-11213508[illegible] | 2 | 0.46 | 0.069 | 4.63E-05 | | |
| chr19:4475058-4475139 | 2 | -0.72 | 0.11 | 4.76E-05 | | |
| chr1:2134991-2135192 | 3 | -0.67 | 0.1 | 4.99E-05 | | |
| chr2:205763364-205764023 | 10 | -0.48 | 0.073 | 5.27E-05 | *NRP2* | exon_13 |
| chr7:2121095-2121236 | 3 | -0.81 | 0.12 | 5.34E-05 | | |
| chr6:169289844-169290176 | 2 | -0.23 | 0.035 | 5.46E-05 | | |
| chr11:8732823-8733223 | 4 | 0.41 | 0.062 | 5.66E-05 | | |
| chr16:2991348-2991584 | 2 | -0.97 | 0.15 | 5.69E-05 | | |
| chr11:66722050-66722210 | 3 | -1.1 | 0.16 | 5.71E-05 | | |
| chr1:15735866-15735950 | 2 | 0.6 | 0.092 | 5.88E-05 | *SLC25A34* | TSS1500 |
| chr5:74652323-74652374 | 2 | 0.7 | 0.11 | 6.01E-05 | | |
| chr15:101174593-10117493[illegible] | 3 | 0.95 | 0.15 | 6.05E-05 | | |
| chr14:95227544-95227644 | 2 | -0.55 | 0.085 | 6.12E-05 | | |
| chr5:74103856-74103876 | 3 | 0.78 | 0.12 | 6.13E-05 | | |
| chr14:105028490-10502858[illegible] | 2 | 0.61 | 0.093 | 6.44E-05 | | |
| chr16:2768494-2768682 | 2 | -0.88 | 0.14 | 6.51E-05 | | |
| chr1:223190137-223190532 | 2 | -0.29 | 0.044 | 6.54E-05 | | |
| chr6:32842965-32843476 | 6 | 0.68 | 0.1 | 6.93E-05 | *PSMB8-AS1* | TSS1500 |
| chr16:56607035-56608086 | 5 | 0.49 | 0.076 | 7.05E-05 | *MT2A* | TSS1500 |
| chr12:10190044-10190117 | 3 | 0.78 | 0.12 | 7.12E-05 | *TMEM52B* | exon_4 |
| chr6:143961894-143962211 | 3 | -0.48 | 0.074 | 7.12E-05 | *PLAGL1* | TSS1500 |
| chr6:33781666-33781843 | 2 | -0.37 | 0.057 | 7.18E-05 | | |
| chr17:75824369-75824597 | 2 | -0.94 | 0.14 | 7.41E-05 | *UNK* | exon_16 |
| chr11:4609064-4609174 | 2 | 1.1 | 0.18 | 7.52E-05 | *TRIM68* | TSS1500 |
| chr20:50590144-50590377 | 2 | -0.87 | 0.13 | 7.53E-05 | | |
| chr20:47487112-47487249 | 2 | -0.82 | 0.13 | 7.97E-05 | | |
| chr20:36173765-36173888 | 2 | -0.63 | 0.096 | 8.52E-05 | *EPB41L1* | 5UTR |
| chr7:100099549-100099702 | 3 | -0.28 | 0.043 | 8.75E-05 | *MCM7* | exon_3 |
| chr2:12077000-12077166 | 2 | -0.57 | 0.089 | 9.15E-05 | | |
| chr17:77375123-77375682 | 5 | -0.47 | 0.072 | 9.67E-05 | *SEPTIN9* | TSS1500 |
| chr22:27669558-27669808 | 2 | -1 | 0.16 | 9.79E-05 | | |
| chr2:43068457-43068615 | 2 | -0.78 | 0.12 | 9.96E-05 | | |
| chr8:38150973-38151112 | 3 | -0.34 | 0.052 | 0.000101 | *STAR* | TSS200 |
| chr1:8154903-8154918 | 2 | -1 | 0.16 | 0.000104 | | |
| chr16:3509551-3509568 | 2 | -0.74 | 0.11 | 0.000107 | *CLUAP1* | TSS1500 |
| chr1:9400145-9400280 | 2 | 0.84 | 0.13 | 0.000116 | | |
| chr2:190838146-190838473 | 2 | -0.6 | 0.094 | 0.000121 | | |
| chr11:64103771-64103793 | 2 | -0.62 | 0.096 | 0.000125 | *FLRT1* | 5UTR |
| chr7:108001045-108001351 | 3 | -0.66 | 0.1 | 0.000126 | | |
| chr12:4278496-4278859 | 5 | -0.68 | 0.11 | 0.000128 | *CCND2* | exon_3 |

| | | | | | | |
|---|---|---|---|---|---|---|
| chr3:14175571-14175970 | 2 | 0.85 | 0.13 | 0.000131 | | |
| chr14:64763569-64763904 | 2 | -0.68 | 0.11 | 0.000134 | | |
| chr11:129947523-12994797€ | 6 | -0.67 | 0.1 | 0.000146 | *PRDM10* | TSS1500 |
| chr18:8778979-8778987 | 2 | -0.77 | 0.12 | 0.000148 | | |
| chr16:27625324-27625336 | 2 | 0.67 | 0.1 | 0.00015 | | |
| chr16:10984660-10984733 | 2 | -0.76 | 0.12 | 0.000154 | | |
| chr17:74044022-74044029 | 2 | 0.83 | 0.13 | 0.000156 | | |
| chr8:40099277-40099760 | 2 | -1.1 | 0.18 | 0.000159 | | |
| chr15:69730833-69730843 | 2 | -0.56 | 0.087 | 0.000161 | | |
| chr18:10453627-10453670 | 2 | -0.97 | 0.15 | 0.000163 | *APCDD1* | TSS1500 |
| chr14:23300256-23300306 | 2 | 0.48 | 0.075 | 0.000168 | | |
| chr1:2128552-2128575 | 2 | -0.61 | 0.096 | 0.000168 | | |
| chr1:2503797-2504116 | 2 | -0.39 | 0.061 | 0.00017 | *PLCH2* | exon_22 |
| chr16:8949378-8949754 | 2 | -0.44 | 0.068 | 0.000172 | *USP7* | TSS200 |
| chr2:217858626-217858904 | 2 | -0.4 | 0.063 | 0.000174 | | |
| chr7:986189-986219 | 3 | -0.73 | 0.12 | 0.00018 | | |
| chr1:1421522-1421981 | 4 | -0.14 | 0.022 | 0.000184 | *ANKRD65* | TSS1500 |
| chr6:43510986-43511091 | 2 | 0.7 | 0.11 | 0.000192 | *LRRC73* | TSS1500 |
| chr9:76297521-76297612 | 2 | -0.96 | 0.15 | 0.000207 | | |
| chr16:27243244-27243251 | 2 | -0.25 | 0.04 | 0.000208 | | |
| chr12:68452005-68452155 | 3 | 0.75 | 0.12 | 0.000208 | *LINC02384* | TSS1500 |
| chr8:1882516-1882602 | 3 | -0.72 | 0.11 | 0.000211 | | |
| chr3:128112418-128112830 | 3 | -0.77 | 0.12 | 0.000225 | | |
| chr7:925743-925898 | 3 | -0.66 | 0.1 | 0.000226 | | |
| chr15:67185698-67185888 | 3 | 0.49 | 0.077 | 0.000228 | | |
| chr1:27350581-27350749 | 2 | -0.68 | 0.11 | 0.000232 | | |
| chr19:35138900-35139451 | 5 | -0.49 | 0.078 | 0.000234 | *FXYD1* | TSS1500 |
| chr20:25214563-25214636 | 2 | -0.47 | 0.074 | 0.000237 | | |
| chr12:10392484-10392872 | 2 | 0.52 | 0.082 | 0.000247 | | |
| chr3:49422624-49422678 | 2 | -0.68 | 0.11 | 0.000249 | *AMT* | TSS1500 |
| chr3:58102121-58102418 | 2 | 1.2 | 0.19 | 0.000261 | | |
| chr12:57894274-57894298 | 2 | -0.65 | 0.1 | 0.000261 | *LOC283387* | exon_1 |
| chr1:6477549-6477725 | 3 | -0.62 | 0.099 | 0.000263 | *PLEKHG5* | exon_2 |
| chr4:104906393-104906501 | 2 | -0.6 | 0.094 | 0.000272 | | |
| chr14:100018883-10001947€ | 5 | -0.41 | 0.065 | 0.000273 | | |
| chr3:181710258-181710909 | 14 | -0.32 | 0.05 | 0.000277 | *SOX2* | TSS1500 |
| chr3:48372217-48372248 | 2 | -0.85 | 0.13 | 0.000282 | *FBXW12* | 5UTR |
| chr22:46063144-46063447 | 2 | 0.83 | 0.13 | 0.00029 | | |
| chr20:21391488-21392225 | 6 | -0.33 | 0.052 | 0.000294 | | |
| chr20:37524784-37524942 | 3 | 0.18 | 0.029 | 0.000297 | *BLCAP* | TSS1500 |
| chr9:123776859-123777048 | 2 | -0.82 | 0.13 | 0.000298 | | |
| chr1:228021653-228021944 | 2 | -0.3 | 0.048 | 0.000299 | | |
| chr16:30845068-30845279 | 2 | 0.81 | 0.13 | 0.000321 | | |
| chr20:57579824-57579841 | 2 | 0.45 | 0.072 | 0.000332 | | |
| chr3:37693736-37694110 | 2 | -0.45 | 0.072 | 0.00034 | | |
| chr8:123202889-123203117 | 3 | -0.39 | 0.061 | 0.000344 | *FAM83A-AS1* | TSS200 |
| chr20:25294505-25294850 | 2 | -0.65 | 0.1 | 0.000345 | *ABHD12* | 3UTR |
| chr1:181096362-181096458 | 2 | -0.65 | 0.1 | 0.000346 | | |
| chr12:56020724-56020914 | 3 | -0.96 | 0.15 | 0.000348 | *IKZF4* | TSS200 |
| chr3:195883360-195883468 | 3 | 0.5 | 0.08 | 0.00035 | *MIR6829* | TSS1500 |
| chr3:184317731-184317739 | 2 | -0.48 | 0.077 | 0.000362 | *EIF4G1* | exon_6 |
| chr11:1927809-1927900 | 4 | -0.76 | 0.12 | 0.000368 | | |
| chr19:51887981-51888004 | 3 | -0.041 | 0.0066 | 0.000388 | *ZNF577* | TSS200 |
| chr10:124035620-12403640C | 4 | -0.38 | 0.06 | 0.000399 | | |
| chr12:121038746-121038917 | 3 | 0.91 | 0.15 | 0.000412 | *OASL* | exon_1 |
| chr19:11195237-11195335 | 3 | -0.7 | 0.11 | 0.000419 | | |
| chr15:78048029-78048040 | 2 | -0.93 | 0.15 | 0.000431 | | |

| | | | | | | |
|---|---|---|---|---|---|---|
| chr12:106140125-10614043¢ | 2 | -0.72 | 0.11 | 0.000436 | *NUAK1* | TSS1500 |
| chr17:77429843-77429940 | 2 | -0.45 | 0.073 | 0.000444 | | |
| chr17:58363352-58363581 | 3 | -0.7 | 0.11 | 0.000446 | *RNF43* | exon_4 |
| chr6:170096333-170096371 | 2 | -0.77 | 0.12 | 0.000453 | | |
| chr15:63998688-63999124 | 2 | 0.77 | 0.12 | 0.000464 | | |
| chr14:75222250-75222363 | 2 | -0.58 | 0.094 | 0.000467 | | |
| chr5:138875324-138875382 | 3 | -0.23 | 0.036 | 0.000484 | *CTN1* | 5UTR |
| chr11:45209624-45209994 | 3 | -0.79 | 0.13 | 0.000488 | | |
| chr7:27030742-27030978 | 2 | -0.6 | 0.096 | 0.000488 | | |
| chr17:3660347-3660484 | 2 | 0.8 | 0.13 | 0.000496 | *CTNS* | 3UTR |
| chr2:1653245-1653690 | 2 | 0.89 | 0.14 | 0.000513 | *PXDN* | exon_16 |
| chr3:58572616-58572648 | 2 | -0.63 | 0.1 | 0.000519 | | |
| chr11:64340850-64341078 | 3 | -0.38 | 0.062 | 0.00054 | *CCDC88B* | exon_3 |
| chr7:116511956-116511967 | 2 | 0.98 | 0.16 | 0.000545 | | |
| chr6:166244191-166244615 | 2 | 0.44 | 0.07 | 0.000548 | | |
| chr7:128211583-128211765 | 2 | -0.53 | 0.085 | 0.000549 | | |
| chr16:2235584-2235774 | 4 | 0.62 | 0.1 | 0.000555 | *E4F1* | 3UTR |
| chr17:80287993-80288408 | 2 | -0.31 | 0.051 | 0.000585 | *RNF213* | exon_4 |
| chr7:137953068-137953484 | 4 | 0.13 | 0.021 | 0.000593 | *:REB3L2-AS1* | TSS200 |
| chr10:98038638-98038658 | 2 | -0.79 | 0.13 | 0.000602 | | |
| chr6:33898332-33898539 | 4 | -0.48 | 0.078 | 0.000607 | *MIR7159* | TSS1500 |
| chr22:27803982-27804323 | 2 | -1.1 | 0.17 | 0.000608 | | |
| chr20:10604625-10604679 | 2 | -0.52 | 0.084 | 0.000621 | | |
| chr1:42152576-42152642 | 2 | 0.65 | 0.11 | 0.000646 | *GUCA2B* | TSS1500 |
| chr9:86078100-86078310 | 3 | -0.24 | 0.039 | 0.000648 | | |
| chr10:104334303-10433440ε | 2 | 0.66 | 0.11 | 0.00066 | *ITPRIP* | TSS1500 |
| chr2:42369080-42369111 | 2 | 0.83 | 0.13 | 0.00066 | *COX7A2L* | TSS200 |
| chr6:43776645-43776651 | 2 | 0.94 | 0.15 | 0.000661 | | |
| chr15:77161019-77161117 | 2 | -0.67 | 0.11 | 0.000664 | | |
| chr7:143392290-143392366 | 2 | -0.38 | 0.061 | 0.000666 | | |
| chr22:45893845-45893867 | 2 | -0.6 | 0.097 | 0.000668 | | |
| chr14:49479563-49479604 | 2 | 0.88 | 0.14 | 0.000683 | | |
| chr2:54663329-54663389 | 2 | -0.5 | 0.082 | 0.000689 | | |
| chr6:29631235-29631401 | 3 | -0.84 | 0.14 | 0.000697 | *GABBR1* | exon_3 |
| chr4:7281278-7281824 | 4 | -0.33 | 0.053 | 0.000706 | | |
| chr3:123993331-123993353 | 2 | -0.85 | 0.14 | 0.000711 | *ROPN1* | TSS1500 |
| chr2:174032094-174032268 | 2 | 0.46 | 0.074 | 0.000721 | | |
| chr1:23436698-23436786 | 3 | -0.71 | 0.12 | 0.000738 | | |
| chr2:20634864-20635267 | 2 | -0.74 | 0.12 | 0.000746 | | |
| chr4:25863556-25864073 | 7 | -0.66 | 0.11 | 0.000767 | *SEL1L3* | TSS1500 |
| chr11:68750061-68750331 | 3 | -0.59 | 0.096 | 0.000785 | *TESMIN* | exon_2 |
| chr8:95072593-95073021 | 2 | 0.34 | 0.056 | 0.000808 | *MIR3150BHG* | TSS200 |
| chr4:154465126-154465195 | 2 | 0.45 | 0.073 | 0.000813 | | |
| chr12:3152579-3153042 | 2 | -0.37 | 0.061 | 0.000816 | | |
| chr10:34937659-34938051 | 2 | -0.4 | 0.064 | 0.000819 | | |
| chr1:84398384-84398480 | 4 | -0.53 | 0.087 | 0.00084 | *DSE2B* | TSS200 |
| chr16:85464324-85464356 | 2 | -0.57 | 0.092 | 0.000847 | | |
| chr22:38751787-38751831 | 2 | 0.59 | 0.095 | 0.000857 | | |
| chr8:29927700-29927829 | 2 | -0.38 | 0.062 | 0.000857 | | |
| chr14:22514124-22514157 | 2 | 0.79 | 0.13 | 0.000875 | | |
| chr16:67399555-67399733 | 2 | -0.68 | 0.11 | 0.000903 | | |
| chr2:74375423-74375463 | 2 | -0.27 | 0.044 | 0.000907 | *DCTN1* | TSS1500 |
| chr1:27351524-27351647 | 2 | -0.62 | 0.1 | 0.000935 | *SYTL1* | exon_13 |
| chr11:12044928-12044941 | 2 | 0.42 | 0.069 | 0.000967 | | |
| chr20:52493482-52493501 | 2 | -1.1 | 0.18 | 0.000999 | | |
| chr13:68711798-68712237 | 5 | -0.8 | 0.13 | 0.00101 | | |
| chr22:30636199-30636236 | 2 | 0.61 | 0.1 | 0.00102 | *SLC35E4* | 5UTR |

| | | | | | | |
|---|---|---|---|---|---|---|
| chr11:63574384-63574604 | 2 | 0.88 | 0.14 | 0.00119 | *PLAAT3* | 3UTR |
| chr11:104059770-10406011€ | 2 | -0.88 | 0.14 | 0.0012 | | |
| chr2:120633298-120633708 | 2 | -0.64 | 0.1 | 0.00125 | | |
| chr16:89321803-89322431 | 4 | -0.59 | 0.097 | 0.00126 | | |
| chr9:132631477-132631607 | 2 | -0.58 | 0.096 | 0.00133 | | |
| chr2:10359339-10359402 | 2 | -0.65 | 0.11 | 0.00135 | | |
| chr11:67467567-67467604 | 2 | 0.44 | 0.073 | 0.00136 | *TMEM134* | exon_3 |
| chr2:38215321-38215456 | 2 | 0.9 | 0.15 | 0.0014 | | |
| chr16:85574618-85574899 | 2 | -0.72 | 0.12 | 0.00143 | | |
| chr1:155940732-155941176 | 4 | -0.61 | 0.1 | 0.00147 | *RXFP4* | TSS1500 |
| chr12:2364298-2364748 | 3 | -0.29 | 0.047 | 0.00147 | | |
| chr20:57390021-57390189 | 2 | -1.1 | 0.18 | 0.00149 | *RBM38* | TSS1500 |
| chr1:165910915-165911187 | 2 | 0.83 | 0.14 | 0.00153 | *UCK2* | 3UTR |
| chr1:119710775-119710854 | 2 | -0.61 | 0.1 | 0.00158 | *PHGDH* | TSS1500 |
| chr12:12534023-12534183 | 2 | -0.64 | 0.11 | 0.0016 | | |
| chr16:69328840-69329111 | 2 | 0.55 | 0.092 | 0.0016 | *COG8* | 3UTR |
| chr17:42509241-42509628 | 2 | 0.74 | 0.12 | 0.00161 | | |
| chr5:150377403-150377693 | 3 | -0.37 | 0.061 | 0.00171 | | |
| chr17:74417606-74417668 | 2 | 0.78 | 0.13 | 0.00174 | | |
| chr6:6971082-6971138 | 3 | 0.66 | 0.11 | 0.00178 | | |
| chr7:74027692-74027781 | 3 | 0.73 | 0.12 | 0.00182 | *ELN* | TSS1500 |
| chr7:374186-374190 | 2 | -0.8 | 0.13 | 0.00186 | | |
| chr11:119664055-11966452ž | 3 | -0.4 | 0.066 | 0.00189 | *NECTIN1* | 3UTR |
| chr6:44226768-44227012 | 2 | -0.28 | 0.046 | 0.0019 | | |
| chr11:62879399-62879805 | 3 | 0.56 | 0.094 | 0.0019 | *SLC3A2* | TSS1500 |
| chr8:30542068-30542147 | 2 | -0.7 | 0.12 | 0.0019 | | |
| chr18:8766441-8766463 | 2 | -0.44 | 0.074 | 0.00194 | | |
| chr12:65827887-65828009 | 2 | -0.49 | 0.082 | 0.00194 | *HMGA2* | exon_2 |
| chr1:160711934-160712218 | 4 | -0.35 | 0.059 | 0.00197 | *CD48* | TSS1500 |
| chr17:29576320-29576433 | 2 | -0.55 | 0.092 | 0.00199 | *GIT1* | exon_14 |
| chr6:33433703-33433743 | 3 | -0.74 | 0.12 | 0.00202 | | |
| chr20:57065893-57065918 | 2 | -0.77 | 0.13 | 0.00206 | | |
| chr7:1549956-1550220 | 4 | -0.63 | 0.11 | 0.00208 | *TMEM184A* | exon_5 |
| chr19:49511921-49512275 | 5 | -0.21 | 0.034 | 0.00225 | *FCGRT* | TSS1500 |
| chr17:897023-897521 | 7 | -0.33 | 0.055 | 0.00228 | | |
| chr15:70062308-70062486 | 3 | -0.31 | 0.052 | 0.00235 | | |
| chr10:86715642-86716014 | 2 | -0.68 | 0.11 | 0.00247 | | |
| chr16:11597803-11597814 | 2 | -0.56 | 0.094 | 0.00255 | | |
| chr11:79269794-79270014 | 2 | -0.73 | 0.12 | 0.00259 | | |
| chr19:11243320-11243456 | 2 | -0.78 | 0.13 | 0.00267 | *DOCK6* | exon_12 |
| chr17:55274659-55274707 | 2 | 0.64 | 0.11 | 0.00269 | | |
| chr16:79669891-79670023 | 2 | 0.74 | 0.13 | 0.00285 | | |
| chr16:67943962-67944089 | 2 | -0.78 | 0.13 | 0.00302 | *SLC12A4* | 3UTR |
| chr7:135147832-135148018 | 3 | 0.81 | 0.14 | 0.00316 | *TMEM140* | TSS200 |
| chr7:1013993-1014715 | 5 | -0.56 | 0.095 | 0.00321 | | |
| chr1:16978758-16979067 | 3 | -0.71 | 0.12 | 0.00323 | | |
| chr9:127950880-127950890 | 2 | -0.88 | 0.15 | 0.00323 | *FAM102A* | TSS1500 |
| chr22:19722173-19722640 | 4 | -0.52 | 0.088 | 0.00328 | *SEPTIN5* | exon_11 |
| chr11:64935720-64936031 | 6 | 0.16 | 0.027 | 0.00329 | *GPHA2* | TSS200 |
| chr9:137812926-137813762 | 3 | -0.47 | 0.08 | 0.00336 | | |
| chr11:69484017-69484056 | 2 | -0.52 | 0.089 | 0.00337 | | |
| chr15:52292882-52292893 | 2 | 0.93 | 0.16 | 0.00348 | | |
| chr1:56509911-56510017 | 2 | -0.81 | 0.14 | 0.00349 | | |
| chr14:22420767-22420905 | 2 | 0.39 | 0.066 | 0.00351 | | |
| chr17:77437111-77437324 | 2 | -0.82 | 0.14 | 0.00351 | | |
| chr6:3386350-3386623 | 2 | 0.52 | 0.089 | 0.00351 | | |
| chr10:72334403-72334414 | 2 | -0.62 | 0.1 | 0.00358 | *DJB12* | 3UTR |

| | | | | | | |
|---|---|---|---|---|---|---|
| chr2:207258358-207258757 | 2 | 0.51 | 0.086 | 0.0037 | | |
| chr11:65790924-65790943 | 2 | 0.12 | 0.02 | 0.00375 | *OVOL1-AS1* | TSS200 |
| chr4:6690686-6691252 | 4 | -0.54 | 0.091 | 0.00385 | *LINC02481* | TSS1500 |
| chr12:1474635-1474908 | 2 | -0.91 | 0.15 | 0.0039 | | |
| chr9:88990950-88991084 | 4 | -0.48 | 0.081 | 0.00392 | *S1PR3* | exon_1 |
| chr3:133455830-133456082 | 3 | -0.46 | 0.078 | 0.00395 | *BFSP2-AS1* | TSS200 |
| chr21:41142442-41142503 | 2 | 0.21 | 0.036 | 0.00398 | *LINC00323* | exon_2 |
| chr3:71893348-71893701 | 2 | -0.59 | 0.1 | 0.00398 | | |
| chr11:130653715-13065385 | 2 | 0.7 | 0.12 | 0.00413 | | |
| chr5:14174443-14174572 | 2 | -0.6 | 0.1 | 0.00415 | | |
| chr3:51974724-51974839 | 5 | 0.32 | 0.055 | 0.00423 | *ABHD14B* | TSS1500 |
| chr16:11355610-11355804 | 2 | -0.68 | 0.12 | 0.00428 | | |
| chr16:19874113-19874232 | 2 | 0.93 | 0.16 | 0.00434 | | |
| chr7:107937954-107938143 | 2 | -0.47 | 0.08 | 0.00439 | | |
| chr5:135330196-135330266 | 2 | -0.53 | 0.089 | 0.00441 | | |
| chr1:15214686-15214721 | 2 | -0.76 | 0.13 | 0.00443 | | |
| chr5:179803096-179803528 | 2 | 0.3 | 0.052 | 0.00461 | *MGAT4B* | TSS1500 |
| chr9:114395050-114395591 | 4 | -0.95 | 0.16 | 0.00473 | *AK* | TSS1500 |
| chr22:24432042-24432073 | 2 | 0.13 | 0.021 | 0.00476 | *ADORA2A* | TSS1500 |
| chr8:143463644-143463811 | 2 | -0.58 | 0.098 | 0.00479 | | |
| chr1:206770732-206770964 | 2 | -0.68 | 0.12 | 0.0048 | *IL10* | exon_1 |
| chr1:15761257-15761507 | 3 | -0.66 | 0.11 | 0.00489 | | |
| chr3:10290292-10290486 | 3 | -0.72 | 0.12 | 0.00493 | | |
| chr3:177349291-177349920 | 3 | -0.76 | 0.13 | 0.00494 | | |
| chr4:77021566-77022028 | 2 | -0.52 | 0.089 | 0.00495 | | |
| chr1:23169811-23170155 | 2 | 0.79 | 0.14 | 0.00504 | *LUZP1* | TSS1500 |
| chr17:57881211-57881439 | 2 | -0.73 | 0.13 | 0.00505 | | |
| chr12:47718466-47718575 | 2 | -0.7 | 0.12 | 0.00506 | | |
| chr19:13012403-13012642 | 3 | 0.71 | 0.12 | 0.00525 | | |
| chr11:77128159-77128171 | 2 | -0.81 | 0.14 | 0.00528 | *MYO7A* | TSS200 |
| chr3:194273990-194274437 | 2 | -0.42 | 0.072 | 0.00534 | | |
| chr20:25032709-25033282 | 8 | 0.19 | 0.032 | 0.00544 | *ACSS1* | TSS200 |
| chr16:85583786-85583852 | 2 | -0.84 | 0.14 | 0.00545 | | |
| chr8:127890392-127890397 | 2 | -0.77 | 0.13 | 0.00553 | | |
| chr20:25055961-25056035 | 2 | -0.62 | 0.11 | 0.00555 | | |
| chr9:95024492-95024597 | 2 | 0.8 | 0.14 | 0.00559 | | |
| chr11:72754743-72755068 | 4 | -0.57 | 0.098 | 0.0056 | *STARD10* | exon_7 |
| chr17:79951323-79951518 | 4 | -0.69 | 0.12 | 0.00584 | *TBC1D16* | TSS1500 |
| chr3:48504607-48504855 | 3 | 0.34 | 0.058 | 0.0061 | *SHISA5* | TSS1500 |
| chr2:202073169-202073250 | 2 | -0.64 | 0.11 | 0.00619 | *KIAA2012* | TSS200 |
| chr16:85444974-85445045 | 2 | -0.35 | 0.061 | 0.00622 | | |
| chr12:47777994-47778307 | 3 | 0.16 | 0.027 | 0.00632 | | |
| chr19:38952772-38953029 | 2 | -0.63 | 0.11 | 0.00633 | *FBXO17* | TSS1500 |
| chr1:33161508-33161554 | 2 | -0.81 | 0.14 | 0.00636 | | |
| chr17:320979-321084 | 3 | -0.52 | 0.09 | 0.00665 | | |
| chr15:36898347-36898804 | 6 | -0.94 | 0.16 | 0.00669 | | |
| chr20:38211884-38211943 | 2 | 0.25 | 0.043 | 0.00673 | *KIAA1755* | 3UTR |
| chr12:107580409-10758051 | 3 | 0.5 | 0.087 | 0.00677 | *BTBD11* | TSS1500 |
| chr4:153080007-153080045 | 2 | -0.67 | 0.12 | 0.00704 | | |
| chr12:121775009-12177504 | 2 | -0.59 | 0.1 | 0.00713 | | |
| chr17:38898096-38898473 | 4 | -0.75 | 0.13 | 0.00718 | *LASP1* | exon_3 |
| chr1:113979153-113979563 | 6 | -0.68 | 0.12 | 0.00724 | *OLFML3* | TSS200 |
| chr3:36908466-36908475 | 2 | 0.87 | 0.15 | 0.00726 | | |
| chr10:132685444-13268586 | 4 | -0.52 | 0.089 | 0.0074 | | |
| chr8:141287022-141287420 | 4 | -0.64 | 0.11 | 0.00745 | | |
| chr12:113160056-11316006 | 2 | -0.46 | 0.079 | 0.00753 | *MIR7106* | TSS1500 |
| chr15:90262121-90262375 | 2 | 0.42 | 0.072 | 0.00754 | *TTLL13* | exon_11 |

| | | | | | | |
|---|---|---|---|---|---|---|
| chr2:6960890-6960979 | 2 | 0.7 | 0.12 | 0.00755 | | |
| chr12:119678783-11967881€ | 2 | -0.96 | 0.17 | 0.00767 | | |
| chr8:27611464-27611484 | 2 | 0.72 | 0.12 | 0.00807 | *CLU* | TSS1500 |
| chr19:18650478-18650982 | 6 | -0.46 | 0.079 | 0.0082 | | |
| chr10:14572165-14572246 | 3 | 0.8 | 0.14 | 0.00824 | *FAM107B* | TSS200 |
| chr3:171466923-171466956 | 3 | -0.68 | 0.12 | 0.00846 | | |
| chr14:69766015-69766123 | 2 | -0.53 | 0.092 | 0.00872 | *SRSF5* | TSS1500 |
| chr22:30606959-30607240 | 7 | -0.72 | 0.13 | 0.00875 | *PES1* | TSS200 |
| chr12:49637557-49637783 | 2 | -0.64 | 0.11 | 0.00876 | *PRPF40B* | exon_17 |
| chr10:80357911-80358102 | 2 | 0.71 | 0.12 | 0.00881 | *DYDC1* | TSS1500 |
| chr2:133433913-133434389 | 3 | -0.69 | 0.12 | 0.00884 | | |
| chr17:75642324-75642331 | 2 | 0.53 | 0.093 | 0.00893 | | |
| chr1:159825472-159825752 | 4 | -0.49 | 0.086 | 0.00894 | *SLAMF8* | TSS1500 |
| chr1:154404888-154404903 | 2 | 0.87 | 0.15 | 0.009 | *IL6R* | TSS1500 |
| chr5:177539236-177539305 | 2 | 0.44 | 0.077 | 0.00916 | | |
| chr11:69654522-69654635 | 3 | 0.66 | 0.11 | 0.00925 | | |
| chr18:58628862-58629217 | 7 | -0.57 | 0.099 | 0.00936 | *ALPK2* | TSS200 |
| chr17:41308761-41309266 | 3 | 0.75 | 0.13 | 0.00958 | *KRTAP16-1* | exon_1 |
| chr7:1043573-1043648 | 3 | 0.24 | 0.041 | 0.00959 | *GPR146* | TSS1500 |
| chr3:53130946-53131334 | 2 | 0.5 | 0.086 | 0.00976 | *RFT1* | TSS1500 |
| chr15:29677027-29677516 | 4 | -0.59 | 0.1 | 0.00978 | *OC10013011* | exon_2 |
| chr17:81406600-81406693 | 2 | -0.37 | 0.065 | 0.00994 | | |
| chr16:56647955-56648033 | 2 | -0.58 | 0.1 | 0.01 | | |
| chr14:21525565-21525643 | 2 | -0.21 | 0.036 | 0.0101 | *SALL2* | exon_2 |
| chr17:50673001-50673066 | 2 | 0.65 | 0.11 | 0.0103 | *ABCC3* | exon_18 |
| chr10:24618421-24618701 | 2 | -0.51 | 0.089 | 0.0109 | | |
| chr1:44806589-44806961 | 2 | 0.79 | 0.14 | 0.011 | *DYNLT4* | exon_3 |
| chr16:27632279-27632371 | 3 | -0.35 | 0.061 | 0.0112 | | |
| chr14:77086862-77087063 | 3 | -0.53 | 0.092 | 0.0112 | | |
| chr2:31384501-31384949 | 2 | -0.77 | 0.13 | 0.0112 | | |
| chr22:45878236-45878600 | 2 | -0.69 | 0.12 | 0.0113 | | |
| chr7:99140991-99141375 | 3 | 0.37 | 0.065 | 0.0113 | | |
| chr17:49992681-49993335 | 3 | -0.33 | 0.057 | 0.0115 | | |
| chr11:44949220-44949498 | 2 | -0.78 | 0.14 | 0.0119 | | |
| chr11:69658486-69658948 | 4 | -0.47 | 0.082 | 0.012 | | |
| chr7:26441023-26441127 | 2 | -0.55 | 0.097 | 0.0122 | | |
| chr17:38415394-38415768 | 2 | -0.7 | 0.12 | 0.0123 | | |
| chr10:86967584-86967975 | 3 | 0.31 | 0.054 | 0.0123 | *ADIRF* | TSS1500 |
| chr5:59356776-59357122 | 4 | -0.8 | 0.14 | 0.0123 | *PDE4D* | TSS200 |
| chr4:41612208-41612380 | 2 | -0.48 | 0.084 | 0.0124 | *LIMCH1* | TSS1500 |
| chr2:105573410-105573470 | 2 | 0.78 | 0.14 | 0.0124 | | |
| chr17:4175967-4176011 | 2 | -0.46 | 0.08 | 0.0126 | | |
| chr9:88537610-88537970 | 2 | -0.52 | 0.092 | 0.0129 | | |
| chr11:45907254-45907347 | 5 | -0.18 | 0.031 | 0.013 | *C11orf94* | TSS200 |
| chr16:67438959-67439028 | 2 | 0.74 | 0.13 | 0.0131 | *ATP6V0D1* | exon_6 |
| chr2:9632246-9632459 | 2 | 0.1 | 0.018 | 0.0133 | *YWHAQ* | TSS1500 |
| chr11:69155893-69155897 | 2 | 0.95 | 0.17 | 0.0133 | *SMIM38* | TSS200 |
| chr4:1802280-1802765 | 4 | -0.44 | 0.077 | 0.0137 | | |
| chr9:136095962-136095972 | 2 | 0.56 | 0.098 | 0.0139 | *CC2* | TSS1500 |
| chr11:69471212-69471584 | 7 | -0.71 | 0.13 | 0.0139 | | |
| chr17:51398070-51398208 | 2 | -0.28 | 0.049 | 0.0141 | | |
| chr1:24352121-24352486 | 2 | -0.7 | 0.12 | 0.0142 | | |
| chr1:204359249-204359715 | 2 | -0.68 | 0.12 | 0.0145 | *PLEKHA6* | 5UTR |
| chr22:18966932-18966971 | 2 | 0.42 | 0.075 | 0.0147 | | |
| chr11:12242240-12242345 | 2 | -0.62 | 0.11 | 0.0148 | *MICAL2* | exon_19 |
| chr11:68118927-68119194 | 2 | -0.65 | 0.11 | 0.0148 | | |
| chr17:82863950-82864304 | 2 | -0.6 | 0.11 | 0.0149 | | |

| | | | | | | |
|---|---|---|---|---|---|---|
| chr16:3645594-3645717 | 2 | -0.53 | 0.094 | 0.015 | | |
| chr1:2505608-2505673 | 2 | -0.55 | 0.097 | 0.0151 | | |
| chr11:68042801-68043438 | 7 | -0.57 | 0.1 | 0.0158 | *TCIRG1* | 5UTR |
| chr11:78698811-78698891 | 2 | 0.47 | 0.083 | 0.016 | | |
| chr11:45156384-45156476 | 2 | -0.23 | 0.042 | 0.0164 | | |
| chr1:1434554-1434570 | 3 | -0.63 | 0.11 | 0.0173 | *LINC01770* | exon_1 |
| chr12:6959261-6959496 | 2 | -0.53 | 0.094 | 0.0174 | | |
| chr1:30879073-30879407 | 2 | -0.69 | 0.12 | 0.0174 | | |
| chr2:219460387-219460721 | 5 | -0.61 | 0.11 | 0.0175 | *SPEG* | TSS1500 |
| chr17:382816-383249 | 2 | -0.61 | 0.11 | 0.0176 | | |
| chr10:124059716-12405971[illegible] | 2 | -0.76 | 0.14 | 0.0182 | | |
| chr10:124307803-12430782[illegible] | 2 | -0.64 | 0.11 | 0.0185 | | |
| chr9:126386463-126386576 | 2 | 0.76 | 0.13 | 0.0188 | *MVB12B* | exon_4 |
| chr7:23287556-23287647 | 2 | -0.62 | 0.11 | 0.0199 | | |
| chr13:40562866-40563227 | 3 | -0.18 | 0.032 | 0.0204 | | |
| chr2:235577683-235577981 | 3 | -0.44 | 0.078 | 0.0206 | | |
| chr11:69464804-69464877 | 2 | 0.65 | 0.12 | 0.0209 | | |
| chr16:56789509-56789604 | 2 | 0.12 | 0.021 | 0.0212 | | |
| chr19:1851751-1852128 | 4 | -0.41 | 0.073 | 0.0212 | | |
| chr12:120759822-12075992[illegible] | 2 | -0.6 | 0.11 | 0.022 | | |
| chr1:64123325-64123678 | 2 | -0.49 | 0.087 | 0.0221 | | |
| chr8:139761715-139761777 | 2 | -0.52 | 0.093 | 0.0223 | | |
| chr19:42278372-42278440 | 2 | 0.37 | 0.066 | 0.0223 | | |
| chr8:49329325-49329381 | 2 | -0.82 | 0.15 | 0.0224 | | |
| chr4:37665553-37665961 | 2 | 0.69 | 0.12 | 0.0228 | | |
| chr13:94416716-94416968 | 2 | -0.78 | 0.14 | 0.0232 | | |
| chr1:24730553-24730573 | 2 | 0.36 | 0.064 | 0.0236 | | |
| chr5:93603-93613 | 2 | -0.67 | 0.12 | 0.0238 | | |
| chr20:2379739-2379750 | 2 | -0.9 | 0.16 | 0.0243 | *TGM6* | TSS1500 |
| chr22:45971737-45972145 | 3 | -0.98 | 0.17 | 0.0244 | | |
| chr20:44714724-44715095 | 4 | 0.19 | 0.034 | 0.0245 | *CCN5* | TSS1500 |
| chr5:77437389-77437459 | 2 | -0.72 | 0.13 | 0.0245 | *WDR41* | exon_11 |
| chr7:1063427-1063541 | 2 | 0.73 | 0.13 | 0.0246 | | |
| chr11:63837947-63838377 | 3 | 0.23 | 0.041 | 0.0246 | *MARK2* | TSS1500 |
| chr16:11299091-11299230 | 2 | -0.54 | 0.097 | 0.025 | | |
| chr16:87723412-87723427 | 2 | -0.55 | 0.098 | 0.0255 | | |
| chr6:28586485-28586557 | 3 | -0.17 | 0.031 | 0.0256 | *ZBED9* | exon_1 |
| chr1:41890733-41890996 | 2 | 0.56 | 0.1 | 0.0258 | | |
| chr6:1575051-1575751 | 3 | -0.41 | 0.074 | 0.0259 | | |
| chr11:68890188-68890397 | 2 | -0.49 | 0.087 | 0.0261 | | |
| chr20:37384193-37384543 | 3 | -0.53 | 0.095 | 0.0264 | *SRC* | exon_4 |
| chr4:1233303-1233888 | 3 | -0.53 | 0.095 | 0.0266 | | |
| chr12:93918393-93918629 | 2 | -0.78 | 0.14 | 0.0267 | | |
| chr8:9073199-9073275 | 2 | -0.68 | 0.12 | 0.027 | | |
| chr16:72931863-72931973 | 2 | -0.54 | 0.098 | 0.0271 | | |
| chr12:124501343-12450157[illegible] | 2 | -0.53 | 0.095 | 0.0271 | | |
| chr1:45863396-45863433 | 2 | -0.48 | 0.087 | 0.0275 | *MAST2* | TSS1500 |
| chr10:4383978-4384066 | 2 | -0.39 | 0.07 | 0.0276 | *LINC00703* | TSS1500 |
| chr16:85573956-85574045 | 2 | -0.75 | 0.13 | 0.0279 | | |
| chr22:29309055-29309167 | 2 | 0.48 | 0.086 | 0.028 | | |
| chr2:222404287-222404585 | 2 | -0.69 | 0.12 | 0.028 | | |
| chr5:67646803-67646855 | 2 | -0.82 | 0.15 | 0.0288 | *LINC02242* | TSS200 |
| chr2:23525234-23525373 | 2 | 0.51 | 0.092 | 0.0293 | | |
| chr16:68084088-68084337 | 4 | -0.42 | 0.076 | 0.0303 | *NFATC3* | TSS1500 |
| chr2:109447051-109447299 | 2 | -0.7 | 0.13 | 0.0303 | | |
| chr10:73915266-73915333 | 2 | 0.4 | 0.072 | 0.0304 | *PLAU* | exon_9 |
| chr9:95265134-95265435 | 2 | -0.77 | 0.14 | 0.0307 | | |

| | | | | | | |
|---|---|---|---|---|---|---|
| chr19:35119196-35119207 | 2 | 0.79 | 0.14 | 0.0312 | *FXYD3* | 5UTR |
| chr2:388372-388418 | 2 | -0.82 | 0.15 | 0.0312 | | |
| chr16:67179441-67179642 | 2 | 0.66 | 0.12 | 0.0315 | *KIAA0895L* | exon_5 |
| chr1:53512182-53512639 | 2 | -0.53 | 0.097 | 0.0317 | | |
| chr12:123268180-12326840[illegible] | 5 | -0.44 | 0.08 | 0.0318 | *CDK2AP1* | 5UTR |
| chr4:7056052-7056091 | 2 | 0.9 | 0.16 | 0.0321 | *TADA2B* | 3UTR |
| chr5:40478003-40478014 | 2 | 0.54 | 0.098 | 0.0323 | | |
| chr9:18976717-18976859 | 2 | -0.84 | 0.15 | 0.0325 | | |
| chr2:112977827-112977833 | 2 | -0.18 | 0.033 | 0.0333 | *IL36G* | TSS200 |
| chr10:33410290-33410472 | 2 | 0.4 | 0.072 | 0.0337 | | |
| chr3:50293347-50293571 | 2 | 0.74 | 0.13 | 0.0341 | *HYAL3* | exon_4 |
| chr12:47808069-47808221 | 2 | -0.65 | 0.12 | 0.0343 | | |
| chr19:3025667-3025687 | 2 | -0.76 | 0.14 | 0.0345 | | |
| chr1:116587802-116587999 | 2 | -0.64 | 0.12 | 0.0345 | | |
| chr7:55526994-55527447 | 3 | -0.38 | 0.068 | 0.0345 | *VOPP1* | TSS1500 |
| chr16:68995544-68995555 | 2 | 0.67 | 0.12 | 0.0349 | | |
| chr19:40802689-40802849 | 2 | 0.7 | 0.13 | 0.0353 | | |
| chr11:72014244-72014252 | 2 | 0.47 | 0.086 | 0.0353 | *NUMA1* | TSS200 |
| chr6:158649459-158649976 | 6 | 0.26 | 0.047 | 0.0358 | *SYTL3* | TSS1500 |
| chr12:53049273-53050093 | 6 | -0.45 | 0.082 | 0.0362 | *TNS2* | TSS1500 |
| chr12:49293018-49293042 | 2 | 0.76 | 0.14 | 0.0364 | *TROAP-AS1* | exon_8 |
| chr3:195258617-195258836 | 2 | -0.81 | 0.15 | 0.0371 | | |
| chr14:105466045-10546647[illegible] | 3 | -0.69 | 0.12 | 0.0372 | *MTA1* | exon_17 |
| chr10:125574192-12557420[illegible] | 2 | -0.46 | 0.084 | 0.0373 | *TEX36-AS1* | TSS200 |
| chr11:12862739-12863584 | 3 | -0.63 | 0.12 | 0.0377 | | |
| chr9:135886516-135886576 | 2 | -0.54 | 0.098 | 0.0378 | | |
| chr7:149855389-149855693 | 2 | -0.47 | 0.086 | 0.039 | | |
| chr13:46666243-46666471 | 2 | -0.55 | 0.1 | 0.0391 | | |
| chr15:99632020-99632335 | 3 | -0.46 | 0.084 | 0.0395 | | |
| chr2:240455896-240456870 | 5 | -0.6 | 0.11 | 0.04 | *OC10013044[illegible]* | TSS200 |
| chr15:67104931-67105356 | 3 | 0.36 | 0.065 | 0.0401 | | |
| chr11:48051925-48052147 | 2 | -0.58 | 0.11 | 0.0403 | | |
| chr15:70569136-70569401 | 2 | -0.43 | 0.078 | 0.0406 | | |
| chr1:85144591-85144774 | 2 | -0.69 | 0.13 | 0.0416 | | |
| chr22:30881629-30881705 | 2 | -0.26 | 0.047 | 0.0418 | *OSBP2* | 5UTR |
| chr5:131999624-131999759 | 3 | -0.31 | 0.057 | 0.0428 | | |
| chr11:64759826-64760374 | 7 | -0.3 | 0.054 | 0.0433 | *PYGM* | exon_1 |
| chr11:64102321-64102590 | 3 | -0.51 | 0.093 | 0.0433 | | |
| chr9:1979164-1979255 | 2 | 0.57 | 0.1 | 0.0434 | | |
| chr15:22917195-22917396 | 4 | -0.62 | 0.11 | 0.0435 | *CYFIP1* | TSS200 |
| chr13:23354949-23355068 | 3 | -0.79 | 0.14 | 0.0444 | *SACS* | exon_6 |
| chr22:27996236-27996318 | 2 | -0.55 | 0.1 | 0.0456 | *TTC28* | exon_16 |
| chr14:104725413-10472593[illegible] | 3 | -0.71 | 0.13 | 0.0465 | | |
| chr11:45907437-45907818 | 3 | 0.24 | 0.044 | 0.0472 | *C11orf94* | TSS1500 |
| chr1:200974808-200975179 | 2 | 0.42 | 0.077 | 0.0481 | *KIF21B* | exon_33 |
| chr11:62042241-62042334 | 2 | 0.67 | 0.12 | 0.0481 | | |
| chr14:103226763-10322693[illegible] | 2 | -0.43 | 0.079 | 0.0487 | | |
| chr13:29435819-29436258 | 2 | 0.69 | 0.13 | 0.0491 | | |
| chr16:3654677-3654800 | 2 | -0.75 | 0.14 | 0.0495 | *DSE1* | exon_1 |
| chr2:26978352-26978402 | 2 | -0.75 | 0.14 | 0.0497 | | |

**Supplementary Table 5**

Differentially methylated regions (DMRs) associated with early preterm birth (ePTB) in girls (N=134). DMRs were selected at regional Bonferroni-adjusted $p < 0.05$.

| DMR | #CpGs | Coefficient | Standard Error | *P* | Near Gene | Genomic context |
|---|---|---|---|---|---|---|
| chr20:1994000-1994726 | 8 | -0.39 | 0.02 | 6.36E-62 | *PDYN* | TSS1500 |
| chr16:85592811-85593506 | 3 | -0.56 | 0.03 | 4.37E-58 | | |
| chr11:2131198-2131347 | 2 | -1.3 | 0.08 | 3.12E-53 | *IGF2* | 3UTR |
| chr17:29572241-29572948 | 6 | -0.35 | 0.02 | 2.23E-48 | *TP53I13* | exon_6 |
| chr9:124800704-124801035 | 5 | 0.11 | 0.01 | 4.13E-44 | *OLFML2A* | TSS200 |
| chr17:82493190-82493473 | 2 | 0.62 | 0.05 | 3.43E-36 | | |
| chr7:100865886-100866137 | 3 | -0.049 | 0.00 | 3.65E-33 | *SLC12A9* | exon_14 |
| chr3:52402589-52402644 | 2 | -1.1 | 0.08 | 1.21E-32 | *BAP1* | exon_16 |
| chr11:118622428-118622681 | 2 | -0.35 | 0.03 | 9.15E-32 | | |
| chr20:46979526-46979898 | 3 | -0.44 | 0.04 | 1.15E-29 | | |
| chr4:56656822-56656889 | 2 | -0.59 | 0.05 | 1.34E-28 | *HOPX* | TSS1500 |
| chr4:3369261-3369825 | 7 | 0.098 | 0.01 | 1.41E-27 | *RGS12* | TSS1500 |
| chr6:30071626-30071699 | 7 | 0.091 | 0.01 | 6.77E-27 | *RNF39* | exon_4 |
| chr21:33419652-33419694 | 2 | -0.48 | 0.04 | 1.07E-26 | | |
| chr2:235088779-235088862 | 2 | -0.4 | 0.03 | 4.36E-26 | | |
| chr7:27144831-27144842 | 2 | 0.48 | 0.04 | 4.61E-26 | *HOXA5* | TSS1500 |
| chr3:44554868-44555020 | 3 | -0.099 | 0.01 | 6.69E-26 | *ZKSCAN7* | TSS1500 |
| chr2:219400113-219400672 | 4 | -0.54 | 0.05 | 7.12E-26 | *DNPEP* | TSS1500 |
| chr5:149605727-149605771 | 2 | -0.51 | 0.04 | 1.12E-25 | | |
| chr5:133266245-133266561 | 3 | -0.43 | 0.04 | 2.51E-25 | | |
| chr6:142302451-142302583 | 3 | 0.23 | 0.02 | 6.60E-25 | | |
| chr14:100680904-100681039 | 2 | -0.46 | 0.04 | 1.04E-24 | | |
| chr7:27144024-27144082 | 5 | 0.23 | 0.02 | 1.04E-24 | *HOXA5* | TSS1500 |
| chr10:944012-944512 | 2 | 0.61 | 0.05 | 1.69E-24 | | |
| chr3:51974522-51974638 | 2 | -0.76 | 0.07 | 2.06E-24 | *ABHD14B* | TSS1500 |
| chr5:135242665-135243162 | 2 | -0.69 | 0.06 | 3.06E-24 | | |
| chr19:42291387-42291640 | 3 | 0.27 | 0.02 | 3.63E-24 | *CIC* | exon_11 |
| chr7:129210434-129210518 | 3 | 0.092 | 0.01 | 1.42E-23 | *SMO* | exon_9 |
| chr22:46074136-46074311 | 2 | -0.18 | 0.02 | 3.04E-23 | | |
| chr16:85394241-85394489 | 2 | -1 | 0.09 | 3.04E-23 | | |
| chr16:69328840-69329333 | 3 | -0.22 | 0.02 | 7.55E-23 | *COG8* | 3UTR |
| chr2:157790278-157790283 | 2 | 0.56 | 0.05 | 1.25E-22 | | |
| chr6:125155085-125155261 | 3 | 0.086 | 0.01 | 2.13E-22 | | |
| chr11:2135464-2135469 | 2 | -1.3 | 0.12 | 2.69E-22 | *IGF2* | exon_2 |
| chr20:37383596-37384196 | 6 | -0.4 | 0.04 | 5.60E-22 | *SRC* | exon_4 |
| chr17:57408523-57408810 | 3 | -0.32 | 0.03 | 1.15E-21 | | |
| chr3:42703844-42704265 | 3 | -0.62 | 0.06 | 1.34E-21 | *HHATL* | TSS1500 |
| chr16:75099572-75099750 | 2 | 0.33 | 0.03 | 1.41E-21 | | |
| chr2:230925272-230926062 | 4 | -0.29 | 0.03 | 1.64E-21 | *GPR55* | TSS1500 |
| chr17:78159962-78160014 | 3 | -0.37 | 0.03 | 2.04E-21 | | |
| chr6:33278213-33278408 | 8 | 0.12 | 0.01 | 2.25E-21 | *B3GALT4* | exon_1 |
| chr16:57372381-57372483 | 3 | 0.11 | 0.01 | 2.26E-21 | *CX3CL1* | TSS200 |

| | | | | | | |
|---|---|---|---|---|---|---|
| chr14:70010013-70010249 | 3 | 0.055 | 0.01 | 2.53E-21 | | |
| chr8:41792158-41792441 | 3 | 0.34 | 0.03 | 5.09E-21 | | |
| chr6:168384844-168385215 | 3 | -0.19 | 0.02 | 5.09E-21 | | |
| chr11:34513734-34513830 | 3 | 0.0057 | 0.00 | 7.08E-21 | *ELF5* | 5UTR |
| chr2:25250912-25251458 | 3 | -0.77 | 0.07 | 8.39E-21 | | |
| chr14:64763904-64764049 | 3 | -0.85 | 0.08 | 1.06E-20 | | |
| chr2:95273650-95273919 | 3 | -0.9 | 0.09 | 2.00E-20 | *PROM2* | TSS1500 |
| chr13:32261649-32261655 | 2 | 0.52 | 0.05 | 2.22E-20 | *FRY* | exon_52 |
| chr6:32127130-32127371 | 4 | -0.22 | 0.02 | 2.66E-20 | *ATF6B* | exon_4 |
| chr12:89379267-89379535 | 2 | -0.36 | 0.03 | 2.82E-20 | | |
| chr6:112236066-112236302 | 4 | -0.48 | 0.05 | 3.82E-20 | *LAMA4-AS1* | TSS1500 |
| chr8:118024153-118024239 | 2 | -0.39 | 0.04 | 3.89E-20 | | |
| chr2:152486238-152486715 | 2 | 0.18 | 0.02 | 5.45E-20 | | |
| chr2:241763138-241763400 | 3 | -0.78 | 0.07 | 5.98E-20 | | |
| chr6:43510717-43510758 | 4 | 0.032 | 0.00 | 1.13E-19 | *LRRC73* | TSS1500 |
| chr20:63537226-63537441 | 5 | -0.032 | 0.00 | 1.73E-19 | *PTK6* | exon_1 |
| chr2:120232317-120232746 | 2 | -0.63 | 0.06 | 2.09E-19 | | |
| chr1:54153556-54153891 | 3 | -0.38 | 0.04 | 3.25E-19 | *CDCP2* | TSS1500 |
| chr12:121958727-121958920 | 4 | 0.09 | 0.01 | 3.31E-19 | | |
| chr15:39976220-39976486 | 3 | 0.42 | 0.04 | 3.86E-19 | *EIF2AK4* | exon_12 |
| chr10:26437875-26438389 | 12 | 0.02 | 0.00 | 4.34E-19 | *APBB1IP* | TSS1500 |
| chr12:48818899-48819298 | 3 | -1.6 | 0.15 | 7.80E-19 | *CACNB3* | 5UTR |
| chr14:65097057-65097199 | 2 | -0.62 | 0.06 | 9.34E-19 | | |
| chr6:37391018-37391086 | 2 | -0.96 | 0.09 | 1.15E-18 | *RNF8* | 3UTR |
| chr3:50268588-50268675 | 3 | 0.089 | 0.01 | 1.17E-18 | *SEMA3B* | TSS1500 |
| chr14:99412862-99413316 | 2 | -0.61 | 0.06 | 1.20E-18 | | |
| chr11:72014244-72014294 | 3 | -1.1 | 0.11 | 2.22E-18 | *NUMA1* | exon_15 |
| chr16:11355610-11355804 | 2 | -0.64 | 0.06 | 2.37E-18 | | |
| chr13:50386833-50386898 | 2 | -0.68 | 0.07 | 2.39E-18 | | |
| chr10:89645928-89646043 | 3 | -0.56 | 0.06 | 2.92E-18 | *PANK1* | TSS1500 |
| chr7:116733589-116734176 | 3 | -0.45 | 0.05 | 3.10E-18 | | |
| chr1:157099450-157099819 | 3 | -0.13 | 0.01 | 5.44E-18 | *ETV3L* | 5UTR |
| chr11:45907347-45907437 | 2 | -0.6 | 0.06 | 7.33E-18 | *C11orf94* | TSS200 |
| chr2:232316147-232316640 | 2 | -0.38 | 0.04 | 9.06E-18 | | |
| chr7:56092716-56092994 | 3 | -0.42 | 0.04 | 1.45E-17 | *PHKG1* | TSS200 |
| chr9:92974381-92974448 | 4 | 0.091 | 0.01 | 1.88E-17 | *FGD3* | TSS200 |
| chr5:157266773-157266804 | 2 | -0.78 | 0.08 | 2.02E-17 | | |
| chr11:126355090-126355381 | 2 | -0.96 | 0.10 | 3.42E-17 | *ST3GAL4* | TSS1500 |
| chr5:1089392-1089607 | 4 | -0.67 | 0.07 | 3.86E-17 | | |
| chr16:56607035-56607189 | 2 | -0.47 | 0.05 | 4.07E-17 | *MT2A* | TSS1500 |
| chr19:6022297-6023014 | 4 | -0.83 | 0.08 | 4.11E-17 | | |
| chr1:156152872-156153029 | 4 | -0.17 | 0.02 | 4.27E-17 | *SEMA4A* | TSS1500 |
| chr8:42208530-42208552 | 2 | -0.085 | 0.01 | 7.01E-17 | *PLAT* | TSS1500 |
| chr4:1221324-1221555 | 2 | -0.05 | 0.01 | 7.10E-17 | | |
| chr22:28917125-28917247 | 2 | -0.44 | 0.04 | 8.69E-17 | *ZNRF3* | TSS1500 |
| chr1:51331278-51331521 | 3 | 0.1 | 0.01 | 1.42E-16 | *TTC39A* | TSS1500 |
| chr11:47378447-47378562 | 2 | -0.67 | 0.07 | 1.64E-16 | *SPI1* | 5UTR |
| chr1:40396308-40396568 | 2 | -0.71 | 0.07 | 1.68E-16 | *SMAP2* | TSS1500 |

| | | | | | | |
|---|---|---|---|---|---|---|
| chr13:43405346-43405726 | 2 | 0.43 | 0.04 | 2.19E-16 | | |
| chr17:50673001-50673066 | 2 | -0.58 | 0.06 | 3.02E-16 | *ABCC3* | exon_18 |
| chr9:136373776-136374446 | 4 | -0.24 | 0.03 | 3.30E-16 | *CARD9* | TSS1500 |
| chr6:135850691-135851088 | 2 | 0.49 | 0.05 | 3.65E-16 | *PDE7B* | TSS1500 |
| chr19:12969407-12969929 | 4 | -0.46 | 0.05 | 3.90E-16 | *DAND5* | exon_1 |
| chr7:32163264-32163681 | 3 | -0.026 | 0.00 | 3.98E-16 | | |
| chr1:116466343-116466465 | 3 | -0.73 | 0.08 | 3.99E-16 | | |
| chr2:42567259-42568122 | 7 | 0.053 | 0.01 | 4.27E-16 | *MTA3* | TSS1500 |
| chr18:31590613-31590641 | 2 | 0.4 | 0.04 | 4.40E-16 | *TTR* | TSS1500 |
| chr2:74375423-74375463 | 2 | 0.17 | 0.02 | 4.58E-16 | *DCTN1* | TSS1500 |
| chr1:109371049-109371117 | 2 | -0.41 | 0.04 | 4.65E-16 | | |
| chr11:67437695-67437820 | 3 | 0.083 | 0.01 | 5.29E-16 | *CORO1B* | 3UTR |
| chr11:61627992-61628317 | 2 | -0.63 | 0.07 | 8.69E-16 | | |
| chr2:10039880-10040161 | 2 | -0.46 | 0.05 | 1.14E-15 | | |
| chr5:172150951-172151087 | 3 | 0.13 | 0.01 | 1.16E-15 | | |
| chr12:7060525-7060553 | 2 | -0.18 | 0.02 | 1.69E-15 | *C1S* | TSS200 |
| chr7:836162-836508 | 2 | -0.38 | 0.04 | 1.92E-15 | | |
| chr11:67438963-67439292 | 2 | -1.4 | 0.15 | 2.38E-15 | *PTPRCAP* | TSS1500 |
| chr1:85578151-85578493 | 4 | 0.17 | 0.02 | 2.69E-15 | *DDAH1* | TSS1500 |
| chr17:57904281-57904744 | 2 | -0.93 | 0.10 | 3.81E-15 | *CUEDC1* | TSS1500 |
| chr4:6633392-6633758 | 2 | -0.28 | 0.03 | 4.60E-15 | | |
| chr2:60353123-60353424 | 4 | 0.11 | 0.01 | 4.71E-15 | | |
| chr17:7383952-7384130 | 2 | 0.62 | 0.07 | 6.22E-15 | *TNK1* | exon_6 |
| chr1:2208218-2208508 | 3 | 0.072 | 0.01 | 6.69E-15 | *FAAP20* | TSS1500 |
| chr14:53615416-53615509 | 2 | -0.47 | 0.05 | 7.75E-15 | | |
| chr11:2141253-2141352 | 5 | -0.68 | 0.07 | 8.47E-15 | *IGF2* | TSS1500 |
| chr1:3689183-3689518 | 5 | -0.49 | 0.05 | 9.76E-15 | *TP73* | TSS1500 |
| chr13:50335014-50335139 | 2 | -0.91 | 0.10 | 1.47E-14 | | |
| chr14:100842584-100843011 | 2 | 0.36 | 0.04 | 3.45E-14 | | |
| chr10:19987108-19987116 | 2 | -0.3 | 0.03 | 3.69E-14 | | |
| chr6:158738149-158738402 | 2 | -0.4 | 0.04 | 3.94E-14 | | |
| chr9:113593778-113593967 | 3 | -1.2 | 0.13 | 3.99E-14 | *RGS3* | TSS1500 |
| chr12:18282440-18282644 | 2 | -0.4 | 0.04 | 4.24E-14 | *PIK3C2G* | exon_2 |
| chr6:32167464-32167508 | 2 | -0.67 | 0.07 | 4.33E-14 | *EGFL8* | exon_8 |
| chr7:137344226-137344351 | 2 | -0.45 | 0.05 | 4.87E-14 | *PTN* | TSS1500 |
| chr1:18927200-18927378 | 2 | 0.16 | 0.02 | 8.57E-14 | | |
| chr7:100866931-100867336 | 6 | -0.55 | 0.06 | 1.00E-13 | *SLC12A9* | 3UTR |
| chr2:98508407-98508810 | 3 | 0.069 | 0.01 | 1.02E-13 | | |
| chr7:23347746-23347764 | 2 | 0.87 | 0.10 | 1.11E-13 | | |
| chr9:114394603-114395050 | 3 | -0.12 | 0.01 | 1.12E-13 | *AK* | TSS1500 |
| chr22:42931092-42931594 | 4 | -0.098 | 0.01 | 1.18E-13 | | |
| chr11:63016071-63016372 | 2 | -0.81 | 0.09 | 1.36E-13 | *SLC22A8* | TSS1500 |
| chr11:35585394-35585439 | 2 | 0.33 | 0.04 | 1.40E-13 | | |
| chr20:36646252-36646312 | 2 | -0.28 | 0.03 | 1.41E-13 | *SLA2* | TSS200 |
| chr11:64268316-64268700 | 2 | -0.48 | 0.05 | 1.63E-13 | *PLCB3* | 3UTR |
| chr22:27996318-27996597 | 2 | -0.6 | 0.07 | 1.66E-13 | | |
| chr7:114082703-114083043 | 5 | 0.014 | 0.00 | 1.72E-13 | | |
| chr11:69445189-69445368 | 2 | -0.26 | 0.03 | 2.08E-13 | | |

| | | | | | | |
|---|---|---|---|---|---|---|
| chr16:31457989-31458051 | 3 | 0.012 | 0.00 | 2.34E-13 | *ARMC5* | TSS1500 |
| chr19:14480391-14480489 | 2 | -2 | 0.23 | 2.40E-13 | *GIPC1* | exon_4 |
| chr9:136694503-136694536 | 3 | 0.087 | 0.01 | 2.59E-13 | | |
| chr1:228650240-228650548 | 3 | 0.23 | 0.03 | 2.60E-13 | *DUSP5P1* | exon_2 |
| chr20:36645878-36646192 | 3 | -0.62 | 0.07 | 2.70E-13 | *SLA2* | 5UTR |
| chr1:184806311-184806565 | 2 | -1.1 | 0.12 | 2.78E-13 | | |
| chr5:150377616-150378423 | 4 | -0.64 | 0.07 | 2.88E-13 | | |
| chr19:1854634-1854820 | 3 | -0.077 | 0.01 | 3.30E-13 | *KLF16* | exon_2 |
| chr10:133237752-133237809 | 5 | 0.26 | 0.03 | 3.44E-13 | *VENTX* | TSS200 |
| chr2:120710287-120710567 | 3 | 0.12 | 0.01 | 3.54E-13 | | |
| chr12:65827887-65828009 | 2 | -0.6 | 0.07 | 3.63E-13 | *HMGA2* | exon_2 |
| chr21:17794010-17794359 | 2 | -0.28 | 0.03 | 4.34E-13 | | |
| chr1:6519855-6520248 | 4 | -0.36 | 0.04 | 4.53E-13 | *PLEKHG5* | 5UTR |
| chr17:82331665-82331755 | 2 | -0.67 | 0.08 | 5.29E-13 | | |
| chr11:119664055-119664444 | 2 | -0.75 | 0.08 | 5.58E-13 | *NECTIN1* | 3UTR |
| chr12:47896219-47896478 | 2 | -0.7 | 0.08 | 5.62E-13 | | |
| chr12:104671250-104671608 | 2 | 0.14 | 0.02 | 6.97E-13 | | |
| chr9:123033004-123033482 | 4 | -0.89 | 0.10 | 7.06E-13 | *GPR21* | TSS1500 |
| chr6:106590795-106590809 | 2 | -0.6 | 0.07 | 7.65E-13 | | |
| chr5:138963988-138964230 | 2 | -1 | 0.12 | 7.86E-13 | | |
| chr3:45554008-45554498 | 3 | -0.19 | 0.02 | 9.22E-13 | | |
| chr11:63504753-63505082 | 2 | -0.051 | 0.01 | 1.20E-12 | *LGALS12* | TSS1500 |
| chr17:74211379-74211456 | 2 | -0.96 | 0.11 | 1.26E-12 | *MGC16275* | exon_2 |
| chr9:136882477-136882661 | 2 | -0.74 | 0.08 | 1.71E-12 | | |
| chr1:43934104-43934197 | 3 | -0.5 | 0.06 | 1.88E-12 | *ARTN* | 5UTR |
| chr16:2687219-2687644 | 4 | -0.016 | 0.00 | 2.02E-12 | | |
| chr11:2533332-2533934 | 4 | -0.43 | 0.05 | 2.04E-12 | | |
| chr16:85485570-85485572 | 2 | -1.4 | 0.16 | 2.21E-12 | | |
| chr2:85391562-85392044 | 2 | -0.99 | 0.11 | 2.35E-12 | *ELMOD3* | 3UTR |
| chr3:37693736-37694110 | 2 | -1.1 | 0.12 | 2.36E-12 | | |
| chr1:228143380-228143414 | 2 | -0.83 | 0.10 | 2.63E-12 | *GUK1* | TSS1500 |
| chr11:34514021-34514370 | 3 | -0.19 | 0.02 | 3.58E-12 | *ELF5* | TSS1500 |
| chr3:52771091-52771320 | 4 | 0.18 | 0.02 | 3.85E-12 | *NEK4* | TSS1500 |
| chr12:117213246-117213494 | 2 | -0.38 | 0.04 | 3.87E-12 | *NOS1* | 3UTR |
| chr11:129313333-129313543 | 3 | 0.14 | 0.02 | 4.11E-12 | | |
| chr6:11849449-11849805 | 2 | -0.83 | 0.10 | 4.20E-12 | | |
| chr19:19460942-19461133 | 2 | -1 | 0.12 | 4.23E-12 | | |
| chr2:42566377-42567220 | 3 | -0.54 | 0.06 | 4.34E-12 | *MTA3* | TSS1500 |
| chr9:34624163-34624590 | 2 | -0.3 | 0.04 | 4.41E-12 | | |
| chr17:81821712-81821724 | 3 | -0.1 | 0.01 | 4.90E-12 | | |
| chr11:62610951-62611128 | 2 | -0.34 | 0.04 | 5.42E-12 | *EML3* | exon_3 |
| chr3:194247181-194247477 | 2 | -0.29 | 0.03 | 6.06E-12 | *LINC02037* | TSS1500 |
| chr17:80187680-80187696 | 2 | -0.046 | 0.01 | 6.14E-12 | *CARD14* | TSS200 |
| chr5:10761565-10761982 | 3 | -1.1 | 0.12 | 6.22E-12 | *DAP* | TSS1500 |
| chr12:6375355-6375547 | 3 | 0.03 | 0.00 | 6.74E-12 | *SCNN1A* | 5UTR |
| chr11:1750776-1751238 | 2 | -0.68 | 0.08 | 7.02E-12 | *IFITM10* | TSS1500 |
| chr5:181198956-181199103 | 2 | -1 | 0.12 | 7.26E-12 | *TRIM7* | exon_4 |
| chr12:121132813-121132949 | 3 | -0.81 | 0.09 | 7.41E-12 | *P2RX7* | TSS200 |

| | | | | | | |
|---|---|---|---|---|---|---|
| chr1:38476641-38476972 | 4 | -0.61 | 0.07 | 7.46E-12 | | |
| chr15:74203262-74203296 | 2 | -0.7 | 0.08 | 7.52E-12 | *STRA6* | TSS1500 |
| chr1:164589061-164589232 | 3 | 0.086 | 0.01 | 7.58E-12 | | |
| chr11:69646468-69646566 | 2 | -0.84 | 0.10 | 8.10E-12 | | |
| chr16:2039896-2040330 | 3 | -0.46 | 0.05 | 8.21E-12 | *NTHL1* | 3UTR |
| chr22:31223651-31224027 | 3 | -0.42 | 0.05 | 8.43E-12 | | |
| chr17:82449873-82449903 | 3 | -0.4 | 0.05 | 8.81E-12 | *CYBC1* | TSS200 |
| chr20:38805160-38805515 | 3 | 0.31 | 0.04 | 9.00E-12 | *PPP1R16B* | TSS1500 |
| chr14:75459517-75459727 | 2 | -0.66 | 0.08 | 1.00E-11 | | |
| chr11:64283580-64283740 | 2 | -0.18 | 0.02 | 1.03E-11 | *GPR137* | TSS1500 |
| chr12:52010546-52010638 | 2 | 2.1 | 0.25 | 1.10E-11 | *TAMALIN* | 5UTR |
| chr1:46449858-46449869 | 2 | -1.4 | 0.16 | 1.12E-11 | *LINC01398* | TSS200 |
| chr8:22052243-22052879 | 3 | -0.31 | 0.04 | 1.21E-11 | *DMTN* | TSS1500 |
| chr6:33315381-33315407 | 3 | -0.36 | 0.04 | 1.22E-11 | *TAPBP* | TSS1500 |
| chr11:124897929-124898078 | 4 | 0.012 | 0.00 | 1.30E-11 | *ROBO4* | TSS200 |
| chr17:81406600-81406906 | 5 | -0.33 | 0.04 | 1.37E-11 | | |
| chr2:216809774-216809943 | 2 | -1.3 | 0.15 | 1.51E-11 | | |
| chr16:2088218-2088380 | 3 | 0.097 | 0.01 | 1.53E-11 | *TSC2* | exon_40 |
| chr15:90063689-90064172 | 2 | -0.65 | 0.08 | 1.56E-11 | | |
| chr11:72012929-72013241 | 2 | -0.34 | 0.04 | 1.85E-11 | *NUMA1* | TSS1500 |
| chr2:223538904-223539368 | 2 | 0.61 | 0.07 | 1.96E-11 | | |
| chr12:50073495-50073517 | 2 | 0.19 | 0.02 | 2.16E-11 | *ASIC1* | TSS200 |
| chr2:216607062-216607183 | 2 | -0.68 | 0.08 | 2.19E-11 | *LINC01280* | TSS1500 |
| chr7:74094538-74094664 | 2 | -0.48 | 0.06 | 2.20E-11 | | |
| chr12:49882729-49883201 | 2 | -0.66 | 0.08 | 2.34E-11 | | |
| chr12:3152579-3153042 | 2 | -0.32 | 0.04 | 2.57E-11 | | |
| chr19:11194555-11195237 | 3 | -0.32 | 0.04 | 2.71E-11 | *KANK2* | 5UTR |
| chr6:30954712-30955026 | 3 | -0.94 | 0.11 | 2.95E-11 | *HCG21* | TSS200 |
| chr17:38508820-38508888 | 2 | -1.7 | 0.20 | 3.35E-11 | | |
| chr8:141219662-141220134 | 4 | -0.79 | 0.09 | 3.61E-11 | *SLC45A4* | exon_3 |
| chr9:134410300-134410385 | 2 | -0.76 | 0.09 | 3.86E-11 | | |
| chr2:128362318-128362748 | 2 | 1.7 | 0.20 | 4.04E-11 | | |
| chr12:81079032-81079455 | 3 | -0.55 | 0.07 | 5.10E-11 | | |
| chr14:50397527-50397697 | 6 | -0.36 | 0.04 | 5.41E-11 | *CDKL1* | TSS1500 |
| chr11:66721167-66721382 | 3 | -0.88 | 0.10 | 6.00E-11 | *SPTBN2* | 5UTR |
| chr5:167754452-167754810 | 4 | -0.45 | 0.05 | 7.08E-11 | *TENM2* | TSS1500 |
| chr11:2301572-2302017 | 6 | 0.17 | 0.02 | 7.90E-11 | *C11orf21* | exon_1 |
| chr11:118605314-118605802 | 2 | -1.2 | 0.14 | 7.94E-11 | *PHLDB1* | TSS1500 |
| chr7:923414-923697 | 2 | -0.63 | 0.08 | 8.98E-11 | | |
| chr20:50019658-50019933 | 2 | -0.27 | 0.03 | 9.21E-11 | | |
| chr10:119516591-119516808 | 2 | -0.73 | 0.09 | 9.62E-11 | | |
| chr16:89694725-89694787 | 2 | 0.6 | 0.07 | 1.01E-10 | *CDK10* | exon_10 |
| chr11:2533973-2534267 | 2 | -0.35 | 0.04 | 1.03E-10 | | |
| chr1:156509810-156509834 | 2 | -0.88 | 0.11 | 1.06E-10 | | |
| chr8:96328042-96328065 | 2 | -0.72 | 0.09 | 1.16E-10 | | |
| chr6:30207528-30207550 | 2 | -0.38 | 0.05 | 1.25E-10 | | |
| chr3:184317390-184317731 | 3 | -0.29 | 0.04 | 1.34E-10 | *EIF4G1* | exon_6 |
| chr17:964542-965041 | 2 | -0.74 | 0.09 | 1.38E-10 | | |

| | | | | | | |
|---|---|---|---|---|---|---|
| chr10:48469913-48470315 | 2 | -0.34 | 0.04 | 1.46E-10 | | |
| chr3:171306918-171307052 | 2 | -0.3 | 0.04 | 1.50E-10 | | |
| chr4:1365350-1365769 | 2 | -0.59 | 0.07 | 1.59E-10 | | |
| chr10:329451-329751 | 2 | -0.074 | 0.01 | 1.70E-10 | *DIP2C* | exon_30 |
| chr6:32081400-32081845 | 7 | -0.44 | 0.05 | 1.77E-10 | *TNXB* | exon_10 |
| chr17:9179427-9179617 | 2 | 0.72 | 0.09 | 1.81E-10 | *.OC10192826(* | TSS1500 |
| chr11:73214443-73214807 | 2 | -0.47 | 0.06 | 2.00E-10 | | |
| chr17:75133132-75133135 | 2 | -0.79 | 0.10 | 2.29E-10 | *NT5C* | TSS1500 |
| chr4:185820293-185820896 | 4 | -0.55 | 0.07 | 2.38E-10 | | |
| chr14:22784024-22784271 | 2 | -0.28 | 0.03 | 2.45E-10 | | |
| chr8:143470616-143470740 | 2 | -0.65 | 0.08 | 2.45E-10 | | |
| chr17:57750677-57750746 | 3 | 0.3 | 0.04 | 3.08E-10 | | |
| chr17:81994652-81994985 | 2 | -0.64 | 0.08 | 3.17E-10 | | |
| chr2:382090-382237 | 2 | -0.54 | 0.07 | 3.20E-10 | | |
| chr11:65386357-65386371 | 2 | -0.74 | 0.09 | 3.32E-10 | *FRMD8* | TSS1500 |
| chr9:75911039-75911409 | 2 | -0.44 | 0.05 | 4.04E-10 | | |
| chr11:35040217-35040699 | 3 | -0.26 | 0.03 | 4.07E-10 | | |
| chr5:10456922-10456967 | 2 | -0.37 | 0.05 | 4.13E-10 | | |
| chr6:30688093-30688247 | 3 | -0.25 | 0.03 | 4.33E-10 | *NRM* | 3UTR |
| chr17:82012259-82012316 | 2 | -0.53 | 0.07 | 4.82E-10 | *ASPSCR1* | exon_11 |
| chr15:81225235-81225340 | 2 | -0.44 | 0.05 | 5.30E-10 | *IL16* | exon_2 |
| chr9:128719478-128719488 | 2 | -0.33 | 0.04 | 6.14E-10 | | |
| chr19:4911046-4911463 | 2 | -1.6 | 0.20 | 6.22E-10 | | |
| chr9:84371964-84371972 | 2 | -0.42 | 0.05 | 6.70E-10 | | |
| chr11:3166520-3166560 | 5 | -0.16 | 0.02 | 6.72E-10 | *OSBPL5* | TSS1500 |
| chr11:1881879-1881946 | 2 | -0.7 | 0.09 | 6.81E-10 | | |
| chr11:112286226-112286518 | 2 | -0.37 | 0.05 | 7.10E-10 | | |
| chr1:184555875-184556014 | 2 | -0.59 | 0.07 | 7.68E-10 | | |
| chr8:1897084-1897278 | 2 | -0.69 | 0.09 | 7.75E-10 | | |
| chr17:8056589-8056774 | 2 | -0.23 | 0.03 | 7.86E-10 | | |
| chr14:99807778-99808216 | 2 | -0.88 | 0.11 | 9.47E-10 | | |
| chr17:55320554-55320712 | 2 | 0.58 | 0.07 | 9.64E-10 | *HLF* | exon_4 |
| chr10:70118351-70118356 | 2 | -0.64 | 0.08 | 9.78E-10 | | |
| chr6:29680498-29680571 | 2 | 0.13 | 0.02 | 9.91E-10 | | |
| chr17:29076870-29077108 | 3 | 0.59 | 0.07 | 1.01E-09 | *TIAF1* | TSS1500 |
| chr12:116318089-116318480 | 3 | -0.31 | 0.04 | 1.04E-09 | | |
| chr8:27838013-27838150 | 4 | 0.34 | 0.04 | 1.13E-09 | *PBK* | TSS1500 |
| chr2:169080584-169080714 | 2 | -0.4 | 0.05 | 1.13E-09 | *DHRS9* | TSS1500 |
| chr19:16326246-16326551 | 2 | -0.75 | 0.09 | 1.16E-09 | | |
| chr19:45117494-45117849 | 2 | 0.24 | 0.03 | 1.16E-09 | | |
| chr22:30881475-30881503 | 2 | -0.69 | 0.09 | 1.20E-09 | *OSBP2* | TSS1500 |
| chr14:22514124-22514157 | 2 | -0.41 | 0.05 | 1.22E-09 | | |
| chr17:36616174-36616560 | 2 | 0.12 | 0.02 | 1.25E-09 | | |
| chr1:2220868-2221241 | 5 | 0.055 | 0.01 | 1.28E-09 | | |
| chr12:92147599-92147678 | 2 | -0.79 | 0.10 | 1.35E-09 | | |
| chr20:32205215-32205551 | 2 | -1.4 | 0.17 | 1.37E-09 | | |
| chr7:105679039-105679372 | 7 | -0.17 | 0.02 | 1.38E-09 | *ATXN7L1* | TSS200 |
| chr19:15464841-15464874 | 2 | -1 | 0.13 | 1.45E-09 | *RASAL3* | TSS1500 |

| | | | | | | |
|---|---|---|---|---|---|---|
| chr1:173207343-173207384 | 3 | 0.063 | 0.01 | 1.51E-09 | *TNFSF4* | TSS200 |
| chr17:7080491-7080868 | 4 | -0.22 | 0.03 | 1.51E-09 | *CLEC10A* | TSS1500 |
| chr17:78774816-78775241 | 3 | -0.26 | 0.03 | 1.60E-09 | | |
| chr19:46303164-46303413 | 2 | -0.45 | 0.06 | 1.62E-09 | *HIF3A* | TSS1500 |
| chr1:91165386-91165765 | 3 | -0.96 | 0.12 | 1.63E-09 | | |
| chr16:2324235-2324473 | 2 | -1.4 | 0.18 | 1.69E-09 | *ABCA3* | exon_6 |
| chr12:59085002-59085176 | 2 | 0.84 | 0.11 | 1.91E-09 | | |
| chr13:40598852-40599303 | 2 | -0.31 | 0.04 | 1.94E-09 | | |
| chr17:49972606-49972615 | 3 | -0.27 | 0.03 | 1.95E-09 | *DLX4* | TSS200 |
| chr12:2364298-2364748 | 3 | -0.32 | 0.04 | 2.04E-09 | | |
| chr8:35377711-35378076 | 2 | -0.58 | 0.07 | 2.08E-09 | | |
| chr1:100719741-100719755 | 3 | 0.46 | 0.06 | 2.11E-09 | *VCAM1* | 5UTR |
| chr9:134404271-134404473 | 2 | -0.46 | 0.06 | 2.19E-09 | | |
| chr1:61851449-61851574 | 2 | -0.55 | 0.07 | 2.22E-09 | | |
| chr8:23538860-23538890 | 2 | -0.38 | 0.05 | 2.30E-09 | | |
| chr3:197520880-197520998 | 2 | -0.36 | 0.05 | 2.31E-09 | | |
| chr5:172800142-172800617 | 2 | -0.074 | 0.01 | 2.43E-09 | | |
| chr19:18649768-18649890 | 2 | -0.33 | 0.04 | 2.44E-09 | | |
| chr16:70634969-70635211 | 2 | -0.44 | 0.06 | 2.45E-09 | | |
| chr17:28712114-28712505 | 3 | -0.39 | 0.05 | 2.47E-09 | *PROCA1* | TSS1500 |
| chr16:74741321-74741382 | 2 | 1 | 0.13 | 2.69E-09 | | |
| chr22:21937278-21937582 | 2 | -0.76 | 0.10 | 2.74E-09 | *PPM1F-AS1* | TSS1500 |
| chr11:90134217-90134252 | 2 | 0.22 | 0.03 | 2.87E-09 | *ALAD2* | TSS1500 |
| chr11:62426707-62426730 | 2 | -0.47 | 0.06 | 2.90E-09 | | |
| chr1:2242961-2243286 | 2 | 0.047 | 0.01 | 2.97E-09 | | |
| chr13:73077993-73078011 | 2 | 0.2 | 0.03 | 3.08E-09 | | |
| chr4:110612727-110612795 | 2 | 0.25 | 0.03 | 3.10E-09 | | |
| chr17:78971613-78972133 | 4 | -0.48 | 0.06 | 3.14E-09 | *LGALS3BP* | exon_6 |
| chr11:120719408-120719425 | 2 | -0.55 | 0.07 | 3.25E-09 | | |
| chr17:40308478-40308965 | 2 | -0.68 | 0.09 | 3.37E-09 | *RARA* | TSS1500 |
| chr6:106509124-106509529 | 4 | 0.084 | 0.01 | 3.51E-09 | | |
| chr10:114881705-114882067 | 2 | 0.61 | 0.08 | 3.54E-09 | | |
| chr4:55794162-55794294 | 3 | 0.2 | 0.03 | 3.57E-09 | | |
| chr5:74103667-74103876 | 4 | -0.15 | 0.02 | 3.63E-09 | | |
| chr8:6563320-6563337 | 2 | -0.71 | 0.09 | 3.72E-09 | *ANGPT2* | TSS200 |
| chr4:142596889-142597183 | 3 | 0.066 | 0.01 | 3.98E-09 | | |
| chr12:132446891-132446915 | 2 | 0.69 | 0.09 | 4.38E-09 | | |
| chr22:30607160-30607296 | 4 | 0.35 | 0.05 | 4.42E-09 | *TCN2* | 5UTR |
| chr17:78680284-78680591 | 2 | -0.35 | 0.05 | 4.61E-09 | *CYTH1* | exon_12 |
| chr15:75626284-75626466 | 6 | 0.36 | 0.05 | 4.68E-09 | *SNUPN* | 5UTR |
| chr17:9791422-9791472 | 2 | -0.59 | 0.08 | 4.74E-09 | *DHRS7C* | 5UTR |
| chr12:11775550-11775748 | 2 | -0.48 | 0.06 | 4.78E-09 | | |
| chr11:64638292-64638521 | 3 | 0.13 | 0.02 | 4.78E-09 | | |
| chr11:57640070-57640369 | 2 | 1.7 | 0.21 | 4.93E-09 | *MIR130A* | TSS1500 |
| chr11:72754914-72754983 | 2 | -0.62 | 0.08 | 5.53E-09 | *STARD10* | exon_7 |
| chr11:64103771-64103793 | 2 | -0.84 | 0.11 | 5.78E-09 | *FLRT1* | 5UTR |
| chr20:2379739-2379750 | 2 | -0.85 | 0.11 | 5.90E-09 | *TGM6* | TSS1500 |
| chr6:33275598-33275614 | 2 | -0.74 | 0.09 | 6.27E-09 | | |

| | | | | | | |
|---|---|---|---|---|---|---|
| chr3:139202451-139202816 | 2 | -0.82 | 0.10 | 6.43E-09 | | |
| chr11:1782993-1783085 | 2 | -0.61 | 0.08 | 6.52E-09 | | |
| chr15:67185698-67185800 | 2 | -0.47 | 0.06 | 6.61E-09 | | |
| chr6:30685414-30685465 | 2 | -1 | 0.13 | 6.81E-09 | *PPP1R18* | exon_1 |
| chr1:962569-963364 | 4 | 0.07 | 0.01 | 6.81E-09 | *KLHL17* | exon_8 |
| chr16:67536959-67537268 | 2 | -0.77 | 0.10 | 6.93E-09 | *RIPOR1* | TSS1500 |
| chr22:19722232-19722568 | 2 | -0.83 | 0.11 | 6.96E-09 | *SEPTIN5* | exon_11 |
| chr2:216081276-216081478 | 2 | -0.21 | 0.03 | 7.04E-09 | *TMEM169* | TSS1500 |
| chr7:76428557-76428973 | 2 | -0.45 | 0.06 | 7.18E-09 | | |
| chr11:16976479-16976748 | 2 | -0.52 | 0.07 | 7.52E-09 | | |
| chr6:44674314-44674790 | 2 | -0.63 | 0.08 | 7.75E-09 | | |
| chr8:41310283-41310329 | 2 | -0.55 | 0.07 | 7.79E-09 | *SFRP1* | TSS1500 |
| chr22:30300175-30300616 | 2 | 0.61 | 0.08 | 7.95E-09 | | |
| chr7:2661933-2662168 | 3 | 0.092 | 0.01 | 8.44E-09 | *TTYH3* | 3UTR |
| chr5:149174199-149174561 | 2 | -0.38 | 0.05 | 8.70E-09 | | |
| chr1:32765508-32765589 | 4 | 0.019 | 0.00 | 9.05E-09 | *KIAA1522* | TSS200 |
| chr11:129947754-129947817 | 2 | -0.71 | 0.09 | 9.55E-09 | *PRDM10* | TSS1500 |
| chr8:49329325-49329770 | 3 | -0.39 | 0.05 | 9.97E-09 | | |
| chr15:63382250-63382724 | 2 | -0.91 | 0.12 | 1.04E-08 | *CA12* | TSS1500 |
| chr3:32036204-32036296 | 2 | -0.24 | 0.03 | 1.05E-08 | | |
| chr1:109624887-109625354 | 3 | -0.75 | 0.10 | 1.07E-08 | *AMPD2* | exon_2 |
| chr19:5913912-5913998 | 3 | 0.86 | 0.11 | 1.11E-08 | *CAPS* | TSS1500 |
| chr14:24313250-24313328 | 2 | 0.25 | 0.03 | 1.15E-08 | *LTB4R* | 5UTR |
| chr3:184319693-184320191 | 4 | 0.089 | 0.01 | 1.16E-08 | *EIF4G1* | exon_7 |
| chr10:71757634-71757640 | 2 | 0.44 | 0.06 | 1.18E-08 | | |
| chr15:92634719-92634984 | 2 | -1.1 | 0.14 | 1.22E-08 | | |
| chr20:62359099-62359544 | 2 | -0.24 | 0.03 | 1.27E-08 | | |
| chr8:41665339-41665417 | 3 | 0.19 | 0.03 | 1.31E-08 | *ANK1* | TSS200 |
| chr7:24758265-24758486 | 2 | -1 | 0.14 | 1.41E-08 | *GSDME* | TSS1500 |
| chr5:174726315-174726793 | 2 | -0.66 | 0.09 | 1.46E-08 | | |
| chr8:125289992-125290154 | 2 | -0.66 | 0.09 | 1.46E-08 | | |
| chr5:135547012-135547230 | 2 | -0.69 | 0.09 | 1.47E-08 | | |
| chr17:29576320-29576433 | 2 | -1.1 | 0.15 | 1.49E-08 | *GIT1* | exon_14 |
| chr7:97025520-97026254 | 6 | 0.0088 | 0.00 | 1.50E-08 | *DLX5* | TSS1500 |
| chr10:113053805-113053999 | 2 | -0.54 | 0.07 | 1.80E-08 | | |
| chr9:109277536-109277610 | 3 | -0.3 | 0.04 | 1.80E-08 | | |
| chr9:113398689-113398967 | 2 | -0.3 | 0.04 | 1.89E-08 | | |
| chr3:37694521-37694554 | 2 | -1.1 | 0.15 | 1.91E-08 | | |
| chr12:1950110-1950119 | 2 | -0.33 | 0.04 | 1.96E-08 | | |
| chr17:57623204-57623323 | 2 | -0.65 | 0.09 | 2.09E-08 | | |
| chr5:172886633-172887130 | 2 | -0.67 | 0.09 | 2.17E-08 | | |
| chr19:3383485-3383596 | 2 | -0.59 | 0.08 | 2.17E-08 | | |
| chr20:50267024-50267409 | 4 | -0.41 | 0.05 | 2.51E-08 | *PELATON* | TSS200 |
| chr2:120278864-120279129 | 2 | 0.32 | 0.04 | 2.52E-08 | | |
| chr12:542244-542288 | 2 | -0.34 | 0.05 | 2.70E-08 | | |
| chr16:30663169-30663212 | 2 | -1.1 | 0.14 | 2.85E-08 | | |
| chr11:62806408-62806866 | 2 | -0.41 | 0.05 | 3.10E-08 | *NXF1* | TSS1500 |
| chr22:29310714-29310764 | 2 | 0.096 | 0.01 | 3.21E-08 | *GAS2L1* | exon_4 |

| | | | | | | |
|---|---|---|---|---|---|---|
| chr1:152810661-152811013 | 2 | -0.21 | 0.03 | 3.23E-08 | *LCE1B* | TSS1500 |
| chr5:14900903-14901063 | 2 | -0.65 | 0.09 | 3.24E-08 | | |
| chr2:60515886-60516354 | 2 | -0.58 | 0.08 | 3.48E-08 | | |
| chr19:45352579-45353021 | 3 | -0.87 | 0.12 | 3.65E-08 | *ERCC2* | exon_20 |
| chr16:85781899-85782094 | 3 | -0.13 | 0.02 | 3.67E-08 | | |
| chr1:27351369-27351381 | 2 | -0.15 | 0.02 | 3.68E-08 | | |
| chr6:34146110-34146155 | 4 | 0.16 | 0.02 | 3.95E-08 | *GRM4* | TSS200 |
| chr19:345778-346278 | 2 | -0.22 | 0.03 | 4.05E-08 | *MIER2* | TSS1500 |
| chr2:100821410-100821742 | 2 | -0.5 | 0.07 | 4.15E-08 | | |
| chr1:41918766-41918820 | 3 | -0.63 | 0.08 | 4.24E-08 | *HIVEP3* | exon_1 |
| chr2:152924635-152925046 | 2 | -0.33 | 0.04 | 4.28E-08 | | |
| chr2:191846487-191846755 | 3 | -0.34 | 0.04 | 4.36E-08 | *CAVIN2* | exon_1 |
| chr6:32842929-32842965 | 2 | -0.69 | 0.09 | 4.37E-08 | *PSMB8-AS1* | TSS1500 |
| chr7:478071-478167 | 2 | -0.87 | 0.12 | 4.63E-08 | | |
| chr1:1122697-1123059 | 2 | -0.93 | 0.12 | 4.83E-08 | | |
| chr9:126661807-126662196 | 3 | 0.24 | 0.03 | 4.85E-08 | | |
| chr16:90010358-90010800 | 2 | -0.59 | 0.08 | 4.95E-08 | *DBNDD1* | TSS1500 |
| chr6:144351751-144351863 | 2 | -0.38 | 0.05 | 5.12E-08 | | |
| chr5:150441728-150442187 | 2 | -0.59 | 0.08 | 5.28E-08 | | |
| chr15:31065854-31066120 | 2 | -0.22 | 0.03 | 5.38E-08 | *TRPM1* | exon_6 |
| chr10:100286986-100286995 | 2 | -0.65 | 0.09 | 5.45E-08 | *BLOC1S2* | TSS1500 |
| chr17:9257010-9257023 | 2 | -0.64 | 0.09 | 6.09E-08 | | |
| chr17:9054347-9054773 | 4 | -0.87 | 0.12 | 6.10E-08 | | |
| chr8:141210230-141210463 | 2 | -0.68 | 0.09 | 6.14E-08 | *SLC45A4* | 3UTR |
| chr3:184426659-184427034 | 2 | 0.35 | 0.05 | 6.22E-08 | *LINC02054* | exon_2 |
| chr13:53028366-53029022 | 3 | -0.39 | 0.05 | 6.24E-08 | *OLFM4* | TSS1500 |
| chr9:136572180-136572321 | 2 | -0.57 | 0.08 | 6.31E-08 | | |
| chr6:33163445-33163497 | 2 | 0.38 | 0.05 | 6.38E-08 | *COL11A2* | 3UTR |
| chr3:127676936-127677412 | 3 | -0.21 | 0.03 | 6.76E-08 | *ABTB1* | exon_8 |
| chr15:74422764-74423158 | 2 | -0.56 | 0.08 | 6.79E-08 | | |
| chr9:95024492-95024609 | 3 | 0.63 | 0.09 | 6.82E-08 | | |
| chr1:53294398-53294665 | 2 | -0.56 | 0.08 | 7.31E-08 | | |
| chr1:204286824-204287021 | 2 | -0.59 | 0.08 | 7.67E-08 | | |
| chr10:102775764-102776035 | 2 | -0.4 | 0.05 | 8.35E-08 | *WBP1L* | TSS1500 |
| chr6:151239846-151239897 | 2 | -0.36 | 0.05 | 8.46E-08 | *AKAP12* | TSS200 |
| chr9:121566988-121567235 | 4 | -0.46 | 0.06 | 9.43E-08 | *DAB2IP* | 5UTR |
| chr14:22492844-22494301 | 6 | -0.097 | 0.01 | 1.01E-07 | | |
| chr4:188031938-188032024 | 3 | 0.23 | 0.03 | 1.10E-07 | | |
| chr2:239337307-239337518 | 3 | 0.33 | 0.04 | 1.11E-07 | | |
| chr2:28644207-28644321 | 2 | 0.92 | 0.12 | 1.14E-07 | | |
| chr7:17274690-17275062 | 2 | -0.26 | 0.04 | 1.14E-07 | | |
| chr16:2033177-2033392 | 2 | -0.42 | 0.06 | 1.18E-07 | *SLC9A3R2* | TSS200 |
| chr3:188901120-188901336 | 4 | -0.16 | 0.02 | 1.18E-07 | | |
| chr19:16442940-16443076 | 2 | -0.81 | 0.11 | 1.20E-07 | | |
| chr1:207064877-207064991 | 2 | -1.3 | 0.17 | 1.24E-07 | | |
| chr1:161201421-161202020 | 7 | -0.099 | 0.01 | 1.25E-07 | *NDUFS2* | TSS1500 |
| chr11:846055-846237 | 2 | -0.66 | 0.09 | 1.32E-07 | | |
| chr11:65917637-65917791 | 2 | -0.52 | 0.07 | 1.35E-07 | *C11orf68* | exon_2 |

| | | | | | | |
|---|---|---|---|---|---|---|
| chr4:185956396-185956624 | 4 | 0.042 | 0.01 | 1.36E-07 | *SORBS2* | TSS200 |
| chr4:168630945-168631195 | 3 | -0.22 | 0.03 | 1.38E-07 | *PALLD* | TSS1500 |
| chr2:127025044-127025075 | 2 | -0.057 | 0.01 | 1.40E-07 | | |
| chr5:159210257-159210676 | 4 | -0.17 | 0.02 | 1.40E-07 | *RNF145* | TSS1500 |
| chr6:15244387-15244462 | 2 | -0.6 | 0.08 | 1.41E-07 | | |
| chr7:55156816-55157389 | 6 | -0.15 | 0.02 | 1.48E-07 | *EGFR* | exon_10 |
| chr7:2438853-2439078 | 2 | -0.35 | 0.05 | 1.54E-07 | *CHST12* | 3UTR |
| chr7:23473420-23473697 | 2 | -0.26 | 0.04 | 1.66E-07 | | |
| chr8:32646591-32646595 | 2 | -1.2 | 0.16 | 1.74E-07 | *NRG1* | TSS1500 |
| chr19:51354975-51355022 | 2 | -0.51 | 0.07 | 1.79E-07 | *ETFB* | TSS200 |
| chr11:2825451-2825702 | 3 | 0.34 | 0.05 | 1.79E-07 | | |
| chr15:65115186-65115549 | 2 | 0.33 | 0.05 | 1.81E-07 | *UBAP1L* | 5UTR |
| chr19:19528759-19528772 | 2 | -0.38 | 0.05 | 1.84E-07 | *YJEFN3* | TSS200 |
| chr2:230870322-230870719 | 2 | -0.64 | 0.09 | 1.84E-07 | | |
| chr2:127455854-127456268 | 2 | -0.54 | 0.07 | 1.91E-07 | | |
| chr15:101188246-101188448 | 4 | -0.11 | 0.02 | 1.92E-07 | | |
| chr14:23366661-23366803 | 2 | -0.17 | 0.02 | 1.98E-07 | *EFS* | TSS1500 |
| chr11:64000940-64001193 | 2 | -1.4 | 0.19 | 2.03E-07 | | |
| chr16:56370921-56370959 | 2 | 0.12 | 0.02 | 2.25E-07 | | |
| chr6:32843476-32843718 | 2 | -1.1 | 0.16 | 2.27E-07 | *PSMB8-AS1* | TSS1500 |
| chr22:39254969-39255359 | 2 | -0.74 | 0.10 | 2.32E-07 | | |
| chr21:36977254-36977980 | 3 | -0.51 | 0.07 | 2.36E-07 | | |
| chr4:40335320-40335426 | 2 | -0.95 | 0.13 | 2.37E-07 | *CHR9* | 5UTR |
| chr1:6477601-6477780 | 3 | -0.59 | 0.08 | 2.41E-07 | *PLEKHG5* | 5UTR |
| chr1:201737893-201737944 | 2 | -0.79 | 0.11 | 2.43E-07 | | |
| chr2:23525234-23525747 | 3 | -0.25 | 0.03 | 2.43E-07 | | |
| chr15:22917599-22917686 | 3 | -0.64 | 0.09 | 2.61E-07 | *CYFIP1* | TSS1500 |
| chr10:72328267-72328431 | 2 | -0.35 | 0.05 | 2.61E-07 | | |
| chr8:22056495-22056776 | 2 | -0.17 | 0.02 | 2.62E-07 | *DMTN* | TSS1500 |
| chr7:27114017-27114044 | 3 | -0.079 | 0.01 | 2.62E-07 | *HOXA3* | TSS200 |
| chr17:1710974-1711206 | 2 | -0.4 | 0.06 | 2.66E-07 | *TLCD2* | TSS1500 |
| chr17:19387449-19387807 | 2 | -0.7 | 0.10 | 2.69E-07 | *MFAP4* | TSS1500 |
| chr3:117885449-117885564 | 2 | -0.61 | 0.08 | 2.71E-07 | | |
| chr4:7393175-7393300 | 2 | -2 | 0.27 | 2.75E-07 | | |
| chr9:86807578-86807688 | 2 | -0.5 | 0.07 | 2.83E-07 | | |
| chr12:57205575-57205934 | 2 | -1.3 | 0.17 | 2.83E-07 | *LRP1* | exon_75 |
| chr10:72089059-72089065 | 2 | -0.27 | 0.04 | 2.83E-07 | *SPOCK2* | TSS200 |
| chr21:44773515-44773926 | 3 | 0.27 | 0.04 | 2.83E-07 | *UBE2G2* | exon_3 |
| chr18:9069787-9069807 | 2 | -0.39 | 0.05 | 2.84E-07 | | |
| chr16:57318453-57318820 | 2 | -0.56 | 0.08 | 2.93E-07 | | |
| chr3:126927654-126927964 | 2 | -0.48 | 0.07 | 3.09E-07 | | |
| chr17:57724236-57724655 | 2 | -0.67 | 0.09 | 3.09E-07 | | |
| chr10:29595440-29595939 | 2 | -0.26 | 0.04 | 3.28E-07 | | |
| chr12:116978024-116978139 | 2 | 2.1 | 0.28 | 3.30E-07 | *.OC10050655[illegible]* | exon_4 |
| chr6:30913802-30913868 | 3 | -0.022 | 0.00 | 3.54E-07 | *VARS2* | TSS1500 |
| chr11:2142069-2142283 | 2 | -0.36 | 0.05 | 3.56E-07 | *IGF2* | TSS1500 |
| chr7:5495832-5495951 | 2 | -0.35 | 0.05 | 3.58E-07 | *MIR589* | TSS200 |
| chr5:181198118-181198506 | 3 | 2.1 | 0.28 | 3.60E-07 | | |

| | | | | | | |
|---|---|---|---|---|---|---|
| chr17:82302075-82302534 | 2 | -0.65 | 0.09 | 3.63E-07 | | |
| chr12:124766190-124766345 | 2 | -0.28 | 0.04 | 3.75E-07 | | |
| chr13:113007273-113007554 | 2 | -1.3 | 0.18 | 4.03E-07 | | |
| chr22:27757333-27757753 | 3 | -0.58 | 0.08 | 4.05E-07 | | |
| chr3:11136300-11136679 | 3 | -0.49 | 0.07 | 4.05E-07 | *HRH1* | TSS1500 |
| chr15:77044232-77044596 | 2 | -0.38 | 0.05 | 4.12E-07 | *TSPAN3* | 3UTR |
| chr17:49578002-49578107 | 2 | -0.7 | 0.10 | 4.28E-07 | | |
| chr5:139794838-139794944 | 2 | -0.54 | 0.07 | 4.29E-07 | *PSD2* | TSS1500 |
| chr2:207766357-207766535 | 2 | -1.6 | 0.23 | 4.53E-07 | *FZD5* | 3UTR |
| chr1:31798759-31798901 | 3 | -0.11 | 0.02 | 4.63E-07 | *SPOCD1* | TSS200 |
| chr18:48827929-48827935 | 2 | -0.53 | 0.07 | 4.66E-07 | | |
| chr2:231393301-231393706 | 3 | -0.78 | 0.11 | 4.69E-07 | | |
| chr9:136858953-136859063 | 2 | 0.31 | 0.04 | 4.83E-07 | *MAMDC4* | exon_24 |
| chr11:1000888-1001163 | 2 | -0.37 | 0.05 | 4.96E-07 | | |
| chr17:80920564-80920593 | 2 | -0.32 | 0.05 | 5.00E-07 | | |
| chr1:1431645-1431877 | 2 | -0.47 | 0.07 | 5.06E-07 | | |
| chr2:16522997-16523180 | 2 | -1.1 | 0.15 | 5.07E-07 | | |
| chr2:120279184-120279215 | 2 | -0.6 | 0.08 | 5.13E-07 | | |
| chr15:81225200-81225215 | 2 | -0.61 | 0.08 | 5.37E-07 | | |
| chr8:9057477-9057638 | 2 | -0.51 | 0.07 | 5.40E-07 | | |
| chr14:67763524-67763752 | 2 | -0.41 | 0.06 | 5.62E-07 | | |
| chr15:73801012-73801090 | 2 | -0.47 | 0.07 | 5.73E-07 | | |
| chr16:74447709-74447764 | 2 | -0.54 | 0.08 | 5.75E-07 | *GLG1* | 3UTR |
| chr7:73827698-73828244 | 3 | -0.63 | 0.09 | 5.81E-07 | | |
| chr1:185557468-185557911 | 2 | -0.22 | 0.03 | 5.89E-07 | | |
| chr11:1008955-1009404 | 3 | -0.71 | 0.10 | 5.89E-07 | *AP2A2* | exon_18 |
| chr17:61486954-61487296 | 2 | -0.91 | 0.13 | 5.92E-07 | | |
| chr12:12736655-12736901 | 2 | -0.96 | 0.13 | 6.05E-07 | | |
| chr11:119653202-119653444 | 2 | -0.9 | 0.12 | 6.09E-07 | | |
| chr17:4786345-4786470 | 5 | 0.036 | 0.00 | 6.12E-07 | *VMO1* | TSS200 |
| chr15:74396664-74397112 | 2 | -0.29 | 0.04 | 6.21E-07 | | |
| chr7:29127038-29127455 | 2 | -0.49 | 0.07 | 6.45E-07 | | |
| chr11:69658522-69658547 | 2 | -1.1 | 0.15 | 6.61E-07 | | |
| chr6:32084512-32084516 | 2 | 0.36 | 0.05 | 6.70E-07 | *TNXB* | exon_8 |
| chr16:86571-86657 | 2 | -0.39 | 0.06 | 6.72E-07 | *NPRL3* | 3UTR |
| chr11:72741437-72741722 | 2 | -0.35 | 0.05 | 6.83E-07 | | |
| chr8:41306534-41306996 | 2 | -1.2 | 0.17 | 6.85E-07 | | |
| chr6:156397128-156397412 | 4 | -0.34 | 0.05 | 7.04E-07 | | |
| chr4:1806859-1807068 | 2 | -1.2 | 0.17 | 7.34E-07 | *FGFR3* | exon_17 |
| chr10:3750047-3750091 | 2 | -0.37 | 0.05 | 7.37E-07 | *.OC10537636(* | exon_2 |
| chr5:95861712-95861772 | 2 | -0.16 | 0.02 | 7.38E-07 | | |
| chr3:129122591-129122604 | 2 | -0.76 | 0.11 | 7.39E-07 | *RAB43* | TSS1500 |
| chr11:71999568-71999597 | 2 | -0.62 | 0.09 | 7.53E-07 | *IL18BP* | 5UTR |
| chr8:130317714-130318003 | 2 | -0.76 | 0.11 | 7.82E-07 | | |
| chr2:4140810-4141127 | 2 | -0.36 | 0.05 | 7.85E-07 | | |
| chr1:19645806-19646134 | 2 | -0.45 | 0.06 | 8.17E-07 | *NBL1* | 5UTR |
| chr1:10813906-10814258 | 2 | -0.44 | 0.06 | 8.20E-07 | | |
| chr11:45209496-45209624 | 2 | -0.85 | 0.12 | 8.39E-07 | | |

| | | | | | | |
|---|---|---|---|---|---|---|
| chr7:75962816-75963100 | 2 | -0.61 | 0.09 | 8.42E-07 | | |
| chr1:6520251-6520327 | 2 | 0.072 | 0.01 | 9.04E-07 | *PLEKHG5* | TSS1500 |
| chr17:45405885-45406289 | 6 | -0.31 | 0.04 | 9.18E-07 | *ARHGAP27* | exon_2 |
| chr16:4306821-4307317 | 2 | -0.42 | 0.06 | 9.46E-07 | | |
| chr19:49862502-49862983 | 3 | -0.26 | 0.04 | 9.51E-07 | | |
| chr11:64760374-64761097 | 3 | -0.16 | 0.02 | 9.55E-07 | *PYGM* | TSS1500 |
| chr3:50246873-50246970 | 2 | -1.1 | 0.16 | 9.80E-07 | *GI2* | 5UTR |
| chr14:68364096-68364493 | 3 | -0.39 | 0.05 | 1.00E-06 | | |
| chr5:138139488-138139599 | 2 | -0.49 | 0.07 | 1.03E-06 | *NME5* | TSS200 |
| chr17:41812996-41813015 | 4 | -1.5 | 0.21 | 1.03E-06 | *FKBP10* | TSS1500 |
| chr1:162984492-162985340 | 3 | -0.2 | 0.03 | 1.06E-06 | | |
| chr16:55701975-55702206 | 2 | -1.1 | 0.16 | 1.07E-06 | | |
| chr9:88537610-88537970 | 2 | -0.7 | 0.10 | 1.09E-06 | | |
| chr5:95823423-95823910 | 4 | 0.096 | 0.01 | 1.13E-06 | *GLRX* | TSS1500 |
| chr12:130017504-130017738 | 4 | 0.0041 | 0.00 | 1.16E-06 | | |
| chr14:105478950-105479362 | 3 | -0.44 | 0.06 | 1.18E-06 | | |
| chr5:174617887-174617960 | 2 | -0.56 | 0.08 | 1.22E-06 | | |
| chr8:37843284-37843497 | 2 | -1.4 | 0.20 | 1.27E-06 | *ADGRA2* | 3UTR |
| chr3:157086117-157086457 | 2 | -0.23 | 0.03 | 1.34E-06 | | |
| chr1:6461078-6461208 | 2 | -0.91 | 0.13 | 1.34E-06 | *TNFRSF25* | 3UTR |
| chr20:36887695-36888168 | 2 | -0.78 | 0.11 | 1.38E-06 | | |
| chr9:133029579-133029703 | 3 | 0.29 | 0.04 | 1.45E-06 | *GTF3C5* | TSS1500 |
| chr17:79924532-79925179 | 3 | -0.24 | 0.03 | 1.48E-06 | | |
| chr1:67647329-67647666 | 2 | -0.3 | 0.04 | 1.51E-06 | | |
| chr1:84398168-84398384 | 2 | -0.62 | 0.09 | 1.54E-06 | *DSE2B* | TSS1500 |
| chr2:238025152-238025204 | 2 | -0.46 | 0.07 | 1.55E-06 | | |
| chr19:44757678-44757747 | 2 | 0.58 | 0.08 | 1.57E-06 | *MIR8085* | TSS1500 |
| chr5:14676351-14676509 | 2 | 0.52 | 0.07 | 1.59E-06 | | |
| chr10:132558343-132558879 | 3 | 0.76 | 0.11 | 1.60E-06 | | |
| chr11:46236488-46237275 | 3 | -0.36 | 0.05 | 1.63E-06 | | |
| chr20:19253274-19253397 | 2 | -0.52 | 0.07 | 1.69E-06 | | |
| chr1:10835460-10835921 | 3 | -0.51 | 0.07 | 1.70E-06 | | |
| chr1:206865239-206865663 | 3 | 0.061 | 0.01 | 1.71E-06 | *IL20* | 5UTR |
| chr7:30996555-30996999 | 2 | -0.28 | 0.04 | 1.72E-06 | | |
| chr1:44238078-44238251 | 2 | -0.23 | 0.03 | 1.75E-06 | | |
| chr8:38402049-38402386 | 2 | -0.47 | 0.07 | 1.77E-06 | | |
| chr4:85930089-85930117 | 2 | -0.51 | 0.07 | 1.80E-06 | *ARHGAP24* | 5UTR |
| chr8:26635599-26635959 | 2 | -0.82 | 0.12 | 1.82E-06 | | |
| chr1:161084348-161084384 | 2 | -0.1 | 0.02 | 1.86E-06 | | |
| chr11:72308857-72308987 | 2 | 0.029 | 0.00 | 1.87E-06 | | |
| chr2:174634555-174634637 | 2 | -0.33 | 0.05 | 1.91E-06 | *WIPF1* | 5UTR |
| chr14:104666006-104666142 | 2 | -0.77 | 0.11 | 1.93E-06 | *LINC02280* | TSS1500 |
| chr19:2016230-2016403 | 3 | 0.3 | 0.04 | 1.94E-06 | *BTBD2* | TSS1500 |
| chr1:59198510-59198758 | 2 | -0.61 | 0.09 | 1.96E-06 | | |
| chr1:161119167-161119312 | 2 | -0.07 | 0.01 | 2.00E-06 | *NIT1* | exon_3 |
| chr19:3539099-3539129 | 2 | -0.81 | 0.11 | 2.05E-06 | *C19orf71* | TSS200 |
| chr15:78066144-78066607 | 2 | -0.66 | 0.09 | 2.31E-06 | *TBC1D2B* | 5UTR |
| chr4:150583035-150583096 | 2 | 0.9 | 0.13 | 2.32E-06 | *MAB21L2* | exon_1 |

| | | | | | | |
|---|---|---|---|---|---|---|
| chr17:63440993-63441601 | 6 | -0.29 | 0.04 | 2.36E-06 | *CYB561* | TSS1500 |
| chr1:15957723-15957962 | 2 | -1.2 | 0.18 | 2.42E-06 | | |
| chr19:40366235-40366379 | 2 | -0.73 | 0.10 | 2.42E-06 | | |
| chr1:225465330-225465498 | 2 | -0.18 | 0.03 | 2.45E-06 | | |
| chr17:50270725-50271204 | 2 | -1 | 0.14 | 2.46E-06 | *TMEM92* | TSS1500 |
| chr3:51942229-51942651 | 4 | -0.98 | 0.14 | 2.48E-06 | *PARP3* | 5UTR |
| chr12:53005309-53005566 | 4 | 0.074 | 0.01 | 2.53E-06 | *EIF4B* | TSS1500 |
| chr17:4719792-4720080 | 2 | -0.65 | 0.09 | 2.56E-06 | | |
| chr8:141331821-141332200 | 3 | -1 | 0.14 | 2.57E-06 | | |
| chr19:1818666-1818887 | 3 | -0.44 | 0.06 | 2.77E-06 | *REXO1* | exon_9 |
| chr12:51390860-51391125 | 2 | 0.5 | 0.07 | 2.80E-06 | *SLC4A8* | TSS1500 |
| chr9:91858129-91858481 | 3 | -0.3 | 0.04 | 2.86E-06 | | |
| chr16:3589687-3589838 | 2 | -0.71 | 0.10 | 2.86E-06 | *SLX4* | exon_12 |
| chr3:184691542-184691958 | 2 | 0.64 | 0.09 | 2.87E-06 | | |
| chr11:67271648-67271983 | 2 | -0.31 | 0.05 | 2.88E-06 | | |
| chr13:112346979-112347034 | 3 | 0.17 | 0.02 | 3.01E-06 | | |
| chr15:52161791-52161951 | 2 | -0.42 | 0.06 | 3.02E-06 | | |
| chr10:132654454-132654932 | 2 | -0.98 | 0.14 | 3.03E-06 | | |
| chr14:21022654-21022886 | 2 | 1 | 0.15 | 3.10E-06 | *NDRG2* | exon_3 |
| chr4:122826378-122826387 | 2 | -0.9 | 0.13 | 3.11E-06 | *FGF2* | TSS1500 |
| chr10:122099184-122099303 | 3 | -0.53 | 0.08 | 3.19E-06 | | |
| chr2:224402016-224402066 | 2 | 0.28 | 0.04 | 3.21E-06 | *FAM124B* | 5UTR |
| chr12:116548079-116548183 | 2 | -0.81 | 0.12 | 3.23E-06 | | |
| chr8:133102590-133102811 | 3 | 0.015 | 0.00 | 3.46E-06 | *SLA* | 5UTR |
| chr8:22057673-22057999 | 2 | -0.46 | 0.07 | 3.57E-06 | *DMTN* | TSS1500 |
| chr4:150209431-150209508 | 3 | -1 | 0.15 | 4.05E-06 | | |
| chr15:43221108-43221306 | 2 | -0.6 | 0.09 | 4.33E-06 | *EPB42* | TSS1500 |
| chr8:48370352-48370615 | 2 | -0.64 | 0.09 | 4.34E-06 | | |
| chr16:3963536-3963658 | 2 | -0.33 | 0.05 | 4.37E-06 | *ADCY9* | 3UTR |
| chr22:38283919-38284088 | 2 | 1.6 | 0.23 | 4.37E-06 | | |
| chr17:38886870-38887510 | 3 | -0.42 | 0.06 | 4.56E-06 | | |
| chr12:24997468-24997596 | 2 | -0.17 | 0.03 | 4.75E-06 | *IRAG2* | TSS200 |
| chr6:170295150-170295155 | 2 | -0.65 | 0.09 | 4.76E-06 | *FAM120B* | TSS200 |
| chr17:44708010-44708303 | 2 | -0.43 | 0.06 | 4.85E-06 | *DBF4B* | TSS1500 |
| chr9:127950104-127950159 | 2 | -0.82 | 0.12 | 5.07E-06 | | |
| chr11:8840432-8840505 | 3 | -0.16 | 0.02 | 5.09E-06 | | |
| chr7:101237832-101238005 | 2 | -0.58 | 0.08 | 5.10E-06 | *CLDN15* | 5UTR |
| chr12:6913902-6914098 | 3 | 0.16 | 0.02 | 5.28E-06 | *LRRC23* | 3UTR |
| chr9:89990492-89990919 | 2 | -0.82 | 0.12 | 5.37E-06 | | |
| chr1:207022091-207022299 | 2 | -0.59 | 0.09 | 5.42E-06 | *C1orf116* | exon_3 |
| chr11:47245016-47245337 | 2 | 0.72 | 0.10 | 5.49E-06 | *ACP2* | exon_6 |
| chr1:112723057-112723323 | 4 | -0.87 | 0.13 | 5.60E-06 | *TAFA3* | exon_4 |
| chr5:139384714-139384801 | 2 | -0.47 | 0.07 | 5.62E-06 | *SLC23A1* | TSS1500 |
| chr12:1616862-1616893 | 3 | -0.55 | 0.08 | 5.68E-06 | *WNT5B* | TSS200 |
| chr1:31769612-31769909 | 2 | -0.59 | 0.09 | 5.73E-06 | | |
| chr9:92717503-92717642 | 2 | -0.48 | 0.07 | 5.85E-06 | | |
| chr1:201737235-201737320 | 2 | -0.55 | 0.08 | 5.87E-06 | | |
| chr17:2237461-2237648 | 4 | -0.51 | 0.07 | 5.91E-06 | *SMG6* | TSS1500 |

| | | | | | | |
|---|---|---|---|---|---|---|
| chr13:23695155-23695361 | 2 | -1.6 | 0.23 | 5.95E-06 | | |
| chr3:58164911-58165303 | 2 | -0.57 | 0.08 | 5.97E-06 | *FLNB-AS1* | exon_2 |
| chr13:105505875-105506203 | 2 | -0.32 | 0.05 | 5.98E-06 | *DAOA-AS1* | TSS1500 |
| chr22:41526983-41527117 | 2 | -0.86 | 0.12 | 6.11E-06 | *POLR3H* | 3UTR |
| chr7:27103668-27103715 | 2 | -0.42 | 0.06 | 6.11E-06 | *HOXA2* | TSS1500 |
| chr17:81405361-81405650 | 2 | -0.74 | 0.11 | 6.16E-06 | | |
| chr16:81888761-81888938 | 2 | -0.51 | 0.08 | 6.17E-06 | | |
| chr9:106616974-106616980 | 2 | -0.38 | 0.06 | 6.30E-06 | | |
| chr4:26197353-26197647 | 2 | -0.71 | 0.10 | 6.33E-06 | | |
| chr19:2986837-2987084 | 2 | -0.74 | 0.11 | 6.39E-06 | *TLE6* | exon_6 |
| chr3:52054123-52054237 | 2 | -0.38 | 0.06 | 6.41E-06 | *DUSP7* | exon_2 |
| chr11:79269794-79270014 | 2 | -0.89 | 0.13 | 6.46E-06 | | |
| chr1:153688812-153688934 | 2 | -0.85 | 0.12 | 6.62E-06 | | |
| chr6:169289844-169290176 | 2 | -0.17 | 0.03 | 6.65E-06 | | |
| chr14:69547831-69547945 | 2 | -1.1 | 0.16 | 6.71E-06 | | |
| chr5:128782142-128782611 | 2 | 0.42 | 0.06 | 6.75E-06 | | |
| chr11:65430062-65430312 | 4 | -0.89 | 0.13 | 6.80E-06 | *NEAT1* | exon_1 |
| chr22:24950224-24950787 | 3 | -0.64 | 0.09 | 7.00E-06 | | |
| chr14:36656563-36656667 | 2 | -0.29 | 0.04 | 7.10E-06 | *PAX9* | TSS1500 |
| chr3:136818765-136818910 | 4 | -0.17 | 0.03 | 7.41E-06 | *SLC35G2* | TSS1500 |
| chr4:8473134-8473460 | 3 | -0.31 | 0.05 | 7.42E-06 | | |
| chr11:61892244-61892718 | 3 | 0.17 | 0.03 | 7.61E-06 | *FADS3* | TSS1500 |
| chr6:34514683-34514702 | 3 | 0.013 | 0.00 | 7.70E-06 | *PACSIN1* | TSS200 |
| chr17:31733557-31733585 | 2 | -1.2 | 0.18 | 8.06E-06 | | |
| chr10:101064055-101064246 | 2 | -0.61 | 0.09 | 8.17E-06 | *KAZALD1* | exon_3 |
| chr7:2362705-2362891 | 2 | -0.18 | 0.03 | 8.21E-06 | *EIF3B* | exon_3 |
| chr19:58357218-58357638 | 4 | -0.061 | 0.01 | 8.28E-06 | *ZNF497* | exon_2 |
| chr12:109727219-109727647 | 3 | -0.2 | 0.03 | 8.30E-06 | | |
| chr16:89321988-89322431 | 2 | -0.57 | 0.08 | 8.56E-06 | | |
| chr16:68740806-68741315 | 3 | -0.28 | 0.04 | 8.88E-06 | | |
| chr15:45117849-45118038 | 2 | 0.54 | 0.08 | 8.96E-06 | *DUOXA1* | 3UTR |
| chr21:42247997-42248449 | 2 | -0.45 | 0.07 | 9.58E-06 | | |
| chr11:65792556-65792703 | 2 | 0.058 | 0.01 | 9.79E-06 | | |
| chr9:128998204-128998850 | 3 | -0.39 | 0.06 | 9.85E-06 | *NUP188* | exon_31 |
| chr12:49963099-49963272 | 2 | -0.41 | 0.06 | 9.98E-06 | *AQP5-AS1* | TSS1500 |
| chr7:99372738-99373120 | 2 | -0.34 | 0.05 | 1.03E-05 | | |
| chr7:111785350-111785372 | 2 | 0.44 | 0.07 | 1.04E-05 | | |
| chr1:118989729-118989919 | 2 | 0.36 | 0.05 | 1.05E-05 | *TBX15* | TSS1500 |
| chr4:108166474-108166824 | 3 | 1 | 0.15 | 1.06E-05 | *LEF1* | TSS1500 |
| chr17:41913934-41914044 | 2 | 0.9 | 0.13 | 1.08E-05 | | |
| chr17:34081057-34081429 | 2 | -0.51 | 0.08 | 1.10E-05 | | |
| chr3:10289993-10290292 | 2 | -0.92 | 0.14 | 1.13E-05 | | |
| chr2:47193315-47193728 | 2 | -0.44 | 0.07 | 1.18E-05 | | |
| chr22:35637979-35638165 | 2 | -0.33 | 0.05 | 1.20E-05 | | |
| chr15:45175419-45175430 | 2 | -0.38 | 0.06 | 1.22E-05 | *SHF* | exon_3 |
| chr11:67054194-67054733 | 3 | -0.35 | 0.05 | 1.23E-05 | | |
| chr11:125351246-125351556 | 4 | -0.31 | 0.05 | 1.24E-05 | | |
| chr1:25806316-25806374 | 2 | -0.56 | 0.08 | 1.26E-05 | | |

| | | | | | | |
|---|---|---|---|---|---|---|
| chr6:30769230-30769678 | 6 | -0.072 | 0.01 | 1.27E-05 | | |
| chr15:73741184-73741462 | 2 | 0.92 | 0.13 | 1.27E-05 | | |
| chr4:625519-625846 | 5 | -0.058 | 0.01 | 1.33E-05 | *PDE6B* | exon_1 |
| chr19:51085493-51085562 | 2 | 0.81 | 0.12 | 1.37E-05 | *KLK14* | TSS1500 |
| chr15:92637871-92637952 | 2 | -0.61 | 0.09 | 1.40E-05 | | |
| chr12:6364145-6364477 | 3 | 0.027 | 0.00 | 1.41E-05 | | |
| chr20:23086396-23086522 | 4 | 0.072 | 0.01 | 1.42E-05 | *CD93* | TSS200 |
| chr1:1542236-1542551 | 2 | -0.57 | 0.09 | 1.49E-05 | | |
| chr12:6326586-6326604 | 2 | -1.4 | 0.21 | 1.49E-05 | | |
| chr17:1650208-1650291 | 2 | -0.057 | 0.01 | 1.50E-05 | *RILP* | TSS1500 |
| chr17:56885167-56885451 | 3 | -0.32 | 0.05 | 1.57E-05 | *MTVR2* | TSS1500 |
| chr5:157269179-157269341 | 2 | -0.58 | 0.09 | 1.59E-05 | *CYFIP2* | 5UTR |
| chr22:23695614-23696336 | 4 | 0.18 | 0.03 | 1.59E-05 | *RGL4* | TSS1500 |
| chr12:119177110-119177548 | 2 | -0.23 | 0.03 | 1.61E-05 | *.OC10537002[illegible]* | TSS1500 |
| chr20:346290-346526 | 3 | -0.67 | 0.10 | 1.64E-05 | *NRSN2* | TSS1500 |
| chr1:205664557-205664631 | 3 | -0.2 | 0.03 | 1.75E-05 | *SLC45A3* | exon_2 |
| chr19:18428601-18428946 | 2 | -0.8 | 0.12 | 1.77E-05 | | |
| chr1:156076987-156077265 | 3 | -0.68 | 0.10 | 1.78E-05 | *MEX3A* | exon_2 |
| chr19:45136729-45137147 | 2 | -0.84 | 0.13 | 1.78E-05 | | |
| chr8:37479942-37480010 | 2 | -0.41 | 0.06 | 1.79E-05 | | |
| chr12:50073271-50073403 | 2 | -0.27 | 0.04 | 1.86E-05 | *ASIC1* | TSS1500 |
| chr15:31339515-31339638 | 2 | 0.074 | 0.01 | 1.88E-05 | | |
| chr11:268923-268950 | 2 | -0.077 | 0.01 | 1.92E-05 | | |
| chr3:71245190-71245229 | 2 | -0.37 | 0.05 | 1.99E-05 | *FOXP1* | TSS200 |
| chr6:32186938-32187364 | 3 | -0.95 | 0.14 | 2.01E-05 | *PBX2* | exon_6 |
| chr17:39165637-39166672 | 7 | -0.37 | 0.06 | 2.06E-05 | *ARL5C* | 5UTR |
| chr4:8228326-8228962 | 4 | -0.22 | 0.03 | 2.07E-05 | *SH3TC1* | exon_12 |
| chr17:14209344-14209456 | 2 | -0.72 | 0.11 | 2.11E-05 | | |
| chr11:46247418-46247764 | 2 | -1.2 | 0.18 | 2.11E-05 | | |
| chr17:39193532-39193999 | 4 | -0.47 | 0.07 | 2.16E-05 | | |
| chr15:63806394-63806667 | 3 | -0.48 | 0.07 | 2.17E-05 | | |
| chr1:10704455-10704829 | 2 | -0.77 | 0.11 | 2.20E-05 | | |
| chr7:5230936-5231297 | 2 | -0.37 | 0.06 | 2.29E-05 | *WIPI2* | 3UTR |
| chr11:8932994-8933023 | 2 | -0.7 | 0.10 | 2.29E-05 | *C11orf16* | TSS200 |
| chr9:135979609-135979749 | 2 | -0.6 | 0.09 | 2.35E-05 | | |
| chr6:111615326-111615474 | 2 | -0.88 | 0.13 | 2.35E-05 | | |
| chr13:20189915-20190187 | 3 | -0.57 | 0.09 | 2.48E-05 | | |
| chr2:38147272-38147331 | 2 | -0.48 | 0.07 | 2.51E-05 | | |
| chr12:47717574-47718575 | 8 | -0.23 | 0.03 | 2.54E-05 | *ENDOU* | exon_2 |
| chr2:43067873-43068168 | 3 | -0.74 | 0.11 | 2.58E-05 | | |
| chr14:67492294-67492679 | 2 | -1.9 | 0.29 | 2.60E-05 | | |
| chr22:42794152-42794226 | 2 | -0.55 | 0.08 | 2.73E-05 | | |
| chr8:133102961-133102966 | 2 | 0.05 | 0.01 | 2.91E-05 | *SLA* | TSS1500 |
| chr8:1862131-1862267 | 2 | -0.93 | 0.14 | 2.93E-05 | | |
| chr2:30230357-30230467 | 2 | -1 | 0.16 | 3.01E-05 | *LBH* | TSS1500 |
| chr16:1769900-1770374 | 3 | -0.47 | 0.07 | 3.04E-05 | *MAPK8IP3* | 3UTR |
| chr1:22216317-22216651 | 2 | -0.52 | 0.08 | 3.04E-05 | | |
| chr14:95257622-95257687 | 2 | -0.72 | 0.11 | 3.08E-05 | | |

| | | | | | | |
|---|---|---|---|---|---|---|
| chr19:42301170-42301662 | 3 | -0.46 | 0.07 | 3.14E-05 | *PRR19* | TSS1500 |
| chr11:73342785-73342803 | 4 | -0.081 | 0.01 | 3.17E-05 | | |
| chr11:72779826-72780294 | 3 | -0.69 | 0.10 | 3.20E-05 | | |
| chr10:5185349-5185794 | 2 | -0.78 | 0.12 | 3.21E-05 | *AKR1C8* | TSS1500 |
| chr1:48680925-48681222 | 2 | 0.56 | 0.08 | 3.38E-05 | | |
| chr11:47378605-47378648 | 3 | -0.61 | 0.09 | 3.49E-05 | *SPI1* | TSS200 |
| chr1:150272362-150272641 | 2 | -1.2 | 0.19 | 3.58E-05 | *C1orf54* | TSS1500 |
| chr7:139110169-139110295 | 3 | -0.83 | 0.13 | 3.62E-05 | *ZC3HAV1* | TSS1500 |
| chr8:126498079-126498116 | 2 | 0.036 | 0.01 | 3.63E-05 | | |
| chr1:1015131-1015591 | 2 | -0.89 | 0.13 | 3.73E-05 | | |
| chr19:48614375-48614857 | 2 | 0.52 | 0.08 | 3.80E-05 | *FAM83E* | 5UTR |
| chr2:120758741-120758957 | 2 | -0.56 | 0.09 | 4.07E-05 | | |
| chr9:125632878-125633104 | 2 | -0.54 | 0.08 | 4.26E-05 | | |
| chr1:202295785-202296022 | 2 | 0.6 | 0.09 | 4.26E-05 | | |
| chr2:16319177-16320075 | 4 | -0.33 | 0.05 | 4.28E-05 | | |
| chr14:68922236-68922317 | 2 | -0.56 | 0.09 | 4.32E-05 | | |
| chr20:46353342-46353391 | 2 | -0.64 | 0.10 | 4.43E-05 | | |
| chr6:32167088-32167141 | 2 | -0.78 | 0.12 | 4.62E-05 | *EGFL8* | exon_6 |
| chr1:11139612-11139648 | 2 | -0.37 | 0.06 | 4.69E-05 | *MTOR* | exon_35 |
| chr8:101631667-101632146 | 3 | -0.35 | 0.05 | 4.76E-05 | *GRHL2* | exon_10 |
| chr1:164625522-164625949 | 2 | -0.46 | 0.07 | 4.87E-05 | | |
| chr4:36026248-36026588 | 2 | -0.097 | 0.02 | 5.08E-05 | | |
| chr1:2505761-2506219 | 2 | -0.34 | 0.05 | 5.14E-05 | | |
| chr17:29150400-29150626 | 2 | -0.56 | 0.09 | 5.23E-05 | | |
| chr14:103712823-103713323 | 2 | -0.45 | 0.07 | 5.24E-05 | | |
| chr7:112790599-112790652 | 2 | -1.4 | 0.21 | 5.25E-05 | *TMEM168* | TSS1500 |
| chr10:24618421-24618701 | 2 | 1 | 0.16 | 5.26E-05 | | |
| chr7:5349150-5349264 | 2 | -0.85 | 0.13 | 5.30E-05 | | |
| chr22:33853743-33853937 | 2 | -0.65 | 0.10 | 5.36E-05 | | |
| chr19:49689285-49689482 | 3 | -0.13 | 0.02 | 5.38E-05 | *ADM5* | TSS1500 |
| chr16:10818642-10818688 | 2 | -0.82 | 0.13 | 5.57E-05 | *TVP23A* | exon_1 |
| chr8:37781866-37782235 | 2 | -0.47 | 0.07 | 5.58E-05 | | |
| chr2:38107036-38107103 | 2 | -0.4 | 0.06 | 5.62E-05 | | |
| chr17:77319026-77319755 | 9 | 0.29 | 0.04 | 5.62E-05 | *SEPTIN9* | TSS200 |
| chr2:60536222-60536307 | 3 | 0.045 | 0.01 | 5.74E-05 | | |
| chr2:27307802-27308016 | 3 | -0.4 | 0.06 | 5.76E-05 | *UCN* | exon_2 |
| chr2:217978712-217978781 | 2 | 0.21 | 0.03 | 5.79E-05 | *TNS1-AS1* | exon_1 |
| chr1:154404927-154404953 | 2 | -2.3 | 0.36 | 5.83E-05 | *IL6R* | TSS1500 |
| chr12:2628107-2628671 | 3 | -0.68 | 0.10 | 5.94E-05 | | |
| chr16:70699929-70700424 | 4 | -0.28 | 0.04 | 5.96E-05 | | |
| chr11:10729343-10729426 | 2 | -0.46 | 0.07 | 6.02E-05 | | |
| chr1:38153305-38153538 | 2 | -0.16 | 0.02 | 6.11E-05 | | |
| chr1:219928935-219929019 | 4 | -0.38 | 0.06 | 6.20E-05 | *SLC30A10* | TSS1500 |
| chr9:136529700-136530190 | 2 | -0.48 | 0.07 | 6.22E-05 | | |
| chr1:204359249-204359715 | 2 | -2 | 0.31 | 6.42E-05 | *PLEKHA6* | 5UTR |
| chr6:168563697-168563830 | 2 | -1.2 | 0.19 | 6.49E-05 | | |
| chr6:16305473-16305865 | 2 | -0.54 | 0.08 | 6.54E-05 | *ATXN1* | 3UTR |
| chr12:124532129-124532588 | 2 | -1.1 | 0.17 | 6.57E-05 | | |

| | | | | | | |
|---|---|---|---|---|---|---|
| chr10:101121357-101121519 | 2 | 0.38 | 0.06 | 6.82E-05 | | |
| chr19:17497301-17497565 | 2 | -0.71 | 0.11 | 6.91E-05 | *SLC27A1* | exon_7 |
| chr11:66386614-66386642 | 2 | 0.33 | 0.05 | 7.02E-05 | | |
| chr11:117831194-117831491 | 2 | -1.4 | 0.21 | 7.04E-05 | | |
| chr19:10353203-10353644 | 2 | -0.72 | 0.11 | 7.27E-05 | *TYK2* | exon_21 |
| chr3:177349291-177349529 | 2 | -0.4 | 0.06 | 7.37E-05 | | |
| chr9:127878346-127878507 | 4 | -0.065 | 0.01 | 7.41E-05 | *AK1* | TSS1500 |
| chr7:4739088-4739250 | 3 | 0.54 | 0.08 | 7.48E-05 | | |
| chr1:202122816-202122877 | 3 | -1.1 | 0.16 | 7.49E-05 | *GPR37L1* | TSS200 |
| chr16:88877125-88877134 | 2 | -0.71 | 0.11 | 7.95E-05 | *CBFA2T3* | exon_11 |
| chr1:155043722-155043887 | 2 | -1.2 | 0.18 | 8.17E-05 | | |
| chr1:153534579-153535303 | 3 | -0.21 | 0.03 | 8.21E-05 | *S100A6* | exon_2 |
| chr9:114894748-114894957 | 2 | 0.53 | 0.08 | 8.22E-05 | | |
| chr16:4335354-4335381 | 2 | -1.2 | 0.18 | 8.66E-05 | *GLIS2* | exon_6 |
| chr19:16066839-16067281 | 3 | -0.17 | 0.03 | 8.85E-05 | *TPM4* | TSS1500 |
| chr1:16981597-16981808 | 2 | -0.42 | 0.06 | 8.89E-05 | *MFAP2* | TSS1500 |
| chr2:222320946-222321019 | 2 | -0.5 | 0.08 | 8.95E-05 | *CT75* | exon_1 |
| chr1:31521188-31521214 | 2 | -0.16 | 0.02 | 8.99E-05 | | |
| chr15:69086857-69086928 | 2 | -0.5 | 0.08 | 9.08E-05 | | |
| chr11:13316185-13316473 | 2 | -0.41 | 0.06 | 9.09E-05 | | |
| chr16:85622767-85623072 | 2 | -0.77 | 0.12 | 9.41E-05 | | |
| chr8:144179035-144179273 | 2 | -0.83 | 0.13 | 9.49E-05 | | |
| chr2:221555386-221555752 | 2 | -0.54 | 0.08 | 9.60E-05 | | |
| chr3:195105663-195105682 | 2 | 0.31 | 0.05 | 9.73E-05 | | |
| chr11:27055256-27055272 | 2 | -0.3 | 0.05 | 9.76E-05 | | |
| chr1:231049504-231049614 | 2 | 0.51 | 0.08 | 9.89E-05 | | |
| chr20:2694051-2694426 | 2 | -0.39 | 0.06 | 9.99E-05 | | |
| chr6:33315315-33315368 | 2 | 0.36 | 0.06 | 0.0001 | *TAPBP* | TSS1500 |
| chr5:180067929-180068374 | 2 | -0.36 | 0.06 | 0.000105 | | |
| chr1:155131093-155131574 | 3 | -0.62 | 0.10 | 0.000105 | *EF1* | exon_2 |
| chr9:137007520-137007531 | 2 | -0.43 | 0.07 | 0.000105 | *ABCA2* | 3UTR |
| chr1:204686116-204686550 | 3 | -0.51 | 0.08 | 0.000106 | *LRRN2* | TSS1500 |
| chr1:94048526-94048992 | 2 | -0.71 | 0.11 | 0.000108 | | |
| chr4:102353956-102353964 | 2 | -0.5 | 0.08 | 0.000109 | | |
| chr6:1607617-1607687 | 2 | 1.3 | 0.20 | 0.000109 | | |
| chr22:46106404-46107245 | 6 | -0.081 | 0.01 | 0.000113 | | |
| chr15:31266252-31266563 | 2 | -0.54 | 0.08 | 0.000117 | | |
| chr1:91830389-91830580 | 2 | -0.61 | 0.10 | 0.000117 | | |
| chr11:76408736-76409069 | 2 | -0.51 | 0.08 | 0.000118 | | |
| chr1:153356984-153357406 | 3 | -0.39 | 0.06 | 0.000118 | *S100A9* | TSS1500 |
| chr2:226795351-226795357 | 2 | -2.2 | 0.34 | 0.000119 | *IRS1* | exon_1 |
| chr17:1896257-1896433 | 2 | 0.96 | 0.15 | 0.000119 | | |
| chr1:1049003-1049048 | 2 | 0.2 | 0.03 | 0.00012 | *AGRN* | exon_24 |
| chr17:29042429-29042746 | 2 | 1.6 | 0.25 | 0.000122 | *PIPOX* | TSS1500 |
| chr18:48119390-48119435 | 2 | -0.59 | 0.09 | 0.000122 | | |
| chr12:48819412-48819587 | 2 | -0.5 | 0.08 | 0.000125 | | |
| chr5:151087278-151087663 | 2 | -0.46 | 0.07 | 0.000126 | *TNIP1* | TSS1500 |
| chr3:49814280-49814288 | 2 | -0.47 | 0.07 | 0.000127 | *UBA7* | TSS1500 |

| | | | | | | |
|---|---|---|---|---|---|---|
| chr11:8658922-8658947 | 2 | -0.83 | 0.13 | 0.000127 | *TRIM66* | TSS200 |
| chr7:108004630-108004895 | 2 | -0.28 | 0.04 | 0.000129 | *LAMB1* | TSS1500 |
| chr6:29680583-29680602 | 2 | 0.01 | 0.00 | 0.00013 | | |
| chr11:68051297-68051663 | 2 | -1.5 | 0.23 | 0.000132 | | |
| chr10:71757200-71757615 | 2 | -0.41 | 0.06 | 0.000135 | | |
| chr4:6917991-6918386 | 3 | -0.27 | 0.04 | 0.000142 | | |
| chr7:74027504-74027659 | 2 | -0.49 | 0.08 | 0.000144 | *ELN* | TSS1500 |
| chr19:859065-859330 | 2 | 0.33 | 0.05 | 0.000147 | *CFD* | TSS1500 |
| chr22:23772503-23772583 | 2 | -0.11 | 0.02 | 0.000148 | *MMP11* | TSS1500 |
| chr20:57410267-57410697 | 2 | -0.71 | 0.11 | 0.000149 | | |
| chr11:66265865-66265985 | 2 | -1 | 0.16 | 0.000161 | *KLC2* | exon_13 |
| chr2:235577683-235577703 | 2 | -0.57 | 0.09 | 0.000163 | | |
| chr6:10585450-10585593 | 2 | -0.76 | 0.12 | 0.000164 | *GCNT2* | TSS1500 |
| chr14:75430673-75431059 | 3 | -0.43 | 0.07 | 0.000165 | *JDP2* | TSS1500 |
| chr7:5497353-5497495 | 2 | -0.55 | 0.09 | 0.000168 | *MIR589* | TSS1500 |
| chr7:151807778-151808030 | 2 | -0.17 | 0.03 | 0.000172 | | |
| chr3:139477649-139477937 | 2 | -0.33 | 0.05 | 0.000172 | *RBP2* | TSS1500 |
| chr15:74787774-74788048 | 2 | 1.6 | 0.25 | 0.000173 | *CSK* | TSS1500 |
| chr8:51961048-51961373 | 2 | 1.1 | 0.17 | 0.000173 | | |
| chr8:29193146-29193276 | 3 | -0.46 | 0.07 | 0.000179 | | |
| chr1:2073621-2073741 | 3 | -0.95 | 0.15 | 0.00019 | *PRKCZ* | exon_1 |
| chr1:215005727-215005848 | 3 | -0.46 | 0.07 | 0.000192 | *KCNK2* | 5UTR |
| chr19:13028773-13028975 | 2 | -0.47 | 0.07 | 0.000198 | | |
| chr4:6417113-6417541 | 2 | -0.93 | 0.15 | 0.000203 | | |
| chr17:81350237-81350241 | 2 | -0.41 | 0.06 | 0.000204 | | |
| chr17:75510149-75510377 | 2 | -0.55 | 0.09 | 0.000205 | *CASKIN2* | TSS1500 |
| chr14:76913339-76913776 | 2 | -0.25 | 0.04 | 0.000208 | | |
| chr9:137425007-137425040 | 2 | -0.28 | 0.04 | 0.000209 | | |
| chr2:28981251-28981334 | 3 | -0.14 | 0.02 | 0.000211 | *TOGARAM2* | 5UTR |
| chr1:26017377-26017752 | 2 | -0.3 | 0.05 | 0.000212 | | |
| chr14:90491752-90491790 | 3 | -0.037 | 0.01 | 0.000212 | | |
| chr16:12436050-12436257 | 2 | -1.9 | 0.29 | 0.000216 | | |
| chr8:38417411-38417674 | 3 | -0.61 | 0.10 | 0.000217 | *FGFR1* | exon_12 |
| chr3:37992443-37992744 | 2 | -0.48 | 0.08 | 0.000237 | *VILL* | TSS1500 |
| chr22:46006305-46006693 | 5 | -0.38 | 0.06 | 0.00024 | *LOC730668* | TSS200 |
| chr9:134134800-134134892 | 2 | -0.61 | 0.10 | 0.000241 | *WDR5* | TSS1500 |
| chr7:149238970-149239248 | 3 | -0.26 | 0.04 | 0.000242 | *ZNF212* | TSS1500 |
| chr6:43644890-43645027 | 3 | -0.74 | 0.12 | 0.000243 | *RSPH9* | TSS200 |
| chr18:44777943-44778225 | 2 | -0.6 | 0.10 | 0.000253 | | |
| chr10:100519946-100520001 | 2 | -1.3 | 0.21 | 0.000254 | *SEC31B* | TSS200 |
| chr18:46946737-46946793 | 4 | -0.67 | 0.10 | 0.000258 | *KATL2* | TSS200 |
| chr10:45000523-45000639 | 3 | -0.39 | 0.06 | 0.00026 | *ZNF22-AS1* | exon_1 |
| chr7:128910913-128911011 | 2 | 1.1 | 0.18 | 0.000261 | *KCP* | TSS1500 |
| chr11:43922213-43922270 | 2 | -0.21 | 0.03 | 0.000262 | *ALKBH3-AS1* | TSS1500 |
| chr5:148850995-148851066 | 2 | -1.6 | 0.25 | 0.000262 | | |
| chr17:1181359-1181729 | 2 | -0.37 | 0.06 | 0.000264 | *ABR* | TSS1500 |
| chr6:41639131-41639799 | 4 | -0.46 | 0.07 | 0.000266 | | |
| chr2:10141399-10141804 | 2 | -0.87 | 0.14 | 0.000267 | | |

| | | | | | | |
|---|---|---|---|---|---|---|
| chr20:45101470-45101600 | 3 | 0.083 | 0.01 | 0.000273 | *KCNS1* | TSS1500 |
| chr11:64284266-64284281 | 2 | 0.14 | 0.02 | 0.000279 | *GPR137* | TSS1500 |
| chr14:96085249-96085599 | 2 | -0.34 | 0.05 | 0.000281 | | |
| chr9:134646930-134647052 | 2 | -0.36 | 0.06 | 0.000289 | | |
| chr16:84054908-84055072 | 2 | -0.81 | 0.13 | 0.00029 | | |
| chr1:201347580-201347940 | 2 | -0.57 | 0.09 | 0.000292 | | |
| chr16:72947304-72947644 | 2 | 0.4 | 0.06 | 0.000292 | | |
| chr9:127528875-127529376 | 3 | -0.88 | 0.14 | 0.000297 | | |
| chr13:77743021-77743027 | 2 | -0.32 | 0.05 | 0.000299 | *SLAIN1* | 5UTR |
| chr2:109350335-109350716 | 2 | -0.49 | 0.08 | 0.000302 | | |
| chr11:625261-625274 | 2 | -1 | 0.16 | 0.000304 | *CDHR5* | TSS1500 |
| chr1:2519600-2519965 | 2 | -1.1 | 0.18 | 0.000306 | | |
| chr17:41522423-41522427 | 2 | -0.3 | 0.05 | 0.000309 | | |
| chr5:135536562-135536949 | 4 | 0.21 | 0.03 | 0.000317 | *NEUROG1* | TSS1500 |
| chr2:54524370-54524848 | 2 | -0.39 | 0.06 | 0.000319 | | |
| chr6:33211948-33211978 | 2 | 0.37 | 0.06 | 0.000322 | *RING1* | exon_6 |
| chr19:39071013-39071022 | 2 | -1.1 | 0.17 | 0.000325 | | |
| chr9:136352922-136352984 | 2 | -0.26 | 0.04 | 0.00034 | *GPSM1* | TSS200 |
| chr6:45473357-45473696 | 2 | -0.84 | 0.13 | 0.000355 | | |
| chr15:80581002-80581039 | 2 | -0.32 | 0.05 | 0.000361 | *MIR5572* | TSS200 |
| chr6:26225975-26226028 | 2 | -0.27 | 0.04 | 0.000362 | *H3C6* | 3UTR |
| chr6:113564619-113564844 | 2 | -0.41 | 0.07 | 0.000363 | | |
| chr11:68371037-68371394 | 2 | -0.5 | 0.08 | 0.000369 | | |
| chr9:129319456-129319904 | 2 | -0.81 | 0.13 | 0.000378 | *LINC02913* | TSS1500 |
| chr2:68832016-68832487 | 3 | -0.62 | 0.10 | 0.000381 | | |
| chr11:13440235-13440255 | 2 | -0.31 | 0.05 | 0.000384 | *BTBD10* | 5UTR |
| chr12:15156809-15156939 | 2 | -1.2 | 0.19 | 0.000385 | | |
| chr1:206035131-206035273 | 2 | 0.83 | 0.13 | 0.000386 | | |
| chr8:22554265-22554300 | 2 | -0.72 | 0.11 | 0.000387 | | |
| chr1:9414040-9414368 | 2 | -0.45 | 0.07 | 0.000396 | | |
| chr3:58572606-58572616 | 2 | -0.73 | 0.12 | 0.0004 | | |
| chr1:1315447-1315637 | 2 | -0.87 | 0.14 | 0.000406 | | |
| chr5:77139104-77139189 | 2 | -0.8 | 0.13 | 0.000407 | | |
| chr9:107260486-107260848 | 2 | -0.52 | 0.08 | 0.000409 | | |
| chr11:2460912-2461334 | 12 | -0.2 | 0.03 | 0.00041 | *KCNQ1* | TSS1500 |
| chr3:101940538-101940786 | 2 | -0.61 | 0.10 | 0.000418 | *LINC02085* | TSS1500 |
| chr7:135147530-135147792 | 3 | 0.025 | 0.00 | 0.000426 | *TMEM140* | TSS1500 |
| chr2:205686742-205687120 | 2 | 0.41 | 0.07 | 0.000437 | | |
| chr11:65310566-65310696 | 2 | -0.8 | 0.13 | 0.000444 | | |
| chr15:89090867-89091106 | 2 | -0.3 | 0.05 | 0.000446 | | |
| chr16:27769904-27769974 | 2 | -0.88 | 0.14 | 0.000449 | *KATNIP* | exon_21 |
| chr7:151736272-151736328 | 2 | -0.015 | 0.00 | 0.000451 | *PRKAG2* | TSS200 |
| chr20:44407396-44407641 | 2 | -0.58 | 0.09 | 0.000452 | *HNF4A* | exon_3 |
| chr9:133421545-133421950 | 6 | -0.35 | 0.06 | 0.000461 | *ADAMTS13* | TSS1500 |
| chr11:75559024-75559169 | 2 | -0.28 | 0.05 | 0.000462 | | |
| chr8:118074471-118074523 | 2 | -0.54 | 0.09 | 0.000483 | | |
| chr5:135534824-135534827 | 2 | -0.15 | 0.02 | 0.000486 | *NEUROG1* | 3UTR |
| chr10:49488308-49488354 | 2 | -1.3 | 0.21 | 0.000502 | | |

| | | | | | | |
|---|---|---|---|---|---|---|
| chr11:781977-782397 | 2 | -0.44 | 0.07 | 0.000531 | | |
| chr16:46885163-46885186 | 2 | -0.65 | 0.10 | 0.000536 | *GPT2* | 5UTR |
| chr10:63703494-63703882 | 2 | -0.49 | 0.08 | 0.000546 | | |
| chr11:62908084-62908139 | 2 | -0.96 | 0.15 | 0.000555 | | |
| chr16:22346505-22346730 | 2 | -0.37 | 0.06 | 0.000573 | *CDR2* | 3UTR |
| chr17:59381937-59382159 | 2 | -0.29 | 0.05 | 0.000573 | | |
| chr19:7255031-7255266 | 2 | -0.4 | 0.06 | 0.000573 | | |
| chr1:89844000-89844439 | 3 | -0.51 | 0.08 | 0.000576 | | |
| chr6:31871196-31871248 | 2 | -0.57 | 0.09 | 0.000581 | | |
| chr4:76948437-76948499 | 2 | -1.2 | 0.20 | 0.000585 | *SEPTIN11* | TSS1500 |
| chr8:144515794-144516029 | 3 | -0.3 | 0.05 | 0.000595 | *RECQL4* | exon_5 |
| chr7:2924069-2924279 | 2 | -0.59 | 0.10 | 0.000596 | *CARD11* | exon_15 |
| chr13:106468446-106468454 | 2 | -0.24 | 0.04 | 0.000599 | | |
| chr17:79949476-79949515 | 2 | -1 | 0.16 | 0.000602 | | |
| chr1:167713411-167713492 | 2 | 0.57 | 0.09 | 0.000604 | | |
| chr3:57948945-57949296 | 2 | -0.7 | 0.11 | 0.000606 | | |
| chr3:9168432-9168472 | 2 | -0.4 | 0.06 | 0.000606 | | |
| chr1:167483754-167484143 | 2 | -0.54 | 0.09 | 0.000621 | | |
| chr10:110658485-110658950 | 2 | 0.26 | 0.04 | 0.000627 | | |
| chr17:8669044-8669099 | 2 | 0.46 | 0.07 | 0.000636 | | |
| chr1:87685382-87685713 | 2 | -0.58 | 0.09 | 0.000636 | | |
| chr14:103698940-103699064 | 2 | -0.26 | 0.04 | 0.000662 | *XRCC3* | exon_10 |
| chr7:43249598-43249678 | 2 | 0.32 | 0.05 | 0.000675 | | |
| chr8:27611464-27611628 | 3 | -0.34 | 0.06 | 0.000685 | *CLU* | TSS1500 |
| chr7:120988820-120988997 | 2 | 0.54 | 0.09 | 0.000689 | *CPED1* | TSS1500 |
| chr1:209768498-209768503 | 2 | 0.24 | 0.04 | 0.000714 | *TRAF3IP3* | TSS200 |
| chr7:135147919-135148018 | 2 | -0.49 | 0.08 | 0.000719 | *TMEM140* | TSS200 |
| chr20:20440618-20440754 | 2 | -1.2 | 0.19 | 0.00073 | | |
| chr9:134049341-134049379 | 2 | -0.37 | 0.06 | 0.000736 | | |
| chr11:67404923-67405055 | 2 | -0.62 | 0.10 | 0.000741 | | |
| chr15:58980171-58980329 | 4 | 0.095 | 0.02 | 0.000745 | | |
| chr22:28953334-28953642 | 2 | -0.57 | 0.09 | 0.000747 | | |
| chr9:135886576-135886838 | 2 | 0.48 | 0.08 | 0.000757 | | |
| chr17:82838867-82839099 | 2 | -0.27 | 0.04 | 0.000777 | | |
| chr17:42681815-42682006 | 2 | -0.39 | 0.06 | 0.000781 | *CCR10* | TSS1500 |
| chr19:52269118-52269213 | 2 | 0.076 | 0.01 | 0.000789 | *ZNF766* | TSS1500 |
| chr16:2989445-2989664 | 2 | -0.62 | 0.10 | 0.000793 | *GREP1* | TSS1500 |
| chr14:90513890-90514226 | 4 | -0.058 | 0.01 | 0.000798 | | |
| chr3:194687827-194688172 | 3 | -0.46 | 0.08 | 0.000802 | *FAM43A* | exon_1 |
| chr5:149271336-149271806 | 6 | -0.35 | 0.06 | 0.000803 | *AFAP1L1* | TSS1500 |
| chr16:68404973-68404989 | 2 | -0.39 | 0.06 | 0.000809 | | |
| chr1:150257698-150257700 | 2 | -0.31 | 0.05 | 0.000819 | *CA14* | TSS200 |
| chr13:113616479-113616974 | 3 | -0.12 | 0.02 | 0.000823 | | |
| chr22:37984432-37984480 | 2 | -0.76 | 0.12 | 0.000836 | *SOX10* | 5UTR |
| chr11:76670104-76670121 | 2 | -1 | 0.17 | 0.000845 | *LRRC32* | TSS200 |
| chr10:80356129-80356201 | 2 | -0.68 | 0.11 | 0.000874 | *DYDC2* | TSS1500 |
| chr11:75337062-75337320 | 2 | 0.89 | 0.15 | 0.000883 | | |
| chr12:41188265-41188335 | 2 | -0.32 | 0.05 | 0.000908 | *PDZRN4* | 5UTR |

| | | | | | | |
|---|---|---|---|---|---|---|
| chr6:36685994-36686196 | 2 | -0.24 | 0.04 | 0.000908 | *CDKN1A* | 3UTR |
| chr3:129688522-129688705 | 5 | 0.023 | 0.00 | 0.000923 | *TMCC1* | exon_1 |
| chr20:25293791-25293795 | 2 | 0.43 | 0.07 | 0.000927 | | |
| chr6:40715522-40715606 | 2 | 0.63 | 0.10 | 0.000939 | | |
| chr2:54557157-54557265 | 2 | -1.2 | 0.19 | 0.000961 | *SPTBN1* | TSS1500 |
| chr15:41881415-41881887 | 3 | -0.11 | 0.02 | 0.000968 | | |
| chr11:47391188-47391536 | 2 | -0.6 | 0.10 | 0.000992 | | |
| chr7:6077237-6077347 | 2 | -0.63 | 0.10 | 0.000996 | | |
| chr1:43704718-43704794 | 2 | -0.34 | 0.06 | 0.00101 | *KDM4A* | 3UTR |
| chr11:8266542-8266951 | 2 | -1.5 | 0.24 | 0.00101 | | |
| chr16:8720644-8720650 | 2 | -1.7 | 0.28 | 0.00101 | *ABAT* | TSS200 |
| chr3:66463856-66464381 | 4 | -0.13 | 0.02 | 0.00101 | | |
| chr2:191845948-191846383 | 3 | 0.21 | 0.04 | 0.00102 | | |
| chr10:34937659-34938051 | 2 | -0.5 | 0.08 | 0.00102 | | |
| chr18:23452102-23452482 | 3 | -0.26 | 0.04 | 0.00106 | *RIOK3* | TSS1500 |
| chr7:8176266-8176442 | 3 | 0.19 | 0.03 | 0.00108 | | |
| chr20:50981482-50981873 | 2 | -0.32 | 0.05 | 0.00108 | | |
| chr13:77696445-77696901 | 2 | 0.36 | 0.06 | 0.00108 | *SLAIN1* | TSS1500 |
| chr9:112098457-112098540 | 2 | -0.55 | 0.09 | 0.00111 | *SUSD1* | exon_10 |
| chr17:6480799-6480995 | 2 | -1 | 0.16 | 0.00111 | | |
| chr1:1779746-1780149 | 3 | -0.32 | 0.05 | 0.00111 | *DK* | TSS1500 |
| chr6:26018306-26018374 | 3 | -0.54 | 0.09 | 0.00113 | *H1-1* | TSS1500 |
| chr6:170297373-170297418 | 2 | -0.53 | 0.09 | 0.00113 | | |
| chr15:22917396-22917510 | 2 | -0.52 | 0.09 | 0.00117 | *CYFIP1* | TSS200 |
| chr14:24369163-24369196 | 2 | -0.64 | 0.10 | 0.00117 | *NFATC4* | 5UTR |
| chr1:36150263-36150427 | 2 | -0.32 | 0.05 | 0.00117 | *TRAPPC3* | TSS1500 |
| chr15:66919737-66919793 | 2 | -0.52 | 0.09 | 0.00118 | | |
| chr2:69619902-69619977 | 2 | -0.36 | 0.06 | 0.00118 | | |
| chr15:90813807-90814291 | 2 | 0.53 | 0.09 | 0.00119 | | |
| chr2:172977304-172977371 | 2 | -0.9 | 0.15 | 0.00119 | | |
| chr9:134401463-134401482 | 2 | -1.1 | 0.18 | 0.00119 | | |
| chr1:201990519-201990609 | 2 | -0.46 | 0.08 | 0.0012 | | |
| chr3:47442501-47443001 | 2 | -0.37 | 0.06 | 0.0012 | *SCAP* | 5UTR |
| chr8:96853255-96854135 | 3 | -0.33 | 0.05 | 0.00122 | | |
| chr11:114366968-114367436 | 2 | -0.25 | 0.04 | 0.00124 | | |
| chr19:44757244-44757392 | 2 | -0.5 | 0.08 | 0.00128 | *MIR8085* | TSS1500 |
| chr2:10285535-10285587 | 2 | -0.75 | 0.12 | 0.0013 | | |
| chr10:69730601-69730982 | 3 | -0.13 | 0.02 | 0.00131 | | |
| chr14:105461947-105462053 | 2 | -0.97 | 0.16 | 0.00132 | | |
| chr2:120766033-120766106 | 3 | -0.48 | 0.08 | 0.00134 | | |
| chr5:172983438-172983486 | 2 | -1.2 | 0.20 | 0.00135 | *ATP6V0E1* | TSS1500 |
| chr9:137113436-137113587 | 2 | -0.44 | 0.07 | 0.00135 | *DPP7* | exon_5 |
| chr14:99414236-99414304 | 2 | 0.42 | 0.07 | 0.00136 | | |
| chr10:104333960-104334061 | 3 | -0.16 | 0.03 | 0.00136 | *ITPRIP* | TSS200 |
| chr1:153628389-153628496 | 2 | -0.92 | 0.15 | 0.0014 | *S100A13* | TSS1500 |
| chr22:38751500-38751831 | 3 | -0.38 | 0.06 | 0.00141 | | |
| chr11:62879399-62879805 | 3 | -0.74 | 0.12 | 0.00141 | *SLC3A2* | TSS1500 |
| chr5:16758113-16758143 | 2 | -0.82 | 0.14 | 0.00143 | *MYO10* | exon_18 |

| | | | | | | |
|---|---|---|---|---|---|---|
| chr6:31792029-31792048 | 2 | -1 | 0.17 | 0.00145 | | |
| chr22:46289831-46289879 | 3 | 0.036 | 0.01 | 0.00145 | *TTC38* | exon_13 |
| chr12:590421-590605 | 4 | 0.069 | 0.01 | 0.00148 | | |
| chr9:34624720-34624735 | 3 | -0.46 | 0.08 | 0.00149 | | |
| chr3:45915774-45915827 | 2 | 1.5 | 0.24 | 0.00154 | *LZTFL1* | TSS200 |
| chr16:57985962-57985989 | 2 | 0.56 | 0.09 | 0.00156 | | |
| chr15:40554081-40554314 | 2 | -1.2 | 0.20 | 0.00156 | *CCDC32* | exon_4 |
| chr2:174326003-174326007 | 2 | 0.16 | 0.03 | 0.0016 | *LINC01305* | TSS200 |
| chr12:56993255-56993373 | 2 | 0.47 | 0.08 | 0.00161 | *GPR182* | TSS1500 |
| chr16:70725192-70725731 | 3 | -0.68 | 0.11 | 0.00163 | | |
| chr3:150248905-150249117 | 2 | -1.5 | 0.25 | 0.00165 | | |
| chr2:16522299-16522703 | 2 | -0.49 | 0.08 | 0.00165 | | |
| chr7:112080753-112081178 | 2 | 0.27 | 0.05 | 0.00171 | | |
| chr6:17505404-17505472 | 2 | -0.58 | 0.10 | 0.00171 | | |
| chr11:64990391-64990459 | 2 | -0.77 | 0.13 | 0.00172 | *BATF2* | TSS200 |
| chr1:27635169-27635235 | 2 | -0.41 | 0.07 | 0.00181 | *FGR* | 5UTR |
| chr11:65592850-65592856 | 2 | 0.12 | 0.02 | 0.00184 | *KCNK7* | 3UTR |
| chr15:65044822-65045087 | 2 | -1.4 | 0.23 | 0.00184 | *SLC51B* | TSS1500 |
| chr11:12461609-12461657 | 2 | -0.83 | 0.14 | 0.00191 | | |
| chr15:99997099-99997400 | 2 | 0.21 | 0.04 | 0.00193 | *ADAMTS17* | exon_19 |
| chr16:58265249-58265741 | 2 | -0.093 | 0.02 | 0.00195 | | |
| chr2:106190101-106190103 | 2 | -0.26 | 0.04 | 0.002 | | |
| chr12:2295723-2296146 | 2 | -0.68 | 0.11 | 0.00202 | | |
| chr12:124543749-124543756 | 2 | -0.44 | 0.07 | 0.00203 | | |
| chr3:10290343-10290486 | 2 | 1.1 | 0.18 | 0.00204 | | |
| chr1:151841570-151841595 | 2 | -0.78 | 0.13 | 0.00206 | *C2CD4D* | TSS1500 |
| chr10:111360406-111360774 | 2 | 0.59 | 0.10 | 0.00206 | | |
| chr4:1805951-1806236 | 5 | -0.24 | 0.04 | 0.00212 | *FGFR3* | exon_14 |
| chr16:31203173-31203286 | 2 | 0.58 | 0.10 | 0.00212 | *PYCARD* | TSS1500 |
| chr11:3671927-3672179 | 3 | -0.59 | 0.10 | 0.00218 | *CHR10* | TSS1500 |
| chr18:77133466-77134079 | 3 | -0.14 | 0.02 | 0.00218 | *MBP* | TSS1500 |
| chr8:60583728-60584016 | 3 | -0.38 | 0.06 | 0.00219 | | |
| chr16:57103058-57103475 | 2 | -0.21 | 0.04 | 0.0022 | | |
| chr19:49690527-49690948 | 4 | -0.29 | 0.05 | 0.00228 | *CPT1C* | TSS1500 |
| chr15:68206119-68206519 | 3 | -0.67 | 0.11 | 0.00229 | *CALML4* | TSS1500 |
| chr2:64987406-64987491 | 2 | -0.22 | 0.04 | 0.0023 | *SLC1A4* | TSS1500 |
| chr1:226163096-226163112 | 2 | -0.41 | 0.07 | 0.00231 | | |
| chr2:235807103-235807641 | 5 | -0.57 | 0.10 | 0.00232 | *AGAP1* | exon_9 |
| chr6:30892353-30892523 | 7 | -0.17 | 0.03 | 0.00232 | *DDR1* | exon_8 |
| chr12:75825748-75826014 | 2 | -0.62 | 0.10 | 0.00232 | | |
| chr1:1138044-1138130 | 2 | -0.4 | 0.07 | 0.00233 | | |
| chr12:52079678-52080263 | 3 | -0.57 | 0.10 | 0.00234 | *SMIM41* | TSS200 |
| chr19:49337995-49338201 | 2 | -0.37 | 0.06 | 0.00239 | *CD37* | exon_5 |
| chr17:7582612-7583019 | 4 | -0.39 | 0.07 | 0.00243 | *MPDU1* | TSS1500 |
| chr10:6148628-6148688 | 2 | 0.1 | 0.02 | 0.00244 | | |
| chr5:124745983-124746257 | 3 | -0.5 | 0.08 | 0.00247 | *ZNF608* | 5UTR |
| chr17:73552606-73552722 | 3 | -0.29 | 0.05 | 0.00248 | | |
| chr1:38815941-38816054 | 2 | -0.71 | 0.12 | 0.00251 | | |

| | | | | | | |
|---|---|---|---|---|---|---|
| chr7:2641735-2641802 | 2 | -0.5 | 0.08 | 0.00251 | | |
| chr8:143813073-143813177 | 2 | -0.34 | 0.06 | 0.00253 | *SCRIB* | exon_7 |
| chr2:97728270-97728410 | 2 | 0.18 | 0.03 | 0.00256 | | |
| chr14:91244130-91244403 | 2 | -0.73 | 0.12 | 0.00257 | *GPR68* | 5UTR |
| chr19:5582306-5582689 | 2 | -0.87 | 0.15 | 0.00258 | | |
| chr20:25221682-25221735 | 3 | 0.39 | 0.07 | 0.00263 | | |
| chr11:43896067-43896466 | 2 | -0.35 | 0.06 | 0.00267 | *SEC14L1P1* | TSS1500 |
| chr20:51306805-51307198 | 2 | -0.39 | 0.07 | 0.00269 | | |
| chr22:17476939-17476983 | 2 | -0.54 | 0.09 | 0.00271 | | |
| chr1:8120851-8121048 | 2 | -0.36 | 0.06 | 0.00271 | | |
| chr1:8883674-8883822 | 2 | -0.54 | 0.09 | 0.00278 | | |
| chr22:46289119-46289575 | 2 | -0.86 | 0.14 | 0.00279 | | |
| chr4:40587709-40587762 | 2 | -0.4 | 0.07 | 0.00281 | | |
| chr17:45910864-45911292 | 2 | -0.63 | 0.11 | 0.00287 | | |
| chr11:68042092-68042360 | 2 | -0.73 | 0.12 | 0.00287 | *TCIRG1* | TSS1500 |
| chr21:34865406-34865589 | 2 | -0.23 | 0.04 | 0.00298 | | |
| chr12:51271158-51271359 | 3 | 0.12 | 0.02 | 0.00299 | *SMAGP* | TSS1500 |
| chr2:201260489-201260587 | 2 | -0.31 | 0.05 | 0.00299 | *CASP8* | 5UTR |
| chr14:100223276-100223752 | 2 | -0.49 | 0.08 | 0.003 | | |
| chr6:30127418-30127471 | 3 | -0.15 | 0.03 | 0.00301 | | |
| chr14:75196474-75196869 | 2 | -0.37 | 0.06 | 0.00301 | | |
| chr14:75206502-75206505 | 2 | -0.79 | 0.13 | 0.00306 | | |
| chr12:27243773-27243832 | 2 | -0.25 | 0.04 | 0.00307 | *STK38L* | TSS1500 |
| chr19:48681300-48681371 | 2 | -0.49 | 0.08 | 0.00307 | *SEC1P* | exon_5 |
| chr20:37519918-37519973 | 2 | -0.75 | 0.13 | 0.00309 | *NT* | TSS1500 |
| chr17:82244469-82244550 | 2 | -0.59 | 0.10 | 0.00311 | *CSNK1D* | 3UTR |
| chr16:55478609-55478812 | 2 | -0.57 | 0.10 | 0.00315 | *MMP2* | TSS1500 |
| chr20:35293022-35293132 | 3 | -0.58 | 0.10 | 0.0032 | *FAM83C* | TSS1500 |
| chr7:2100596-2100686 | 2 | 0.95 | 0.16 | 0.0032 | | |
| chr9:69205076-69205083 | 2 | -0.02 | 0.00 | 0.00332 | *TJP2* | TSS200 |
| chr10:103737686-103737934 | 2 | -0.52 | 0.09 | 0.00334 | | |
| chr12:124043925-124044127 | 2 | -1.3 | 0.22 | 0.00334 | | |
| chr10:6383051-6383300 | 2 | -0.27 | 0.05 | 0.0034 | | |
| chr22:46625327-46625870 | 4 | -0.35 | 0.06 | 0.00344 | | |
| chr14:69726759-69726779 | 2 | -0.66 | 0.11 | 0.00349 | | |
| chr6:57013659-57013867 | 2 | -0.32 | 0.05 | 0.00355 | | |
| chr2:98518394-98518551 | 2 | -0.68 | 0.11 | 0.00361 | | |
| chr11:2271951-2271971 | 2 | -1.9 | 0.32 | 0.00362 | *ASCL2* | TSS1500 |
| chr15:91627584-91627863 | 2 | -0.55 | 0.09 | 0.00368 | | |
| chr3:47892446-47892770 | 2 | 1.4 | 0.24 | 0.00369 | | |
| chr15:101348500-101348783 | 2 | -0.58 | 0.10 | 0.00373 | | |
| chr8:38414050-38414246 | 2 | 2.5 | 0.42 | 0.00377 | *FGFR1* | exon_16 |
| chr9:129580292-129580373 | 2 | -0.59 | 0.10 | 0.00378 | | |
| chr1:47190465-47190751 | 3 | -0.57 | 0.10 | 0.0038 | *PDZK1IP1* | TSS1500 |
| chr2:96695156-96695297 | 2 | -0.49 | 0.08 | 0.00384 | | |
| chr17:27532356-27532416 | 2 | -0.21 | 0.04 | 0.00386 | | |
| chr1:21279724-21279821 | 2 | -0.87 | 0.15 | 0.00386 | *ECE1* | TSS1500 |
| chr1:219323703-219324310 | 3 | -0.23 | 0.04 | 0.00388 | | |

| | | | | | | |
|---|---|---|---|---|---|---|
| chr3:36908466-36908475 | 2 | -0.3 | 0.05 | 0.00403 | | |
| chr1:202122752-202122805 | 2 | -1.4 | 0.24 | 0.00404 | *GPR37L1* | TSS200 |
| chr9:105278819-105279123 | 3 | 0.016 | 0.00 | 0.00408 | | |
| chr20:19658037-19658232 | 2 | -0.52 | 0.09 | 0.0041 | | |
| chr14:64539298-64539452 | 5 | 0.15 | 0.03 | 0.00414 | *HSPA2* | TSS1500 |
| chr16:56353609-56353721 | 3 | -0.49 | 0.08 | 0.00418 | | |
| chr7:2104251-2104307 | 2 | -0.45 | 0.08 | 0.00429 | | |
| chr5:95730532-95731088 | 7 | -0.27 | 0.05 | 0.00431 | *RHOBTB3* | TSS1500 |
| chr1:178724479-178724762 | 2 | -0.86 | 0.15 | 0.00437 | *RALGPS2* | TSS1500 |
| chr10:98391523-98391853 | 2 | -0.4 | 0.07 | 0.00441 | | |
| chr20:49668623-49668723 | 2 | -0.54 | 0.09 | 0.00444 | | |
| chr15:47845116-47846016 | 4 | -0.099 | 0.02 | 0.00449 | | |
| chr2:134206534-134206657 | 2 | -0.46 | 0.08 | 0.00452 | | |
| chr11:114257595-114257795 | 2 | -0.48 | 0.08 | 0.00453 | *NNMT* | TSS1500 |
| chr1:2073785-2073821 | 2 | -2.1 | 0.36 | 0.00459 | *PRKCZ* | exon_1 |
| chr16:56782122-56782169 | 2 | -0.38 | 0.07 | 0.00462 | *NUP93* | 5UTR |
| chr11:34310001-34310361 | 2 | -0.67 | 0.11 | 0.00463 | | |
| chr16:85574452-85574618 | 2 | -0.9 | 0.15 | 0.00465 | | |
| chr6:33898332-33898512 | 3 | -0.57 | 0.10 | 0.00466 | *MIR7159* | TSS1500 |
| chr15:58552301-58552384 | 2 | 0.38 | 0.06 | 0.00466 | | |
| chr22:37691211-37691241 | 2 | 0.36 | 0.06 | 0.00469 | *NOL12* | exon_6 |
| chr10:102379314-102379353 | 2 | -0.59 | 0.10 | 0.0047 | *GBF1* | exon_34 |
| chr1:37947016-37947072 | 3 | 0.17 | 0.03 | 0.00476 | *INPP5B* | exon_1 |
| chr18:58047139-58047161 | 2 | 0.67 | 0.11 | 0.00478 | *NEDD4L* | TSS200 |
| chr5:139903614-139903677 | 2 | -0.89 | 0.15 | 0.00479 | | |
| chr11:76604469-76604724 | 2 | -0.37 | 0.06 | 0.00484 | | |
| chr22:24066594-24066815 | 2 | -0.49 | 0.08 | 0.00484 | | |
| chr21:44910988-44911101 | 2 | -0.36 | 0.06 | 0.00491 | | |
| chr10:43340782-43340812 | 2 | 0.0067 | 0.00 | 0.00492 | | |
| chr18:58629102-58629117 | 2 | -0.25 | 0.04 | 0.00498 | *ALPK2* | TSS200 |
| chr6:30988202-30988620 | 4 | -0.31 | 0.05 | 0.00499 | *MUC21* | exon_3 |
| chr4:3373040-3373103 | 2 | -0.16 | 0.03 | 0.00499 | | |
| chr11:72727357-72727726 | 3 | -0.6 | 0.10 | 0.00502 | | |
| chr1:10960800-10960886 | 3 | -1.1 | 0.19 | 0.00511 | | |
| chr7:47291248-47291413 | 3 | -0.12 | 0.02 | 0.00516 | | |
| chr17:75239041-75239098 | 2 | -1.3 | 0.22 | 0.00518 | *GGA3* | exon_14 |
| chr2:46073730-46073944 | 2 | -0.74 | 0.13 | 0.00522 | | |
| chr2:85434804-85435242 | 2 | -0.57 | 0.10 | 0.00522 | | |
| chr2:101968034-101968120 | 2 | -0.21 | 0.04 | 0.00524 | | |
| chr1:31689638-31690066 | 2 | -0.78 | 0.13 | 0.00525 | | |
| chr21:16589509-16589676 | 2 | -0.72 | 0.12 | 0.00526 | *MIR125B2* | TSS1500 |
| chr22:38423253-38423697 | 2 | -0.34 | 0.06 | 0.00526 | | |
| chr17:82900530-82900663 | 2 | -0.65 | 0.11 | 0.00526 | *TBCD* | exon_18 |
| chr8:38644553-38644934 | 2 | -0.32 | 0.06 | 0.00528 | | |
| chr11:64368617-64368791 | 3 | -0.68 | 0.12 | 0.00537 | *RPS6KA4* | exon_12 |
| chr2:202090836-202091062 | 2 | -0.55 | 0.10 | 0.00551 | *KIAA2012* | exon_3 |
| chr5:138139011-138139216 | 2 | -0.39 | 0.07 | 0.00551 | | |
| chr6:32167593-32167602 | 2 | -1.3 | 0.23 | 0.00556 | *EGFL8* | exon_8 |

| | | | | | | |
|---|---|---|---|---|---|---|
| chr2:70095478-70095527 | 2 | -0.62 | 0.11 | 0.00564 | | |
| chr5:102865433-102865717 | 2 | -0.6 | 0.10 | 0.00573 | *PAM* | TSS1500 |
| chr11:61608169-61608213 | 2 | -1.1 | 0.19 | 0.00578 | | |
| chr8:100336131-100336411 | 6 | -0.1 | 0.02 | 0.00602 | *RNF19A* | TSS200 |
| chr15:39195544-39195608 | 2 | 0.44 | 0.08 | 0.00606 | | |
| chr17:62895428-62895693 | 2 | -0.83 | 0.14 | 0.00611 | | |
| chr17:79986027-79986208 | 2 | -0.77 | 0.13 | 0.00621 | | |
| chr20:35225433-35226012 | 3 | -0.42 | 0.07 | 0.00621 | *MMP24* | TSS1500 |
| chr14:100323172-100323287 | 3 | -1.2 | 0.21 | 0.0063 | *SLC25A47* | TSS200 |
| chr7:38929518-38929753 | 2 | 0.31 | 0.05 | 0.0064 | | |
| chr10:102776278-102776295 | 2 | -0.46 | 0.08 | 0.00642 | *WBP1L* | 5UTR |
| chr12:56020658-56020749 | 4 | -0.61 | 0.11 | 0.00648 | *IKZF4* | TSS1500 |
| chr12:6020770-6020995 | 2 | -0.74 | 0.13 | 0.00649 | | |
| chr14:99414059-99414069 | 2 | -0.3 | 0.05 | 0.00655 | | |
| chr2:223047698-223047824 | 2 | -0.45 | 0.08 | 0.00659 | | |
| chr17:73261906-73261962 | 2 | -0.63 | 0.11 | 0.00659 | *CPSF4L* | TSS200 |
| chr21:33401066-33401291 | 2 | -0.4 | 0.07 | 0.00663 | | |
| chr17:19363341-19363779 | 2 | -0.49 | 0.08 | 0.00666 | *B9D1* | TSS1500 |
| chr2:120796788-120796842 | 2 | -0.6 | 0.10 | 0.00669 | | |
| chr16:8949378-8949754 | 2 | -0.23 | 0.04 | 0.00686 | *USP7* | TSS200 |
| chr7:11130062-11130251 | 2 | -1.4 | 0.24 | 0.00694 | | |
| chr3:50125836-50126568 | 3 | -0.85 | 0.15 | 0.0071 | | |
| chr8:124247449-124247573 | 2 | -0.26 | 0.04 | 0.00725 | *.OC10192758[illegible]* | TSS200 |
| chr15:80838856-80839214 | 3 | -0.49 | 0.09 | 0.00728 | | |
| chr3:64684783-64684839 | 2 | -0.11 | 0.02 | 0.00747 | *[illegible]DAMTS9-AS[illegible]* | TSS200 |
| chr7:148817867-148818044 | 2 | -0.54 | 0.09 | 0.00748 | *EZH2* | exon_10 |
| chr16:66521956-66522230 | 2 | -0.47 | 0.08 | 0.00761 | | |
| chr3:16329788-16329832 | 2 | -0.86 | 0.15 | 0.00773 | | |
| chr2:151640330-151640605 | 2 | -0.29 | 0.05 | 0.00773 | *NEB* | exon_61 |
| chr17:6894389-6894452 | 2 | -0.4 | 0.07 | 0.0079 | *ALOX12P2* | exon_2 |
| chr1:51009526-51009898 | 3 | -0.19 | 0.03 | 0.00801 | | |
| chr10:86955145-86955560 | 2 | -0.52 | 0.09 | 0.00802 | *SNCG* | TSS1500 |
| chr16:88498339-88498526 | 2 | -0.95 | 0.16 | 0.00809 | | |
| chr3:114759114-114759409 | 3 | -0.5 | 0.09 | 0.00837 | *ZBTB20* | TSS200 |
| chr7:137970504-137970776 | 2 | -0.16 | 0.03 | 0.00838 | | |
| chr1:160062339-160062345 | 2 | -0.93 | 0.16 | 0.00848 | | |
| chr1:24761915-24762364 | 2 | -0.54 | 0.09 | 0.00854 | | |
| chr6:33163542-33164386 | 8 | 0.093 | 0.02 | 0.00854 | *COL11A2* | 3UTR |
| chr8:133249670-133249685 | 2 | -0.82 | 0.14 | 0.00896 | | |
| chr11:75151956-75152145 | 2 | -0.22 | 0.04 | 0.00903 | | |
| chr10:16522621-16522672 | 2 | 0.3 | 0.05 | 0.00911 | *C1QL3* | TSS1500 |
| chr5:177357776-177357964 | 3 | -0.028 | 0.00 | 0.00912 | *RGS14* | 5UTR |
| chr8:27725809-27725836 | 2 | -0.57 | 0.10 | 0.00914 | | |
| chr7:3880296-3880374 | 2 | -0.28 | 0.05 | 0.00932 | | |
| chr9:79974823-79975095 | 2 | -0.26 | 0.05 | 0.00947 | *LINC01507* | TSS1500 |
| chr15:74202513-74202520 | 2 | -0.25 | 0.04 | 0.00948 | *STRA6* | exon_1 |
| chr5:10330733-10330872 | 2 | 0.25 | 0.04 | 0.00951 | | |
| chr16:77223410-77223509 | 3 | -0.43 | 0.07 | 0.00958 | | |

| | | | | | | |
|---|---|---|---|---|---|---|
| chr8:763642-763913 | 2 | -0.34 | 0.06 | 0.00961 | | |
| chr1:26289990-26290043 | 2 | -0.34 | 0.06 | 0.00962 | | |
| chr21:46083534-46083543 | 2 | -0.8 | 0.14 | 0.00963 | | |
| chr16:31711804-31712149 | 2 | -0.4 | 0.07 | 0.00972 | *KRBOX5* | TSS1500 |
| chr17:79979289-79979305 | 2 | -0.33 | 0.06 | 0.00972 | | |
| chr7:151728781-151728821 | 2 | -1.3 | 0.22 | 0.00985 | | |
| chr11:108788642-108788997 | 2 | -0.54 | 0.10 | 0.00986 | | |
| chr10:13101286-13101470 | 2 | -0.74 | 0.13 | 0.00986 | *CCDC3* | TSS1500 |
| chr9:137410658-137410779 | 2 | -0.73 | 0.13 | 0.00988 | | |
| chr6:110903077-110903408 | 2 | -0.92 | 0.16 | 0.00994 | | |
| chr17:42687625-42687748 | 2 | -0.62 | 0.11 | 0.0101 | *CNTP1* | exon_8 |
| chr1:180188266-180188373 | 2 | -0.51 | 0.09 | 0.0101 | | |
| chr16:85444974-85445045 | 2 | 0.54 | 0.09 | 0.0102 | | |
| chr11:65867920-65868041 | 2 | -0.73 | 0.13 | 0.0104 | *EFEMP2* | exon_10 |
| chr19:16719935-16719938 | 2 | -0.28 | 0.05 | 0.0106 | *NWD1* | 5UTR |
| chr5:34756491-34756784 | 2 | -0.52 | 0.09 | 0.0106 | | |
| chr1:204446545-204446850 | 2 | -0.39 | 0.07 | 0.0106 | | |
| chr11:65593038-65593149 | 2 | -0.14 | 0.02 | 0.0107 | *KCNK7* | 3UTR |
| chr14:105476299-105476706 | 2 | -0.43 | 0.07 | 0.0107 | *CRIP2* | TSS1500 |
| chr6:31766112-31766137 | 4 | 0.32 | 0.06 | 0.0109 | *SAPCD1-AS1* | TSS1500 |
| chr22:46073915-46074062 | 2 | -0.015 | 0.00 | 0.0111 | | |
| chr11:60785282-60785312 | 2 | 0.079 | 0.01 | 0.0111 | *MS4A10* | TSS200 |
| chr16:2078706-2079360 | 6 | 0.2 | 0.04 | 0.0112 | *TSC2* | exon_27 |
| chr17:81870006-81870480 | 2 | -0.35 | 0.06 | 0.0114 | | |
| chr6:143961894-143962532 | 4 | -0.18 | 0.03 | 0.0114 | *PLAGL1* | TSS1500 |
| chr6:34277491-34277564 | 2 | -0.6 | 0.11 | 0.0115 | | |
| chr7:56093044-56093073 | 2 | -0.21 | 0.04 | 0.0116 | *PHKG1* | TSS200 |
| chr4:85930258-85930272 | 2 | -0.29 | 0.05 | 0.0116 | *ARHGAP24* | 5UTR |
| chr2:237056461-237056707 | 2 | -0.81 | 0.14 | 0.0118 | *LOC93463* | TSS1500 |
| chr1:26225291-26225611 | 2 | -1.1 | 0.19 | 0.012 | *ZPLD2P* | TSS200 |
| chr19:45234225-45234227 | 2 | -0.74 | 0.13 | 0.012 | *EXOC3L2* | exon_3 |
| chr10:5526387-5526457 | 2 | -0.55 | 0.10 | 0.0121 | *CALML3* | 3UTR |
| chr6:31156812-31157201 | 2 | -0.79 | 0.14 | 0.0122 | *CCHCR1* | exon_3 |
| chr11:35138764-35138853 | 3 | -1.3 | 0.22 | 0.0122 | *CD44* | TSS1500 |
| chr13:40188315-40188332 | 2 | 0.25 | 0.04 | 0.0122 | | |
| chr7:157151156-157151183 | 2 | 0.89 | 0.16 | 0.0123 | | |
| chr20:39645263-39645483 | 2 | 0.7 | 0.12 | 0.0125 | | |
| chr1:16157163-16157199 | 2 | -0.65 | 0.11 | 0.0127 | *EPHA2* | TSS1500 |
| chr10:619054-619364 | 3 | 0.84 | 0.15 | 0.0127 | | |
| chr3:52495069-52495377 | 5 | -0.51 | 0.09 | 0.0128 | *STAB1* | TSS200 |
| chr17:17757247-17757746 | 2 | -0.36 | 0.06 | 0.0128 | | |
| chr11:131408309-131408746 | 2 | -0.2 | 0.04 | 0.0129 | | |
| chr17:1934318-1934554 | 5 | -0.97 | 0.17 | 0.0135 | | |
| chr4:7287780-7288131 | 3 | -0.34 | 0.06 | 0.0137 | | |
| chr13:30905866-30906088 | 4 | 0.088 | 0.02 | 0.0141 | *MEDAG* | TSS1500 |
| chr7:144842525-144842779 | 2 | -0.28 | 0.05 | 0.0146 | | |
| chr12:105676692-105676749 | 2 | -0.44 | 0.08 | 0.0148 | | |
| chr10:5659174-5659468 | 2 | -1 | 0.18 | 0.0153 | | |

| | | | | | | |
|---|---|---|---|---|---|---|
| chr1:17123141-17123570 | 2 | 0.12 | 0.02 | 0.0153 | | |
| chr15:60763649-60763715 | 2 | -0.29 | 0.05 | 0.0155 | | |
| chr1:3663309-3663414 | 2 | -0.2 | 0.04 | 0.0156 | | |
| chr3:169134002-169134013 | 2 | -0.54 | 0.10 | 0.0158 | | |
| chr10:128959353-128959493 | 3 | -1.2 | 0.22 | 0.0159 | | |
| chr16:30948271-30948341 | 2 | -0.64 | 0.11 | 0.0159 | *FBXL19* | 3UTR |
| chr4:7460741-7461411 | 5 | -0.26 | 0.05 | 0.016 | | |
| chr21:46220237-46220296 | 2 | -0.67 | 0.12 | 0.0163 | | |
| chr6:33433759-33433765 | 2 | -0.52 | 0.09 | 0.0163 | | |
| chr17:18978697-18978826 | 2 | -0.35 | 0.06 | 0.0166 | *FAM83G* | exon_5 |
| chr10:79094180-79094228 | 2 | -0.44 | 0.08 | 0.0168 | | |
| chr6:32168026-32168275 | 2 | -0.55 | 0.10 | 0.0171 | *AGPAT1* | 3UTR |
| chr11:61480457-61480745 | 4 | 0.35 | 0.06 | 0.0172 | *PPP1R32* | TSS1500 |
| chr12:47758421-47758645 | 5 | -1.1 | 0.20 | 0.0172 | *RAPGEF3* | TSS200 |
| chr14:91870265-91870380 | 3 | -0.96 | 0.17 | 0.0174 | *FBLN5* | exon_12 |
| chr6:44219372-44219395 | 2 | -2.8 | 0.50 | 0.0177 | *SLC29A1* | TSS1500 |
| chr22:39466987-39467125 | 2 | 0.59 | 0.11 | 0.0181 | | |
| chr1:164775063-164775073 | 2 | 0.54 | 0.10 | 0.0184 | *PBX1-AS1* | TSS1500 |
| chr6:32843056-32843404 | 2 | -1.6 | 0.28 | 0.0184 | *PSMB8-AS1* | TSS1500 |
| chr20:56676133-56676153 | 2 | -0.72 | 0.13 | 0.0186 | | |
| chr20:32182116-32182217 | 2 | -0.66 | 0.12 | 0.0187 | | |
| chr11:67440103-67440313 | 2 | 1.1 | 0.19 | 0.0187 | | |
| chr12:116558968-116558985 | 2 | -0.23 | 0.04 | 0.0188 | *MAP1LC3B2* | TSS1500 |
| chr1:235867796-235868127 | 2 | -0.33 | 0.06 | 0.0189 | *LYST* | TSS1500 |
| chr1:53328316-53328539 | 3 | -0.68 | 0.12 | 0.0189 | *LRP8* | TSS1500 |
| chr14:104883118-104883439 | 5 | -0.57 | 0.10 | 0.0189 | *CEP170B* | exon_7 |
| chr16:85642920-85643255 | 2 | 0.23 | 0.04 | 0.0193 | | |
| chr15:40258345-40258827 | 2 | -0.47 | 0.08 | 0.0198 | | |
| chr17:79940167-79940363 | 2 | -0.4 | 0.07 | 0.0198 | *TBC1D16* | 3UTR |
| chr12:55931131-55931231 | 2 | -0.39 | 0.07 | 0.0199 | *DGKA* | TSS1500 |
| chr16:678470-678516 | 2 | -0.55 | 0.10 | 0.02 | | |
| chr1:20185915-20185970 | 2 | -0.59 | 0.11 | 0.02 | *UBXN10* | TSS200 |
| chr1:13582666-13583027 | 2 | -0.4 | 0.07 | 0.0201 | *PDPN* | TSS1500 |
| chr8:546327-546440 | 2 | -0.66 | 0.12 | 0.0202 | *TDRP* | TSS1500 |
| chr10:103759664-103759838 | 2 | -0.85 | 0.15 | 0.0203 | | |
| chr1:83860779-83860941 | 3 | -0.78 | 0.14 | 0.0205 | *.OC10192756(* | TSS1500 |
| chr8:141429180-141429389 | 2 | -0.96 | 0.17 | 0.0205 | | |
| chr9:137189500-137189836 | 2 | -1 | 0.18 | 0.0207 | *APC2* | TSS1500 |
| chr11:7512872-7513098 | 2 | -0.26 | 0.05 | 0.0208 | *PPFIBP2* | TSS1500 |
| chr15:62165499-62165684 | 2 | -0.52 | 0.09 | 0.0209 | *C2CD4B* | TSS1500 |
| chr11:35116226-35116537 | 2 | -0.1 | 0.02 | 0.0215 | | |
| chr17:30380291-30380443 | 2 | -0.58 | 0.10 | 0.0218 | *CPD* | TSS200 |
| chr17:82419423-82419771 | 2 | -0.25 | 0.04 | 0.0219 | *HEXD* | 5UTR |
| chr2:25204239-25204245 | 2 | 0.3 | 0.05 | 0.0219 | *LINC01381* | TSS200 |
| chr1:21264712-21264840 | 2 | -0.6 | 0.11 | 0.022 | | |
| chr16:29859657-29860085 | 3 | 0.87 | 0.15 | 0.0223 | | |
| chr12:124847513-124847637 | 2 | -0.46 | 0.08 | 0.0223 | | |
| chr7:108003804-108004125 | 2 | -0.28 | 0.05 | 0.0224 | *LAMB1* | TSS1500 |

| | | | | | | |
|---|---|---|---|---|---|---|
| chr1:51404632-51404907 | 2 | -0.35 | 0.06 | 0.0224 | | |
| chr6:683859-684090 | 2 | -0.23 | 0.04 | 0.0224 | | |
| chr14:68599307-68599416 | 2 | 0.42 | 0.08 | 0.0228 | | |
| chr20:25553662-25554149 | 2 | -0.58 | 0.10 | 0.0235 | | |
| chr16:2044666-2044877 | 3 | -0.55 | 0.10 | 0.0237 | *NTHL1* | exon_3 |
| chr16:81220405-81220604 | 5 | 0.067 | 0.01 | 0.0239 | *PKD1L2* | TSS200 |
| chr2:47002904-47003211 | 3 | -0.17 | 0.03 | 0.0241 | | |
| chr4:7940125-7940293 | 3 | -0.46 | 0.08 | 0.0243 | *AFAP1* | TSS1500 |
| chr17:3646033-3646200 | 2 | -0.71 | 0.13 | 0.0245 | | |
| chr19:50414848-50414919 | 2 | 0.59 | 0.10 | 0.0246 | *POLD1* | exon_20 |
| chr2:136645467-136645597 | 2 | -0.51 | 0.09 | 0.0249 | | |
| chr5:116502777-116502824 | 2 | -1.1 | 0.20 | 0.025 | | |
| chr7:2043470-2043546 | 2 | -0.18 | 0.03 | 0.0256 | | |
| chr19:38655537-38655807 | 2 | 0.17 | 0.03 | 0.0257 | | |
| chr16:2715842-2716246 | 3 | -0.44 | 0.08 | 0.0258 | *PRSS27* | exon_3 |
| chr1:43206561-43206951 | 2 | -0.39 | 0.07 | 0.0261 | | |
| chr17:17785528-17785763 | 3 | -0.67 | 0.12 | 0.0264 | | |
| chr6:32952790-32953103 | 5 | 0.16 | 0.03 | 0.0266 | *HLA-DMA* | exon_1 |
| chr3:124780806-124780982 | 2 | 0.91 | 0.16 | 0.0273 | | |
| chr18:61333265-61333269 | 2 | -0.56 | 0.10 | 0.0274 | *CDH20* | TSS200 |
| chr11:2272030-2272075 | 2 | -1.3 | 0.23 | 0.0275 | *ASCL2* | TSS1500 |
| chr9:127906146-127906163 | 2 | 0.65 | 0.12 | 0.0276 | *ST6GALC6* | TSS1500 |
| chr16:3099263-3099396 | 3 | 0.052 | 0.01 | 0.0277 | *ZSCAN10* | 5UTR |
| chr11:66547768-66547827 | 2 | -0.69 | 0.12 | 0.0277 | | |
| chr11:66280725-66280765 | 2 | -1.5 | 0.27 | 0.0278 | | |
| chr7:150385247-150385377 | 2 | 0.28 | 0.05 | 0.028 | | |
| chr20:24144502-24144523 | 2 | 0.47 | 0.08 | 0.0281 | | |
| chr20:41087094-41087218 | 2 | -0.37 | 0.07 | 0.0282 | | |
| chr4:165379536-165379924 | 2 | -0.88 | 0.16 | 0.0284 | | |
| chr2:156330717-156330829 | 2 | -0.23 | 0.04 | 0.0285 | *NR4A2* | 5UTR |
| chr3:50322547-50322666 | 2 | 0.28 | 0.05 | 0.0286 | *HYAL2* | 5UTR |
| chr8:144550989-144551313 | 2 | -0.64 | 0.12 | 0.0288 | | |
| chr18:46412551-46412680 | 2 | -0.49 | 0.09 | 0.0289 | | |
| chr14:104799572-104799829 | 2 | -0.86 | 0.16 | 0.0289 | *ZBTB42* | TSS1500 |
| chr16:28984949-28985037 | 3 | -0.037 | 0.01 | 0.0297 | *LAT* | TSS200 |
| chr12:53102945-53103058 | 2 | -0.45 | 0.08 | 0.0297 | *SOAT2* | TSS1500 |
| chr12:116319143-116319240 | 2 | -0.25 | 0.04 | 0.0299 | | |
| chr12:123268304-123268406 | 3 | -0.25 | 0.04 | 0.0301 | *CDK2AP1* | 5UTR |
| chr5:3633997-3634317 | 2 | -1.1 | 0.20 | 0.0304 | | |
| chr11:20112974-20113138 | 2 | -1 | 0.19 | 0.0305 | | |
| chr12:117025488-117025752 | 2 | -1.2 | 0.22 | 0.031 | | |
| chr20:25310864-25311476 | 3 | -0.27 | 0.05 | 0.032 | | |
| chr4:37567057-37567450 | 2 | -0.53 | 0.10 | 0.0322 | | |
| chr12:49961057-49961179 | 2 | -0.3 | 0.05 | 0.0323 | *AQP5* | TSS1500 |
| chr6:10420836-10421227 | 3 | 0.38 | 0.07 | 0.0323 | *TFAP2A* | TSS1500 |
| chr19:16418053-16418115 | 2 | -0.68 | 0.12 | 0.0324 | *EPS15L1* | exon_11 |
| chr13:112702689-112703140 | 4 | -0.31 | 0.06 | 0.0324 | | |
| chr10:34792119-34792156 | 2 | 0.57 | 0.10 | 0.0329 | | |

| | | | | | | |
|---|---|---|---|---|---|---|
| chr1:207248838-207248911 | 2 | -0.45 | 0.08 | 0.0332 | | |
| chr19:1355919-1356154 | 4 | 0.39 | 0.07 | 0.0333 | *PWWP3A* | TSS1500 |
| chr19:5029927-5030047 | 2 | -1 | 0.18 | 0.0334 | | |
| chr3:114383768-114384220 | 4 | 0.4 | 0.07 | 0.0336 | *ZBTB20* | TSS1500 |
| chr6:157275390-157275536 | 3 | 0.16 | 0.03 | 0.0338 | | |
| chr5:134115783-134115934 | 2 | -0.63 | 0.11 | 0.0339 | *TCF7* | 5UTR |
| chr12:116520780-116520935 | 2 | -0.26 | 0.05 | 0.0345 | | |
| chr7:151062546-151062576 | 2 | -0.87 | 0.16 | 0.0349 | *SLC4A2* | TSS200 |
| chr10:70759956-70760117 | 2 | -0.63 | 0.11 | 0.0349 | | |
| chr19:48747304-48747431 | 2 | 0.59 | 0.11 | 0.035 | *IZUMO1* | TSS1500 |
| chr2:226795735-226796497 | 3 | -0.65 | 0.12 | 0.0352 | *IRS1* | exon_1 |
| chr21:34865955-34865975 | 2 | 0.74 | 0.13 | 0.0353 | | |
| chr8:17002324-17002415 | 3 | 0.41 | 0.07 | 0.0353 | *FGF20* | 5UTR |
| chr11:65204756-65204932 | 2 | -0.69 | 0.13 | 0.0353 | *CAPN1* | exon_11 |
| chr2:218382129-218382246 | 2 | -1.5 | 0.28 | 0.0353 | *SLC11A1* | TSS200 |
| chr1:2115540-2115662 | 2 | -1 | 0.19 | 0.0355 | | |
| chr8:124655098-124655544 | 3 | 0.1 | 0.02 | 0.0358 | | |
| chr3:195545919-195546236 | 2 | -0.2 | 0.04 | 0.0362 | | |
| chr10:124016262-124016640 | 2 | -0.93 | 0.17 | 0.0366 | | |
| chr10:71719757-71719939 | 2 | -1 | 0.19 | 0.0371 | *C10orf105* | 5UTR |
| chr17:40060143-40060259 | 2 | -0.48 | 0.09 | 0.0372 | | |
| chr5:92082516-92082531 | 2 | -0.23 | 0.04 | 0.0374 | | |
| chr3:127037022-127037303 | 2 | -0.41 | 0.08 | 0.0377 | *PLX1* | 3UTR |
| chr4:787441-787484 | 2 | -0.88 | 0.16 | 0.038 | | |
| chr14:23376684-23376787 | 2 | -0.4 | 0.07 | 0.0384 | *CMTM5* | TSS1500 |
| chr15:43516242-43516608 | 2 | -0.57 | 0.10 | 0.0386 | *MAP1A* | TSS1500 |
| chr14:22560719-22561595 | 3 | -0.28 | 0.05 | 0.0389 | | |
| chr3:71728813-71729241 | 2 | -0.58 | 0.11 | 0.039 | *EIF4E3* | TSS200 |
| chr16:85062827-85063207 | 3 | -0.56 | 0.10 | 0.0391 | *KIAA0513* | TSS200 |
| chr15:101047533-101047678 | 2 | -0.64 | 0.12 | 0.0395 | | |
| chr9:85508795-85508994 | 2 | -0.41 | 0.08 | 0.0396 | | |
| chr16:2936884-2937034 | 2 | -0.24 | 0.04 | 0.0398 | | |
| chr7:98348877-98349418 | 4 | 0.25 | 0.05 | 0.0403 | | |
| chr17:76480883-76480896 | 2 | -0.13 | 0.02 | 0.0414 | | |
| chr7:127593826-127593975 | 2 | -0.73 | 0.13 | 0.0416 | *FSCN3* | exon_1 |
| chr6:109465889-109466036 | 2 | -0.33 | 0.06 | 0.0418 | *MICAL1* | exon_7 |
| chr17:62965442-62965532 | 2 | -0.27 | 0.05 | 0.0423 | *TANC2* | TSS1500 |
| chr12:114669735-114670120 | 3 | -0.13 | 0.02 | 0.0423 | | |
| chr17:80105809-80105816 | 2 | -0.19 | 0.04 | 0.0428 | *GAA* | exon_3 |
| chr1:1282014-1282131 | 5 | -0.12 | 0.02 | 0.0429 | | |
| chr11:68042954-68043275 | 2 | -0.86 | 0.16 | 0.0429 | *TCIRG1* | 5UTR |
| chr12:53051331-53051415 | 2 | -0.42 | 0.08 | 0.043 | | |
| chr6:31877895-31877944 | 2 | -0.72 | 0.13 | 0.0431 | *SLC44A4* | TSS1500 |
| chr12:109814811-109815026 | 6 | 0.3 | 0.06 | 0.0433 | *TRPV4* | TSS200 |
| chr18:58629312-58629375 | 2 | -0.28 | 0.05 | 0.0436 | *ALPK2* | TSS1500 |
| chr3:149936752-149937045 | 2 | 1.2 | 0.22 | 0.0441 | | |
| chr16:88741299-88741463 | 2 | -0.32 | 0.06 | 0.0444 | *LOC339059* | TSS1500 |
| chr11:64138447-64138632 | 4 | 0.0083 | 0.00 | 0.0448 | | |

| | | | | | | |
|---|---|---|---|---|---|---|
| chr16:30429355-30429455 | 2 | 0.84 | 0.15 | 0.0451 | | |
| chr19:19439934-19439990 | 2 | -1.4 | 0.25 | 0.0453 | | |
| chr10:77147398-77147625 | 2 | -1.5 | 0.26 | 0.0462 | | |
| chr5:168654315-168654456 | 2 | -0.84 | 0.15 | 0.0462 | | |
| chr3:134367821-134368308 | 2 | -0.85 | 0.16 | 0.0462 | *MIR6827* | TSS1500 |
| chr2:38945305-38945563 | 2 | -0.47 | 0.09 | 0.0468 | | |
| chr17:34254201-34254447 | 2 | -0.49 | 0.09 | 0.0468 | *CCL2* | TSS1500 |
| chr7:105298884-105299372 | 3 | -0.29 | 0.05 | 0.047 | *SRPK2* | TSS1500 |
| chr12:6839950-6840096 | 3 | -1.2 | 0.22 | 0.047 | *GNB3* | TSS1500 |
| chr19:5932403-5932465 | 2 | -0.4 | 0.07 | 0.0473 | *RANBP3* | exon_6 |
| chr17:81459133-81459327 | 2 | -0.4 | 0.07 | 0.0476 | *BAHCC1* | exon_22 |
| chr14:104694459-104694851 | 2 | -0.33 | 0.06 | 0.0477 | | |
| chr1:248818433-248818542 | 2 | -0.54 | 0.10 | 0.048 | | |
| chr1:1168885-1168986 | 3 | -1.9 | 0.35 | 0.0484 | *MIR429* | TSS200 |
| chr17:76716967-76717199 | 2 | -0.81 | 0.15 | 0.0488 | | |

**Supplementary Table 6**

Functional enrichment analysis for top CpGs identified in the EWAS of early preterm birth (ePTB) in boys (n=318 CpGs at p < 9.4x10-8). Analysis performed using the *KnowyourCG* R package with functional terms observed across multiple databases. Enrichment of a functional term based on FDR < 0.05.

| Source Database | Term name | Term size | Query size | Overlap | Adjusted P |
|---|---|---|---|---|---|
| GO:BP | neuron development | 1167 | 125 | 27 | 1.5E-06 |
| GO:BP | neuron projection development | 1010 | 125 | 25 | 1.8E-06 |
| GO:BP | cell morphogenesis involved in neuron differentiation | 584 | 125 | 18 | 1.8E-05 |
| GO:BP | neuron differentiation | 1444 | 125 | 27 | 1.3E-04 |
| GO:BP | cell development | 2842 | 125 | 40 | 1.4E-04 |
| GO:BP | plasma membrane bounded cell projection organization | 1582 | 125 | 28 | 2.3E-04 |
| GO:CC | cytoplasm | 12601 | 132 | 102 | 2.3E-04 |
| GO:BP | generation of neurons | 1523 | 125 | 27 | 3.8E-04 |
| GO:BP | cell projection organization | 1628 | 125 | 28 | 4.1E-04 |
| GO:BP | neuron projection morphogenesis | 648 | 125 | 17 | 4.9E-04 |
| GO:BP | regulation of signaling | 3473 | 125 | 44 | 5.0E-04 |
| GO:BP | regulation of cell communication | 3480 | 125 | 44 | 5.3E-04 |
| GO:BP | plasma membrane bounded cell projection morphogenesis | 664 | 125 | 17 | 6.8E-04 |
| GO:BP | cell morphogenesis | 998 | 125 | 21 | 6.9E-04 |
| GO:BP | cell projection morphogenesis | 669 | 125 | 17 | 7.6E-04 |
| MIRNA | hsa-mir-543 | 1569 | 131 | 31 | 1.0E-03 |
| MIRNA | hsa-mir-338-5p | 1349 | 131 | 28 | 1.5E-03 |
| GO:CC | cytosol | 5607 | 132 | 57 | 1.7E-03 |
| GO:BP | neurogenesis | 1769 | 125 | 28 | 2.2E-03 |
| MIRNA | hsa-mir-3928 | 1145 | 131 | 25 | 2.4E-03 |
| GO:CC | synapse | 1608 | 132 | 25 | 2.6E-03 |
| GO:BP | nervous system development | 2597 | 125 | 35 | 3.3E-03 |
| GO:BP | axon development | 513 | 125 | 14 | 3.7E-03 |
| GO:MF | protein binding | 15082 | 131 | 118 | 4.0E-03 |
| GO:BP | system development | 4023 | 125 | 46 | 4.6E-03 |
| GO:BP | axonogenesis | 452 | 125 | 13 | 4.9E-03 |
| GO:BP | cell communication | 6598 | 125 | 64 | 5.2E-03 |

| | | | | | |
|---|---|---|---|---|---|
| GO:BP | regulation of signal transduction | 3031 | 125 | 38 | 5.9E-03 |
| MIRNA | hsa-mir-4760-3p | 1288 | 131 | 26 | 6.1E-03 |
| TF | Factor: ZXDL; motif: GSGSCNNGGGMRGCNCCGGGS; match class: 1 | 6630 | 140 | 73 | 6.2E-03 |
| GO:BP | regulation of plasma membrane bounded cell projection organization | 623 | 125 | 15 | 7.3E-03 |
| GO:BP | signal transduction | 6063 | 125 | 60 | 7.5E-03 |
| MIRNA | hsa-mir-4261 | 1223 | 131 | 25 | 7.7E-03 |
| GO:BP | regulation of response to stimulus | 3993 | 125 | 45 | 9.4E-03 |
| GO:BP | signaling | 6558 | 125 | 63 | 9.7E-03 |
| GO:BP | regulation of cell projection organization | 639 | 125 | 15 | 9.8E-03 |
| MIRNA | hsa-mir-3140-3p | 1408 | 131 | 27 | 1.0E-02 |
| GO:BP | multicellular organism development | 4690 | 125 | 50 | 1.1E-02 |
| TF | Factor: MAZ; motif: NKGGGAGGGGRGGR | 7258 | 140 | 77 | 1.1E-02 |
| MIRNA | hsa-mir-4795-3p | 1340 | 131 | 26 | 1.2E-02 |
| GO:BP | regulation of neuron projection development | 426 | 125 | 12 | 1.5E-02 |
| GO:CC | cell junction | 2370 | 132 | 30 | 1.6E-02 |
| GO:BP | regulation of cellular component organization | 2427 | 125 | 32 | 1.6E-02 |
| GO:BP | cellular response to stimulus | 7425 | 125 | 68 | 1.6E-02 |
| GO:CC | cytoskeleton | 2544 | 132 | 31 | 2.3E-02 |
| GO:BP | intracellular signal transduction | 2963 | 125 | 36 | 2.4E-02 |
| TF | Factor: E2F3; motif: NNRGMKGGAR | 12627 | 140 | 112 | 2.4E-02 |
| MIRNA | hsa-mir-1294 | 986 | 131 | 21 | 2.5E-02 |
| WP | Factors and pathways affecting insulin like growth factor IGF1 Akt signaling | 36 | 68 | 4 | 2.8E-02 |
| GO:BP | cell differentiation | 4443 | 125 | 47 | 3.0E-02 |
| GO:BP | cellular developmental process | 4444 | 125 | 47 | 3.0E-02 |
| TF | Factor: CKROX; motif: SCCCTCCCC; match class: 1 | 2689 | 140 | 38 | 3.3E-02 |
| GO:CC | neuron projection | 1321 | 132 | 20 | 3.4E-02 |
| GO:CC | glutamatergic synapse | 548 | 132 | 12 | 3.5E-02 |
| GO:MF | cytoskeletal protein binding | 996 | 131 | 18 | 4.2E-02 |
| GO:BP | developmental process | 6535 | 125 | 61 | 4.3E-02 |
| GO:BP | anatomical structure development | 5948 | 125 | 57 | 4.6E-02 |

GO:BP = Gene ontology biological process; GO:CC = Gene ontology cellular component; GO:MF = Gene ontology molecular function; TF = transcription factor; MIRNA = micro RNAs; WP = Curated Wiki pathways.

**Supplementary Table 7**

Functional enrichment analysis for top CpGs identified in the EWAS of early preterm birth (ePTB) in girls (n=542 CpGs at $p < 9.4 \times 10^{-8}$). Analysis performed using the *KnowyourCG* R package. Enrichment deemed at FDR < 0.05.

| Source Database | Term name | Term size | Query size | Overlap | Adjusted P |
|---|---|---|---|---|---|
| GO:CC | cytoplasm | 12601 | 263 | 204 | 4.6E-10 |
| TF | Factor: ZXDL; motif: GSGSCNNGGGMRGCNCCGGGS; match class: 1 | 6630 | 274 | 145 | 2.0E-08 |
| TF | Factor: AP-2alpha; motif: NGCCYSNNGSN; match class: 1 | 4535 | 274 | 112 | 2.2E-08 |
| TF | Factor: ZXDL; motif: GSGSCNNGGGMRGCNCCGGGS | 10669 | 274 | 197 | 3.6E-07 |
| TF | Factor: AP-2alpha; motif: NGCCYSNNGSN | 9244 | 274 | 177 | 1.1E-06 |
| TF | Factor: RERE; motif: CNGCNSCNNGSRCRGSGSS; match class: 1 | 5959 | 274 | 129 | 2.3E-06 |
| TF | Factor: MAZ; motif: GGGMGGGGS | 9467 | 274 | 178 | 5.0E-06 |
| TF | Factor: ZNF138; motif: GCAGCRSCNSGSNCMGSGCS; match class: 1 | 5518 | 274 | 121 | 5.8E-06 |
| TF | Factor: GKLF; motif: NNNRGGNGNGGSN; match class: 1 | 10807 | 274 | 195 | 7.2E-06 |
| GO:CC | cytosol | 5607 | 263 | 107 | 1.0E-05 |
| TF | Factor: HA95; motif: CCSNSSCCNSCNCCWSCCNS; match class: 1 | 8444 | 274 | 163 | 1.1E-05 |
| TF | Factor: AP-2beta; motif: GCNNNGGSCNGVGGGN; match class: 1 | 3086 | 274 | 80 | 1.2E-05 |
| TF | Factor: TIEG1; motif: NCCCNSNCCCCGCCCCC | 12566 | 274 | 216 | 1.2E-05 |
| MIRNA | hsa-mir-326 | 1354 | 249 | 47 | 4.0E-05 |
| TF | Factor: PHB; motif: NCCCAGCCCCY; match class: 1 | 4876 | 274 | 108 | 5.1E-05 |
| TF | Factor: ZBED4; motif: CCCGCYCCGC; match class: 1 | 3784 | 274 | 90 | 5.9E-05 |
| TF | Factor: KLF15; motif: NCCMCGCCCMCN | 11280 | 274 | 198 | 6.9E-05 |
| TF | Factor: ZNF219; motif: SNNCAGCACCNNGGNCAGCGSC; match class: 1 | 6609 | 274 | 134 | 7.5E-05 |
| TF | Factor: Egr-2; motif: GNGRRNGWGKGGGNGGRG | 6616 | 274 | 134 | 8.1E-05 |
| TF | Factor: ETF; motif: GVGGMGG; match class: 1 | 7033 | 274 | 140 | 8.8E-05 |
| TF | Factor: TIEG1; motif: NCCCNSNCCCCGCCCCC; match class: 1 | 8323 | 274 | 158 | 1.1E-04 |
| TF | Factor: ZNF232; motif: NCAGCASCNNGGNCAGCGCC | 6249 | 274 | 128 | 1.2E-04 |
| TF | Factor: MOVO-B; motif: GNGGGGG; match class: 1 | 5667 | 274 | 119 | 1.3E-04 |
| MIRNA | hsa-mir-330-5p | 1340 | 249 | 45 | 2.1E-04 |

| | | | | | |
|---|---|---|---|---|---|
| TF | Factor: Zbtb37; motif: NYACCGCRNTCACCGCR; match class: 1 | 1753 | 274 | 52 | 2.1E-04 |
| TF | Factor: ZNF232; motif: NCAGCASCNNGGNCAGCGCC; match class: 1 | 2122 | 274 | 59 | 2.3E-04 |
| TF | Factor: MOVO-B; motif: GNGGGGG | 10403 | 274 | 185 | 2.4E-04 |
| TF | Factor: WT1; motif: CGCCCCCNCN; match class: 1 | 5335 | 274 | 113 | 2.4E-04 |
| TF | Factor: BTEB3; motif: CCNNSCCNSCCCCKCCCCC | 11717 | 274 | 201 | 4.1E-04 |
| MIRNA | hsa-mir-138 | 895 | 249 | 34 | 5.2E-04 |
| TF | Factor: ZNF253; motif: SNGNSCGNGGNGCKGNN; match class: 1 | 6348 | 274 | 127 | 6.1E-04 |
| TF | Factor: PATZ; motif: GGGGNGGGGGMKGGRRNGGNRN | 8545 | 274 | 158 | 8.7E-04 |
| TF | Factor: BTEB1; motif: GGGGGCGGGGCNGSGGGNGS; match class: 1 | 6055 | 274 | 122 | 9.2E-04 |
| TF | Factor: KLF14; motif: NRCCACRCCCMCN | 6962 | 274 | 135 | 1.2E-03 |
| TF | Factor: ZNF253; motif: SNGNSCGNGGNGCKGNN | 10979 | 274 | 190 | 1.2E-03 |
| TF | Factor: E2F-3; motif: GGCGGGN; match class: 1 | 9478 | 274 | 170 | 1.4E-03 |
| TF | Factor: TF3C-beta; motif: CCNGGAGGGCTTCCTGGAGGAG | 13779 | 274 | 224 | 1.6E-03 |
| HPA | Hippocampus | 6820 | 178 | 138 | 1.6E-03 |
| TF | Factor: BTEB3; motif: CCNNSCCNSCCCCKCCCCC; match class: 1 | 7629 | 274 | 144 | 1.8E-03 |
| TF | Factor: E2F-3:FOXO6; motif: NAATGACACGCGCCCMC; match class: 1 | 7849 | 274 | 147 | 1.9E-03 |
| TF | Factor: WT1; motif: GNGGGGGCGGGG | 8428 | 274 | 155 | 2.0E-03 |
| TF | Factor: ZNF614; motif: NCYCWGCCYYNNN; match class: 1 | 4204 | 274 | 92 | 2.1E-03 |
| TF | Factor: MAZ; motif: GGGMGGGGS; match class: 1 | 4394 | 274 | 95 | 2.1E-03 |
| TF | Factor: KLF15; motif: RCCMCRCCCMCN | 12970 | 274 | 214 | 2.2E-03 |
| TF | Factor: Kaiso; motif: GCMGGGRGCRGS; match class: 1 | 9168 | 274 | 165 | 2.3E-03 |
| TF | Factor: MAZ; motif: GGGMGGGGSSGGGGGGGGGGG; match class: 1 | 14348 | 274 | 230 | 2.5E-03 |
| TF | Factor: Miz-1; motif: NNRGGWGGGGGAGGGGMRR; match class: 1 | 3981 | 274 | 88 | 2.7E-03 |
| TF | Factor: WT1; motif: CGCCCCCNCN | 9882 | 274 | 174 | 3.4E-03 |
| TF | Factor: ZNF670; motif: SNGGGCRGRG; match class: 1 | 8277 | 274 | 152 | 3.5E-03 |
| TF | Factor: DB1; motif: GGRRRRGRRGGAGGGGGNGRRR; match class: 1 | 2696 | 274 | 66 | 3.6E-03 |
| TF | Factor: RERE; motif: CNGCNSCNNGSRCRGSGSS | 10357 | 274 | 180 | 4.1E-03 |
| HPA | Cerebral cortex; neuronal cells[>=Low] | 6698 | 178 | 135 | 4.2E-03 |
| TF | Factor: GKLF; motif: NNNRGGNGNGGSN | 14949 | 274 | 236 | 4.5E-03 |
| TF | Factor: ZNF37A; motif: CCYYGGCTCCNTSCCMN; match class: 1 | 5630 | 274 | 113 | 4.7E-03 |
| MIRNA | hsa-mir-4504 | 816 | 249 | 30 | 4.9E-03 |

| | | | | | |
|---|---|---|---|---|---|
| TF | Factor: E2F-3; motif: GGCGGGN | 13411 | 274 | 218 | 5.3E-03 |
| MIRNA | hsa-mir-1207-3p | 1558 | 249 | 46 | 5.4E-03 |
| TF | Factor: ZNF460; motif: NNACNCCCCCCNN | 5915 | 274 | 117 | 5.6E-03 |
| TF | Factor: CNOT3; motif: GGCCGCGSSS | 3626 | 274 | 81 | 6.1E-03 |
| TF | Factor: AP2; motif: GCCYGSGGSN | 10181 | 274 | 177 | 6.1E-03 |
| TF | Factor: AP-2beta; motif: NSCCNNNGGSN; match class: 1 | 8133 | 274 | 149 | 6.3E-03 |
| TF | Factor: BTEB2; motif: RGGGNGKGGN | 8639 | 274 | 156 | 6.5E-03 |
| TF | Factor: ZNF148; motif: NNNNNNCCNNCCCCTCCCCCACCCN | 7087 | 274 | 134 | 6.7E-03 |
| TF | Factor: Egr-1; motif: CCGCCCMCG | 5144 | 274 | 105 | 6.7E-03 |
| MIRNA | hsa-mir-4779 | 1571 | 249 | 46 | 6.7E-03 |
| HPA | Hippocampus; neuronal cells[>=Low] | 6215 | 178 | 127 | 7.6E-03 |
| TF | Factor: KLF16; motif: GMCACGCCCCC | 3413 | 274 | 77 | 8.4E-03 |
| MIRNA | hsa-mir-4436b-3p | 1537 | 249 | 45 | 8.6E-03 |
| TF | Factor: ZNF93; motif: GGYAGCRGCAGCGGYG; match class: 1 | 1297 | 274 | 39 | 8.7E-03 |
| TF | Factor: DB1; motif: GGRRRRGRRGGAGGGGGNGRRR | 5898 | 274 | 116 | 8.8E-03 |
| TF | Factor: Sp2; motif: GGSNNGGGGGCGGGGCCNGNGS; match class: 1 | 2039 | 274 | 53 | 9.6E-03 |
| TF | Factor: MAZ; motif: GGGGGAGGGGGNGRGRRRGNRG | 9715 | 274 | 170 | 9.7E-03 |
| TF | Factor: ZNF93; motif: GGYAGCRGCAGCGGYG | 2548 | 274 | 62 | 1.0E-02 |
| GO:CC | nucleoplasm | 4334 | 263 | 79 | 1.0E-02 |
| TF | Factor: ZBED4; motif: CCCGCYCCGC | 7698 | 274 | 142 | 1.0E-02 |
| TF | Factor: KLF15; motif: RGGGMGGRGNNGGGGGNGG | 3317 | 274 | 75 | 1.1E-02 |
| TF | Factor: PHB; motif: NCCCAGCCCCY | 10410 | 274 | 179 | 1.1E-02 |
| TF | Factor: Churchill; motif: CGGGNN; match class: 1 | 10486 | 274 | 180 | 1.1E-02 |
| TF | Factor: MAZ; motif: GGGGGAGGGGGNGRGRRRGNRG; match class: 1 | 5660 | 274 | 112 | 1.1E-02 |
| MIRNA | hsa-mir-1294 | 986 | 249 | 33 | 1.2E-02 |
| MIRNA | hsa-mir-1225-3p | 897 | 249 | 31 | 1.2E-02 |
| TF | Factor: AP2; motif: GCCYGSGGSN; match class: 1 | 5141 | 274 | 104 | 1.2E-02 |
| TF | Factor: AP-2; motif: MKCCCSCNGGCG; match class: 1 | 6476 | 274 | 124 | 1.3E-02 |
| MIRNA | hsa-mir-320c | 1321 | 249 | 40 | 1.4E-02 |
| GO:CC | cell junction | 2370 | 263 | 50 | 1.4E-02 |
| MIRNA | hsa-mir-765 | 1617 | 249 | 46 | 1.4E-02 |

| | | | | | |
|---|---|---|---|---|---|
| MIRNA | hsa-mir-4316 | 952 | 249 | 32 | 1.5E-02 |
| TF | Factor: CKROX; motif: SCCCTCCCC | 7397 | 274 | 137 | 1.6E-02 |
| MIRNA | hsa-mir-320a | 1683 | 249 | 47 | 1.8E-02 |
| TF | Factor: EZI; motif: GGGGAGGGGRRGGRGRRGGRRR | 2141 | 274 | 54 | 1.8E-02 |
| MIRNA | hsa-mir-4660 | 1153 | 249 | 36 | 2.0E-02 |
| TF | Factor: BTEB1; motif: GGGGGCGGGGCNGSGGGNGS | 10190 | 274 | 175 | 2.1E-02 |
| TF | Factor: E2F-3:FOXI1; motif: NGACACCGCGCCCAC; match class: 1 | 7158 | 274 | 133 | 2.1E-02 |
| MIRNA | hsa-mir-4288 | 750 | 249 | 27 | 2.2E-02 |
| MIRNA | hsa-mir-637 | 1399 | 249 | 41 | 2.3E-02 |
| MIRNA | hsa-mir-320d | 1302 | 249 | 39 | 2.3E-02 |
| MIRNA | hsa-mir-4429 | 1303 | 249 | 39 | 2.3E-02 |
| MIRNA | hsa-mir-632 | 753 | 249 | 27 | 2.4E-02 |
| TF | Factor: Sp1; motif: NGGGGCGGGGN | 10814 | 274 | 183 | 2.4E-02 |
| TF | Factor: Pax-5; motif: GTYAYGCTTSRCTGVNYN | 5288 | 274 | 105 | 2.5E-02 |
| TF | Factor: KLF15; motif: NCCMCGCCCMCN; match class: 1 | 6085 | 274 | 117 | 2.5E-02 |
| TF | Factor: CP2; motif: NRRRCCGGCYNRARCCGGNN | 11130 | 274 | 187 | 2.6E-02 |
| MIRNA | hsa-mir-4766-5p | 1456 | 249 | 42 | 2.6E-02 |
| TF | Factor: CPBP; motif: GNNRGGGHGGGGNNGGGRN | 11055 | 274 | 186 | 2.6E-02 |
| MIRNA | hsa-mir-6782-5p | 59 | 249 | 7 | 2.6E-02 |
| TF | Factor: AP-2alpha; motif: NSCCNCRGGSN | 8604 | 274 | 153 | 2.7E-02 |
| TF | Factor: MAZ; motif: NKGGGAGGGGRGGR | 7258 | 274 | 134 | 2.7E-02 |
| MIRNA | hsa-mir-4677-3p | 1029 | 249 | 33 | 2.7E-02 |
| TF | Factor: Egr-1; motif: GCGTGGGCGT | 5897 | 274 | 114 | 2.8E-02 |
| TF | Factor: Sp1; motif: NGGGGGCGGGGCCNGGGGGGG | 8609 | 274 | 153 | 2.8E-02 |
| TF | Factor: KLF15; motif: RGGGMGGRGNNGGGGGNGG; match class: 1 | 1016 | 274 | 32 | 3.0E-02 |
| GO:BP | organelle organization | 3573 | 258 | 71 | 3.0E-02 |
| MIRNA | hsa-mir-499-5p | 722 | 249 | 26 | 3.1E-02 |
| TF | Factor: ZBP89; motif: CCCCKCCCCCNN | 7277 | 274 | 134 | 3.2E-02 |
| TF | Factor: ZNF383; motif: SSNGGGMGGNGSNGGS; match class: 1 | 4346 | 274 | 90 | 3.2E-02 |
| MIRNA | hsa-mir-1252 | 1471 | 249 | 42 | 3.3E-02 |
| GO:BP | cellular response to stimulus | 7425 | 258 | 124 | 3.4E-02 |

| | | | | | |
|---|---|---|---|---|---|
| TF | Factor: RNF96; motif: BCCCGCRGCC | 8564 | 274 | 152 | 3.5E-02 |
| MIRNA | hsa-mir-3918 | 1232 | 249 | 37 | 3.6E-02 |
| MIRNA | hsa-mir-4453 | 685 | 249 | 25 | 3.6E-02 |
| MIRNA | hsa-mir-4538 | 685 | 249 | 25 | 3.6E-02 |
| TF | Factor: AP-2alpha; motif: GCCNNNRGS | 5133 | 274 | 102 | 3.6E-02 |
| WP | KCNQ2 and KCNQ3 related epilepsy | 31 | 154 | 5 | 3.7E-02 |
| TF | Factor: ZNF138; motif: GCAGCRSCNSGSNCMGSGCS | 9668 | 274 | 167 | 3.7E-02 |
| MIRNA | hsa-mir-4456 | 910 | 249 | 30 | 4.0E-02 |
| TF | Factor: Sp1; motif: GGGGCGGGGC | 6344 | 274 | 120 | 4.0E-02 |
| TF | Factor: SP1; motif: GGGGYGGGGNS | 8082 | 274 | 145 | 4.0E-02 |
| TF | Factor: GKLF; motif: NNCCMCRCCCN; match class: 1 | 5016 | 274 | 100 | 4.1E-02 |
| TF | Factor: Churchill; motif: CGGGNN | 14432 | 274 | 227 | 4.1E-02 |
| TF | Factor: AP-2; motif: MKCCCSCNGGCG | 11119 | 274 | 186 | 4.1E-02 |
| HPA | Cerebral cortex; neuronal cells[>=Medium] | 4984 | 178 | 105 | 4.2E-02 |
| TF | Factor: KLF15; motif: RCCMCRCCCMCN; match class: 1 | 8164 | 274 | 146 | 4.3E-02 |
| TF | Factor: Sp2; motif: NYSGCCCCGCCCCCY | 7812 | 274 | 141 | 4.4E-02 |
| TF | Factor: WT1; motif: NGCGGGGGGGTSMMCYN | 5618 | 274 | 109 | 4.4E-02 |
| TF | Factor: Egr-1; motif: GCGCATGCG; match class: 1 | 10295 | 274 | 175 | 4.6E-02 |
| TF | Factor: TCF-1; motif: ACATCGRGRCGCTGW; match class: 1 | 13360 | 274 | 214 | 4.7E-02 |
| TF | Factor: AP-2beta; motif: NSCCNNNGGSN | 10078 | 274 | 172 | 4.8E-02 |
| TF | Factor: E2F-1; motif: NGGGCGGGARV; match class: 1 | 10913 | 274 | 183 | 4.9E-02 |
| TF | Factor: PATZ; motif: GGGGNGGGGGMKGGRRNGGNRN; match class: 1 | 4974 | 274 | 99 | 5.0E-02 |

GO:BP = Gene ontology biological process; GO:CC = Gene ontology cellular component; GO:MF = Gene ontology Molecular function; TF = transcription factor; MIRNA = micro RNAs; WP = Curated Wiki pathways; HPA= Human Protein Atlas.

### Supplementary Table 8

Look-up of CpGs in common between our sex-stratified EWAS of early preterm birth (ePTB), and the EWAS of gestational age and pre-menstrual age (PMA) among very preterm infants (< 30 weeks) reported by Hodge *et al.* 2024

| CpG | Near Gene | ePTB EWAS analysis | Coefficient ePTB boys | Coefficient ePTB girls | Coefficient GA Hodge | Directionality ePTB Boys-Girls-Hodge | Gene Name Hodge |
|---|---|---|---|---|---|---|---|
| *Gestational age signals in Hodge et al. 2024* | | | | | | | |
| cg02603733 | *POGZ;CGN* | boys | -0.11 | -0.07 | -0.11 | --- | |
| cg04813347 | *LINC01921;DIRC3-AS1* | boys | -0.12 | -0.06 | -0.10 | --- | *RUFY4;RUFY4* |
| cg22482278 | *RPL23AP87;NONE* | boys | 0.14 | -0.09 | 0.09 | +-+ | *SECTM1* |
| cg07920092 | *DGKZ* | boys | -0.11 | -0.07 | -0.09 | --- | *DGKZ* |
| cg13832290 | *SIT1* | boys | 0.10 | -0.09 | 0.13 | +-+ | *SIT1* |
| cg08224960 | *NOL8* | boys | 0.12 | -0.08 | 0.10 | +-+ | *C9orf3;C9orf3* |
| cg26683426 | *SORL1;MIR100HG* | boys | -0.10 | -0.04 | -0.10 | --- | |
| cg07164639 | *GPR6;WASF1* | boys | 0.10 | 0.08 | 0.09 | +++ | *DDO;DDO* |
| cg24867727 | *SERTAD2;LINC01800* | boys | 0.15 | -0.04 | 0.11 | +-+ | *SLC1A4;SLC1A4* |
| cg21658153 | *OR10H4;LINC00661* | boys | 0.11 | -0.08 | 0.11 | +-+ | *TPM4* |
| cg18039268 | *RNF24* | boys | 0.14 | -0.06 | 0.10 | +-+ | *RNF24;RNF24;RNF24* |
| cg05883794 | *C14orf177;BCL11B* | boys | -0.16 | -0.08 | -0.09 | --- | *SETD3;SETD3* |
| cg00187327 | *LINC02554;MN1* | boys | -0.12 | 0.08 | -0.09 | -+- | *MN1* |
| cg27548752 | *IGFL1;HIF3A* | boys | 0.12 | 0.09 | 0.09 | +++ | |
| cg22206745 | *FGF3;LINC02753* | boys | 0.13 | 0.05 | 0.13 | +++ | *ORAOV1* |
| cg07649206 | *TNIK* | boys | -0.10 | -0.04 | -0.09 | --- | *SLC2A2* |
| cg25212737 | *SPIDR* | boys | 0.13 | 0.03 | 0.15 | +++ | |
| cg08474901 | *CCDC3* | boys | -0.13 | -0.02 | -0.16 | --- | |
| cg14022913 | *SPCS3;VEGFC* | boys | 0.13 | -0.06 | 0.07 | +-+ | *NEIL3* |
| cg18168435 | *RBFOX2* | boys | 0.10 | -0.05 | 0.09 | +-+ | *APOL3* |
| cg08935541 | *LINC01589;ATXN10* | boys | 0.10 | 0.05 | 0.11 | +++ | *LINC00899* |
| cg07595197 | *MAF;MAFTRR* | boys | 0.11 | 0.06 | 0.09 | +++ | |
| cg05899788 | *ABLIM2;SH3TC1* | girls | 0.08 | 0.12 | 0.06 | +++ | *SH3TC1* |

| | | | | | | | |
|---|---|---|---|---|---|---|---|
| cg20254998 | *BCLAF1;MAP7* | girls | -0.09 | 0.11 | -0.08 | -+- | *MAP3K5* |
| cg09734183 | *RIN2* | girls | 0.10 | 0.13 | 0.09 | +++ | *RIN2;RIN2* |
| cg12148678 | *RBM42* | girls | -0.07 | -0.13 | -0.08 | --- | *TBCB;TBCB* |
| cg06670039 | *SNX29* | girls | -0.07 | 0.15 | -0.07 | -+- | *SNX29* |
| cg14864829 | *EIF2AK2* | girls | 0.10 | 0.12 | 0.10 | +++ | *QPCT* |
| cg17175421 | *TLE2* | girls | -0.06 | -0.13 | -0.10 | --- | *TLE2;TLE2;TLE2;TLE2* |
| cg03091585 | *CHD9* | girls | -0.05 | -0.14 | -0.07 | --- | *CHD9* |
| cg10153714 | *CAPZA2* | girls | 0.06 | -0.11 | 0.12 | +-+ | |
| cg06417243 | *LLGL2* | girls | 0.05 | -0.11 | 0.10 | +-+ | *SDK2* |
| cg02069095 | *GPR39* | joint | -0.12 | -0.14 | -0.07 | --- | *NCKAP5* |
| cg04067341 | *IRS2;LINC00396* | joint | 0.12 | 0.11 | 0.13 | +++ | *COL4A2-AS1;COL4A2* |
| cg08849509 | *SBF2-AS1* | joint | 0.11 | -0.13 | 0.11 | +-+ | *SBF2* |
| *Post-menstrual age signals in Hodge et al. 2024* | | | | | | | |
| cg00861010 | *SLFNL1-AS1* | boys | 0.13 | 0.05 | -0.16 | ++- | |
| cg11343820 | *LINC01951;DRD1* | boys | -0.17 | 0.08 | 0.10 | -++ | |
| cg07222631 | *LINC02554;MN1* | boys | -0.18 | -0.06 | 0.14 | --+ | |
| cg14871997 | *LINC01951;DRD1* | boys | -0.11 | -0.05 | 0.10 | --+ | |
| cg02458632 | *B3GNT2* | boys | 0.16 | -0.05 | -0.14 | +-- | |
| cg13413384 | *UCK1;PRRT1B* | boys | 0.13 | -0.09 | -0.10 | +-- | *RXRA* |
| cg01714756 | *PRIMA1;FAM181A-AS1* | boys | 0.16 | -0.09 | -0.16 | +-- | *SERPINA6* |
| cg27293447 | *LINC01799* | boys | -0.12 | -0.04 | 0.11 | --+ | *LOC102800447* |
| cg11712893 | *INHBB;LINC01101* | boys | 0.10 | -0.03 | -0.16 | +-- | |
| cg06325472 | *LINC01951;DRD1* | boys | -0.12 | -0.10 | 0.11 | --+ | |
| cg02603733 | *POGZ;CGN* | boys | -0.11 | -0.07 | 0.17 | --+ | |
| cg03862381 | *PGPEP1* | boys | 0.10 | -0.09 | -0.14 | +-- | *ELL* |
| cg03515773 | *EIF2D;DYRK3* | boys | -0.18 | -0.03 | 0.07 | --+ | |
| cg21361437 | *NCK2;ECRG4* | boys | 0.10 | 0.09 | -0.11 | ++- | |
| cg04813347 | *LINC01921;DIRC3-AS1* | boys | -0.12 | -0.06 | 0.09 | --+ | *RUFY4;RUFY4* |
| cg23982461 | *IQCJ-SCHIP1;SCHIP1* | boys | -0.13 | -0.06 | 0.09 | --+ | *IQCJ;IQCJ* |
| cg23565609 | *RPL23AP87;NONE* | boys | 0.13 | -0.04 | -0.15 | +-- | |
| cg22482278 | *RPL23AP87;NONE* | boys | 0.14 | -0.09 | -0.15 | +-- | *SECTM1* |

| | | | | | | | |
|---|---|---|---|---|---|---|---|
| cg06204531 | *GIMAP2* | boys | -0.13 | -0.02 | 0.14 | --+ | *ZNF775* |
| cg01206832 | *MIR4424;RALGPS2* | boys | 0.11 | 0.09 | -0.15 | ++- | |
| cg16041784 | *PLEKHA6* | boys | 0.11 | -0.06 | -0.10 | +-- | *PLEKHA6* |
| cg07920092 | *DGKZ* | boys | -0.11 | -0.07 | 0.15 | --+ | *DGKZ* |
| cg03837680 | *DCXR* | boys | -0.13 | -0.04 | 0.18 | --+ | *TBC1D16* |
| cg13832290 | *SIT1* | boys | 0.10 | -0.09 | -0.12 | +-- | *SIT1* |
| cg14880229 | *LOC101929710* | boys | 0.10 | -0.02 | -0.12 | +-- | |
| cg19580937 | *TLE3;LINC02205* | boys | -0.14 | -0.05 | 0.13 | --+ | |
| cg00120416 | *MCM4;PRKDC* | boys | 0.11 | 0.08 | -0.07 | ++- | |
| cg19034820 | *MN1* | boys | -0.10 | -0.03 | 0.12 | --+ | *TTC28* |
| cg03142049 | *PCDHA1* | boys | 0.11 | -0.06 | -0.10 | +-- | *CYSTM1* |
| cg09144398 | *DDX6;CXCR5* | boys | 0.12 | 0.09 | -0.17 | ++- | *TREH* |
| cg23173566 | *RBMS3* | boys | -0.16 | 0.05 | 0.10 | -++ | *RBMS3;RBMS3;RBMS3* |
| cg08583624 | *LINC02267* | boys | -0.11 | 0.05 | 0.11 | -++ | *STPG2-AS1* |
| cg23173746 | *RFX8;MAP4K4* | boys | 0.11 | -0.05 | -0.05 | +-- | *IL1RL2* |
| cg08780680 | *LINC00327;TNFRSF19* | boys | 0.11 | -0.07 | -0.12 | +-- | |
| cg15060258 | *RCSD1* | boys | 0.13 | 0.08 | -0.13 | ++- | *RCSD1* |
| cg09750115 | *DUSP26;LINC01288* | boys | 0.12 | -0.06 | -0.09 | +-- | *RNF122* |
| cg08224960 | *NOL8* | boys | 0.12 | -0.08 | -0.14 | +-- | *C9orf3;C9orf3* |
| cg20463533 | *TCERG1L* | boys | 0.11 | 0.08 | -0.13 | ++- | |
| cg24541835 | *VPS13D;DHRS3* | boys | 0.14 | -0.05 | -0.15 | +-- | *DHRS3* |
| cg15661950 | *TMC3-AS1;MEX3B* | boys | 0.12 | 0.05 | -0.15 | ++- | |
| cg26683426 | *SORL1;MIR100HG* | boys | -0.10 | -0.04 | 0.15 | --+ | |
| cg23873636 | *ADGRF1;TNFRSF21* | boys | 0.12 | -0.08 | -0.16 | +-- | |
| cg09276050 | *ALKBH4* | boys | -0.12 | -0.10 | 0.12 | --+ | *CUX1* |
| cg09364402 | *ANKHD1;ANKHD1-EIF4EBP3* | boys | 0.10 | -0.07 | -0.09 | +-- | *NRG2* |
| cg19004811 | *EIF2B2* | boys | -0.11 | 0.07 | 0.10 | -++ | |
| cg05906420 | *TNFRSF21;CD2AP* | boys | 0.11 | -0.03 | -0.12 | +-- | *TNFRSF21* |
| cg24765446 | *EYA2* | boys | 0.11 | 0.10 | -0.15 | ++- | *WFDC6* |
| cg00984412 | *GCDH* | boys | 0.12 | 0.06 | -0.12 | ++- | *NFIX* |
| cg02451350 | *ST7* | boys | 0.10 | -0.06 | -0.12 | +-- | |

| | | | | | | | |
|---|---|---|---|---|---|---|---|
| cg07164639 | *GPR6;WASF1* | boys | 0.10 | 0.08 | -0.18 | ++- | *DDO;DDO* |
| cg10424513 | *LOC100129620;PLPPR4* | boys | 0.14 | 0.07 | -0.10 | ++- | *PALMD* |
| cg09031352 | *DGKI* | boys | -0.17 | -0.03 | 0.04 | --+ | *PTN* |
| cg19238575 | *COL22A1;KCNK9* | boys | -0.15 | -0.07 | 0.09 | --+ | *TRAPPC9* |
| cg24867727 | *SERTAD2;LINC01800* | boys | 0.15 | -0.04 | -0.12 | +-- | *SLC1A4;SLC1A4* |
| cg25909582 | *HORMAD2* | boys | 0.11 | -0.02 | -0.11 | +-- | *SEC14L4;SEC14L4* |
| cg27609217 | *LOC101928894;CREB3L1* | boys | -0.11 | -0.04 | 0.06 | --+ | |
| cg04342509 | *MTX2* | boys | 0.16 | 0.09 | -0.07 | ++- | |
| cg21956384 | *APBB2* | boys | -0.11 | -0.06 | 0.06 | --+ | *APBB2* |
| cg07532353 | *MIR5093;GSE1* | boys | -0.12 | 0.05 | 0.06 | -++ | |
| cg16003049 | *INS-IGF2* | boys | 0.16 | -0.07 | -0.08 | +-- | *TH;TH;TH* |
| cg11889960 | *EXT1* | boys | 0.11 | -0.05 | -0.06 | +-- | *COLEC10* |
| cg06593560 | *SASH1* | boys | -0.12 | -0.03 | 0.10 | --+ | *UST* |
| cg18039268 | *RNF24* | boys | 0.14 | -0.06 | -0.10 | +-- | *RNF24* |
| cg05883794 | *C14orf177;BCL11B* | boys | -0.16 | -0.08 | 0.16 | --+ | *SETD3;SETD3* |
| cg00187327 | *LINC02554;MN1* | boys | -0.12 | 0.08 | 0.16 | -++ | *MN1* |
| cg13907014 | *LINC02801* | boys | -0.11 | 0.02 | 0.13 | -++ | |
| cg00251358 | *XRCC4* | boys | -0.13 | -0.04 | 0.10 | --+ | |
| cg09999715 | *SNX18;LOC102467080* | boys | -0.10 | -0.06 | 0.09 | --+ | *ARL15* |
| cg21919117 | *DDX47* | boys | -0.10 | 0.03 | 0.07 | -++ | *LOC100506314;HEBP1* |
| cg01890836 | *COPS3* | boys | 0.10 | -0.03 | -0.15 | +-- | *MPRIP;MPRIP* |
| cg10644659 | *ALKBH4* | boys | -0.11 | -0.04 | 0.11 | --+ | *CUX1* |
| cg14153983 | *ZHX3* | boys | 0.16 | -0.06 | -0.10 | +-- | |
| cg17470895 | *RNF182;CD83* | boys | -0.13 | -0.02 | 0.06 | --+ | |
| cg13538965 | *EPS8* | boys | -0.13 | 0.07 | 0.06 | -++ | |
| cg16813564 | *PPP1R9A* | boys | -0.14 | -0.04 | 0.11 | --+ | *SGCE* |
| cg20599118 | *MTUS1* | boys | 0.12 | -0.05 | -0.10 | +-- | *MTUS1* |
| cg24493244 | *FAM230G* | boys | -0.11 | -0.06 | 0.05 | --+ | *MED15* |
| cg26897909 | *CTSE;SRGAP2* | boys | -0.10 | -0.04 | 0.05 | --+ | *SRGAP2* |
| cg08248287 | *TNIK;PLD1* | boys | 0.12 | -0.04 | -0.09 | +-- | *TNIK* |
| cg27548752 | *IGFL1;HIF3A* | boys | 0.12 | 0.09 | -0.14 | ++- | |

| | | | | | | | |
|---|---|---|---|---|---|---|---|
| cg27244773 | *EFCAB13* | boys | -0.11 | -0.04 | 0.08 | --+ | *ARHGAP27* |
| cg22866411 | *CACNA2D3* | boys | -0.13 | 0.07 | 0.13 | -++ | *CACNA2D3-AS1;CACNA2D3* |
| cg24793045 | *DDR2* | boys | 0.10 | 0.06 | -0.09 | ++- | *DDR2* |
| cg18774577 | *LINC00562* | boys | 0.14 | -0.04 | -0.14 | +-- | *RCBTB2* |
| cg22206745 | *FGF3;LINC02753* | boys | 0.13 | 0.05 | -0.19 | ++- | *ORAOV1* |
| cg22797643 | *LINC02077;ROBO1* | boys | -0.11 | 0.07 | 0.07 | -++ | |
| cg23825480 | *SEC14L6* | boys | -0.10 | 0.02 | 0.08 | -++ | *MORC2* |
| cg24401660 | *ZWILCH* | boys | 0.11 | -0.09 | -0.07 | +-- | |
| cg25368317 | *CRTAC1* | boys | 0.13 | -0.09 | -0.08 | +-- | *ENTPD7* |
| cg26980152 | *RPTOR* | boys | -0.14 | -0.05 | 0.08 | --+ | *CYTH1* |
| cg07649206 | *TNIK* | boys | -0.10 | -0.04 | 0.12 | --+ | *SLC2A2* |
| cg27438425 | *SLFNL1-AS1* | boys | -0.10 | -0.04 | 0.10 | --+ | |
| cg13420277 | *CREB5* | boys | 0.14 | 0.07 | -0.15 | ++- | *CREB5* |
| cg25274540 | *HSD17B12;ALKBH3* | boys | -0.11 | 0.00 | 0.07 | --+ | *ALKBH3;LOC729799* |
| cg25212737 | *SPIDR* | boys | 0.13 | 0.03 | -0.21 | ++- | |
| cg23543824 | *UGT2B15* | boys | -0.16 | -0.02 | 0.09 | --+ | *UGT2B4* |
| cg08406348 | *TACR1;EVA1A* | boys | -0.10 | -0.03 | 0.13 | --+ | |
| cg06013117 | *LINC01951;DRD1* | boys | -0.15 | -0.09 | 0.12 | --+ | *MSX2* |
| cg23656872 | *RCSD1* | boys | 0.13 | 0.06 | -0.10 | ++- | *RCSD1* |
| cg04800041 | *LOC105373782;FSIP2-AS2* | boys | 0.12 | 0.05 | -0.07 | ++- | |
| cg00191665 | *EFCC1* | boys | -0.14 | -0.07 | 0.09 | --+ | *RAB7A* |
| cg19014858 | *MYBPC2* | boys | -0.10 | -0.05 | 0.12 | --+ | *KLK6* |
| cg19864972 | *MIR4675;NEBL* | boys | 0.11 | -0.04 | -0.09 | +-- | *NEBL* |
| cg20087585 | *GIMAP2* | boys | -0.12 | -0.03 | 0.11 | --+ | *ZNF775* |
| cg08474901 | *CCDC3* | boys | -0.13 | -0.02 | 0.15 | --+ | |
| cg10307152 | *VPS13D;DHRS3* | boys | -0.15 | -0.02 | 0.04 | --+ | *DHRS3* |
| cg23941537 | *DACH1;MZT1* | boys | -0.13 | -0.04 | 0.07 | --+ | |
| cg06764229 | *LINC01108;JARID2* | boys | -0.11 | 0.05 | 0.05 | -++ | |
| cg16871855 | *LINC00824;LINC00976* | boys | -0.11 | 0.07 | 0.07 | -++ | *FAM49B;FAM49B* |
| cg17744019 | *GRAMD4* | boys | -0.10 | -0.04 | 0.09 | --+ | *TBC1D22A* |
| cg07317567 | *BCKDHB;TENT5A* | boys | -0.13 | -0.05 | 0.08 | --+ | |

| | | | | | | | |
|---|---|---|---|---|---|---|---|
| cg25832180 | *MAGI2* | boys | -0.11 | -0.08 | 0.09 | --+ | *MAGI2* |
| cg14022913 | *SPCS3;VEGFC* | boys | 0.13 | -0.06 | -0.05 | +-- | *NEIL3* |
| cg20016416 | *PC* | boys | -0.10 | -0.03 | 0.08 | --+ | *SPTBN2* |
| cg13908635 | *TNS1;LOC105373878* | boys | 0.10 | -0.04 | -0.08 | +-- | *CYP27A1* |
| cg26169783 | *STARD13* | boys | 0.11 | -0.03 | -0.15 | +-- | |
| cg18168435 | *RBFOX2* | boys | 0.10 | -0.05 | -0.11 | +-- | *APOL3* |
| cg03242909 | *LINC01951;DRD1* | boys | -0.12 | -0.06 | 0.10 | --+ | |
| cg14538218 | *IL21-AS1* | boys | -0.12 | -0.08 | 0.04 | --+ | |
| cg08935541 | *LINC01589;ATXN10* | boys | 0.10 | 0.05 | -0.14 | ++- | *LINC00899* |
| cg01049503 | *KALRN* | boys | -0.11 | 0.04 | 0.09 | -++ | |
| cg17883068 | *ZNF280D* | boys | 0.15 | -0.05 | -0.06 | +-- | *TCF12* |
| cg18522395 | *ZNF433-AS1* | boys | 0.12 | 0.00 | -0.09 | +-- | *ZNF788* |
| cg17083209 | *ADGRL2* | boys | -0.13 | 0.06 | 0.06 | -++ | *LPHN2* |
| cg07141356 | *HAAO;LINC01819* | boys | -0.12 | -0.05 | 0.08 | --+ | |
| cg27060294 | *LDB2* | boys | 0.10 | -0.04 | -0.07 | +-- | *LDB2;LDB2* |
| cg01942816 | *FBXL18* | boys | 0.10 | -0.04 | -0.12 | +-- | *MIR589;FBXL18* |
| cg02605091 | *CAMTA1* | boys | 0.10 | -0.08 | -0.10 | +-- | *CAMTA1* |
| cg20324727 | *C5orf63;MEGF10* | boys | 0.13 | 0.00 | -0.11 | ++- | *PHAX* |
| cg25114899 | *MIR1269A;LOC101927237* | boys | -0.14 | -0.03 | 0.11 | --+ | *GNRHR;GNRHR* |
| cg13632752 | *MIR1268A* | boys | -0.10 | -0.03 | 0.07 | --+ | |
| cg19374905 | *LINC01921;DIRC3-AS1* | boys | -0.10 | 0.02 | 0.06 | -++ | |
| cg01451808 | *PRKDC* | boys | -0.12 | -0.05 | 0.06 | --+ | |
| cg04302055 | *SUMO1P1;BCAS1* | boys | -0.11 | -0.01 | 0.12 | --+ | |
| cg04111054 | *C4orf33;LINC02466* | boys | -0.13 | -0.02 | 0.11 | --+ | |
| cg23063382 | *MIR3144;TBC1D32* | boys | 0.12 | -0.07 | -0.13 | +-- | *TBC1D32;TBC1D32* |
| cg26873042 | *PIK3R1;LINC02198* | boys | -0.10 | 0.05 | 0.05 | -++ | |
| cg07478690 | *SLC35F1* | boys | 0.12 | -0.05 | -0.11 | +-- | |
| cg11902170 | *ANKRD22* | boys | -0.11 | -0.08 | 0.07 | --+ | |
| cg14980042 | *LINC01736;GALNT2* | boys | -0.12 | -0.09 | 0.06 | --+ | *GALNT2* |
| cg13495130 | *LINC01736;GALNT2* | boys | -0.11 | -0.04 | 0.08 | --+ | *GALNT3* |
| cg08315524 | *RAB3C;PDE4D* | boys | -0.11 | -0.02 | 0.09 | --+ | |

| | | | | | | | |
|---|---|---|---|---|---|---|---|
| cg20485781 | *LBHD2* | boys | 0.11 | -0.04 | -0.05 | +-- | |
| cg07595197 | *MAF;MAFTRR* | boys | 0.11 | 0.06 | -0.13 | ++- | |
| cg05688017 | *ATP6V0A1* | boys | -0.11 | -0.07 | 0.09 | --+ | |
| cg13286635 | *LINC01491;SLC24A5* | boys | 0.10 | -0.07 | -0.13 | +-- | |
| cg10387952 | *CSGALNACT1* | boys | -0.10 | -0.03 | 0.05 | --+ | |
| cg12678751 | *SOX1-OT* | boys | -0.12 | -0.02 | 0.07 | --+ | *ATP11A;ATP11A* |
| cg03078488 | *IGF2BP3* | boys | -0.13 | -0.05 | 0.07 | --+ | *IGF2BP3* |
| cg08257257 | *MRGPRD;MRGPRF* | boys | -0.11 | -0.05 | 0.04 | --+ | *MTL5;MTL5* |
| cg17326611 | *EGLN3;SPTSSA* | boys | -0.11 | 0.09 | 0.06 | -++ | |
| cg11201229 | *PIK3C2A* | boys | -0.10 | -0.01 | 0.14 | --+ | *PIK3C2A* |
| cg17889259 | *KCNMA1* | boys | 0.12 | -0.03 | -0.10 | +-- | *ZMIZ1* |
| cg06679296 | *LINC01150* | boys | -0.11 | -0.04 | 0.18 | --+ | *TNNT3* |
| cg09340066 | *MAPKAP1* | boys | 0.10 | 0.02 | -0.08 | ++- | *SLC27A4* |
| cg19743728 | *KLHDC2* | boys | -0.11 | -0.03 | 0.08 | --+ | *L2HGDH* |
| cg03728930 | *KCNQ3* | boys | 0.17 | -0.03 | -0.06 | +-- | *NDRG1* |
| cg22749750 | *NONE;SLITRK1* | boys | 0.12 | -0.06 | -0.18 | +-- | |
| cg02806631 | *PARD3* | boys | -0.10 | -0.04 | 0.08 | --+ | |
| cg21159128 | *GLIS1;NDC1* | boys | -0.10 | -0.05 | 0.07 | --+ | *SSBP3;SSBP3;SSBP3* |
| cg16283919 | *LOC441087;SV2C* | boys | 0.12 | 0.03 | -0.10 | ++- | |
| cg10347254 | *MERTK* | boys | 0.11 | -0.03 | -0.12 | +-- | |
| cg00314861 | *TOR1AIP2* | boys | -0.11 | -0.02 | 0.06 | --+ | |
| cg22413388 | *FBLN1* | boys | -0.11 | -0.03 | 0.11 | --+ | *WNT7B* |
| cg05297739 | *CLIC6* | boys | 0.11 | -0.05 | -0.12 | +-- | *CBR1* |
| cg02046552 | *BMP1* | boys | -0.12 | -0.03 | 0.19 | --+ | *[illegible]PB49;EPB49;EPB49;EPB4[illegible]* |
| cg01423044 | *ERICH6B* | boys | -0.13 | -0.02 | 0.08 | --+ | *LCP1* |
| cg05527149 | *RCC1L* | boys | -0.10 | 0.07 | 0.07 | -++ | *GTF2IRD1* |
| cg02865822 | *DGKZ* | girls | -0.09 | -0.12 | 0.12 | --+ | *DGKZ* |
| cg17266233 | *DGKZ* | girls | -0.07 | 0.11 | 0.10 | -++ | *DGKZ* |
| cg19752649 | *CLEC16A* | girls | -0.09 | 0.12 | 0.06 | -++ | *CLEC16A;CLEC16A* |
| cg06285241 | *CDCA4* | girls | -0.06 | -0.11 | 0.08 | --+ | *CRIP2* |
| cg02635407 | *ABLIM2;SH3TC1* | girls | 0.06 | -0.13 | -0.05 | +-- | *SH3TC1* |

| | | | | | | | |
|---|---|---|---|---|---|---|---|
| cg00810262 | *LINC01982;LINC02089* | girls | 0.05 | -0.13 | -0.09 | +-- | *ABCC3* |
| cg21370660 | *PRDM11* | girls | 0.06 | -0.11 | -0.07 | +-- | *SYT13;SYT13* |
| cg09878710 | *GPC6* | girls | -0.06 | 0.14 | 0.05 | -++ | |
| cg27605998 | *CREB3L1* | girls | -0.10 | -0.12 | 0.06 | --+ | *DGKZ* |
| cg05899788 | *ABLIM2;SH3TC1* | girls | 0.08 | 0.12 | -0.06 | ++- | *SH3TC1* |
| cg04266340 | *SLC26A9;FAM72A* | girls | -0.07 | 0.13 | 0.10 | -++ | *C1orf186* |
| cg02769133 | *ITIH5* | girls | 0.05 | -0.15 | -0.09 | +-- | *ITIH5;ITIH5* |
| cg25766247 | *LOC286178;LINC01602* | girls | -0.04 | -0.10 | 0.05 | --+ | |
| cg03185144 | *KIAA0513* | girls | 0.06 | -0.13 | -0.07 | +-- | *KIAA0513* |
| cg01599826 | *LPAR1;MIR7702* | girls | 0.08 | 0.11 | -0.05 | ++- | *ZNF618* |
| cg16619139 | *DGKZ* | girls | -0.08 | 0.11 | 0.09 | -++ | *DGKZ* |
| cg11767915 | *USP25;MIR99AHG* | girls | -0.07 | -0.11 | 0.06 | --+ | *LINC01549;LINC01549* |
| cg12429481 | *LINC01441;CBLN4* | girls | 0.12 | -0.11 | -0.06 | +-- | |
| cg20254998 | *BCLAF1;MAP7* | girls | -0.09 | 0.11 | 0.11 | -++ | *MAP3K5* |
| cg25193276 | *CITED4;CTPS1* | girls | -0.07 | -0.14 | 0.09 | --+ | |
| cg21017775 | *ELF5* | girls | -0.07 | 0.14 | 0.09 | -++ | *ELF5* |
| cg09734183 | *RIN2* | girls | 0.10 | 0.13 | -0.12 | ++- | *RIN2;RIN2* |
| cg06170720 | *HES2* | girls | -0.08 | 0.12 | 0.12 | -++ | *PLEKHG5* |
| cg00760938 | *LINC00607* | girls | -0.09 | -0.11 | 0.09 | --+ | |
| cg23599026 | *HPSE2* | girls | -0.03 | 0.12 | 0.03 | -++ | *SEC31B* |
| cg23604266 | *LINC00487;NRIR* | girls | 0.08 | -0.11 | -0.18 | +-- | *RNF144A* |
| cg24435571 | *SLF2* | girls | -0.08 | -0.11 | 0.10 | --+ | *ARL3* |
| cg17499490 | *PARP1;STUM* | girls | -0.04 | 0.11 | 0.07 | -++ | *C1orf95* |
| cg03163262 | *LINC01173;SH3BP4* | girls | -0.06 | -0.11 | 0.09 | --+ | *AGAP1;AGAP1;AGAP1* |
| cg10922765 | *LMCD1* | girls | -0.07 | 0.10 | 0.08 | -++ | |
| cg09825286 | *MGLL;KBTBD12* | girls | -0.03 | -0.12 | 0.05 | --+ | *TPRA1;TPRA1;TPRA1* |
| cg21737106 | *LOC105375075;SUPT3H* | girls | -0.09 | -0.10 | 0.13 | --+ | |
| cg16412082 | *ADRA2A;HEAT2* | girls | -0.03 | -0.12 | 0.05 | --+ | *LOC143188* |
| cg10094994 | *TRAF3IP1* | girls | -0.03 | 0.14 | 0.05 | -++ | *HDAC4* |
| cg19547694 | *LINC01310;NONE* | girls | -0.04 | 0.13 | 0.06 | -++ | *BRD1;LOC90834* |
| cg01948724 | *FLYWCH1* | girls | -0.07 | 0.12 | 0.11 | -++ | |

| | | | | | | | |
|---|---|---|---|---|---|---|---|
| cg23659250 | *LINC01310;NONE* | girls | -0.06 | -0.10 | 0.07 | --+ | *BRD1;LOC90834* |
| cg24986815 | *ITPKB;PSEN2* | girls | 0.06 | -0.13 | -0.05 | +-- | *ADCK3* |
| cg05205239 | *LOC102724265;LINC01488* | girls | -0.09 | -0.12 | 0.11 | --+ | |
| cg21406409 | *KIF1B* | girls | -0.05 | 0.11 | 0.08 | -++ | *KIF1B* |
| cg26701500 | *ACTR5* | girls | -0.03 | -0.10 | 0.06 | --+ | *SRC;SRC* |
| cg12494143 | *SMOX* | girls | -0.06 | 0.11 | 0.07 | -++ | *SMOX* |
| cg14080475 | *LRRC37A5P;DNAJC25* | girls | -0.05 | 0.10 | 0.12 | -++ | *AKNA* |
| cg11359720 | *UBASH3A* | girls | -0.05 | 0.13 | 0.04 | -++ | |
| cg22705929 | *CTNND1;OR9Q1* | girls | -0.06 | -0.18 | 0.07 | --+ | *YPEL4* |
| cg15422377 | *AIF1L* | girls | -0.03 | -0.11 | 0.08 | --+ | *VAV2;VAV2* |
| cg12148678 | *RBM42* | girls | -0.07 | -0.13 | 0.06 | --+ | *TBCB;TBCB* |
| cg14010305 | *MAD1L1* | girls | -0.06 | -0.12 | 0.13 | --+ | *MAD1L1;MAD1L1;MAD1L1* |
| cg20789819 | *MKLN1* | girls | 0.04 | -0.16 | -0.07 | +-- | |
| cg10259077 | *DECR2* | girls | -0.02 | -0.11 | 0.06 | --+ | *RAB11FIP3* |
| cg03662398 | *MSI2* | girls | -0.07 | -0.12 | 0.06 | --+ | |
| cg17716056 | *KCNQ5* | girls | 0.08 | 0.10 | -0.08 | ++- | |
| cg17364007 | *MYO7B* | girls | -0.07 | 0.13 | 0.07 | -++ | |
| cg09253641 | *SMIM2* | girls | -0.03 | -0.10 | 0.06 | --+ | |
| cg10369363 | *SCN8A* | girls | -0.03 | 0.11 | 0.05 | -++ | *GRASP* |
| cg24038105 | *KAZN* | girls | 0.06 | -0.10 | -0.07 | +-- | *FHAD1* |
| cg08900426 | *MAP6;MOGAT2* | girls | -0.03 | -0.13 | 0.09 | --+ | |
| cg27526774 | *PP2672;LOC101927851* | girls | -0.10 | -0.13 | 0.05 | --+ | |
| cg19206131 | *ASXL2* | girls | 0.07 | 0.12 | -0.07 | ++- | |
| cg06670039 | *SNX29* | girls | -0.07 | 0.15 | 0.07 | -++ | *SNX29* |
| cg23657589 | *ELF5* | girls | -0.05 | 0.13 | 0.09 | -++ | |
| cg26867637 | *ABR* | girls | -0.08 | -0.10 | 0.07 | --+ | *ABR;ABR;ABR;ABR* |
| cg18978125 | *GET4* | girls | -0.11 | -0.10 | 0.12 | --+ | *ADAP1* |
| cg19938826 | *TCERG1* | girls | -0.10 | -0.11 | 0.10 | --+ | |
| cg11037750 | *CYP2T1P;CYP2A6* | girls | -0.02 | 0.12 | 0.03 | -++ | *TGFB1* |
| cg16736924 | *CD200;BTLA* | girls | -0.08 | 0.12 | 0.04 | -++ | *C3orf52;C3orf52* |
| cg12828504 | *FAAP20* | girls | -0.07 | -0.13 | 0.07 | --+ | *RKCZ;PRKCZ;PRKCZ;PRKC* |

| | | | | | | | |
|---|---|---|---|---|---|---|---|
| cg12223498 | *LINC01370;MAFB* | girls | 0.06 | -0.17 | -0.07 | +-- | |
| cg04744624 | *NRCAM* | girls | -0.06 | -0.17 | 0.11 | --+ | *LAMB1* |
| cg07585771 | *CNOT10;TRIM71* | girls | -0.05 | -0.13 | 0.08 | --+ | *TRIM71* |
| cg05315047 | *NME9* | girls | -0.06 | -0.14 | 0.05 | --+ | |
| cg13037081 | *SLC22A12* | girls | -0.04 | 0.15 | 0.06 | -++ | *PS6KA4;MIR1237;RPS6KA* |
| cg25289862 | *CCDC77* | girls | 0.10 | 0.11 | -0.07 | ++- | *B4GALNT3* |
| cg06906523 | *RAB3GAP2* | girls | 0.11 | 0.11 | -0.13 | ++- | |
| cg07904475 | *IQCM* | girls | -0.03 | -0.13 | 0.05 | --+ | *LRBA* |
| cg18245735 | *TTLL4* | girls | -0.09 | 0.13 | 0.09 | -++ | |
| cg14864829 | *EIF2AK2* | girls | 0.10 | 0.12 | -0.09 | ++- | *QPCT* |
| cg20744302 | *CNOT1;SLC38A7* | girls | 0.09 | -0.11 | -0.07 | +-- | *SLC38A7;SLC38A7* |
| cg06513971 | *ZBTB25* | girls | 0.08 | -0.11 | -0.11 | +-- | *RAB15;CHURC1-FNTB* |
| cg15150970 | *DNAJC27-AS1* | girls | -0.04 | -0.11 | 0.05 | --+ | *NMT3A;DNMT3A;DNMT3A* |
| cg09336794 | *HES5;TNFRSF14-AS1* | girls | -0.05 | -0.11 | 0.08 | --+ | *PLCH2;PLCH2;PLCH2* |
| cg27199647 | *ATR* | girls | -0.06 | -0.14 | 0.06 | --+ | |
| cg17331032 | *ROR2* | girls | -0.02 | -0.11 | 0.06 | --+ | *FBP1;FBP1* |
| cg17175421 | *TLE2* | girls | -0.06 | -0.13 | 0.10 | --+ | *TLE2;TLE2;TLE2;TLE2* |
| cg19302297 | *SPRED2;MIR4778* | girls | 0.05 | 0.11 | -0.10 | ++- | |
| cg08362543 | *TSC22D2;SERP1* | girls | -0.05 | -0.13 | 0.05 | --+ | |
| cg15318882 | *LINC00173;MAP1LC3B2* | girls | -0.03 | 0.11 | 0.04 | -++ | *LOC100506551;FBXW8* |
| cg23045350 | *UVRAG* | girls | -0.01 | -0.10 | 0.06 | --+ | |
| cg16498391 | *PRELID3A* | girls | -0.05 | 0.10 | 0.11 | -++ | *SLMO1;SLMO1* |
| cg20924966 | *SCN8A* | girls | -0.02 | 0.12 | 0.06 | -++ | *GRASP;GRASP;GRASP* |
| cg06661904 | *DYNLL1;COQ5* | girls | -0.07 | -0.10 | 0.06 | --+ | |
| cg10729019 | *SLC24A3* | girls | -0.08 | 0.12 | 0.13 | -++ | *SLC24A3* |
| cg09694782 | *GPAT2;ADRA2B* | girls | -0.05 | 0.10 | 0.05 | -++ | |
| cg02172262 | *MARS* | girls | -0.10 | -0.11 | 0.07 | --+ | |
| cg17806418 | *MOG* | girls | -0.05 | 0.12 | 0.07 | -++ | *GABBR1;GABBR1* |
| cg13555278 | *MAN1C1* | girls | -0.03 | -0.12 | 0.05 | --+ | *EXTL1;EXTL1* |
| cg13545247 | *KRT87P;KRT86* | girls | -0.03 | -0.11 | 0.08 | --+ | |
| cg19352801 | *RPS6KA4;LINC02724* | girls | -0.03 | -0.13 | 0.06 | --+ | *STIP1* |

| | | | | | | | |
|---|---|---|---|---|---|---|---|
| cg23449231 | *GFPT2;CNOT6* | girls | 0.09 | -0.14 | -0.09 | +-- | *SQSTM1;SQSTM1* |
| cg06130893 | *RPH3A* | girls | -0.06 | -0.11 | 0.06 | --+ | *SLC8B1* |
| cg02116058 | *CEP63* | girls | -0.05 | 0.10 | 0.11 | -++ | |
| cg00409842 | *PDE8A* | girls | 0.10 | -0.11 | -0.08 | +-- | *AKAP13;AKAP13* |
| cg14177325 | *WDR81* | girls | -0.07 | -0.14 | 0.04 | --+ | *SCARF1* |
| cg00819441 | *RNF213-AS1* | girls | -0.08 | -0.10 | 0.08 | --+ | *PGS1;PGS1;PGS1;PGS1* |
| cg07560287 | *FOXP4* | girls | -0.04 | -0.11 | 0.07 | --+ | *FOXP4;FOXP4;FOXP4* |
| cg07244414 | *AFAP1* | girls | -0.10 | -0.12 | 0.06 | --+ | *AFAP1;AFAP1* |
| cg03091585 | *CHD9* | girls | -0.05 | -0.14 | 0.10 | --+ | *CHD9* |
| cg20858526 | *TMEM51-AS1* | girls | -0.02 | -0.12 | 0.05 | --+ | *CTRC* |
| cg02298220 | *ARHGAP40;SLC32A1* | girls | -0.08 | -0.12 | 0.08 | --+ | |
| cg08333564 | *PARP11;CCND2-AS1* | girls | -0.05 | 0.10 | 0.04 | -++ | *CCND2* |
| cg27072594 | *ATP5MC3;LNPK* | girls | 0.05 | 0.10 | -0.07 | ++- | |
| cg09013732 | *EML1* | girls | -0.04 | -0.13 | 0.05 | --+ | *WDR25;WDR25* |
| cg04512899 | *RNGTT* | girls | -0.05 | -0.15 | 0.05 | --+ | *ANKRD6;ANKRD6;ANKRD6* |
| cg07937270 | *ERI3* | girls | 0.08 | -0.12 | -0.07 | +-- | *TCTEX1D4;TCTEX1D4* |
| cg10978403 | *PRDX5;CCDC88B* | girls | -0.05 | -0.12 | 0.07 | --+ | *FLRT1;MACROD1* |
| cg24196622 | *LINC00276;LRATD1* | girls | -0.03 | 0.11 | 0.08 | -++ | |
| cg02600660 | *SLC9A9* | girls | -0.03 | -0.10 | 0.05 | --+ | *SLC9A9* |
| cg00109834 | *XRN1* | girls | -0.09 | 0.10 | 0.06 | -++ | *TFDP2;TFDP2* |
| cg26782908 | *HAS2-AS1;SMILR* | girls | 0.06 | -0.14 | -0.11 | +-- | *LINC01151* |
| cg14008271 | *IFNG-AS1;IFNG* | girls | 0.09 | -0.10 | -0.08 | +-- | *LOC100507195* |
| cg25183214 | *MIR4268;EPHA4* | girls | -0.06 | -0.10 | 0.06 | --+ | *EPHA4* |
| cg01152374 | *GLCE* | girls | -0.03 | -0.11 | 0.10 | --+ | |
| cg25498638 | *BLOC1S5-TXNDC5* | girls | -0.04 | -0.13 | 0.05 | --+ | *BLOC1S5-TXNDC5* |
| cg10553109 | *TBX5-AS1;TBX3* | girls | 0.04 | 0.11 | -0.06 | ++- | |
| cg23093954 | *CERS6* | girls | -0.05 | -0.10 | 0.05 | --+ | *KLHL41* |
| cg02519744 | *TSNAX-DISC1;SIPA1L2* | girls | 0.05 | 0.11 | -0.09 | ++- | |
| cg26954761 | *TSHR* | girls | -0.07 | 0.11 | 0.11 | -++ | *LOC100506700* |
| cg21023952 | *SMIM15-AS1* | girls | -0.06 | -0.10 | 0.07 | --+ | *DE4D;PART1;PART1;PART* |
| cg07606115 | *ZBTB44-DT* | girls | -0.06 | 0.12 | 0.06 | -++ | *ST14* |

| | | | | | | | |
|---|---|---|---|---|---|---|---|
| cg20045250 | *LINC01695;LINC00161* | girls | -0.10 | 0.11 | 0.05 | -++ | *GRIK1;GRIK1* |
| cg04372478 | *EPB41L5* | girls | -0.04 | -0.10 | 0.08 | --+ | *GLI2* |
| cg00220152 | *RSPH6A* | girls | -0.10 | -0.11 | 0.10 | --+ | *IF3A;HIF3A;HIF3A;RNU6-66* |
| cg20569075 | *LINC01206;LINC01994* | girls | -0.04 | -0.11 | 0.05 | --+ | |
| cg25930644 | *MYH10;CCDC42* | girls | -0.03 | -0.13 | 0.08 | --+ | *MYH10* |
| cg10153714 | *CAPZA2* | girls | 0.06 | -0.11 | -0.09 | +-- | |
| cg07846061 | *RFWD3* | girls | 0.08 | -0.11 | -0.10 | +-- | *MLKL;MLKL* |
| cg20405062 | *ALK;YPEL5* | girls | -0.03 | 0.11 | 0.07 | -++ | |
| cg06564875 | *LINC02227;EBF1* | girls | -0.08 | -0.10 | 0.05 | --+ | |
| cg00560277 | *GLI3* | girls | -0.03 | 0.11 | 0.08 | -++ | *GLI3* |
| cg17306116 | *ESRRB;VASH1* | girls | -0.04 | -0.12 | 0.06 | --+ | |
| cg12915526 | *RORA* | girls | -0.03 | -0.17 | 0.07 | --+ | *RORA* |
| cg26222075 | *NMUR2;LINC01470* | girls | -0.08 | -0.11 | 0.10 | --+ | *CTB-12O2.1* |
| cg04316096 | *CEP128;TSHR* | girls | -0.05 | 0.15 | 0.05 | -++ | *STON2;STON2* |
| cg18994049 | *ANO1* | girls | -0.04 | -0.14 | 0.07 | --+ | |
| cg02057912 | *JDP2;BATF* | girls | -0.03 | -0.10 | 0.07 | --+ | *TGFB3* |
| cg06417243 | *LLGL2* | girls | 0.05 | -0.11 | -0.08 | +-- | *SDK2* |
| cg21117965 | *CNOT9* | girls | -0.07 | -0.10 | 0.09 | --+ | *SPEG* |
| cg13002603 | *TSC22D2;SERP1* | girls | -0.04 | -0.11 | 0.04 | --+ | |
| cg14593760 | *LINC01924;CDH7* | girls | 0.07 | -0.10 | -0.07 | +-- | |
| cg22016234 | *TMCC2;NUAK2* | girls | -0.11 | -0.12 | 0.07 | --+ | *TMCC2* |
| cg14735070 | *C8orf37-AS1* | girls | -0.08 | 0.13 | 0.06 | -++ | *SDC2* |
| cg02733814 | *PTPRG* | girls | 0.05 | 0.13 | -0.08 | ++- | *PTPRG* |
| cg14056226 | *PRICKLE2;ADAMTS9* | girls | 0.05 | -0.13 | -0.05 | +-- | |
| cg22698115 | *KRTAP5-6;IFITM10* | girls | 0.08 | -0.11 | -0.08 | +-- | *HCCA2* |
| cg01868577 | *NR2F1* | girls | -0.07 | 0.12 | 0.07 | -++ | |
| cg04404543 | *RARG* | girls | 0.03 | -0.10 | -0.03 | +-- | *ATF7;ATF7* |
| cg07464291 | *KLHL11* | girls | -0.05 | -0.14 | 0.07 | --+ | *SF3;CSF3;CSF3;CSF3;CSF* |
| cg12764291 | *AIG1* | girls | -0.03 | -0.10 | 0.05 | --+ | |
| cg18966495 | *CASC15;PRL* | girls | -0.03 | -0.10 | 0.04 | --+ | |
| cg18270687 | *MIR3659;LINC01343* | girls | -0.06 | -0.10 | 0.09 | --+ | |

| | | | | | | | |
|---|---|---|---|---|---|---|---|
| cg13955644 | *VWCE* | girls | -0.02 | 0.11 | 0.05 | -++ | |
| cg22049450 | *MCRS1* | girls | -0.03 | -0.11 | 0.05 | --+ | *AQP2;LOC101927318* |
| cg02556345 | *LINC01561;WDR11-AS1* | girls | -0.03 | -0.12 | 0.04 | --+ | *PLEKHA1* |
| cg22389845 | *GRIP2* | girls | -0.03 | -0.11 | 0.05 | --+ | *GRIP2* |
| cg16109426 | *RNF38;MELK* | joint | 0.11 | 0.12 | -0.14 | ++- | |
| cg18295744 | *KCNMA1-AS3* | joint | 0.12 | -0.11 | -0.17 | +-- | *ZMIZ1* |
| cg10355819 | *LINC02578* | joint | 0.12 | 0.12 | -0.09 | ++- | *GGTA1P;GGTA1P* |
| cg02723558 | *LOC105376671;LIN7C* | joint | 0.17 | -0.11 | -0.12 | +-- | *BDNFOS* |
| cg03876023 | *UBR5* | joint | -0.22 | 0.15 | 0.07 | -++ | |
| cg05616242 | *HOXC-AS2* | joint | 0.10 | -0.11 | -0.07 | +-- | *ZNF385A;LOC102724050* |
| cg19944895 | *SLC22A24;SLC22A25* | joint | -0.12 | 0.11 | 0.12 | -++ | *CHRM1* |
| cg03715269 | *GPC6* | joint | -0.10 | -0.11 | 0.07 | --+ | |
| cg08431289 | *LINC01194* | joint | -0.20 | -0.14 | 0.11 | --+ | *LINC01194* |
| cg02069095 | *GPR39* | joint | -0.12 | -0.14 | 0.09 | --+ | *NCKAP5;NCKAP5* |
| cg04067341 | *IRS2;LINC00396* | joint | 0.12 | 0.11 | -0.12 | ++- | *COL4A2-AS1;COL4A2* |
| cg05268743 | *LOC643339* | joint | 0.11 | -0.10 | -0.12 | +-- | |
| cg22026677 | *MIR8056;LOC285593* | joint | 0.10 | -0.11 | -0.16 | +-- | *ERGIC1* |
| cg16943230 | *RPS6KA5* | joint | 0.14 | 0.21 | -0.05 | ++- | *PPP4R3A* |
| cg21503763 | *B3GNT2* | joint | 0.13 | -0.15 | -0.12 | +-- | |
| cg08849509 | *SBF2-AS1* | joint | 0.11 | -0.13 | -0.15 | +-- | *SBF2* |

**Supplementary Table 9**

Look-up of CpGs in common between our sex-stratified EWAS of ePTB and the meta-analysis of gestational age reported by Merid *et al.* 2020

| CpG | Near Gene | ePTB EWAS analysis | Coefficient ePTB boys | Coefficient ePTB girls | Coefficient GA Merid | Directionality ePTB Boys-Girls-Merid | Gene Name Merid |
|---|---|---|---|---|---|---|---|
| cg01890836 | *COPS3* | boys | 0.100 | -0.029 | -0.0002 | +-- | *MPRIP* |
| cg23825480 | *SEC14L6* | boys | -0.103 | 0.021 | -0.001 | -+- | *MORC2-AS1* |
| cg02635407 | *ABLIM2* | girls | 0.064 | -0.126 | -0.0004 | +-- | *SH3TC1* |
| cg08655953 | *TNFRSF14* | girls | -0.070 | -0.223 | -0.0003 | --- | *PLCH2* |
| cg03662398 | *MSI2* | girls | -0.070 | -0.117 | 0.0004 | --+ | *HLF* |
| cg04193923 | *KLHL11* | girls | -0.021 | -0.103 | 0.0005 | --+ | *CSF3* |
| cg08888956 | *ALX1* | girls | -0.014 | -0.103 | 0.001 | --+ | *NTS* |
| cg04744624 | *NRCAM* | girls | -0.056 | -0.174 | 0.0004 | --+ | *LAMB1* |
| cg27071707 | *TCF3* | girls | -0.020 | 0.109 | 0.001 | -++ | *TCF3* |
| cg17806418 | *MOG* | girls | -0.046 | 0.119 | 0.001 | -++ | *GABBR1* |
| cg13555278 | *MAN1C1* | girls | -0.032 | -0.121 | 0.001 | --+ | *EXTL1* |
| cg13922488 | *LINC01841* | girls | 0.036 | 0.118 | -0.0005 | ++- | *PKN1* |
| cg23904955 | *ZFYVE27* | girls | -0.032 | -0.105 | 0.001 | --+ | *DQ372722* |
| cg19143990 | *GLDN* | girls | 0.018 | 0.106 | -0.0003 | ++- | *LYSMD2* |
| cg08449748 | *ECI1* | girls | -0.026 | -0.114 | 0.0004 | --+ | *ABCA3* |
| cg02556345 | *LINC01561* | girls | -0.030 | -0.124 | -0.001 | --- | *MIR3941* |
